# Genomic landscape of SARS-CoV-2 pandemic in Brazil suggests an external P.1 variant origin

**DOI:** 10.1101/2021.11.10.21266084

**Authors:** Camila P. Perico, Camilla R. De Pierri, Giuseppe P. Neto, Danrley R. Fernandes, Fabio O. Pedrosa, Emanuel M. de Souza, Roberto T. Raittz

**Author notes:** Corresponding authors: Roberto T. Raittz.

## Abstract

Brazil was the epicenter of worldwide pandemics at the peak of its second wave. The genomic/proteomic perspective of the COVID-19 pandemic in Brazil can bring new light to understand the global pandemics behavior. In this study, we track SARS-CoV-2 molecular information in Brazil using real-time bioinformatics and data science strategies to provide a comparative and evolutive panorama of the lineages in the country. SWeeP vectors represented the Brazilian and worldwide genomic/proteomic data from GISAID between 02/2020 – 08/2021. Clusters were analyzed and compared with PANGO lineages. Hierarchical clustering provided phylogenetic and evolutionary analysis of the lineages, and we tracked the P.1 (Gamma) variant origin. The genomic diversity based on Chao’s estimation allowed us to compare richness and coverage among Brazilian states and other representative countries. We found that epidemics in Brazil occurred in two distinct moments, with different genetic profiles. The P.1 lineages emerged in the second wave, which was more aggressive. We could not trace the origin of P.1 from the variants present in Brazil in 2020. Instead, we found evidence pointing to its external source and a possible recombinant event that may relate P.1 to the B.1.1.28 variant subset. We discussed the potential application of the pipeline for emerging variants detection and the stability of the PANGO terminology over time. The diversity analysis showed that the low coverage and unbalanced sequencing among states in Brazil could have allowed the silenty entry and dissemination of P.1 and other dangerous variants. This comparative and evolutionary analysis may help to understand the development and the consequences of the entry of variants of concern (VOC).

## Introduction

The current pandemic of the SARS-CoV-2 virus (Severe Acute Respiratory Syndrome Coronavirus 2), which causes the disease known as Corona Virus Disease 2019 (COVID-19) (1), had its first cases notified in Brazil in February 2020. Brazil was the pandemic’s epicenter at the peak of its second wave, around April 2021.

New variants continually emerge, many of them considered Variants of Concern (VOC) such as the British B.1.1.7 (Alpha), the South African B.1.351 (Beta), the Indian B.1.617.2 (Delta), and the P.1 (Gamma), first identified in Brazil in November 2020 (2). In addition, variants acquire mutations that make them more adapted, transmissible and challenging to detect by the immune system (3; 4; 5). Therefore, virus monitoring is essential to diagnose, improve treatment, characterize strains and sub-strains, and thus understand their dynamics and dispersion (6). It is also of utmost importance in health policy decisions. International and domestic travel without quarantine is a significant vehicle for spreading potentially dangerous variants, as occurred at the beginning of the pandemic in 2020 before air travel restrictions (7). Proper quarantine use positively impacted case reduction, and neglected quarantine caused exponential growth in infected curves (8).

Franceschi *et al*. (9) brought the Brazilian panorama until February 2021, when it completed a year of the pandemic in Brazil. The authors analyzed mutations, phylogeny, and phylogeography of the virus in the Brazilian context by exploiting conventional bioinformatics tools, with a genomic focus, analyzing 2,732 sequences. The viral sequence data is immense, reaching more than 3.4 million genomes sequenced worldwide by September 2021 in the GISAID Database (10), numbers that are in constant growth.

Current methods based on sequence alignment cannot process large volumes of data due to the exponential growth of the computational cost. Conventional bioinformatics is not enough to thoroughly analyze large volumes of data. However, data mining and machine learning methods can be decisive in extensive data analysis, providing reliable and fast results. These methods are already widely used, with several applications in different areas, including the taxonomic classification of coronavirus genomes (11; 12).

Previous studies have shown that alignment-free methods, particularly vector representation of biological sequences, are fast, scalable, and effective in analyzing SARS-CoV-2 sequences and efficient in associating with machine learning methods (12; 13; 14; 15). Vector representation of biological sequences is an emerging method that facilitates the implementation of data science techniques and has already proven effective in applications in bioinformatics (14; 15; 16; 17; 18).

This study intended to understand how the emergence and extinction of SARS-CoV-2 lineages occur and verify if the variants in the databases are correctly defined. As suggested in the correlated studies, the terminology PANGO (or PANGOLIN) was adopted (19; 20). We constructed a pipeline in R language based on the application of vector representation, data mining, and machine learning methods to obtain the current panorama of the pandemic in real-time and to understand the evolution of the virus in Brazil. To understand the virus evolutive process, we tested the hypothesis of the external origin of the P.1 variant and the possibility of whether or not a recombination event was involved in its origin. Furthermore, to facilitate monitoring and adequate decision-make action, we investigated whether the our pipeline is suitable for the early detection of the emergence of new strains.

## Materials and Methods

Figure **S1** presents the pipeline constructed in R language that is available at https://github.com/CamilaPPerico/SARS-CoV-2_Brazil_Landscape/, as well as the other results of this research. The sequences used in this paper, excepting the Wuhan reference sequence, were downloaded from the GISAID database and represented into vectors. Euclidean is the adopted metric for distance in this study. We ran the analysis on a Xeon server with 251Gb of RAM and 40 threads.

### Obtaining and pre-processing of SARS-CoV-2 sequences

We downloaded the proteomes of SARS-CoV-2 and the sequences corresponding to the Brazilian genomes from GISAID (https://gisaid.org/) (21). The PANGO nomenclature^1^ (22) was adopted. The sequences were obtained from GISAID in three different moments, with its corresponding PANGO designation: a) initial analysis (GISAID release 409, PANGO v.2.3.8 2021-04-20); b) principal analysis (release 609, PANGO v3.0.5 2021-06-04); and c) final update (release 829, PANGO v.3.1.11 2021-08-09). The Wuhan reference sequence (NC 045512.2) is from the NCBI^2^ database.

The addressed sequences from Brazil, Italy, India, Germany, and England, correspond to the period from the pandemic onset to the end of May 2021, while other worldwide considered sequences were from 2020 only. All incomplete proteomes and the sequences with misreading were not considered. However, when only one protein was absent, it was accepted (23). Table 1 shows the number of samples before and after filtering.

**Table 1.** Relationship between the number of sequences analyzed per country and the computational time for the vectorization. Total: total number of samples present in GISAID in release 609 (excepting World 2020 from release 409); Quality: number of filtered vectors used for analysis; Non-redundant: number of unique vectors; rSWeeP: computational time in minutes to project the filtered sequences.

| Country | total | quality | non-redundant | rSWeeP (min) |
| --- | --- | --- | --- | --- |
| Brazil | 13 395 | 8720 | 6146 | 12.2 |
| India | 18 558 | 7154 | 5493 | 17.4 |
| Italy | 33 014 | 12 784 | 6617 | 18.4 |
| Germany | 126 794 | 51 880 | 22 048 | 74.1 |
| England | 353 330 | 199 110 | 62 643 | 274.3 |
| World (release 409) | 1 000 558 | 493 080 | 312 224 | 22 hours |

### Mutations

We searched for Brazilian sequences mutations using the web platform Nextstrain^3^ from FASTA nucleotide files (24). Mutations statistics by cluster and by lineage were performed, considering only the Brazilian context. The characteristic mutations for each group were considered when present in more than 75% of the respective cluster or lineage samples.

### Sequences vectorial representation

Protein sequences were concatenated (with border delimiters) into proteomes which were represented in vectors using the SWeeP tool (Spaced Words Projection) (14). The R version of SWeeP tool, used for the proteome vectorization, is available in the Bioconductor Platform^4^ for R version 3.12 (25). Finally, we made the vector projection of the Brazilian genomes (coded in DNA) in the SWeeP tool in Matlab^@^ (14) with its default parameters.

The 1,000,588 (1M) of SARS-CoV-2 worldwide proteomes were vectorized, totaling 9.97 billion amino acids, including the reference sequence of Wuhan and the spike protein of Brazilian samples separately integrated into the comparative study. The proteomes of Brazil, Germany, India, Italy, England, and World-2020 were vectorized and considered as independent sets. The same orthonormal base, with the SWeeP default parameters (length 600 and mask [1 1 0 1 1]), was employed to project all sequences into compacted vectors.

### Cluster analysis and visualization

Brazilian proteomes were clustered using the ConsensusClusterPlus package version 1.54.0 from Bioconductor (26) and the kmedoids method (Partitioning Around Medoids, PAM), in procedures with 1,000 replicates for each cycle, testing 2-20 as the number of clusters. For the spike proteins, 2-10 sets were tested. As a criterion for selecting the best number of groups, in both cases, was considered the best convergence in the Consensus Cumulative Distribution Function (CDF) with the smallest number of clusters. We visualized and compared the clustering results using two approaches of dimensionality reduction: Principal Component Analysis (PCA) and the t-Distributed Stochastic Neighbor Embedding (t-SNE) (27). The t-SNE diagrams were constructed in the Rtsne package^5^, with its default parameters.

Information on the number of COVID-19 cases in Brazil was available at the official website https://covid.saude.gov.br/. In addition, the mapping of temporal and spatial evolution was carried out based on information obtained from the metadata provided by the GISAID platform.

### Diversity Analysis

Coverage and richness of viral subvariants (unique and non-redundant sequences) were estimated via the Chao 1 richness estimator, given by the equation 1 (28; 29).

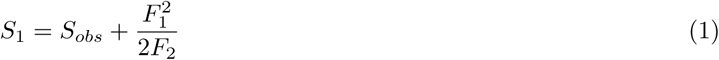

In equation 1, *S*_*obs*_ is the number of distinct vectorized proteomes observed, *F*_1_ is the number of singletons (single-occurring vectors), and *F*_2_ is the number of doubletons (two-occurring vectors). Thus, the coverage is given by:

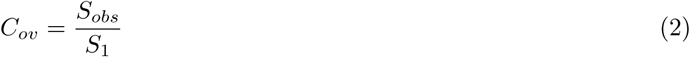

### Phylogenetic analisys

All proteomic phylogenetic trees were built using the Neighbor-Joining (NJ) method through the Ape version 5.5 package (30), performing bootstrap with 1000 replicas. Only branches with bootstrap greater than 70% were considered. For previous studies employing bootstrap calculation in tree construction in alignment-free analyses, see references (31; 32).

We built a consensus phylogenetic tree for the 8,720 Brazilian proteomes based on the proteome’s vectors distance matrix. Also, phylogenetic trees were built for cluster and lineage centroids, selecting the sequence closest to each corresponding centroid and taking it as a representative vector. The centroids were obtained by the average of the vectors within the cluster/lineage.

The proteomic results were compared to a phylogenetic tree with the aligned genomes of the clusters and lineages centroids. We also aligned the specific sequences and constructed genome trees to analyze the origin of the P.1 variant. For this step, the Maximum Likelihood method of the MEGAX 10.2.6 (33) tool, with a 500 bootstrap of size, was performed using the Jukes-Cantor nucleotide substitution model (34). The alignment was made using the Nextclade online tool (24). All the phylogenetic trees were rooted using the Wuhan reference sequence (NC 045512.2) as the outgroup. Finally, we visualize the trees in the iTOL tool^6^ (35) and with the ggtree package (36).

### P.1 variant origin analysis

Intending to determine the origin of the P.1 variant, whether internal or external to Brazil, we obtained the 70 closest worldwide samples to each of the 50 P.1 Brazilian samples in 2020 by distance, resulting in 91 unique vectors whose phylogeny by alignment was analyzed. We also searched for occurrences of sequences like P.1 in the World before its emergence in Brazil. Finally, we assessed the involvement of the P.1 variant in possible recombination events.

### Machine Learning for P.1 search

An ensemble of 50 feed-forward neural networks (multilayer perceptron, MLP) was trained using the vectors of the Brazilian sequences classified as P.1 and non-P.1 utilizing data from release 609 with classification PANGO v.3.0.5. Data division was 70:30 for training and testing sets, respectively, randomly divided for each neural network training. Each MLP network contained the layers input, middle, and output with 600,3,1 neurons, respectively. Only networks with an f1-score higher than 90% as threshold compose the ensemble. A majority vote decided classification. We tested the model with the complete set of Brazilian vectors of accuracy, f1-score, recall, and precision through cross-validation.

### Recombinant’s detection

Possible recombinants were detected from aligned genomes using RAPR (37) and RDP4 (38) tools. RDP4 provides the methods RDP (39), BOOTSCAN (40), MAXCHI (41), CHIMAERA (42), 3SEQ (43), GENECONV (44), LARD (45), and SISCAN (46), applied in this task. The confirmation test for the recombinant events was performed by analyzing the phylogenetic trees of genomes. The genomes were split into two parts at the breaking points of the aligned sequences and, we phylogenetically analyzed the relative position between supposed recombinants and their parents. Finally, the recombinants that presented a distinct relative position between the trees of each segment were validated (47).

## Results

From the 1,000,558 samples worldwide in release 409 on the GISAID platform (21), 49% of proteomes remained after filtering (493,080 sequences), of which 260,759 from 2020. In Brazil, 65% of samples were considered, a quality percentage higher than the world average and the other countries studied, as shown in Table 1. The 8,720 vectorized Brazilian sequences were analyzed and discussed below. More detailed information on the results is in Section **2** of Supplementary **1**. The complete metadata of the Brazilian sequences and the metadata referring to other countries and the World is available at the Github link.

### Landscape in Brazil

The ConsensusClusterPlus analysis returned 15 clusters representing the epidemic proteomes in Brazil from 25 February 2020 to the end of May 2021. More than 15 clusters do not provide a considerable increase in the consensus value of the CDF curve (less than 5% – Figure **S2**). The main lineages identified in Brazil according to the PANGO nomenclature are: P.1 (3572 – 40.9%), P.4 (1274 – 14.6%), P.2 (1132 – 13.0%), B.1.1.33 (909 – 10.4%), B.1.1.28 (864 – 9.9%), B.1.1.7 (248 – 2.8%), B.1.1 (186 – 2.1%), P.1.2 (153 – 1.7%), N.9 (81 – 0.9%), B.1 (65 – 0.7%), B.1.195 (54 – 0.6%), other (178 – 2.0%). Some variants were completely grouped in single clusters (B.1.1.7, P.2, P.1.2, B.1.1.33), while others occurred in various groups divided into subvariants (B.1.1.28 in clusters 1, 5, and 6; P.1 and P.4 in clusters 3,7,9,10,14 and 15). Rarer variants were mainly grouped in clusters 2 and 6. Cluster 2 is composed of lineages less frequent in Brazil, including the basal lineages A.1, A.2, and B and B.1, which have 1,3,3 and 59 samples, respectively. The Wuhan reference sequence belongs to cluster 2 and is highlighted in Figure 2.

The analysis showed that the clustering approach respects evolutionary similarity among the sequences. Moreover, the clustering results match the PANGO division, as viewed in the t-SNE diagram, PCA, and clusters/lineage centroids heatmap (Figures 1, 2). Tables 2 and **S2** show the relationship between the clusters and their main composition. Other results are presented in the supplementary material (Table **S1**).

**Figure 1.**
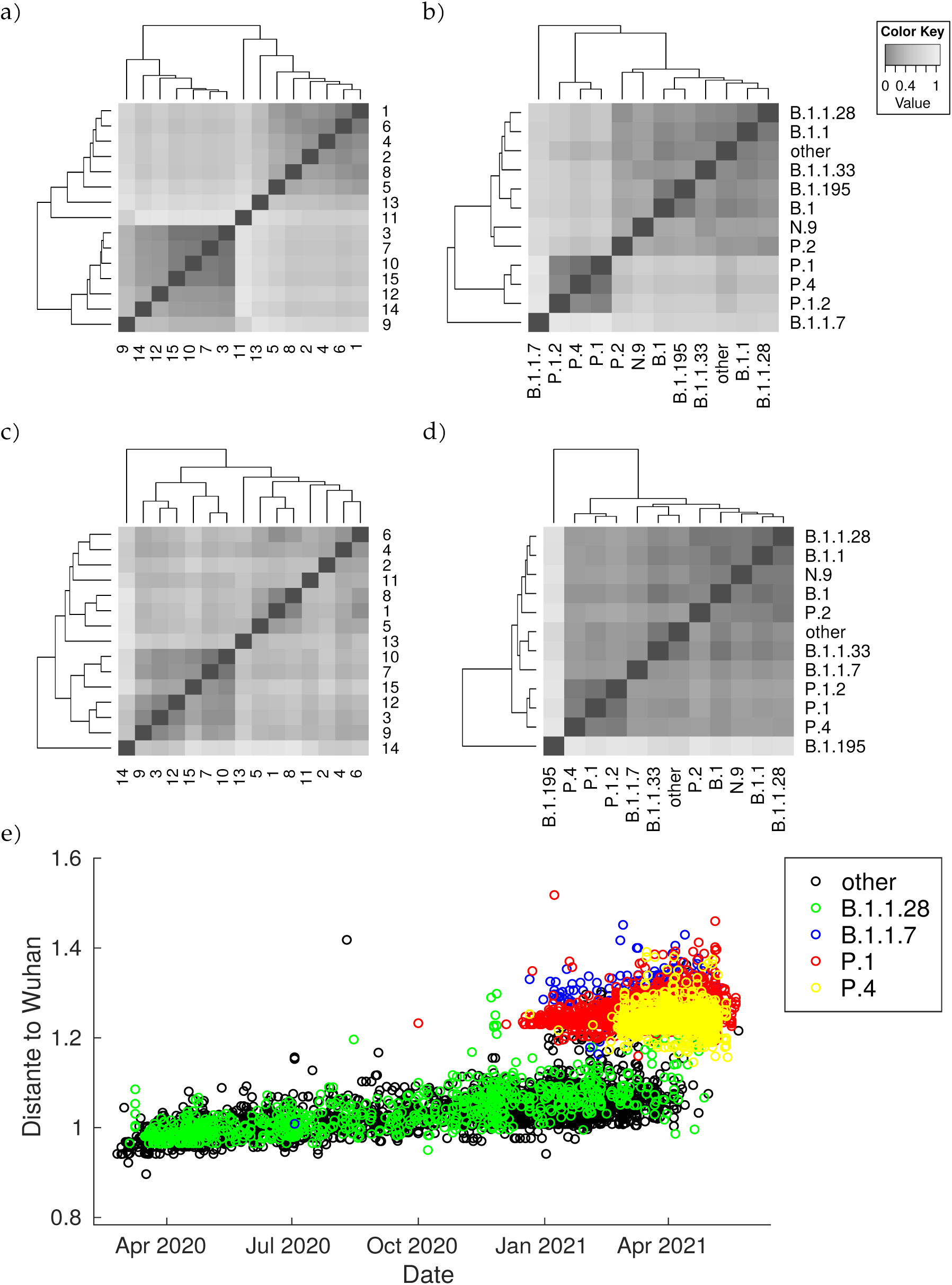
Heatmap of the centroid distance matrix. Distances regarding genomes and proteomes were analyzed and grouped by lineages and by clusters. The images below correspond to **a)** proteomes by clusters; **b)** proteomes by lineages; **c)** genomes by clusters; **d)** genomes by lineages. The image **e)** corresponds to the Euclidean distance between the Wuhan vectorized sample and the Brazilian ones against time. There is a considerable gap between the Brazilian sequences in general (B.1.1.28 and other variants from T0) to the TP1 group (P.1 and P.4 according to PANGO v3.0.5) and different imported sequences (B.1.1.7). The outlier sequences were removed from the visualization.

**Figure 2.**
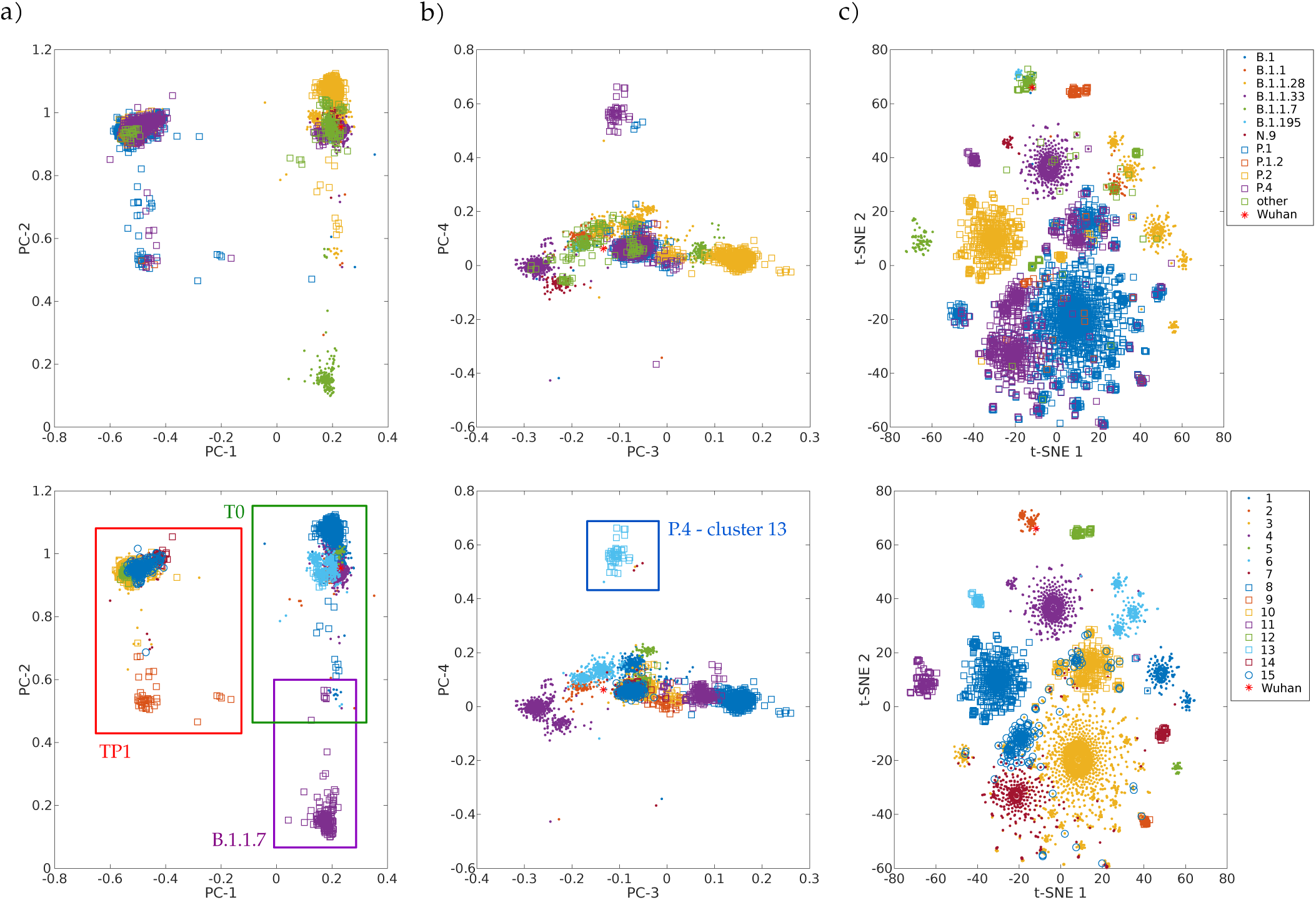
Overview of the relationship between SARS-CoV-2 proteomes of Brazilian samples. The colors represent main lineages (above) and clusters (below) for the Brazilian sequences for each image pair. The red asterisk highlights the position of the Wuhan reference sequence. **a)** components 1 and 2 of PCA graph – note the cluster on the left is the TP1 group, composed of sequences from P.1, P.1.1, P.1.2, and P.4; **b)** components 3 and 4 of PCA graph – the cluster 13, isolated above, corresponds to the 79 samples identified as P.4 by PANGO (v3.0.5). The TP1 group is more central, and the other clusters are around it; c) t-SNE graph – note clustering coincides with classification by lineages.

**Table 2.** Division of lineages into clusters using the complete vectorized proteome. The predominant strains in each cluster are listed with the date of their first and last sample. The complete list of observed lineages by cluster is available in Table **S2**.

| cluster | predominant lineage | number of samples | first case | last case |
| --- | --- | --- | --- | --- |
| groups related to the P.1 variant – TP1 |  | 5000 | 2020-10-01 | 2021-05-20 |
| 3 | P.1 | 3062 | 2020-10-01 | 2021-05-20 |
| 7 | P.4 | 754 | 2020-12-21 | 2021-05-07 |
| 9 | P.1 + P.4 | 50 | 2020-12-23 | 2021-05-06 |
| 10 | P.1 + P.4 | 665 | 2021-01-11 | 2021-05-10 |
| 12 | P.1.2 | 104 | 2021-02-22 | 2021-05-17 |
| 14 | P.1 | 81 | 2021-01-16 | 2021-05-13 |
| 15 | P.4 | 284 | 2021-02-19 | 2021-05-19 |
| early group – T0 |  | 3391 | 2020-02-25 | 2021-05-22 |
| 1 | B.1.1.28 | 477 | 2020-03-09 | 2021-04-27 |
| 2 | B.1.195 + other | 179 | 2020-02-25 | 2021-05-22 |
| 4 | B.1.1.33 + N.9 | 1037 | 2020-03-01 | 2021-04-19 |
| 5 | B.1.1.28 | 77 | 2020-07-31 | 2021-04-03 |
| 6 | B.1.1.28 + B.1.1 + other | 531 | 2020-02-28 | 2021-04-25 |
| 8 | P.2 | 1090 | 2020-04-13 | 2021-04-30 |
| others |  | 329 | 2020-12-21 | 2021-05-14 |
| 11 | B.1.1.7 | 250 | 2020-12-21 | 2021-05-06 |
| 13 | P.4 | 79 | 2021-02-17 | 2021-05-14 |

### Groups analysis

The Brazilian samples are divided into two main groups: the early Brazilian group with 3,391 representatives (here named T0); and the representatives related to the P.1 variant with 5,000 samples (TP1). In addition, two specific clusters that consistently drift apart from the other samples are cluster 13 – emerging P.4, within 79 pieces; and cluster 11 – imported variant B.1.1.7, within 250 samples. The t-SNE diagram (Figure 2c) shows partially overlapping clusters 3,7,10, and 15 – composed mainly of variants P.4 and P.1 – this likely occurred due to dimensionality reduction. The clusters 1, 2,4,5,6 and 8 compose the T0 group (Figure 2a, right) and the clusters 3,7,9,10,12,14 and 15 TP1 (left). In particular, the 1×2 components of PCA shows group B.1.1.7 below far apart, and 3×4 components of PCA (Figure 2b) show cluster 13 above, far away from the others.

We also vectorized and clustered spike proteins sequences which derived eight consensus clusters (Figure **S3**, Tables **S3**,**S4**). The clustering of the spike proteins was similar to those of the complete proteomes but with fewer divisions. Nevertheless, the division into two larger groups is maintained, and clusters 11 and 13 are still differentiable, as shown in the PCA (Figure **S3b**).

The consensus mutations for all SARS-CoV-2 Brazilian samples, the characteristic mutations for the TP1 group, and for the other clusters are presented in Tables **S5, S6** and **S7**, respectively, and can be visualized in the heatmaps of Figures 3 and **S5** for clusters and lineages. More detailed information is available in Section **2.2** of Supplementary **1**.

**Figure 3.**
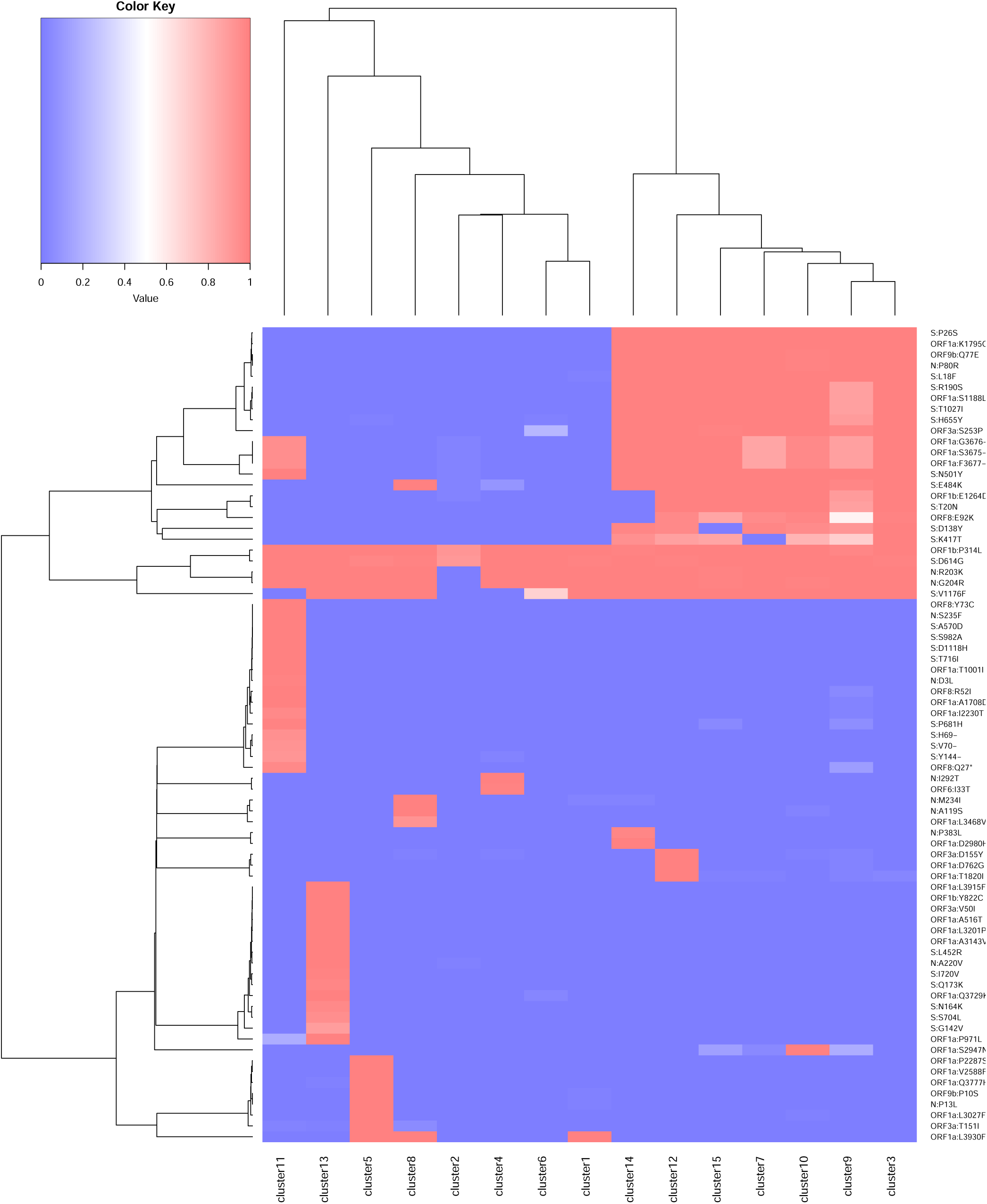
Heatmap of mutations by cluster. Mutations present in 75% of the samples from one or more clusters are listed. The value 1 (red) represents the presence of the mutation in 100% of the cluster samples, and the value 0 (blue) indicates the absence. Values are normalized by cluster.

#### Early group (T0)

The T0 group (clusters 1,2,4,5,6 and 8) is composed of clusters of sequences from the early entry of the virus in Brazil at the beginning of 2020, and daughter lineages evolved locally. The group is mainly composed of B.1.1, B.1.1.28, B.1.1.33, P.2, N.9, and N.10 (Table **S2**). These are the older groups that predominated in Brazil in 2020, but are almost extinct, giving way to the TP1 group (Figure 4). There are no consensus mutations characteristic in T0 (Tables **S5, S7**); each cluster represents an individual lineage or a group of lineages less frequent in Brazil. One example of a variant belonging to T0 is the B.1.1.33, which stood out the most in 2020 in Brazil. Franceschi *et al*. (9) suggest that this variant (B.1.1.33) probably was originated in Europe and later spread into America. The Brazilian B.1.1.33 sequences are closer to the B.1.1 sequences found in Switzerland (EPI_ISL_415454, EPI_ISL_524474, EPI_ISL_415700, EPI_ISL_415457, EPI_ISL_429203), Czech Republic (EPI_ISL_416743, EPI_ISL_895731) and Netherlands (EPI_ISL_454750), corroborating its European origin (9).

**Figure 4.**
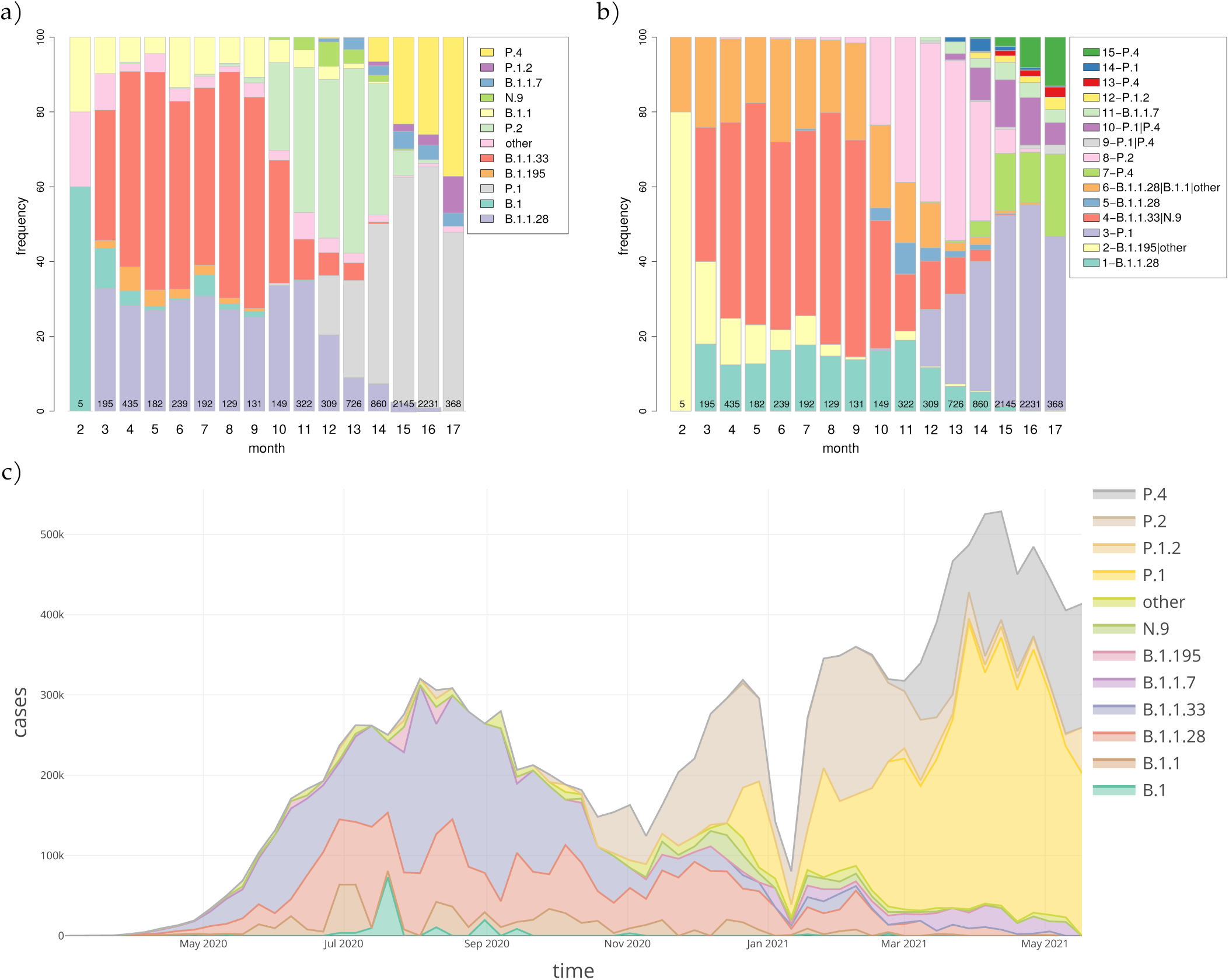
Temporal distribution of **a)** lineages and **b)** clusters in Brazil are represented as proportional stacked bar charts divided by months from February 2020 to May 2021. Additionally, in **c)**, the number of COVID-19 cases registered in Brazil proportionally divided by lineages. Currently, in Brazil, there is a considerable increase in the number of clusters possibly linked to the diversification of variants, and the strains P.1, P.4, and P.1.2 stand out. The values inside the bars indicate the number of sequencing performed each month. PANGO version 3.0.5 was used.

#### Groups related to variant P.1 (TP1)

TP1 comprises clusters within variants P.1, P.4, P.1.1, and P.1.2 (clusters 3,7,9,10,12,14 and 15). The first P.1 was notified in 12/2020, though previous studies estimate that P.1 origin in Brazil occurred between early October and mid-November 2020 (2). The P.1 sample (EPI_ISL_2241496) dated 2020-10-01 from Paraíba State corroborates this hypothesis. October/2020 was one of the months with the lowest sequencing in Brazil, which can be the sake of underreporting of P.1 related cases. Remarkably, this month was a period of flexibilization of international flights in Brazil (48).

Phylogenies show that P.1 and P.4 variants mix themselves among and inside clusters in TP1 (Figure **S6**). In t-SNE, PCA, and heatmaps, P.1 and P.4 are hardly distinguishable, either in clusters or lineages (Figures 1 and 2). Furthermore, the TP1 clusters share many non-synonymous mutations (Table **S6** and Figure 3). At least five of these mutations are in the spike protein, conferring the adaptive virus advantage (E484K, N501Y, K417T, H655Y, and L18F) (49; 50; 51; 52). The sum of these characteristics suggests that the TP1 group could be seen as a single lineage, divided into sublineages. As stated before, it is also remarkable that clusters within TP1 do not correspond perfectly to the P.1 and P.4 subdivision provided by PANGO.

#### Cluster 11 **–** variant B.1.1.7

Cluster 11 is composed of B.1.1.7, totaling 250 sequences, which had its first case identified in Brazil in 2020-12-21. The characteristic mutations of the group correspond to those found in the literature (53) (Table **S7**). Furthermore, the smallest distances between cluster 11 and world-2020 samples indicate their closest similarity with sequences from England (EPI_ISL_799516, EPI_ISL_1248398, EPI_ISL_760286, EPI_ISL_797822, EPI_ISL_799518), all belonging to the British B.1.1.7 variant. Therefore, it reinforces the possibility that the entry of the variant in Brazil occurred directly from England.

#### Cluster 13 – variant P.4

Cluster 13 comprises 79 sequences classified as P.4, as designated by PANGO v3.0.5 (2021-06-04); however, mutations do not correspond to the TP1 group to which the P.4 variant belongs (Figure 3). Later modifications in the PANGO nomenclature (v3.1.11 2021-08-09) changed P.4 classification that will be covered in more detail in the discussion. This cluster is an attention-grabbing group because it contains many unique mutations, three of which are of concern (Table **S7**). This group of mutations was not found in other locations but only in Brazil (according to a search carried out on Outbreak.info). The 3×4 components of PCA (Figure 2b) place the cluster 13 group away from the other clusters in the same way that occurred with the samples from B.1.1.7, indicating a possible late entry, but we couldn’t track its origin.

### Phylogenetic Trees

Although most analyzes of this study were performed based on proteomes samples, complete genome DNA trees were built for comparison. The results showed that the proteome and genome derived trees with 8,720 samples generally agree (Figure **S6**) – complete trees are available in Supplementary **5** and **6**.

The consensus tree consistently grouped the monophyletic branch of the TP1 group with 100% bootstrap (BP). The branch containing cluster 12 (P.1.2), internal to the branch of the TP1 group, is monophyletic and obtained a 100% BP. The cluster 11 (B.1.1.7) – with BP=87% – and the lineages N.9 and N.10 of cluster 4 – with BP=100% both – are also monophyletic (tree available in Github as SARS_NJ_Consensus_BP.nwk). Centroids-based phylogenies provided a cleaner and more reliable evolutionary overview of the groups with high bootstrap (Figure 5). TP1 group appears together in all tested centroid trees with high bootstrap.

**Figure 5.**
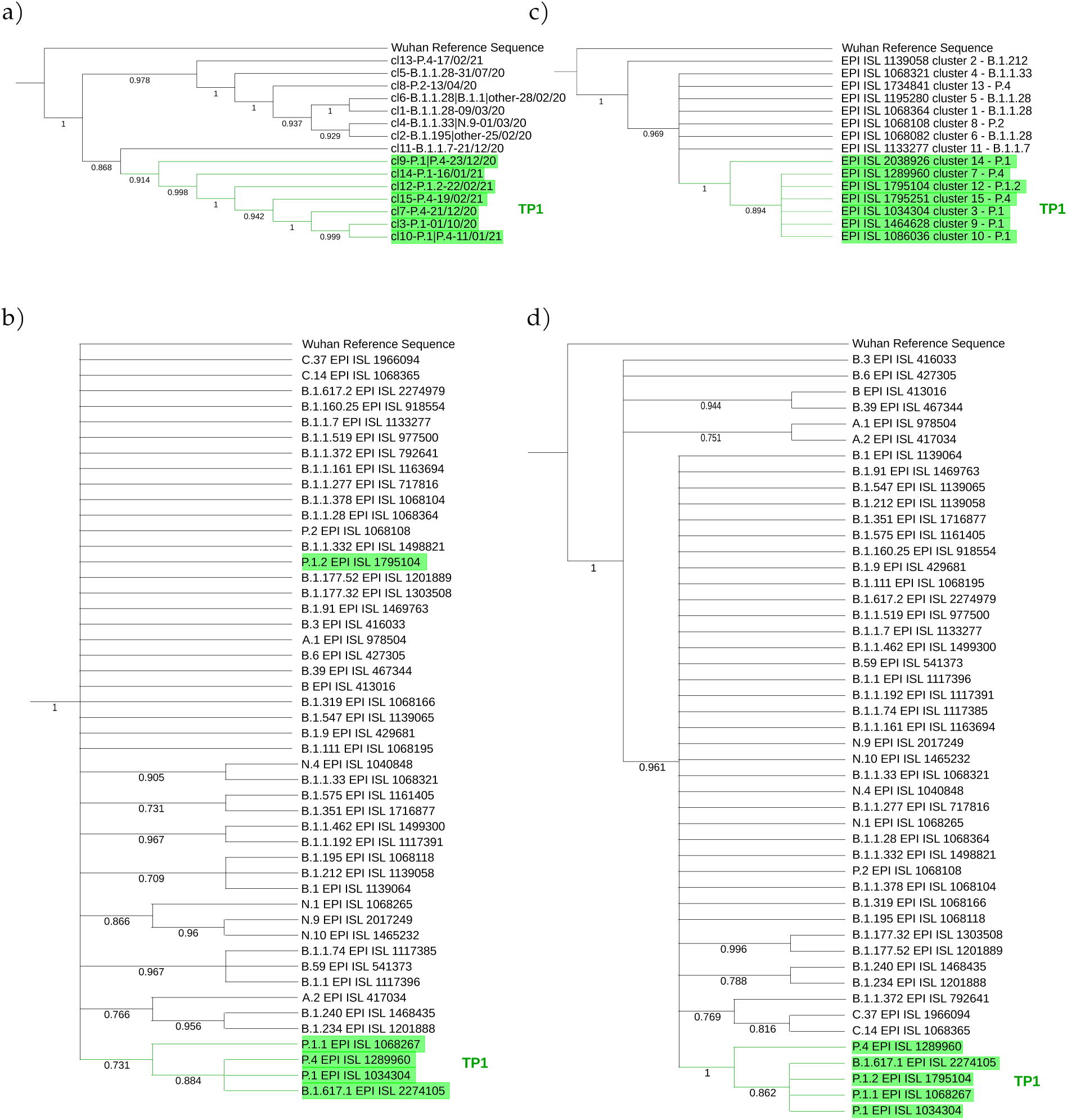
Phylogenetic trees by cluster and lineage centroids (relative to PANGO v3.0.5) with a bootstrap of 1000 replicas. **Left**, the Neighbor-Joining method with Euclidean distance was used on the proteome centroids corresponding to each **a)** cluster, **b)** lineage. **Right**, centroid trees per cluster by DNA via Maximum Likelihood with JukesCantor method, by **c)** cluster, lineage. The tree **a)** is divided into two main branches: the TP1 group, with cluster 11 of B.1.1.7 as more basal, followed by cluster 9; and the other main branch, the T0 with cluster 13 as the most basal. All branches reach bootstrap greater than 85%. The **b)** is less consistent, but the TP1 group reaches a 73% bootstrap, except the branch of P.1.2, which does not get high BP. The **c)** consistently inserts cluster 14 as basal in the TP1 group and cluster 2 as more basal in Brazil. Finally, tree **d)** obtains BP of 100% for the branch of the TP1 group and inserts as basal the variants A.1, A.2, B, B.3, B.6, and B.39 consistently.

The TP1 group is cohesive and monophyletic in all approaches, and the P.4 lineage does not differ from P.1, as there is an alternation of branches in all trees, both in genome and proteome (Figure **S6**). The basal cluster of the TP1 group is uncertain, varying between cluster 14 in the genomic approach and cluster 9 in the proteomic one. Cluster-based methods reached higher bootstrap values compared to those based on the lineage (Figure 5).

Kappa variant B.1.617.1 samples appear together within the TP1 group in the DNA and the protein trees (Figures 5b and d), which probably consists of annotation errors once these samples have the characteristic mutations of the P.1 variant rather than B.1.617.1 (Table **S1**).

### P.1 variant origin analisys

We investigated three hypotheses for the P.1 variant origin: a) it evolved locally, i.e., from T0, b) it had a later entry external origin (came from abroad); and c) P.1 is derived from some recombination event.

From each of the 50 P.1 samples (Brazilian), we take the 70 closest vectors in the set of World proteomes of 2020. This search identified 91 unique sequences, including 6 Peruvians P.1 and 17 Brazilian B.1.1.28 samples, and others from several countries – listed in Table **S8**. This isolated information would indicate that P.1 is closely related to the B.1.1.28 sequences from the Pará (PA) and São Paulo (SP) States, supporting the local ancestry hypothesis previously reported (54). However, the two samples here nominated as PA-TP1 (EPI_ISL_1068256 and EPI_ISL_1261122) have as closest sequences only foreign samples Table 3. These samples also appeared close to all other searched P.1 samples then we deepened the analysis. The PA-TP1 genomes have 10 of the 17 characteristic mutations of the P.1 group, and both instances belong to cluster 9. Thus, the phylogeny suggests that PA-TP1 may be the precursors of the TP1 group in Brazil (Figures 6 and **S7**), and cluster 9 may be ancestral of the P.1 variant.

**Table 3.** Worldwide sequences close to P.1 ancestors (PA-TP1) in 2020. The list was obtained using the distance from PA-TP1 to the 2020 world samples.

| epi | ID | date | variant |
| --- | --- | --- | --- |
| EPI_ISL_831339 | hCoV-19/USA/NC-UNC-0017/2020 | 2020-04-00 | B.1.1.1 |
| EPI_ISL_530145 | hCoV-19/USA/WA-S2788/2020 | 2020-08-12 | B.1.1 |
| EPI_ISL_530128 | hCoV-19/USA/WA-S2771/2020 | 2020-08-01 | B.1.1 |
| EPI_ISL_525755 | hCoV-19/USA/WA-S2765/2020 | 2020-08-03 | B.1.1 |
| EPI_ISL_954139 | hCoV-19/NorthMacedonia/29205/2020 | 2020-12-23 | B.1.1.428 |
| EPI_ISL_555709 | hCoV-19/England/ALDP-952525/2020 | 2020-06-09 | B.1.1 |
| EPI_ISL_1301549 | hCoV-19/Mexico/HID-InDRE-IBT-66/2020 | 2020-06-02 | B.1.1 |
| EPI_ISL_729470 | hCoV-19/Germany/SH-ChVir8194/2020 | 2020-07-19 | B.1.1 |
| EPI_ISL_700185 | hCoV-19/India/MH-ACTREC-539/2020 | 2020-08-25 | B.1.1.306 |
| EPI_ISL_745223 | hCoV-19/Russia/MOS-CRIE-7182855/2020 | 2020-08-24 | B.1.1 |

**Figure 6.**
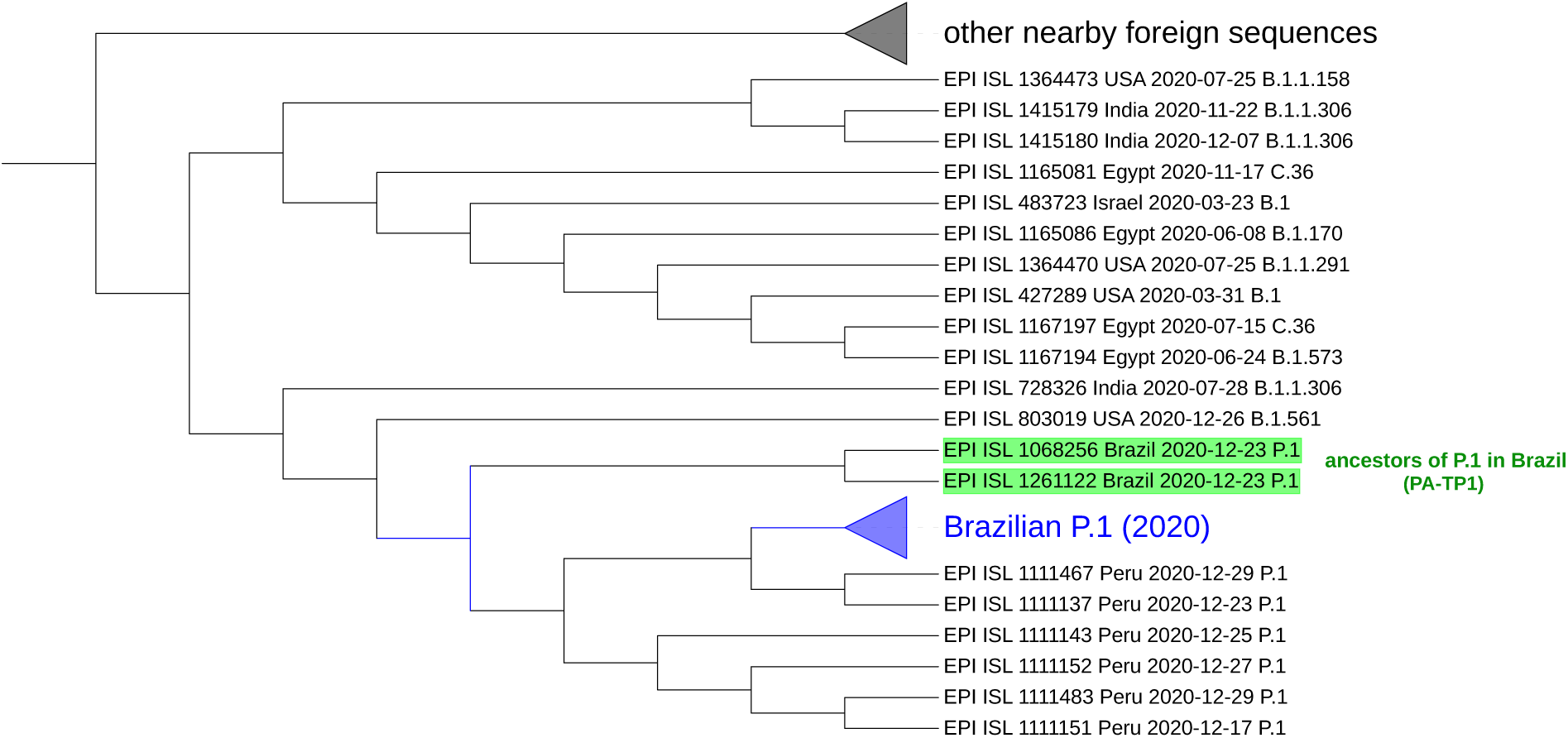
Sequences similar to P.1 detected by the neural network ensemble. The two identified ancestral sequences from the P.1 lineage in Brazil (PA-TP1) are highlighted green. The collapsed branch in blue groups 48 of the 50 Brazilian P.1 samples and is close to Peruvian. No instance of B.1.1.28 was identified in the group, nor other Brazilian sequences besides P.1. The collapsed branch above groups different foreign arrangements similar to P.1. The complete tree is available in Supplementary **7**. Graphic obtained with the iTOL tool – https://itol.embl.de/(35)

In the tree based on aligned genomes (Figure **S7**), we included the 50 Brazilian P.1 sequences from 2020, the 91 closest samples, including B.1.1.28 from cluster 6 and the B.1.1.28 sequences (EPI_ISL_1068137, EPI_ISL_801387, EPI_ISL_801397, EPI_ISL_801398, EPI_ISL_801389, EPI_ISL_801392, EPI_ISL_801394, EPI_ISL_801395, EPI_ISL_801399 EPI_ISL_801401) indicated by Naveca *et al*. (54) as belonging to the ancestral clade of the P.1 lineage. The P.1 group achieved a bootstrap of 100% in its branch (Figure **S7**) and is a sister group of a branch divided into a consistent branch of B.1.1.198 and another branch that includes samples of B.1.1 and B.1.1.192. The P.4 variant, corresponding to cluster 7, is a descendant of P.1 strain (cluster 3) since the first sample of P.4 (hCoV-19/Brazil/AM-CD1739/2020 – EPI_ISL_2233906) is a sister group of an Amazonian strain of P.1, indicating its probable place of origin, with BP=86%. The 100% bootstrap corroborates that PA-TP1 are ancestors of the P.1 lineage in Brazil. However, it was impossible to confirm the ancestor of the TP1 group since the BP was low in all other more basal branches, including the samples indicated by Naveca *et al*. (54) as the P.1 ancestral clade. This same phylogeny suggests that the Peruvian lineage of P.1 descended from the Brazilian P.1 lineage.

We measured the distances between proteomes from Brazilian samples and that of the Wuhan reference proteome over the pandemic period (Figure 1e). The differences between P.1 and P.4 and the Wuhan reference are much higher than the distance among the T0 group to Wuhan. From the beginning of 2021, the average distance among the Brazilian samples concerning the Wuhan sample leaped with the TP1 emergence as T0 group variants became extinct. Therefore, the distancing of the TP1 group from T0 is abrupt and not gradual.

The objective to construct the neural network ensemble was to search for P.1 like sequences in worldwide samples of 2020, before the emergence of P.1 in Brazil. After all results of the cross-validation over the complete set of Brazilian proteomes samples using the trained ensemble of neural networks for the P.1-true and P.1-false classes, we obtained: f1-score of 99.39%, accuracy of 99.5%, precision of 99.05%, and recall of 99.72%. These results confirm the separability of the P.1 samples from other Brazilian strains. The search in World-2020 data found 129 records of P1-like organisms, including the 50 Brazilian P.1 from 2020 and an additional 79 from other countries. This shows there were already in 2020 viruses like the Brazilian P.1 variant circulating the World before its emergence in Brazil. The phylogenetic analysis of these samples presented the same PA-TP1 samples, mentioned above with 10/17 mutations, as ancestral of P.1 variant (Figure 6). The proteomes closest to the origin of P.1, from those identified by the network, are one from USA (EPI_ISL_803019) labeled B.1.561, and one from India (EPI_ISL_728326) identified as B.1.1.306.

The proximity of the 48/50 P.1 sequences to a particular subgroup of B.1.1.28 samples in Brazil, above mentioned, led us to consider a possible recombination event involving B.1.1.28 and P.1 variants. Therefore, we provide a list with the 91 closest samples, the 50 Brazilian samples, and the ones suggested by Naveca *et al*. (54) for the recombinant event search tools RDP4 and RAPR.

The RDP4 found one possible recombinant event: hCoV-19/Lithuania/MR-LUHS-Eilnr352/2020 (EPI_ISL_636871 – B.1.1.280) as a recombinant sample, hCoV-19/Brazil/AM-CD1739/ 2020 (EPI_ISL_2233906 – P.4) as minor parental, and hCoV-19/England/OXON-AD15D/2020 (EPI_ISL_448567 – B.1.1.10) as major parental. This indicative comes with the observation that the recombinant may be a parent since the “minor parental” has not been precisely identified. The methods applied by RDP4, and their respective p-values are RDP (3.92E-04), GENECONV (6.26E-03), Bootscan (2.46E-03), Maxchi (1.35E-02), Chimaera (6.28E-03), 3Seq (1.36E-05).

RAPR results (Table **S9**) suggested the hypothesis that the proximity between P.1 to few samples of B.1.1.28 from cluster 6 may be due to a recombination event between a Brazilian P.1 and a foreign strain, close to hCoV-19/USA/NC-UNC-0017/2020 (EPI_ISL_831339 – B.1.1.1), which originated this group of B.1.1.28. Among the samples indicated as recombinant is the Brazil/AM-FIOCRUZ-20890261MV (EPI_ISL_801402 – B.1.1.28 – Table **S9**)). This sample belongs to clade 28-AM-II (A6613G) of B.1.1.28, indicated as the ancestor of lineage P.1 according to Naveca *et al*. (54) (Figure **S7**). Such clade has the A6613G mutation, a characteristic mutation of the TP1 group, present in 99.9% of the samples in the group. Therefore, to reinforce the recombination possibility, we built phylogenetic trees with the sequence before the alignment breakpoint and the other after this point. However, the trees did not reach a high enough bootstrap to confirm or rule out any recombination events suggested by the tools (Figure **S7**).

### Temporal and spatial distribution of lineages

The distribution of variants by state (Figure **S8**) shows that only 4 of the 27 states had samples continuously sequenced along with pandemics till May 2021: SP, RJ, RS, and BA. In other states, there were months without sequencing or simply one-off analyses. In the first stage of the pandemic, the B.1.1.28 and B.1.1.33 variants predominated until October 2020, when the P.2 variant became the predominant strain. Thus, from December 2020 until March 2021, the variant grew to become the primary variant in the country, followed by February 2021 by variant P.4 (Figure 4). Detailed information for the Brazilian States is available in Section **2.3** of Supplementary **1**.

The second epidemic wave of SARS-CoV-2 was more significant than the first, and its beginning coincides with the emergence and rise of P.1 (Figure 4c), as already reported (9; 54). See also Supplementary **4**. Over time, the lineages and clusters graphs illustrate how the T0 group prevalence decreased and was probably extinct (or occurred in small quantity), with variants TP1 and the imported groups, B.1.1.7 and the new variant of cluster 13, became dominants in Brazil (Figure 4). This transition is more evident when the evolution of the pandemics over time in the PCA and t-SNE graphs is viewed (3D graph of Figure **S9**, and the Supplementaries **3** and **4**). Furthermore, looking at the development of the lineages over time, we notice a pattern in the origin of new variants, characterized by the formation of new clusters (discussed later).

### Diversity of SARS-CoV-2 proteomes in Brazil

The study of SARS-CoV-2 diversity permitted both: i) understanding the distribution of the variants in the viral population in Brazil (richness), and ii) verifying and comparing the degree of sequencing in different Brazilian states and to compare with other countries (coverage). Here we exploited the concepts of richness and coverage as defined in Methods. São Paulo State (SP) presents the higher diversity (richness value), followed by RJ and RS with 10,221, 4,073, and 3,011 estimated distinct proteomes, respectively. The lower richness is in AC, with 50, followed by DF with 99 estimated proteomes (Table **S10**).

Regarding coverage, the states of SP, RO, and AM had the highest coverage (30.3%, 28.9%, and 26.9% respectively), while MA, AP, AL, MG, and SC had the lowest coverage (4.4%, 7.2%, 7.2%, 7.8%, and 8.9% respectively). Unfortunately, we could not analyze the states of RR, PI, RN, and MS due to low sampling.

The TP1 group presented almost half of the richness in Brazil, with 13,194 estimated distinct proteomes representing about 47.6% of the total local diversity in Brazil (Table **S11**). The TP1 also presented a high coverage (27.10% on average), which may be corresponding to the increase in sequencing in 2021; clusters 11 and 13 also had high coverages of 33.67% and 25.56%, respectively. T0 had lower coverages with 19.20% on average due to less sequencing at the beginning of 2020. We observed an increase in sampling between 2020 to 2021 and an increase in proteome quality (proteome complete and without misreadings). In 2020, 4,402 samples were sequenced, of which 52% (2,293 sequences) with quality, while in 2021, 8,993, of which 71% have good quality (6,427 instances).

Brazilian coverage (23.3%) is low compared to Italy, Germany, and England, but it is superior to India. However, the estimated richness of Brazil (26,370) is lower than the other countries in this study except for Italy (Table 4,Figure **S10**).

**Table 4.** Diversity comparison in selected countries. Richness and coverage metrics were calculated by Chao 1 method (28). Ncases = number of cumulative cases in the country (in millions); Nseq = number of quality collected sequences (analyzed); Nunique = number of non-redundant samples.

| Country | Ncases (millions) | Nseq | Nunique | Chao richness | Chao Coverage (%) |
| --- | --- | --- | --- | --- | --- |
| Brazil | 16.55 | 8,720 | 6,164 | 26,370 | 23.3 |
| India | 28.18 | 7,154 | 5,493 | 33,505 | 16.4 |
| Italy | 4.22 | 12,784 | 6,617 | 21,946 | 30.1 |
| Germany | 3.69 | 51,880 | 22,048 | 63,263 | 34.8 |
| England | 4.50 | 199,110 | 62,643 | 164,070 | 38.2 |
| World (until march 21) | 83.56 | 493,080 | 312,224 | 2,534,900 | 12.3 |

## Discussion

The pandemic of COVID-19 is a catastrophic event with severe consequences, leading to losses in almost all human activities, mainly health and the economy. On the other hand, we have a rare opportunity to observe the evolution process in almost real-time; since it promotes a rush for genomes sequencing of a single virus species never seen before. Recent bioinformatics technology provides resources to analyze the big data provided by these efforts and allows us to draw a panoramic view of the SARS-CoV-2 evolution in Brazil and worldwide.

The pandemic in Brazil had two moments (Figure 4):

1. T0 - the early entry of SARS-CoV-2, which occurred in early 2020, characterizing the T0 group in this study. The T0 group embodied many lineages that disappeared over time, and the prevailing lineage were B.1.1.28 and B.1.1.33, and later the P.2.
2. TP1 - detected between Dec 2020 and Feb 2021, is characterized by groups related to the P.1 (Gamma) variant and other late imported sequences, including B.1.1.7 and P.4 of cluster 13.

We observed that strains tend to be extinct and replaced by newer and more adapted strains holding more advantageous mutations, as observed in other studies (20; 54). This lineage substitution process was followed in Brazil on several occasions, as in the emergence of the P.2 variant and later of the TP1 group (Figure 4).

Our proposed pipeline^7^ (Figure **S1**) allowed us to recognize the appearance of new variants. New variants emerge by moving away from the parental lineage, in a process called “exploitation of the mutational space”, which becomes graphically visible by the methods of PCA and t-SNE (Figure **S4**), followed by the establishment of a new cluster. We observed a remarkable variant emergence event during the analysis, the origin of cluster 7, a sublineage of P.1 (cluster 10) composed of 126 samples from the State of São Paulo (called P.1-SP at first moment – Figure **S4**). These samples had their designation updated from P.1 to P.4 between v2.3.8 and v3.0.5 of PANGO.

There is no reliable phylogenetic analysis of SARS-CoV-2 in the literature. Furthermore, the high mutation rate associated with the large volume of circulating viruses strains in the world entails frequent cases of parallel and backward mutations, resulting in inconsistencies in the determination of lineages and hindering the reconstruction of their evolutionary relationships (20), difficulties also reported in the survey by Morel *et al*. (55). Therefore, we performed a phylogenetic analysis of complete SARS-CoV-2 genomes/proteomes based on vectorial distances matrices. We compared trees based on representing lineage centroids with those based on representing cluster centroids. The cluster centroid-based phylogenetic trees showed to be consistent (bootstrap greater than 85% in all branches), while the lineage centroid-based tree presented a much lower BP and did not show clear differentiation in the evolutionary history of the lineages. It led us to conclude that the division of specimens by clustering is more reliable to the evolutionary mapping and that some inconsistencies may be present in SARS-CoV-2 classification by PANGO. Therefore, the clustering approach presented in this study may help revise the lineages nomenclature process. Additionally, the use of proteomes (amino acid representation) in the evolutionary analyses and the heatmaps (Figures 1 and 5) showed more consistency than use of genomes (DNA) in both cluster and lineage divisions. As a consensus across all methods, the TP1 group consistently clusters in a single branch, away from the other Brazilian variants.

Based on the results, we suggest three plausible hypotheses for P.1 variant origin: (a) the origin from variant B.1.1.28 in Brazil, as reported by Naveca *et al*. (54), (b) a foreign origin from a late entry strain, and (c) P.1 was originated by some recombinant event.

The phylogeny in Figure **S7** does not support the lineage B.1.1.28 as ancestor of P.1. We cannot, however, conclusively rule out the possibility of a Brazilian origin for P.1 since there is a gap in the sampling in the period of the emergence of P.1 in Brazil around October 2020. However, the accumulating body of evidence consistently points to an external P.1 origin:

1. The considerable distance (Euclidean and phylogenetic) and different clustering between samples of P.1 and the previously reported ancestor B.1.1.28 (Figures 1a-d, 2, 5, **S6** and **S7**);
2. Foreign sequences are closer to PA-TP1 than any Brazilian samples of the T0 group (Table 3);
3. The distance from the Wuhan reference sample is much higher to P.1 than to the other Brazilian instances in 2020 (Figure 1e);
4. There are many accumulated mutations in P.1 without intermediate sequences detected in Brazil (Figures 3 and **S5**);
5. The machine learning approach found P.1-like SARS-CoV-2 samples circulating the World before the variant emergence in Brazil (Figure 6).

The external VOC P.1 entry in Brazil may have been favored by the flexibilization of measures regarding international flights in Brazil in October 2020 (48), the period of entry/emergence of P.1 in Brazil, also suggested by Faria *et al*. (2). After the entry of P.1 in Brazil, the mutations S:H655Y, S:T1027I, S:R190S, S:T20N, ORF1a:S1188L, ORF8:E92K and ORF1b:E1264D probably originated in Brazil, since they are not present in the ancestral PA-TP1, assuming these samples as reference. Among these mutations, the S:H655Y promotes escape from the immune system (51).

Recombination is a common phenomenon in the Coronaviridae family (47); however, there are indications that recombinant events between SARS-CoV-2 strains are rarer than expected (56). Our results indicate no recombination event in the origin of the P.1 variant; however, such an event can relate to B.1.1.28 and P.1 variants. The RAPR tool results indicate that a subgroup of B.1.1.28, a subset of cluster 6 in our study, the same group identified as 28-AM-II (A6613G) clade by Naveca *et al*. (54), was originated by recombination between a P.1 and a foreign sample close to the hCoV-19/USA/NC-UNC-0017 (EPI_ISL_831339 – B.1.1.1) (Table **S9**). Thus, our analysis points to the possibility that clade 28-AM-II comes from recombination, but it is not an ancestor of P.1.

We propose Cluster 9 as the probable ancestral cluster of the TP1 group (Figure 5). It contains the PA-TP1 samples, the sequenced strains closest to the ancestors of the P.1 lineage. Furthermore, the hypothesis is supported by the PCA (Figure 2a), which shows cluster 9 as the furthest apart among the TP1 clusters, dispersed as in the described “exploitation of the mutational space” during the origin of new variants, forming a bridge between itself and the other TP1s.

The diversity analysis revealed that coverage of viral subvariants is low in all Brazilian states (Table **S10**), and 13 of the 27 Brazilian states had less than 100 quality samples until May 2021. São Paulo (SP) and Rio de Janeiro (RJ) States present more sequencing and had 4,386 and 1,170 sequenced samples, respectively. The state of SP is the national center of the pandemic, having the highest virus richness. In addition, SP is the the main hub for national and international travel, representing more than 70% of international flights from/to Brazil (7). Therefore, it was expected a large circulation of different viral variants in this state. Candido *et al*. (7) indicate that, like SP, the states MG, CE, and RJ are major international travel entry centers. For these states, the estimated richness also presented high values (Table **S10**), except CE, that appear to have underestimated richness, likely due to the low sampling.

Wealthier countries, such as those in Europe, also presented the highest richness estimations despite the lower number of COVID-19 cases (Table 4). Thus, we hypothesize that European countries, receiving more international inflows, have a higher chance of variants entering, increasing their viral richness, such as in the State of São Paulo in Brazil.

The disproportion in sampling between states (Table **S12**) makes it difficult to compare the evolutionary history of the virus among states. In addition, the low coverage in regions may hide VOCs, making their tracking hard (9). Also, globally, there is a concentration of sequencing. Ten countries account for 85% of the GISAID samples and only 35% of the World’s cases. Disproportion in sampling between different countries results in strains remaining undetectable until they become widely spread, and then it is no longer possible to effectively control their dispersion (20). We think that, similarly, the subsampling in Brazilian states corroborated the sudden spreading of the P.1 lineage.

### Comments on PANGO and GISAID database updates

In the current version of PANGO in v3.1.11, all P.4 samples from all clusters have been reclassified as P.1 (excepting those of cluster 13). It came in accord with our findings and explains the overlapping observed in Figure 2c concerning the clusters of the TP1 group, based on the previous terminology (Figures 3, 1a-d). Also, concerning the T0 group, cluster 5 has 77 samples of B.1.1.28 consistently separated from the others, which had its designation updated to variant P.7, which agrees with our analysis. More about updates in PANGO in Table **S13**.

## Conclusion

Here we present a data-science-based overview of the SARS-CoV-2 pandemics in Brazil. Brazil has a scenario divided into two moments. The first, which occurs throughout 2020 to early 2021, is composed of several variants from the initial entry of SARS-CoV-2 strains in Brazil (Figure 4). The primary early entry samples are the B.1.1.1, B.1.1.28, and B.1.1.33 variants, and later occurs the emergence of P.2 and the recently named P.7. The second moment sees the emerging of P.1 (and correlated variants) and the entry of foreign strains – late 2020 and early 2021 – a period when preventive measures were relaxed (48).

The classification performed by PANGO generally corresponds to our analyses.Most of the recent updates of the PANGO nomenclature (v3.1.11) are supported by our findings; however, constant changes impair the comparability of studies. Our analyses showed that the clustering method groups the sequences by evolutionary similarity, making it suitable to classify tasks even for nomenclature purposes. Additionally, the results of proteomic evolutionary analyses showed to be more consistent than genomic ones and thus ideal for this analysis. Therefore, the proposed pipeline is based on proteomic sequences.

The process of emergence and extinction of new lineages occurs continuously and gradually (54). New strains arise through a process of “genomic exploration”, distancing from the parents and consolidating (Figure **S4**). This process is visible graphically by t-SNE and PCA on the vectorized and clustered sequences, methods that detect emerging variants as demonstrated in the notable case of cluster 7.

The considerable distance between the sequences of the TP1 group and the other Brazilian sequences led us to question the Brazilian origin of the lineage P.1. We could not completely discard the Brazilian source from group T0. Still, we have assembled a set of evidence that suggests its external origin (Figures 3, 5). The recombination analysis did not detect a recombination event in the root of P.1; however, it shows a possible recombination event between a Brazilian P.1 specimen and a B.1.1.1 from the USA, generating a subgroup of B.1.1.28, interpreted in previous studies as the ancestral clade of P.1 (54).

A considerable increase in sequencing has occurred in 2021 in Brazil; however, the disproportion between states remains. Disproportionate monitoring, both nationally and internationally (20), allows dangerous variants to remain undetectable until they become widespread. In addition, the richness estimations point out the existence of a much larger number of variants that are not yet sequenced (Table **S10**, Figure **S12**). The analyses indicate that Chao’s metric combined with vector representation for proteomes is a suitable method for viral diversity analysis (28).

Worldwide sequencing is scarce, around 12% (Table 4). We have seen that the lack of monitoring by sequencing in Brazil has allowed P.1 to spread silently; moreover, we could not trace the origin of its large number of accumulated mutations (17 in all), mutations which make this VOC dangerous. The low sequencing associated with a great richness of variants, observed in countries like India, may lead to the emergence of new VOCs, such as the Indian B.1.617.2 (Delta). Therefore, sequencing should increase, and border control measures help control the spread of dangerous variants.

The variant Delta is replacing the P.1 lineage in Brazil (Figure **S11**) (24). Based on this study, we suspect that the emergence and domination of Delta in the World follow an analogous way that P.1 dominated in Brazil. Moreover, due to some countries’ low sampling and vaccination, new VOC can still emerge and even replace Delta itself. We hope our study can bring new lights to better understanding such viral evolving behavior, now and in other eventual future outbreaks.

## Supporting information

Supplementary2_gisaid_hcov-19_acknowledgement_table

Supplementary3_tSNE_by_week

Supplementary4_PCA_by_week

Supplementary5_6_and_7_phylogenetics_trees

## Data Availability

All data produced are contained in the manuscript or available online at https://github.com/CamilaPPerico/SARS-CoV-2_Brazil_Landscape/tree/main

## Acknowledgments

The authors are deeply grateful to all the researchers and organizations collaborating to maintain and share SARS-CoV-2 genomic data on the GISAID Platform (See Supplementary **2**). The authors thank the group of Artificial Intelligence Applied to Bioinformatics of Federal University of Paraná and CAPES (Coordination for the Improvement of Higher Education Personnel) for the financial support.

## Author contributions

R.T.R. and C.P.P. designed and implemented the analysis. G.P.N. contributed to the search and analysis. D.R.F. coded the R version of SWeeP. C.P.P. wrote the original draft of the manuscript. C.R.D.P, F.O.P., and E.M.S. made substantial contributions, revisions and approved the final manuscript. R.T.R. supervised the whole project. All authors contributed thoughts and advice, discussed the results, and contributed to writing the final manuscript.

## Data availability

Genomes and proteomes were obtained at Global Initiative on Sharing Avian Influenza Data (GISAID), available at https://gisaid.org/. Complete information about the Brazilian, Indian, German, Italian, British, and World-2020 sequences analyzed has been deposited at https://github.com/CamilaPPerico/SARS-CoV-2_Brazil_Landscape/tree/main/metadata. The Wuhan reference sequence (NC 045512.2) was obtained from the National Center for Biotechnology Information (NCBI) https://www.ncbi.nlm.nih.gov/nuccore/1798174254. Information regarding the number of COVID-19 cases in Brazil was collected from official website https://covid.saude.gov.br/. Further data and materials produced in this study are available in the supplementary information (the contents of each file are described in Supplementary **1**) and at https://github.com/CamilaPPerico/rSWeeP_case_study

## Code availability

The pipeline developed in this study, coded in R language, is available at https://github.com/CamilaPPerico/SARS-CoV-2_Brazil_Landscape/tree/main/scripts Code and instructions for reproducing our main results are described in Supplementary **1**. The R version of the SWeeP tool is deposited at Bioconductor platform, available for version R 3.12 (25), at https://bioconductor.org/packages/release/bioc/html/rSWeeP.html.

Previous versions of PANGOLIN are available at https://github.com/cov-lineages/pangolin/releases (22). The web platform Nextstrain, used for genome alignment, can be accessed at https://clades.nextstrain.org/ (24). For the recombination analysis, the RDP4 software is available for download at http://web.cbio.uct.ac.za/darren/rdp.html (38), and the RAPR web tool is available at https://www.hiv.lanl.gov/content/sequence/RAP2017/rap.html (37). The MEGAX 10.2.6 tool, used for the phylogenetic analysis genome-based, can be downloaded from https://www.megasoftware.net/ (33). Finally, for visualization of phylogenetic trees, the iTOL online tool can be accessed at https://itol.embl.de/ (35).

## Footnotes

1 PANGO lineages – https://cov-lineages.org/

2 Wuhan reference sequence NC_045512.2 – https://www.ncbi.nlm.nih.gov/nuccore/1798174254

3 Nextclade – https://clades.nextstrain.org/

4 rSWeeP Bioconductor – https://bioconductor.org/packages/release/bioc/html/rSWeeP.html

5 Rtsne package – https://github.com/jkrijthe/Rtsne

6 iTOL tool – https://itol.embl.de/

7 Pipeline available at:https://github.com/CamilaPPerico/SARS-CoV-2_Brazil_Landscape/

