## Supplementary1_Figures_and_Tables for "Genomic landscape of SARS-CoV-2 pandemic in Brazil suggests an external P.1 variant origin"

November 10, 2021

1. Laboratory of Artificial Intelligence Applied to Bioinformatics - SEPT, Federal University of Paraná, Curtiba, Paraná, Brazil
2. Graduate Program in Bioinformatics - SEPT, Federal University of Paraná, Curtiba, Paraná, Brazil
3. Department of Biochemistry and Molecular Biology, Federal University of Paraná, Curtiba, Paraná, Brazil

**Corresponding Author:**
Roberto T. Raittz:

**This PDF file includes:**

- Description of Supplementary 2-7 (section 1) and other Supplementary material available at https://github.com/CamilaPPerico/SARS-CoV-2_Brazil_Landscape/
- Results in detail (section 2)
- Supplementary Figures and Tables (section 3)
- References

### 1 Supplementary Description

**Supplementary 2:** (Supplementary2_gisaid_hcov-19_acknowledgement_table.pdf)
Acknowledgements to all researchers and professionals who contributed to the GISAID genome database and for this study.

**Supplementary 3:** (Supplementary3_tSNE_by_week.pdf)
Lineage evolution over time through visualization with t-SNE graph. Each slide adds the samples of a new week. The colors represent each cluster obtained for Brazilian sequences. Here was adopted the nomenclature PANGO v3.0.5. The key shows the main lineages that each cluster represents. The emergence, growth and extinction of the groups is visible, as well as the gradual replacement of the T0 group variants by the TP1 group.

**Supplementary 4:** (Supplementary4_PCA_by_week.pdf)
Lineage evolution over time through visualization with PCA graph. Each slide adds the samples of a new week. The colors represent each cluster obtained for Brazilian sequences. Here was adopted the nomenclature PANGO v3.0.5. The key shows the main lineages that each cluster represents. The emergence, growth and extinction of the groups is visible, as well as the gradual replacement of the T0 group variants by the TP1 group. Note that the second wave was bigger and started with the emergence of P.1 lineage. The information on the number of COVID-19 cases in Brazil was obtained at the official website `https://covid.saude.gov.br/`.

**Supplementary 5:** (Supplementary5_BRtree8kProt_160k.nwk)
Complete phylogenetic tree of 8720 Brazilian samples from GISAID release 609 of 2021-06-06 generated by Neighbor-Joining method from the vectorized proteomes (PANGO nomenclature v3.0.5 of 2021-06-04)

**Supplementary 6:** (Supplementary6_BRtree8kDNA_65k.nwk)
Complete phylogenetic tree of 8720 Brazilian samples from GISAID release 609 of 2021-06-06 generated by Neighbor-Joining method from the vectorized genomes (PANGO nomenclature v3.0.5 of 2021-06-04)

**Supplementary 7:** (Supplementary7_tree_near_P1_world_sequences_from_ANN.nwk)
Phylogenetic Tree of sequences similar to P.1 obtained through the neural network ensemble. The probable ancestral samples PA-TP1 (EPI ISL 1068256 and EPI ISL 1261122) are highlighted in green in the Figure S7. The remaining 48 samples of Brazilian P.1 are close to the Peruvian samples. No sample of B.1.1.28 was identified in the group, nor other Brazilian sequences besides P.1. The branch above groups other foreign sequences similar to P.1. The 'No' in labels indicates that they are not of the P.1 lineage.

**Additional data:** Supplementary material available at `https://github.com/CamilaPPerico/SARS-CoV-2_Brazil_Landscape/`:

- The Pipeline is divided into 9 codes, in order of use, in the "scripts" folder. In all steps, parameters and the name of the input/output files can be edited.

  **script 1** Extraction of the proteomes of interest from GISAID's fasta file.
  The input sequence for this step is the file `allprot(release).fasta`, available at GISAID (`https://gisaid.org`). This file contains the proteome of each world sample of SARS-CoV-2. The input for all the following steps are produced by the scripts themselves. In this step, are extracted only the sequences of the country of interest.

**script 2** Concatenation of sequences in multifasta file.
Concatenates each of the proteins of the country of interest and creates a unique complete proteome sequence (multifasta output). In this step, a file listing the headers and a file with the EPI codes are provided.

**script 3** Selection of quality sequences.
From the multifasta, are selected only sequences without misreadings (presence of X) and with complete proteomes (with 26 or more proteins). In this step is create only the index list of the quality sequences.

**script 4** Filtering quality sequences.
The quality sequences index list is used to create a second multifasta, EPI and headers lists only with filtered sequences.

**script 5** SWeeP projection of sequences in vectors.
The proteomes from filtered multifasta are vectorized by the SWeeP tool. A matrix is obtained in which each row corresponds to a sample. Each sample vector has a length of 600 dimension.

**script 6** Consensus Clustering step.
The vectorized sequences are grouping by similarity using the clustering method. Parameters of resampling number, maximum number of clusters, clustering algorithm, among others, can be adjusted as needed.

**script 7** Visualization of clusters step.
The visualization of the sequences by PCA and t-SNE is performed using the vectorized data (SWeeP) based on the division by clusters (Consensus Clustering).

**script 8** Consensus Phylogenetic tree step.
Obtaining of the consensus phylogenetic tree, with bootstrap values, by the Neighbor-Joining method.

**script 9** Consensus Phylogenetic tree visualization step.
The visualization of the consensus phylogenetic tree, obtained in step 8. The division by clusters (Consensus Clustering) is highlighted by the colors of the branches. The bootstrap values are shown at the base of each branch.

- A sample of GISAID release 609 to test the scripts: Sample_GISAID_Release0609.fasta
- The metadata of the analyzed sequences of Brazil, Germany, India, Italy, UK and World-2020 are available in the "metadata" folder: Metadata_Brazil.csv, Metadata_Germany.csv, Metadata_India.csv, Metadata_Italy.csv, Metadata_UK.csv (in 7 parts), and Metadata_World_2020.csv.
- Consensus tree of vectorized proteomes, with bootstrap of 1'000 replicas, 8'720 samples, Neighbor-Joining method: SARS_NJ_Consensus_BP.nwk

#### 2 Detailed results

We obtained 1'000'558 samples worldwide in release 409 on the GISAID platform (1), and 49% of proteomes remained after filtering (493'080 sequences, of these 260'759 from 2020). Corresponding to the release of 609, Brazil had 65% of quality sequences, a percentage higher than the world average and other countries studied (India with 38%, Italy with 39%, Germany with 41% and England with 56% of the sequences with quality) . The rSWeeP projection process ran in about 0.08s per sequence. For the 1 million sequences worldwide it took 22h using 21Gb of RAM, as shown in Table **1** (paper).

#### 2.1 Landscape in Brazil

Based on the established consensus criteria (Figure S2), we identified 15 clusters representing the epidemic in Brazil from 25 February 2020 to the end of May 2021. All 15 clusters obtained around 80% consensus of each point in its respective cluster, for all sequences. More than 15 clusters do not provide considerable increase (less than 5%) in the consensus value of the CDF curve.

According to the PANGO v3.0.5 nomenclature of 2021-06-04, 11 main lineages were identified in Brazil: P.1(3572 – 40.9%), P.4(1274 – 14.6%), P.2(1132 – 13.0%), B.1.1.33(909 – 10.4%), B.1.1.28(864 – 9.9%), B.1.1.7(248 – 2.8%), B.1.1(186 – 2.1%), P.1.2(153 – 1.7%), N.9(81 – 0.9%), B.1(65 – 0.7%), B.1.195(54 – 0.6%), other (178 – 2.0%).

As mentioned in the main text, the clustering results match with the division of the observed groups by t-SNE, PCA, distance matrix and PANGO nomenclature (Figures **1** and **2**), except for particular cases (Table S1).

#### 2.2 Mutation profile of the groups

We found the most common mutations in Brazil, present in almost all 8720 samples, and listed them in Table S5. We determined the characteristic mutations for the TP1 group in Table S6, and for the other clusters in Table S7. Mutations are visually presented in the heatmap of Figure **3**.

The spike mutation D614G is a globally dominant allele (2), present in 98.9% of the samples already collected in Brazil. It is already established that such mutation generates an increase in infectivity due to a large conformational change of the spike protein and due a higher viral load in relation to the ancestral (3; 4).

As previously verified, mutations in the Spike protein have a strong impact on the definition of lineages and variants of SARS-CoV-2, being responsible for much of the division between the clusters. Mutations and indels that occur in the Spike protein change the protein's conformation, affecting its stability and functionality, as well as its ability to infect the host (5).

Non-characteristic samples from each group were analyzed in greater detail. The Table S1 presents samples that have probable lineage annotation error by PANGO (v3.0.5) or possible unidentified recombination events, as already reported (6).

##### 2.2.1 cluster 11 - variant B.1.1.7

B.1.1.7 of cluster 11 has 3 deletions and 1 mutation shared with TP1 group (ORF1a:G3676–, S3675–, F3677– and S:N501Y), 4 mutations that are consensus in Brazil (ORF1b:P314L, S:D614G, N:R203K and N:G204R), and 13 mutations and 3 unique deletions from the group (ORF8:Y73C, N:S235F, S:A570D, S:S982A, S:D1118H, S:T716I , ORF1a:T1001I, N:D3L, ORF8:R52I, ORF1a:A1708D, ORF1a:I2230T, ORF8:Q27*, S:P681H, S:H69–, S:V70–, S:Y144–). These mutations correspond to those found in the literature (7).

##### 2.2.2 cluster 13 - variant P.4

Cluster 13 has 5 mutations that are consensus in Brazil (ORF1b:P314L, S:D614G, N:R203K, N:G204R, S:V1176F) and has 15 unique mutations (ORF1a:L3915F, ORF1b:Y822C, ORF3a:V50I, ORF1a:A516T, ORF1a:L3201P, ORF1a:A3143V, S:L452R, N:A220V, S:I720V, S:Q173K, ORF1a:Q3729K, S:N164K, S:S704L, S:G142V, ORF1a:P971L). As detailed in Table S7, the S:L452R mutation makes the virus 20% more transmissible, increases its infectivity (*in vitro*), and reduces antibody neutralization (8), giving low vaccine efficiency (9). This mutation occurs in more than 103'000 samples worldwide, but it is rare in Brazil.

The N:A220V mutation alters the conformation of the protein and potentially causes changes in pathogenicity and/or response to vaccination, affecting the activity of the protein's binding site. It is a growing mutation, with 163'000 occurrences in the world, one of the top 10 most frequent in the world, but rare in Brazil (10; 11). Additionally, the S:Q173K mutation has the potential to increase the fitness of the (12) virus, but there is still no concrete evidence.

##### 2.2.3 Early group (T0) - clusters 1,2,4,5,6 e 8

The group does not have consensus mutations characteristic of the group, as they are not a coherent group like the TP1 group. Each cluster represents an individual lineage, or a group of less frequent lineages in Brazil. The mutations shared by this group are the Brazilian consensus mutations (e.g. ORF1b:P314L, S:D614G), as detailed in Table S5.

##### 2.2.4 Groups related to variant P.1 (TP1) - clusters 3,7,9,10,12,14 e 15

The TP1 group are divided into 7 clusters: the oldest are cluster 3 (composed of sequences from P.1) and cluster 7 (composed of P.4), the only clusters with samples in 2020. In total, there are two clusters of variant P.1 (clusters 3 and 14), two of variant P.4 (clusters 7 and 15), two containing variants P.1 and P.4 (clusters 9 and 10), and one composed of the P.1.2 variant (cluster 12).

These clusters have a particular set of non-synonymous mutations shared by the entire group, in addition to the deletions of nucleotides 11288-11296, and the insertion 28263:AACA (Table S6). Most of these consensus mutations are located in the spike protein and confer an adaptive advantage: mutations H655Y and L18F promote immune response escape (13; 14); mutations E484K and N501Y strengthen the binding with hACE2, providing high transmissibility and low vaccine efficiency (9); the association K417T, E484K and N501Y promotes high affinity with ACE2 (40-50% greater than wild type) and resistance to antibodies (15). It is believed that some mutations such as E484K and N501Y in the spike protein were positively selected for conferring such adaptive advantages (16).

#### 2.3 Temporal and spatial distribution of strains detailing

About the distribution of variants by state (Figure S8), in more detail, we observed:

- No conclusions can be drawn from the distribution of variants in the following states: Acre (Ac), Ceará (CE), Distrito Federal (DF), Mato Grosso (MT), Piauí (PI), Rondônia (RO), Roraima (RR), Tocantins (TO).
- The states of Mato Grosso do Sul (MS), Pará (PA), Rio de Janeiro (RJ), Paraná (PR), Rio Grande do Norte (RN) and Rio Grande do Sul (RS) behaved as the Brazilian average described .
- The states of Alagoas (AL), Amapá (AP), Maranhão (MA), Minas Gerais (MG) and Santa Catarina (SC) do not provide enough data to infer about the variants in the state in 2020. In late 2020 (November and December) and early 2021 there was a predominance of variant P.2 which was gradually replaced by P.1. Additionally, in MA there was a predominance of N.9 in December 2020; and in MG there was a growth of the P.4 variant from March 2021 onwards..
- Amazonas (AM): until april/2020 the B.1.195 variant predominated. Between May and August/2020 there were variants B.1.195, B.1.1.28 and B.1.1.33 representatively. The B.1.1.378 variant was present from the beginning in a smaller amount until the end of 2020. From December 2020 the P.1 variant became predominant in the state.
- Bahia (BA): In November the N.9 became significant but its frequency dropped until February/2021, when the P.2 variant became representative.
- Espírito Santo (ES): as of February/2021, variants B.1.1.7 and P.1 become predominant.
- Goiás (GO) and São Paulo (SP): behave as the Brazilian average described, additionally, from February 2021, P.4 and P.1.2 are growing.
- Paraíba (PB): The year 2020 is mainly composed of variants B.1.1 and B.1.1.33.

- Pernambuco (PE): the beginning of 2020, between April and July 2020, the most representative variant was B.1.1.

- Sergipe (SE): in the first months of 2020 (until April 2020) there was a varied distribution of variants (B.1, B.1.1, B.1.1.28, B.1.1.33 and others). In early 2021 there was also a diversity of variants which was quickly replaced by P.1 from February 2021 onwards. In the other months there was no sampling.

### 3 Supplementary Figures and Tables

**Figure S1:** Pipeline. The method starts by concatenating viral proteome sequences (with border delimiters) and filtering out the quality sequences (removing incomplete proteomes and sequences with misreading). Then, the sequences are projected in numerical vectors by the rSWeeP tool resulting in a numerical matrix that is used for the following analyses: phylogenetic tree construction, clustering, PCA, t-SNE, and other analyses.

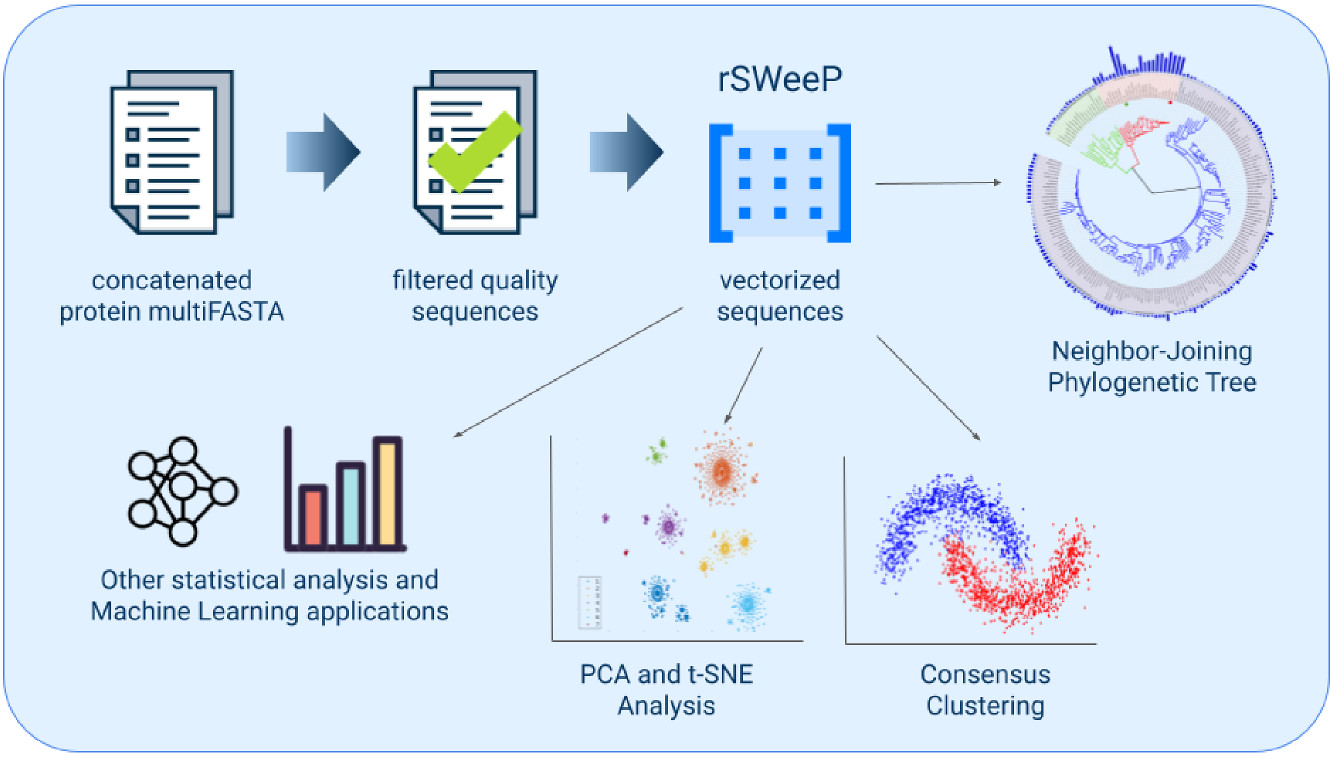

**Figure S2:** Consensus Cumulative Distribution Function. It is observed that above 15 clusters there is no appreciable increase in the value of the consensus between the clusters (less than 5%). With 15 clusters, a value of about 80% of consensus is obtained for all samples in their respective clusters.

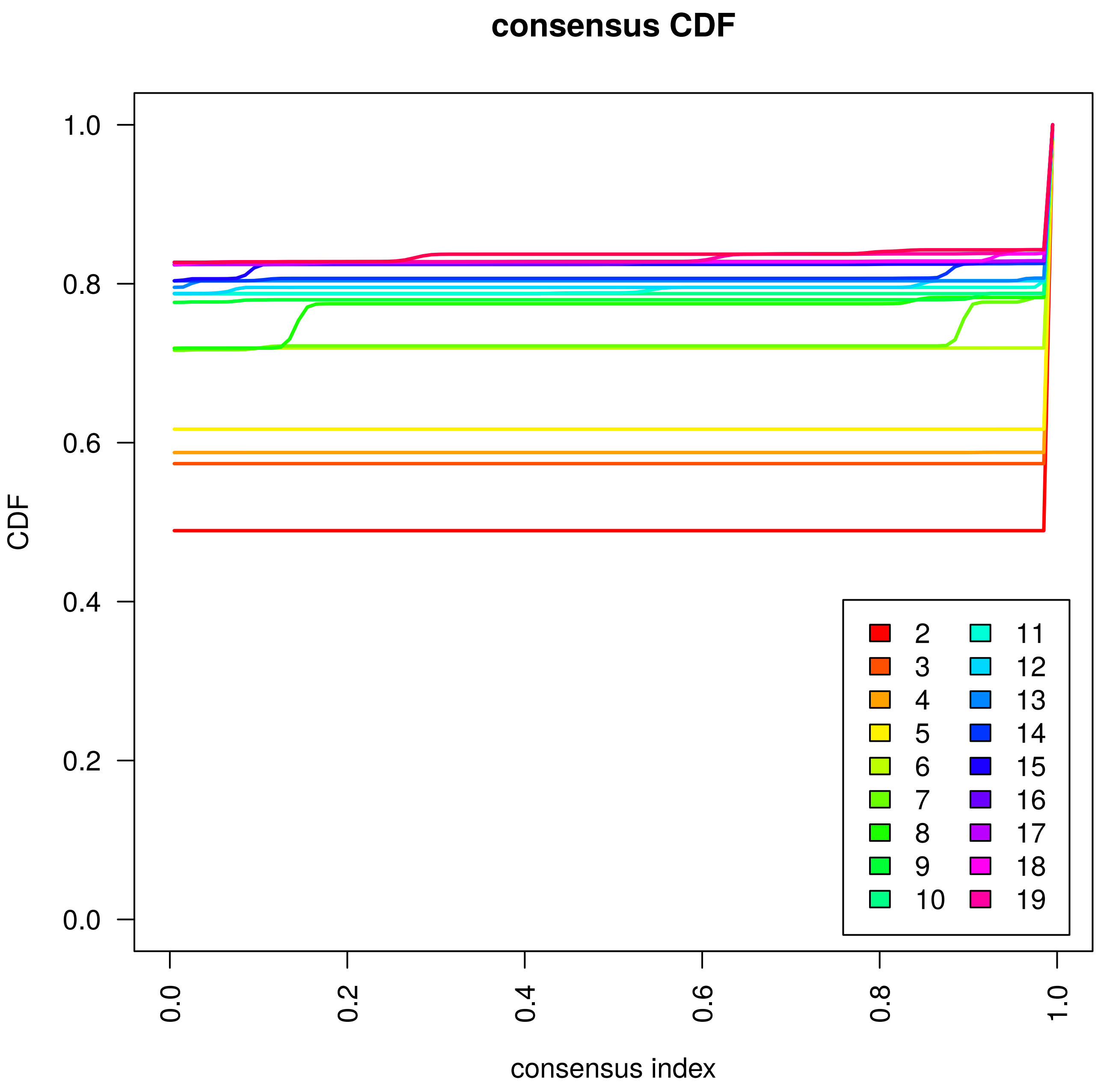

**Figure S3:** Clustering of vectorized spike proteins. We obtained 8 well-defined clusters, however there is a greater degree of mixing of the strains within the clusters due to the lower resolution provided by the protein in relation to the complete proteome. **a)** t-SNE and **b)** PCA of spike sequences. Divides the samples similarly to the complete proteome: TP1 group on the left, B.1.1.7 below on the right, and the remaining sequences on the right.

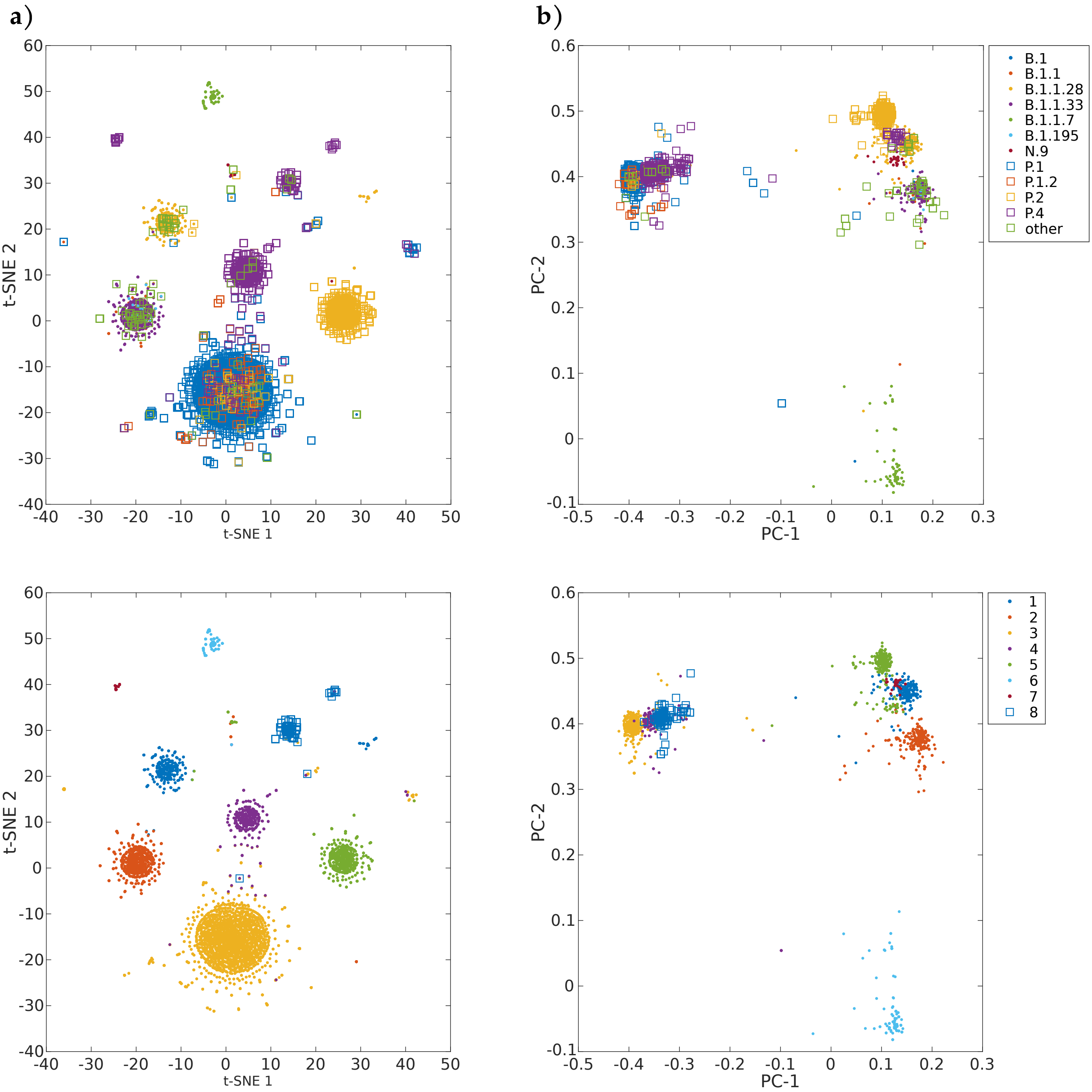

**Figure S4:** Detection of the emerging variant P.1-SP. A subset of variant P.1 (data from GISAID release 409 using PANGO v.2.3.8) was later identified as new variant P.4 (GISAID release 609 using PANGO v.3.0.5). **a)** t-SNE with data from GISAID release 409, until 2021-03-19. Clusters correspond to: (1) P.1; (2) B.1.1.33; (3) B.1.1/B.1.1.28; (4)B.1.1.28; (5)B.1/B.1.195; (6) P.2; (7) B.1.1.7 (PANGO nomenclature v2.3.8 of 2021-04-02). Circled in red is P.1-SP group, emerging variant group P.4, according to PANGO v.3.0.5. **b)** t-SNE with data from GISAID release 609, until 2021-05-29. Highlighted in red triangles the emerging variant P.4, as circled in **a)**.

**a)**

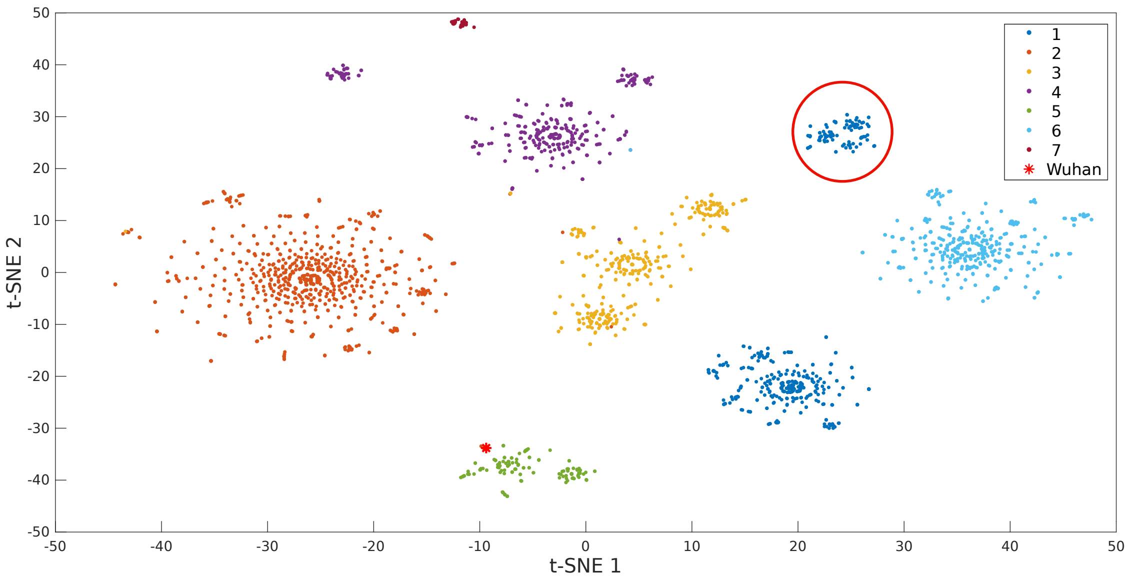


**b)**

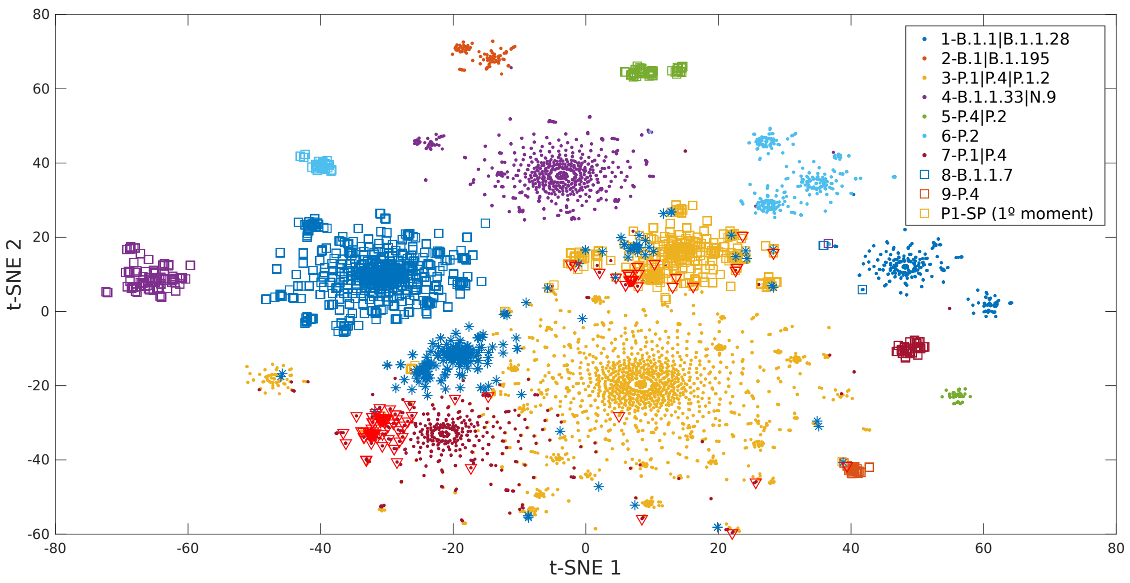

**Figure S5:** Heatmap of mutations by lineage (relative to PANGO v3.0.5). Mutations present in 75% samples from one or more lineages are listed. The value 1 (red) represents the presence of the mutation in 100% of the lineage samples, and the value 0 (blue) indicates the absence. Values are normalized by lineage.

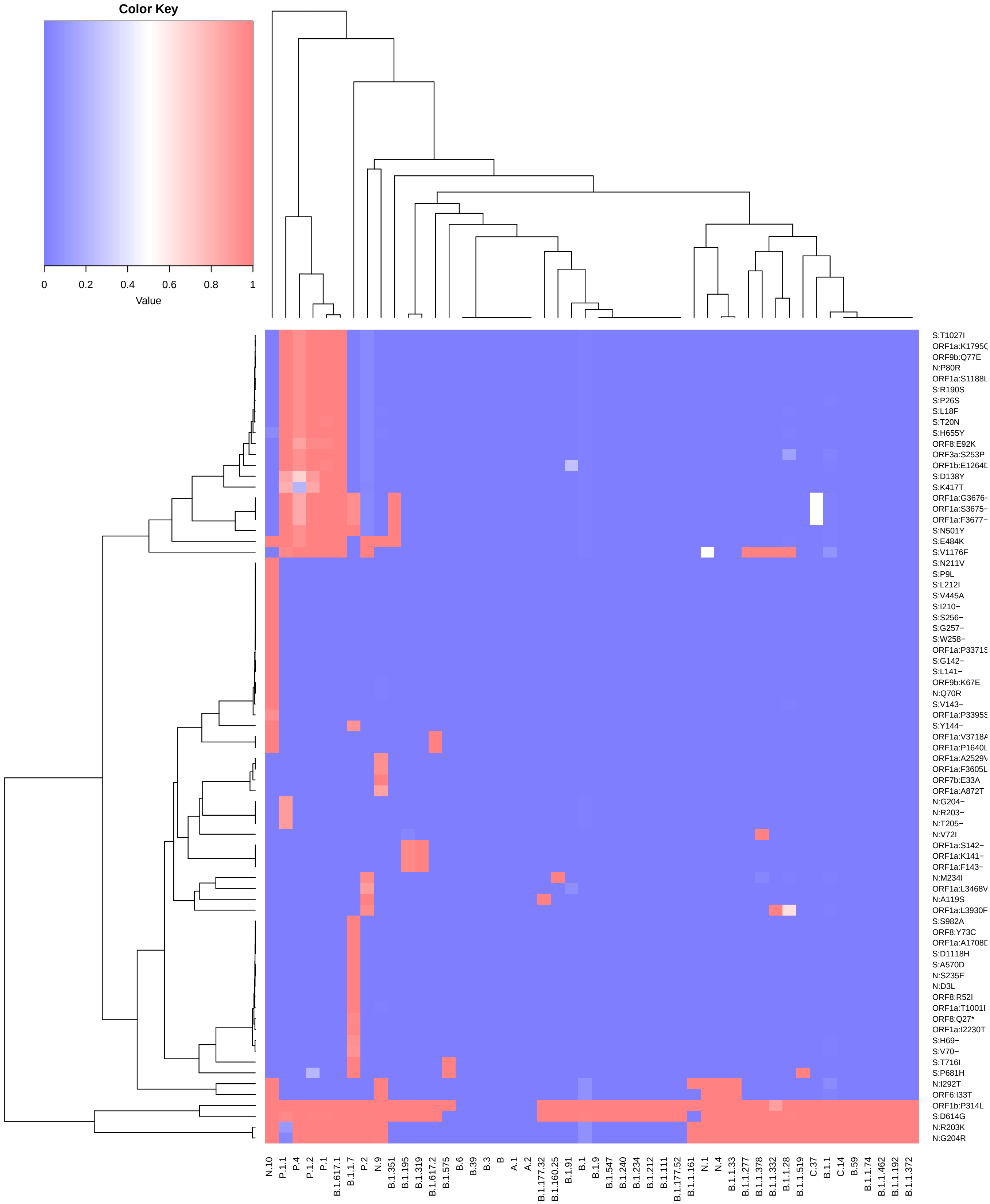

**Figure S6:** One of the possible phylogenetic trees for SARS-CoV-2 in Brazil, generated by the Neighbor-Joining method from the **a)** proteomes and **b)** genomes. Total of 8720 samples, data from release 609 of GISAID, using PANGO nomenclature v3.0.5. Complete trees in newick format available in the **Suplementary 5** and **Suplementary 6**

**a)** Proteome

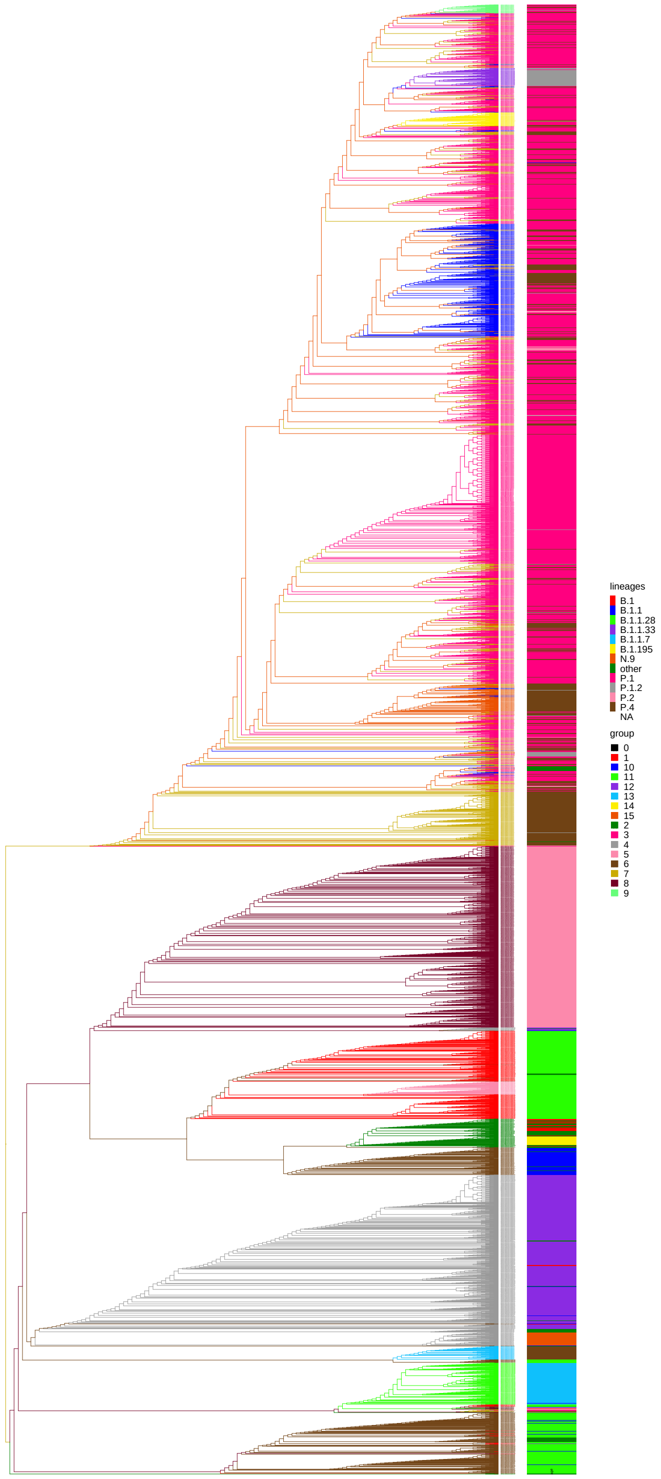


**b)** Genome

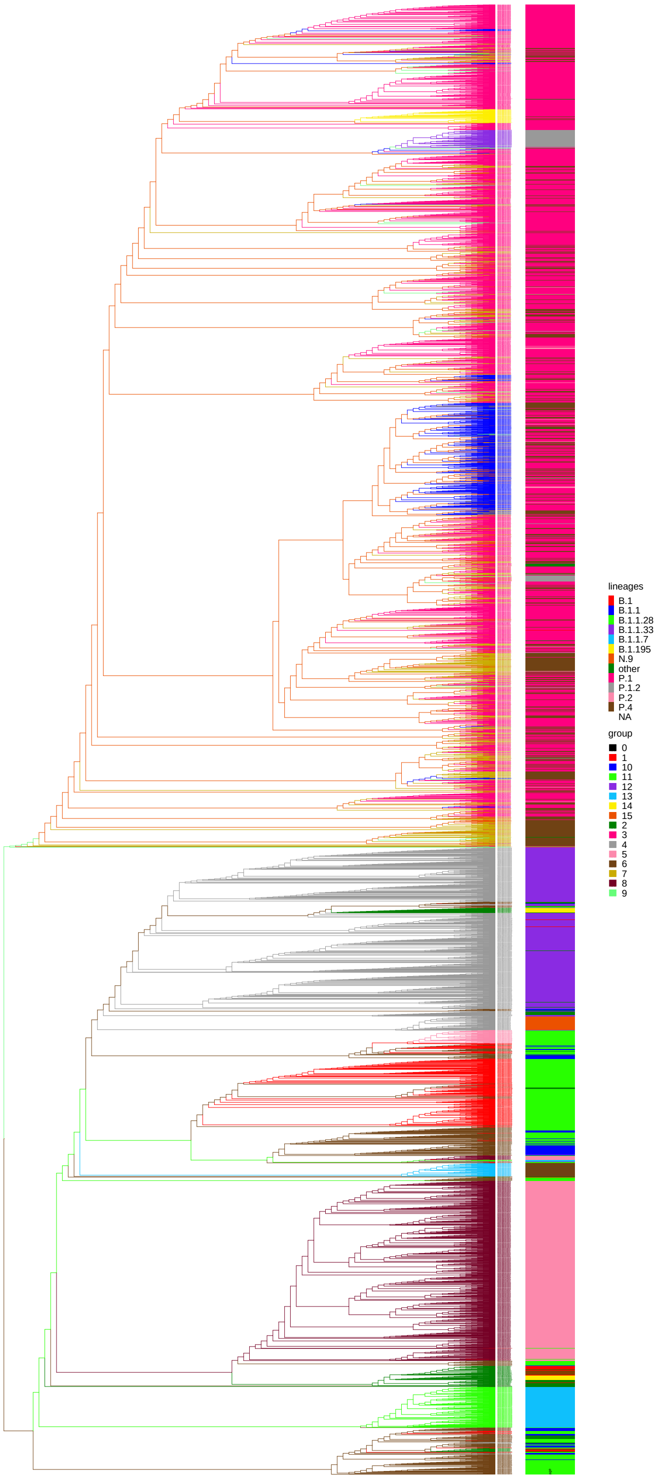

**Figure S7:** Phylogenetic tree of the origin of P.1 lineage in 2020. Phylogeny composed of the samples closest to P.1 samples from 2020 selected by distance matrix. In blue are the 50 Brazilian and 6 Peruvian P.1 samples from 2020, in green the two ancestral samples of the P.1 variant. In pink the samples indicated by Naveca *et al* (17) as ancestors of P.1. In red the Wuhan reference sample. Complete tree in newick format available in the **Suplementary 7**.

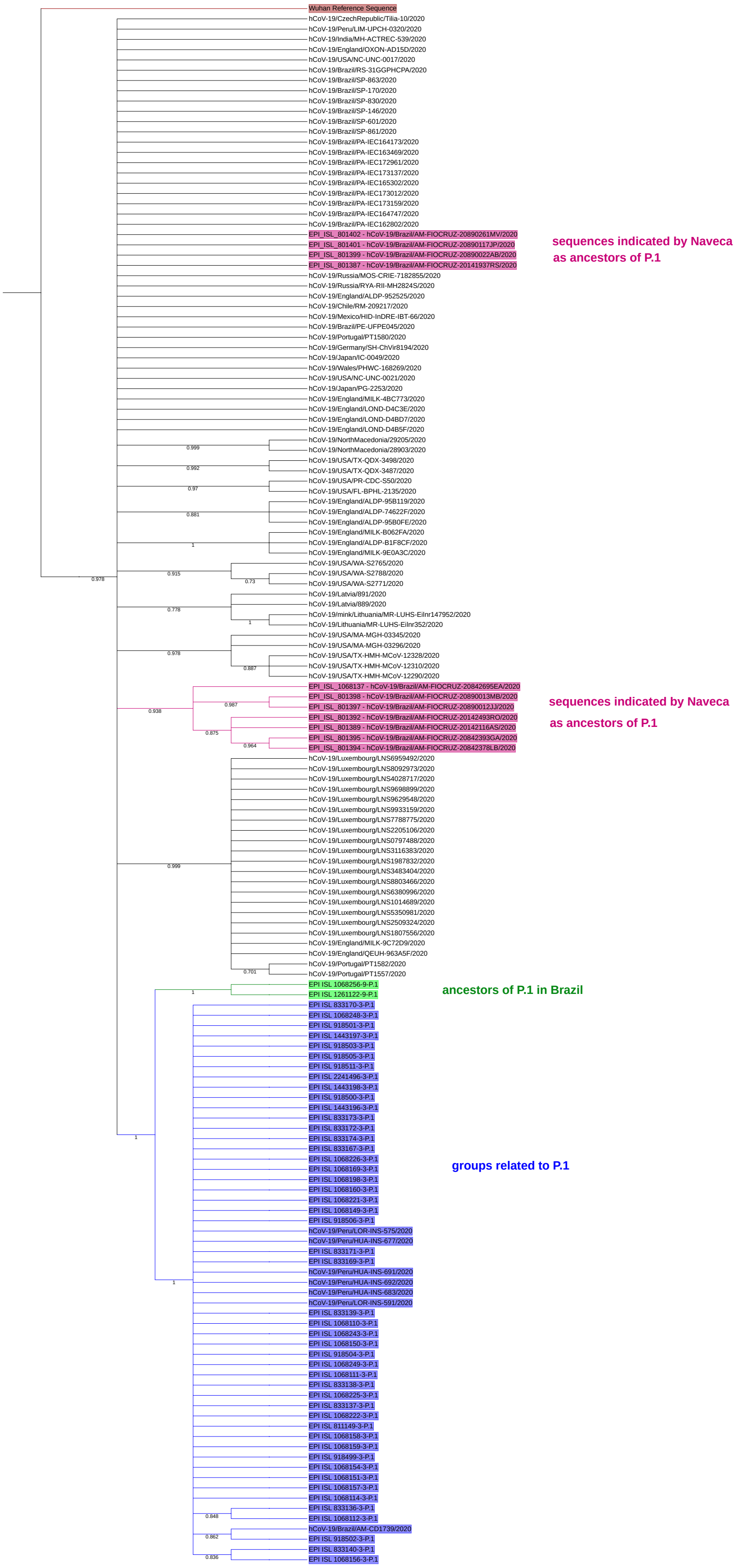

**Figure S8:** Proportional stacked bar chart representing proportional abundance of variants by month for each Brazilian state. Month 2 corresponds to February 2020, and 13 to January 2021.

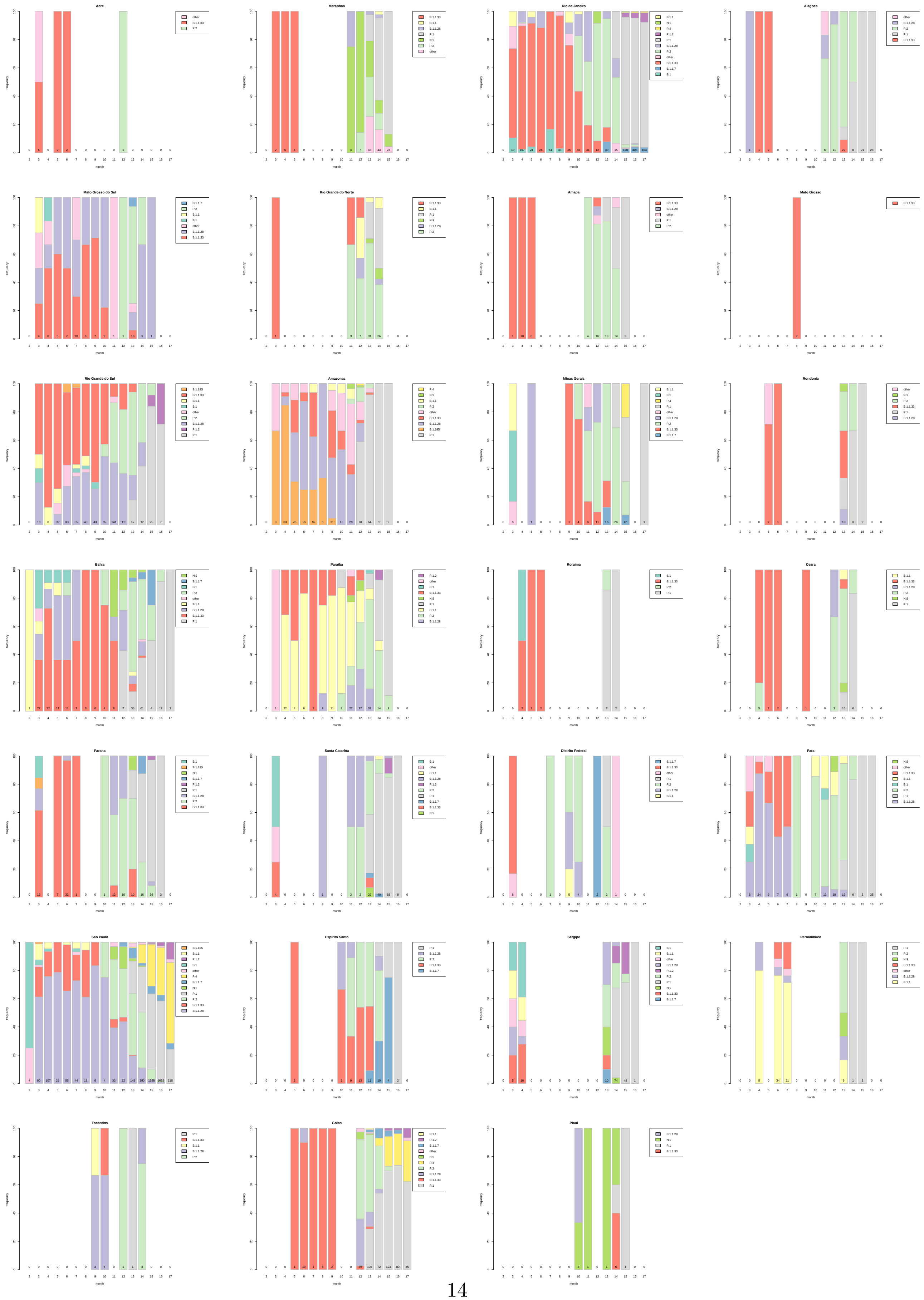

**Figure S9:** t-SNE in time. Time scale in weeks, since the first week of 2020. The TP1 group currently predominates and the older clusters are almost extinct.

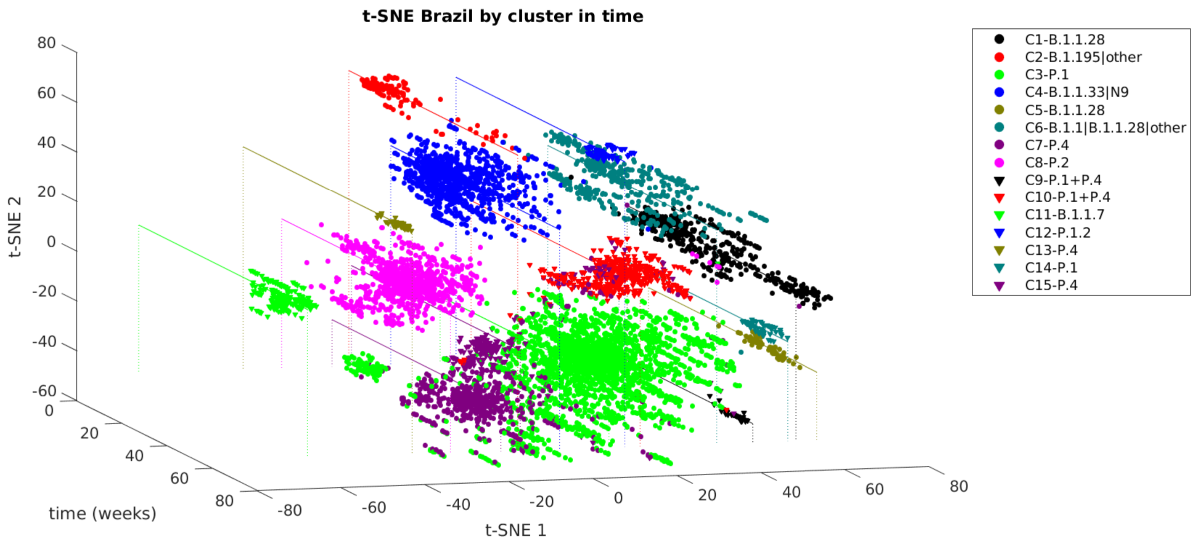

**Figure S10:** Richness by country until the end of May 2021. Despite having a much higher number of cases than European countries, Brazil and India have a much smaller sample.

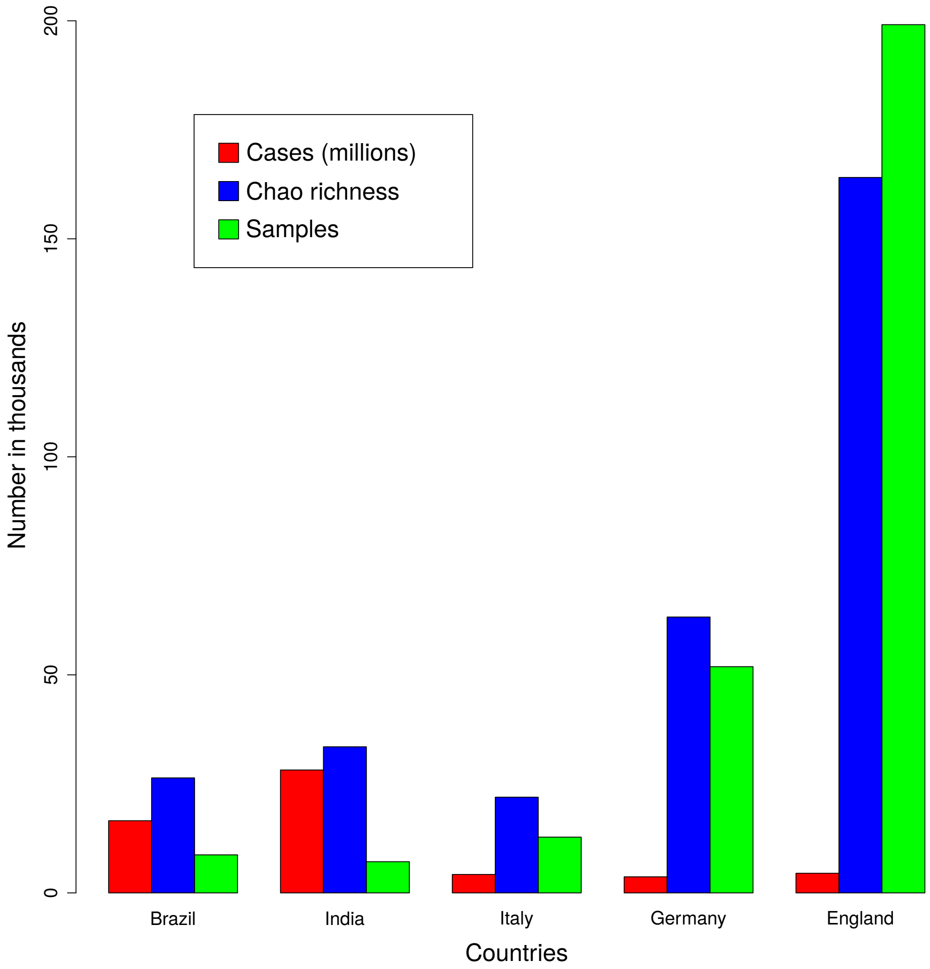

**Figure S11:** Proportional stacked bar chart representing proportional abundance of lineage by months for the year 2021. We use the PANGO nomenclature version 3.1.11 (2021-08-09). Histogram normalized by months, from January to August 2021. The numbers inside the bars indicate the number of sequencing performed each month.

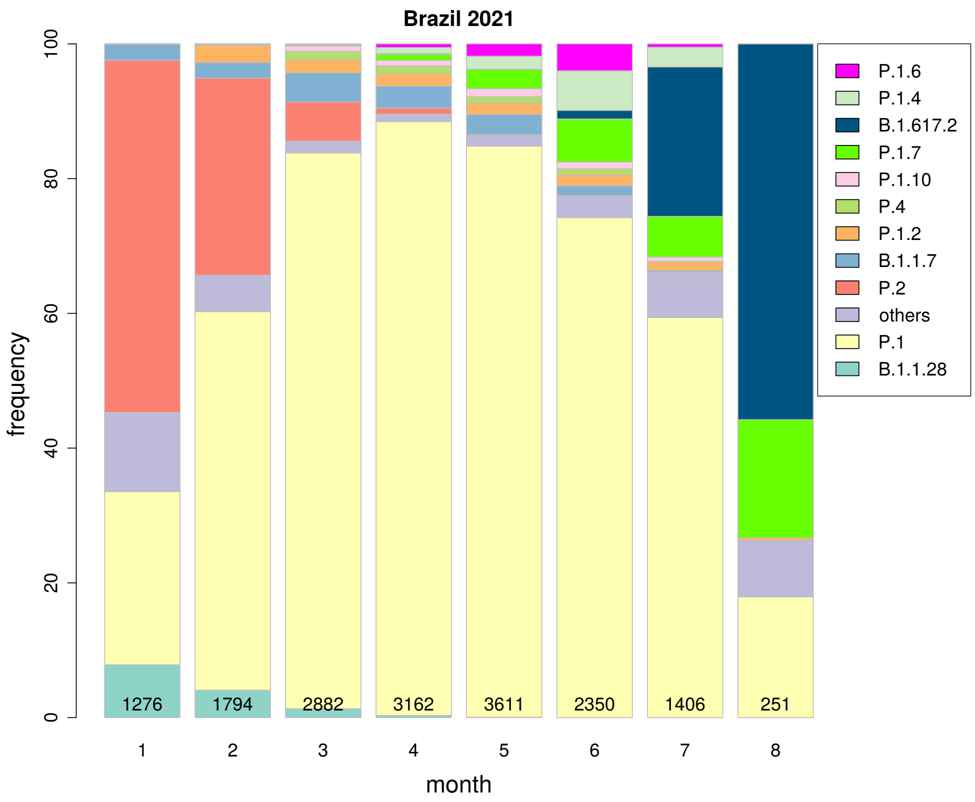

**Figure S12:** Coverage and Richness by state by the Chao metric. The coverage of the states of RR, RN, PI and MS could not be calculated due to low sampling.

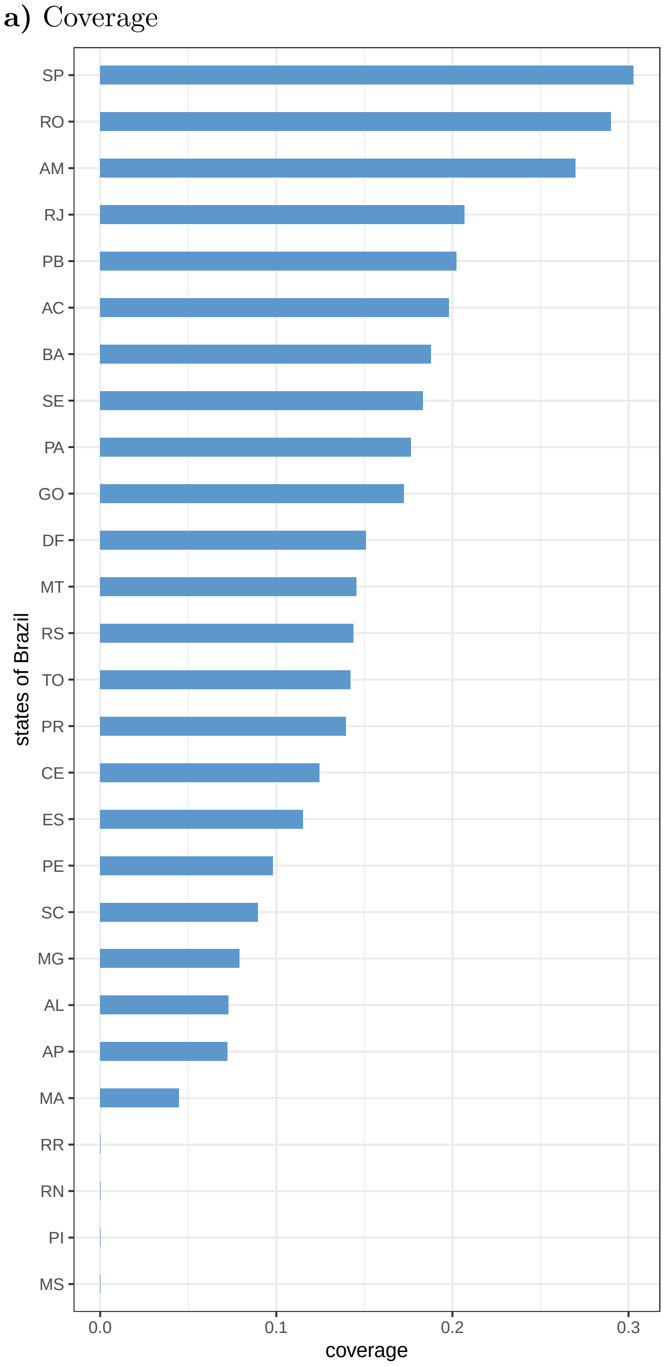


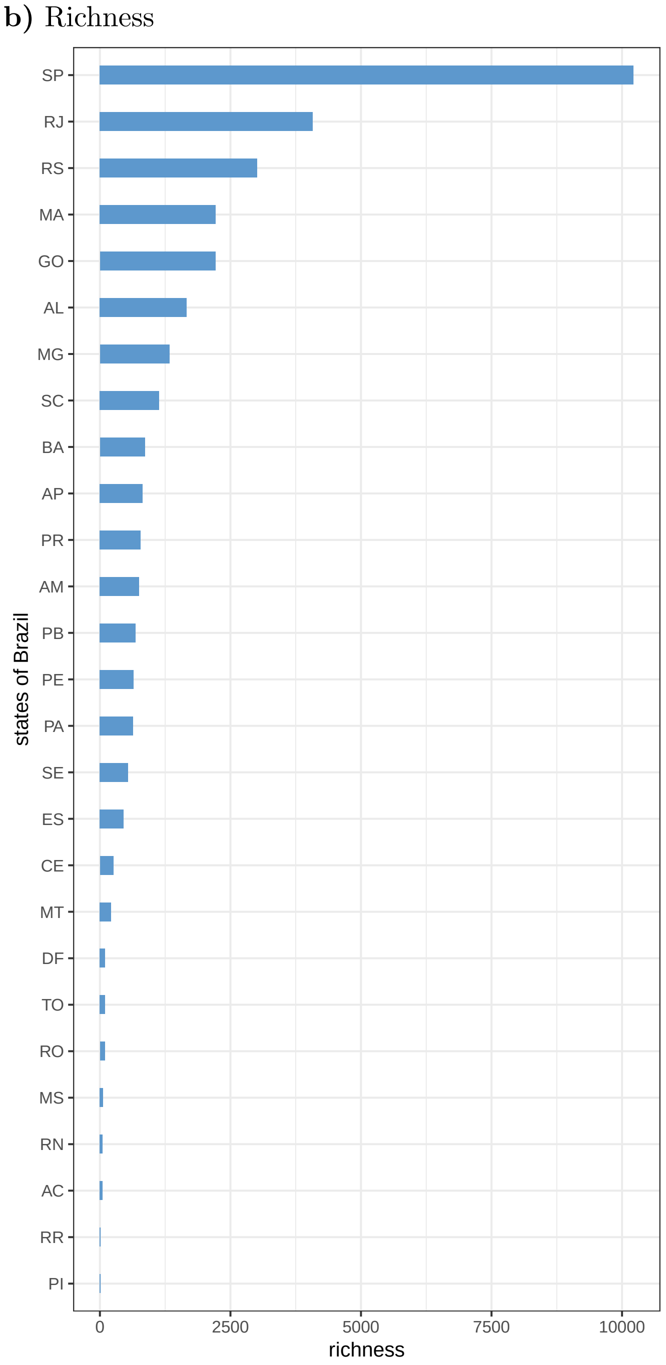

**Table S1:** Problable annotation errors of variants. Possible recombination events have not been discarded for the listed samples.

| EPI code | cluster | PANGO variant | rectification | justification |
|---|---|---|---|---|
| EPI_ISL_2188014, EPI_ISL_2274105, EPI_ISL_1628358 | 3 | B.1.617.1 | P.1 | samples have characteristic mutations of the P.1 variant of the cluster 3, and does not have the characteristics of lineage B.1.617.1. Is inserted next to the branch of P.1 in the genomic and proteomic trees. |
| EPI_ISL_2344990 | 3 | B.1 | P.1.1 | the sample is inserted in the tree of P.1 in the branch of P.1.1 and has mutations characteristic of P.1.1. |
| EPI_ISL_2209844 | 7 | P.1 | P.4 | the sample is inserted in the tree of P.4 and has mutations characteristic of P.4 of the cluster 7. |
| EPI_ISL_1445131, EPI_ISL_1795191, EPI_ISL_1966175, EPI_ISL_1966818, EPI_ISL_1966819, EPI_ISL_1966821 | 7 | P.2 | P.4 | the sample is inserted in the branch of P.4 (100 % bootstrap in the TP1 group) and has mutations characteristic of P.4 of cluster 7. But they have an N:A119S mutation characteristic of P.2. |
| EPI_ISL_2308409, EPI_ISL_2308456, EPI_ISL_2308471, EPI_ISL_2308473 | 12 | P.1 | P.1.2 | have the non-synonymous mutations characteristic of P.1.2 (ORF3a:D155Y,ORF1a:D762G and ORF1a:T1820I) and are inserted within the branch of P.1.2 (with bootstrap of 100%) |

**Table S2:** Distribution of PANGO lineages (v 3.0.5) in each cluster.

| clusters | pangolin lineage | frequency |
|---|---|---|
| 1 | B.1.1 | 2 |
| 1 | B.1.1.28 | 467 |
| 1 | B.1.1.332 | 8 |
| 2 | A.1 | 1 |
| 2 | A.2 | 3 |
| 2 | B | 3 |
| 2 | B.1 | 59 |
| 2 | B.1.111 | 5 |
| 2 | B.1.160.25 | 1 |
| 2 | B.1.177.32 | 1 |
| 2 | B.1.177.52 | 1 |
| 2 | B.1.195 | 54 |
| 2 | B.1.212 | 15 |
| 2 | B.1.234 | 2 |
| 2 | B.1.240 | 1 |
| 2 | B.1.319 | 1 |
| 2 | B.1.351 | 5 |
| 2 | B.1.547 | 1 |
| 2 | B.1.575 | 1 |
| 2 | B.1.617.2 | 1 |
| 2 | B.1.9 | 1 |
| 2 | B.1.91 | 15 |
| 2 | B.3 | 2 |
| 2 | B.39 | 5 |
| 2 | B.6 | 1 |
| 3 | B.1 | 1 |
| 3 | B.1.1 | 1 |
| 3 | B.1.617.1 | 2 |
| 3 | P.1 | 2966 |
| 3 | P.1.1 | 18 |
| 3 | P.1.2 | 26 |
| 3 | P.2 | 27 |
| 3 | P.4 | 21 |
| 4 | B.1 | 4 |
| 4 | B.1.1 | 11 |
| 4 | B.1.1.161 | 7 |
| 4 | B.1.1.33 | 909 |
| 4 | N.1 | 2 |
| 4 | N.10 | 19 |
| 4 | N.4 | 4 |
| 4 | N.9 | 81 |
| 5 | B.1.1.28 | 77 |
| 6 | B.1 | 1 |
| 6 | B.1.1 | 171 |
| 6 | B.1.1.192 | 1 |
| 6 | B.1.1.277 | 3 |
| 6 | B.1.1.28 | 317 |
| 6 | B.1.1.372 | 1 |
| 6 | B.1.1.378 | 29 |
| 6 | B.1.1.462 | 1 |
| 6 | B.1.1.519 | 2 |
| 6 | B.1.1.74 | 1 |
| 6 | B.59 | 1 |
| 6 | C.14 | 1 |
| 6 | C.37 | 2 |
| 7 | P.1 | 4 |
| 7 | P.1.1 | 4 |
| 7 | P.1.2 | 8 |
| 7 | P.2 | 6 |
| 7 | P.4 | 732 |

| continuation... | | |
|---|---|---|
| clusters | pangolin lineage | frequency |
| 8 | B.1.1.28 | 1 |
| 8 | P.2 | 1089 |
| 9 | P.1 | 31 |
| 9 | P.1.2 | 4 |
| 9 | P.4 | 15 |
| 10 | P.1 | 491 |
| 10 | P.1.1 | 2 |
| 10 | P.1.2 | 3 |
| 10 | P.2 | 14 |
| 10 | P.4 | 155 |
| 11 | B.1.1 | 1 |
| 11 | B.1.1.7 | 248 |
| 11 | P.1 | 1 |
| 12 | P.1 | 4 |
| 12 | P.1.2 | 100 |
| 13 | P.4 | 79 |
| 14 | B.1.1.28 | 2 |
| 14 | P.1 | 74 |
| 14 | P.4 | 5 |
| 15 | P.1 | 1 |
| 15 | P.1.1 | 4 |
| 15 | P.1.2 | 12 |
| 15 | P.4 | 267 |

**Table S3:** Division of lineages into clusters using only the vectorized spike protein. The predominant lineages in each cluster are listed. The complete listing of observed lineages by cluster is available in Table S4.

| cluster | predominant lineage | number of samples | first case |
|---|---|---|---|
| groups related to the P.1 variant – TP1 | | 4999 | 2020-10-01 |
| 3 | P.1 + P.1.2 | 3767 | 2020-10-01 |
| 4 | P.4 | 902 | 2020-12-21 |
| 8 | P.4 | 330 | 2021-02-19 |
| early group – T0 | | 3391 | 2020-02-25 |
| 1 | B.1.1.28 | 895 | 2020-03-05 |
| 2 | B.1.1.33 + B.1.1 + B.1.195 | 1307 | 2020-02-25 |
| 5 | P.2+N.9 | 1189 | 2020-04-13 |
| others | | 330 | 2020-12-21 |
| 6 | B.1.1.7 | 251 | 2020-12-21 |
| 7 | P.4 | 79 | 2021-02-17 |

**Table S4:** Distribution of PANGO lineages (v 3.0.5) in each cluster, using only spike proteins.

| cluster | pangolin lineage | frequency |
|---|---|---|
| 1 | B.1.1 | 15 |
| 1 | B.1.1.277 | 3 |
| 1 | B.1.1.28 | 837 |
| 1 | B.1.1.33 | 1 |
| 1 | B.1.1.332 | 8 |
| 1 | B.1.1.378 | 29 |
| 1 | N.1 | 1 |
| 1 | P.1 | 1 |
| 2 | A.1 | 1 |
| 2 | A.2 | 3 |
| 2 | B | 3 |
| 2 | B.1 | 63 |
| 2 | B.1.1 | 169 |
| 2 | B.1.1.161 | 7 |
| 2 | B.1.1.192 | 1 |
| 2 | B.1.1.33 | 908 |
| 2 | B.1.1.372 | 1 |
| 2 | B.1.1.462 | 1 |
| 2 | B.1.1.519 | 2 |
| 2 | B.1.1.74 | 1 |
| 2 | B.1.111 | 5 |
| 2 | B.1.160.25 | 1 |
| 2 | B.1.177.32 | 1 |
| 2 | B.1.177.52 | 1 |
| 2 | B.1.195 | 54 |
| 2 | B.1.212 | 15 |
| 2 | B.1.234 | 2 |
| 2 | B.1.240 | 1 |
| 2 | B.1.319 | 1 |
| 2 | B.1.351 | 5 |
| 2 | B.1.547 | 1 |
| 2 | B.1.575 | 1 |
| 2 | B.1.617.2 | 1 |
| 2 | B.1.9 | 1 |
| 2 | B.1.91 | 15 |
| 2 | B.3 | 2 |
| 2 | B.39 | 5 |
| 2 | B.59 | 1 |
| 2 | B.6 | 1 |
| 2 | C.14 | 1 |
| 2 | C.37 | 2 |
| 2 | N.1 | 1 |
| 2 | N.10 | 19 |
| 2 | N.4 | 4 |
| 2 | N.9 | 6 |
| 3 | B.1 | 1 |
| 3 | B.1.1 | 1 |
| 3 | B.1.617.1 | 2 |
| 3 | P.1 | 3563 |
| 3 | P.1.1 | 19 |
| 3 | P.1.2 | 116 |
| 3 | P.2 | 41 |
| 3 | P.4 | 24 |
| 4 | P.1 | 7 |
| 4 | P.1.1 | 5 |
| 4 | P.1.2 | 21 |
| 4 | P.2 | 6 |
| 4 | P.4 | 863 |
| 5 | B.1.1.28 | 24 |
| 5 | N.9 | 75 |
| 5 | P.2 | 1089 |
| 5 | P.4 | 1 |
| 6 | B.1 | 1 |
| 6 | B.1.1 | 1 |
| 6 | B.1.1.28 | 1 |
| 6 | B.1.1.7 | 248 |
| 7 | P.4 | 79 |
| 8 | B.1.1.28 | 2 |
| 8 | P.1 | 1 |
| 8 | P.1.1 | 4 |
| 8 | P.1.2 | 16 |
| 8 | P.4 | 307 |

**Table S5:** Consensus mutations in the Brazilian context. The frequency is relative to the 8720 quality sequences analyzed.

| mutation | frequence (seq.) | remark |
|---|---|---|
| ORF1b:P314L | 8660 | present in all clusters |
| S:D614G | 8629 | present in all clusters – provides high transmissibility, low vaccine efficiency (9) and increases the infectivity rate due to a large conformational change of the protein (18; 3). Presented high levels of viral load compared to the original Wuhan strain (4) |
| N:R203K | 8512 | present in all clusters, except in cluster 2 – suggested as a critical region for virus maturation and assembly; highly observed in the world (18) |
| N:G204R | 8501 | present in all clusters, except in cluster 2 – suggested as a critical region for virus maturation and assembly; highly observed in the world (18) |
| S:V1176F | 7080 | present in all clusters significantly, except in clusters 2, 4 e 11 |

**Table S6:** Consensus mutations of TP1 group in the Brazilian context. The frequency is relative to the 5000 quality sequences analyzed belonging to the group, for general mutations. For specific clusters, the frequency is relative to their own number of sequences.

| mutation | frequence (seq.) | remark |
|---|---|---|
| S:E484K | 4992 | present in all clusters of the TP1 group – the mutation strengthens the link with hACE2, providing high transmissibility and low vaccine efficiency (9). The association S:K417T, S:E484K and S:N501Y promotes high affinity with ACE2 (40-50% greater than the wild type) and resistance to antibodies (15) |
| S:N501Y | 4990 | present in all clusters of the TP1 group – the mutation strengthens the link with hACE2, providing high transmissibility and low vaccine efficiency (9). The association S:K417T, S:E484K and S:N501Y promotes high affinity with ACE2 (40-50% greater than the wild type) and resistance to antibodies (15) |
| S:H655Y | 4988 | present in all clusters of the TP1 group; promotes escape from the immune system (13) |
| S:L18F | 4991 | present in all clusters of the TP1 group; compromises the binding of neutralizing antibodies and the immune response (14) |
| S:T1027I | 4984 | present in all clusters of the TP1 group |
| S:P26S | 4987 | present in all clusters of the TP1 group |
| S:R190S | 4964 | present in all clusters of the TP1 group |
| N:P80R | 4977 | present in all clusters of the TP1 group |
| ORF1a:K1795Q | 4992 | present in all clusters of the TP1 group |
| ORF1a:S1188L | 4960 | present in all clusters of the TP1 group |
| ORF3a:S253P | 4973 | present in all clusters of the TP1 group |
| ORF9b:Q77E | 4977 | present in all clusters of the TP1 group |
| S:K417T | 4035 | absent in the cluster 7; the association S:K417T, S:E484K and S:N501Y promotes high affinity with ACE2 (40-50% greater than the wild type) and resistance to antibodies (15) |
| S:D138Y | 4656 | absent in the cluster 15 |
| ORF8:E92K | 4763 | absent in the cluster14 |
| ORF1b:E1264D | 4904 | absent in the cluster14 |
| S:T20N | 4908 | absent in the cluster14 |
| ORF1a:S3675- | 4840 | present in all clusters of the TP1 group |
| ORF1a:G3676- | 4840 | present in all clusters of the TP1 group |
| ORF1a:F3677- | 4839 | present in all clusters of the TP1 group |
| **cluster 10 (665 samples)** | | |
| ORF1a:S2947N | 665 | |
| **cluster 12 (104 samples)** | | |
| ORF3a:D155Y | 104 | |
| ORF1a:D762G | 104 | |
| ORF1a:T1820I | 103 | |
| ORF3a:L83F | 49 | |
| **cluster 14 (81 samples)** | | |
| ORF1a:D2980H | 81 | |
| N:P383L | 79 | |

**Table S7:** Characteristic mutations of each cluster of T0 group.

| mutation | frequence (seq.) | remark |
|---|---|---|
| **cluster 1 (477 samples)** | | |
| ORF1a:L3930F | 476 | shared with clusters 5 and 8 |
| S:S689I | 103 | |
| ORF3a:G224V | 103 | |
| S:M153T | 97 | |
| **cluster 2 (179 samples)** | | |
| ORF1a:S142- | 53 | present in the B.1.195 samples of the cluster |
| ORF1a:K141- | 53 | present in the B.1.195 samples of the cluster |
| ORF1a:F143- | 53 | present in the B.1.195 samples of the cluster |
| **cluster 4 (1037 samples)** | | |
| N:I292T | 1033 | possible highly stabilizing mutation (18) |
| ORF6:I33T | 1017 | |
| S:E484K | 101 | shared with cluster 8 and TP1 group – the mutation strengthens the link with hACE2, providing high transmissibility and low vaccine efficiency (9). The association S:K417T, S:E484K and S:N501Y promotes high affinity with ACE2 (40-50% greater than the wild type) and resistance to antibodies (15) |
| **cluster 5 (77 samples)** | | |
| ORF9b:P10S | 77 | |
| ORF3a:T151I | 77 | |
| ORF1a:V2588F | 77 | |
| ORF1a:Q3777H | 77 | |
| ORF1a:P2287S | 77 | |
| ORF1a:L3930F | 77 | shared with clusters 1 and 8 |
| ORF1a:L3027F | 77 | |
| N:P13L | 77 | provides stabilization (19), highly observed in the world (18), potentially increases transmissibility and death rate (20) |
| **cluster 6 (531 samples)** | | |
| ORF3a:S253P | 118 | characteristic of TP1 group |
| **cluster 8 (1090 samples)** | | |
| ORF1a:L3930F | 1089 | shared with clusters 1 and 5 |
| N:A119S | 1089 | |
| S:E484K | 1088 | characteristic of TP1 group, shared with cluster 4 – the mutation strengthens the link with hACE2, providing high transmissibility and low vaccine efficiency (9). The association S:K417T, S:E484K and S:N501Y promotes high affinity with ACE2 (40-50% greater than the wild type) and resistance to antibodies (15) |
| N:M234I | 1085 | |
| ORF1a:L3468V | 1012 | |
| **cluster 11 (250 samples)** | | |
| S:N501Y | 250 | characteristic of TP1 group – the mutation strengthens the link with hACE2, providing high transmissibility and low vaccine efficiency (9). The association S:K417T, S:E484K and S:N501Y promotes high affinity with ACE2 (40-50% greater than the wild type) and resistance to antibodies (15) |
| ORF1a:T1001I | 249 | |
| S:T716I | 248 | |
| S:S982A | 248 | in association of S:S982A and S:D614G is related to the conformational variation of the spike, promoting high binding affinity with ACE2 and increased replication (21) |
| S:D1118H | 248 | |
| S:A570D | 248 | in association of S:S982A and S:D614G is related to the conformational variation of the spike, promoting high binding affinity with ACE2 and increased replication (21) |
| ORF8:Y73C | 248 | |
| ORF8:R52I | 248 | |
| ORF1a:A1708D | 248 | |
| N:S235F | 248 | S:P681H together with N:S235F potentially characterizes immune escape (22) |
| S:P681H | 247 | causes greater infectivity due to changes in protein conformation that increase affinity with the furin enzyme (Mohammad,etal,2021). S:P681H together with N:S235F potentially characterizes immune escape (22) |
| N:D3L | 246 | |
| ORF8:Q27* | 241 | |
| ORF1a:I2230T | 241 | |
| ORF1a:S3675- | 237 | characteristic of TP1 group |
| ORF1a:G3676- | 237 | characteristic of TP1 group |
| ORF1a:F3677- | 237 | characteristic of TP1 group |
| S:H69- | 233 | the pair of H69/V70 inserts increases infectivity (23) |
| S:V70- | 231 | the pair of H69/V70 inserts increases infectivity (23) |
| S:Y144- | 230 | |
| **cluster 13 (79 samples)** | | |
| S:L452R | 79 | mutation makes the virus 20% more transmissible, increases *in vitro* infectivity, and reduces antibody neutralization (8), conferring low vaccine efficacy (9) |
| ORF3a:V50I | 79 | |
| ORF1b:Y822C | 79 | |
| ORF1a:Q3729K | 79 | |
| ORF1a:P971L | 79 | |
| ORF1a:L3915F | 79 | |
| ORF1a:L3201P | 79 | |
| ORF1a:A516T | 79 | |
| ORF1a:A3143V | 79 | |
| N:A220V | 79 | changes the conformation of proteins with the potential to alter pathogenicity and/or response to vaccination, affecting the activity of the protein binding site. A mutation with 163k cases worldwide and increasing, one of the 10 most frequent in the world, but rare in Brazil (10; 11). |
| S:I720V | 78 | |
| S:Q173K | 77 | the mutation potentially increases the fitness of the virus (12) |
| S:N164K | 76 | |
| S:S704L | 75 | |
| S:G142V | 70 | |

**Table S8:** Worldwide sequences near to P.1 in 2020, defined by Euclidean distance. There are listed the 91 unique sequences closest to the 50 Brazilian samples of P.1 from 2020.

| ID | variant |
|---|---|
| hCoV-19/Peru/HUA-INS-677/2020 | P.1 |
| hCoV-19/Peru/LOR-INS-575/2020 | P.1 |
| hCoV-19/Peru/HUA-INS-691/2020 | P.1 |
| hCoV-19/Peru/LOR-INS-591/2020 | P.1 |
| hCoV-19/Peru/HUA-INS-692/2020 | P.1 |
| hCoV-19/Brazil/AM-CD1739/2020 | P.4 |
| hCoV-19/Peru/HUA-INS-683/2020 | P.1 |
| hCoV-19/Brazil/PA-IEC173137/2020 | B.1.1.28 |
| hCoV-19/Brazil/PA-IEC164747/2020 | B.1.1.28 |
| hCoV-19/Brazil/SP-146/2020 | B.1.1.28 |
| hCoV-19/Brazil/SP-601/2020 | B.1.1.28 |
| hCoV-19/Brazil/SP-861/2020 | B.1.1.28 |
| hCoV-19/Brazil/SP-863/2020 | B.1.1.28 |
| hCoV-19/Brazil/SP-830/2020 | B.1.1.28 |
| hCoV-19/Brazil/PA-IEC164173/2020 | B.1.1.28 |
| hCoV-19/Brazil/PA-IEC172961/2020 | B.1.1.28 |
| hCoV-19/Brazil/PA-IEC173012/2020 | B.1.1.28 |
| hCoV-19/Brazil/SP-170/2020 | B.1.1.28 |
| hCoV-19/Brazil/PA-IEC162802/2020 | B.1.1.28 |
| hCoV-19/Brazil/AM-FIOCRUZ-20890261MV/2020 | B.1.1.28 |
| hCoV-19/Brazil/PA-IEC173159/2020 | B.1.1.28 |
| hCoV-19/Brazil/PA-IEC163469/2020 | B.1.1.28 |
| hCoV-19/Brazil/PA-IEC165302/2020 | B.1.1.28 |
| hCoV-19/England/MILK-4BC773/2020 | B.1.1 |
| hCoV-19/England/LOND-D4B5F/2020 | B.1.1 |
| hCoV-19/England/LOND-D4C3E/2020 | B.1.1 |
| hCoV-19/England/LOND-D4BD7/2020 | B.1.1 |
| hCoV-19/USA/WA-S2765/2020 | B.1.1 |
| hCoV-19/Japan/PG-2253/2020 | B.1.1.48 |
| hCoV-19/India/MH-ACTREC-539/2020 | B.1.1.306 |
| hCoV-19/USA/NC-UNC-0021/2020 | B.1.1 |
| hCoV-19/England/ALDP-952525/2020 | B.1.1 |
| hCoV-19/Mexico/HID-InDRE-IBT-66/2020 | B.1.1 |
| hCoV-19/England/OXON-AD15D/2020 | B.1.1.10 |
| hCoV-19/Portugal/PT1580/2020 | B.1.1.421 |
| hCoV-19/USA/WA-S2788/2020 | B.1.1 |
| hCoV-19/USA/WA-S2771/2020 | B.1.1 |
| hCoV-19/Russia/RYA-RII-MH2824S/2020 | B.1.1 |
| hCoV-19/Peru/LIM-UPCH-0320/2020 | B.1.1.485 |
| hCoV-19/USA/MA-MGH-03345/2020 | B.1.1.192 |
| hCoV-19/USA/MA-MGH-03296/2020 | B.1.1.192 |
| hCoV-19/CzechRepublic/Tilia-10/2020 | B.1.1 |
| hCoV-19/USA/TX-HMH-MCoV-12328/2020 | B.1.1.192 |
| hCoV-19/USA/TX-HMH-MCoV-12310/2020 | B.1.1.192 |
| hCoV-19/USA/TX-HMH-MCoV-12290/2020 | B.1.1.192 |
| hCoV-19/Russia/MOS-CRIE-7182855/2020 | B.1.1 |
| hCoV-19/England/ALDP-95B0FE/2020 | B.1.1 |
| hCoV-19/England/ALDP-74622F/2020 | B.1.1 |
| hCoV-19/England/ALDP-95B119/2020 | B.1.1 |
| hCoV-19/Brazil/PE-UFPE045/2020 | B.1.1 |
| hCoV-19/USA/NC-UNC-0017/2020 | B.1.1.1 |
| hCoV-19/England/MILK-9C72D9/2020 | B.1.1.198 |
| hCoV-19/Luxembourg/LNS1014689/2020 | B.1.1.198 |
| hCoV-19/Luxembourg/LNS8803466/2020 | B.1.1.198 |
| hCoV-19/Luxembourg/LNS8092973/2020 | B.1.1.198 |
| hCoV-19/Luxembourg/LNS9698899/2020 | B.1.1.198 |
| hCoV-19/Luxembourg/LNS7788775/2020 | B.1.1.198 |
| hCoV-19/Luxembourg/LNS6380996/2020 | B.1.1.198 |
| hCoV-19/Luxembourg/LNS3116383/2020 | B.1.1.198 |
| hCoV-19/Luxembourg/LNS1807556/2020 | B.1.1.198 |
| hCoV-19/Luxembourg/LNS9933159/2020 | B.1.1.198 |
| hCoV-19/Portugal/PT1557/2020 | B.1.1.198 |
| hCoV-19/Portugal/PT1582/2020 | B.1.1.198 |
| hCoV-19/Luxembourg/LNS6959492/2020 | B.1.1.198 |
| hCoV-19/Luxembourg/LNS9629548/2020 | B.1.1.198 |
| hCoV-19/Luxembourg/LNS0797488/2020 | B.1.1.198 |
| hCoV-19/Luxembourg/LNS5350981/2020 | B.1.1.198 |
| hCoV-19/Luxembourg/LNS1987832/2020 | B.1.1.198 |
| hCoV-19/Luxembourg/LNS2205106/2020 | B.1.1.198 |
| hCoV-19/Luxembourg/LNS2509324/2020 | B.1.1.198 |
| hCoV-19/Luxembourg/LNS4028717/2020 | B.1.1.198 |
| hCoV-19/England/QEUH-963A5F/2020 | B.1.1.198 |
| hCoV-19/Luxembourg/LNS3483404/2020 | B.1.1.198 |
| hCoV-19/NorthMacedonia/29205/2020 | B.1.1.428 |
| hCoV-19/Japan/IC-0049/2020 | B.1.1.1 |
| hCoV-19/Germany/SH-ChVir8194/2020 | B.1.1 |
| hCoV-19/NorthMacedonia/28903/2020 | B.1.1.428 |
| hCoV-19/Wales/PHWC-168269/2020 | B.1.1 |
| hCoV-19/USA/TX-QDX-3487/2020 | B.1.1 |
| hCoV-19/Chile/RM-209217/2020 | N.4 |
| hCoV-19/England/MILK-9E0A3C/2020 | B.1.1.198 |
| hCoV-19/England/MILK-B062FA/2020 | B.1.1.46 |
| hCoV-19/USA/TX-QDX-3498/2020 | B.1.1 |
| hCoV-19/mink/Lithuania/MR-LUHS-Eilnr147952/2020 | B.1.1.464 |
| hCoV-19/Brazil/RS-31GGPHCPA/2020 | B.1.1.28 |
| hCoV-19/England/ALDP-B1F8CF/2020 | B.1.1.46 |
| hCoV-19/Lithuania/MR-LUHS-Eilnr352/2020 | B.1.1.280 |
| hCoV-19/USA/FL-BPHL-2135/2020 | B.1 |
| hCoV-19/USA/PR-CDC-S50/2020 | B.1 |
| hCoV-19/Latvia/889/2020 | B.1.1.280 |
| hCoV-19/Latvia/891/2020 | B.1.1.280 |

**Table S9:** List of recombinants indicated by the RAPR tool for the Brazilian P.1 sequences in 2020. The asterisk indicates the events that potentially generated B.1.1 and B.1.1.28 cluster 6 recombinants from P.1 and the USA sequence of B.1.1.1.

| recombinant sample | parental sequence 1 | parental sequence 2 | p-value |
|---|---|---|---|
| *Brazil/PA-IEC162802 ( EPI_ISL_458140 - B.1.1.28 ) | Brazil/AM-FIOCRUZ-20143088ER ( EPI_ISL_1068110 - P.1 ) | USA/NC-UNC-0017 ( EPI_ISL_831339 - B.1.1.1 ) | $5{,}12 \times 10^{-6}$ |
| *Brazil/AM-FIOCRUZ-20890261MV ( EPI_ISL_801402 - B.1.1.28 ) | Brazil/AM-FIOCRUZ-20143088ER ( EPI_ISL_1068110 - P.1 ) | USA/NC-UNC-0017 ( EPI_ISL_831339 - B.1.1.1 ) | $3{,}01 \times 10^{-6}$ |
| Brazil/RS-31GGPHCPA ( EPI_ISL_1163736 - B.1.1.28 ) | Brazil/PB-BA1200-250009713 ( EPI_ISL_2241496 - P.1 ) | Lithuania/MR-LUHS-Eilnr352 ( EPI_ISL_636871 - B.1.1.280 ) | $4{,}805 \times 10^{-4}$ |
| Brazil/PE-UFPE045 ( EPI_ISL_1117381 - B.1.1 ) | USA/NC-UNC-0017 ( EPI_ISL_831339 - B.1.1.1 ) | Brazil/PA-IEC173159 ( EPI_ISL_848562 - B.1.1.28 ) | $3{,}996 \times 10^{-4}$ |
| Wales/PHWC-168269 ( EPI_ISL_494217 - B.1.1 ) | Brazil/AM-FIOCRUZ-20143088ER ( EPI_ISL_1068110 - P.1 ) | USA/NC-UNC-0017 ( EPI_ISL_831339 - B.1.1.1 ) | $4{,}96 \times 10^{-7}$ |
| Russia/RYA-RII-MH2824S ( EPI_ISL_639968 - B.1.1 ) | Brazil/AM-FIOCRUZ-20143088ER ( EPI_ISL_1068110 - P.1 ) | USA/NC-UNC-0017 ( EPI_ISL_831339 - B.1.1.1 ) | $2{,}08 \times 10^{-5}$ |
| Chile/RM-209217 ( EPI_ISL_1167679 - N.4 ) | Brazil/PB-BA1200-250009713 ( EPI_ISL_2241496 - P.1 ) | USA/NC-UNC-0017 ( EPI_ISL_831339 - B.1.1.1 ) | $1{,}46 \times 10^{-7}$ |
| Russia/MOS-CRIE-7182855 ( EPI_ISL_745223 - B.1.1 ) | Brazil/AM-FIOCRUZ-20143088ER ( EPI_ISL_1068110 - P.1 ) | USA/NC-UNC-0017 ( EPI_ISL_831339 - B.1.1.1 ) | $2{,}08 \times 10^{-5}$ |
| Peru/LIM-UPCH-0320 ( EPI_ISL_812477 - B.1.1.485 ) | Brazil/AM-FIOCRUZ-20143103JT ( EPI_ISL_833136 - P.1 ) | USA/NC-UNC-0017 ( EPI_ISL_831339 - B.1.1.1 ) | $3{,}82 \times 10^{-7}$ |
| CzechRepublic/Tilia-10 ( EPI_ISL_906062 - B.1.1 ) | Peru/HUA-INS-683 ( EPI_ISL_1111143 - P.1 ) | USA/NC-UNC-0017 ( EPI_ISL_831339 - B.1.1.1 ) | $2{,}33 \times 10^{-7}$ |
| Mexico/HID-InDRE-IBT-66 ( EPI_ISL_1301549 - B.1.1 ) | Japan/IC-0049 ( EPI_ISL_591391 - B.1.1.1 ) | USA/NC-UNC-0017 ( EPI_ISL_831339 - B.1.1.1 ) | $1{,}748 \times 10^{-4}$ |
| England/MILK-9E0A3C ( EPI_ISL_581366 - B.1.1.46 ) | Russia/RYA-RII-MH2824S ( EPI_ISL_639968 - B.1.1 ) | USA/NC-UNC-0017 ( EPI_ISL_831339 - B.1.1.1 ) | $2{,}057 \times 10^{-4}$ |
| Lithuania/MR-LUHS-Eilnr352 ( EPI_ISL_636871 - B.1.1.280 ) | Luxembourg/LNS5350981 ( EPI_ISL_744221 - B.1.1.98 ) | USA/NC-UNC-0017 ( EPI_ISL_831339 - B.1.1.1 ) | $3{,}702 \times 10^{-4}$ |
| USA/FL-BPHL-2135 ( EPI_ISL_653288 - B.1 ) | Brazil/AM-FIOCRUZ-20890261MV ( EPI_ISL_801402 - B.1.1.28 ) | USA/NC-UNC-0017 ( EPI_ISL_831339 - B.1.1.1 ) | $9{,}991 \times 10^{-4}$ |
| Portugal/PT1580 ( EPI_ISL_693533 - B.1.1.421 ) | USA/NC-UNC-0017 ( EPI_ISL_831339 - B.1.1.1 ) | Brazil/PA-IEC173159 ( EPI_ISL_848562 - B.1.1.28 ) | $3{,}996 \times 10^{-4}$ |
| Germany/SH-ChVir8194 ( EPI_ISL_729470 - B.1.1 ) | Brazil/AM-FIOCRUZ-20890261MV ( EPI_ISL_801402 - B.1.1.28 ) | USA/NC-UNC-0017 ( EPI_ISL_831339 - B.1.1.1 ) | $9{,}99 \times 10^{-4}$ |
| Luxembourg/LNS1014689 ( EPI_ISL_739870 - B.1.1.198 ) | England/OXON-AD15D ( EPI_ISL_448567 - B.1.1.10 ) | USA/NC-UNC-0017 ( EPI_ISL_831339 - B.1.1.1 ) | $9{,}99 \times 10^{-4}$ |
| USA/TX-HMH-MCoV-12290 ( EPI_ISL_789934 - B.1.1.192 ) | England/MILK-9E0A3C ( EPI_ISL_581366 - B.1.1.46 ) | USA/NC-UNC-0017 ( EPI_ISL_831339 - B.1.1.1 ) | $2{,}057 \times 10^{-4}$ |
| mink/Lithuania/MR-LUHS-Eilnr147952 ( EPI_ISL_851056 - B.1.1.464 ) | England/ALDP-B1F8CF ( EPI_ISL_646057 - B.1.1.46 ) | USA/NC-UNC-0017 ( EPI_ISL_831339 - B.1.1.1 ) | $7{,}14 \times 10^{-5}$ |
| USA/TX-QDX-3498 ( EPI_ISL_877125 - B.1.1 ) | Russia/RYA-RII-MH2824S ( EPI_ISL_639968 - B.1.1 ) | USA/NC-UNC-0017 ( EPI_ISL_831339 - B.1.1.1 ) | $2{,}057 \times 10^{-4}$ |
| Luxembourg/LNS6380996 ( EPI_ISL_744406 - B.1.1.198 ) | England/OXON-AD15D ( EPI_ISL_448567 - B.1.1.10 ) | USA/NC-UNC-0017 ( EPI_ISL_831339 - B.1.1.1 ) | $9{,}990 \times 10^{-4}$ |
| Latvia/889 ( EPI_ISL_1312400 - B.1.1.280 ) | England/MILK-9E0A3C ( EPI_ISL_581366 - B.1.1.46 ) | Lithuania/MR-LUHS-Eilnr352 ( EPI_ISL_636871 - B.1.1.280 ) | $5{,}97 \times 10^{-5}$ |

**Table S10:** Richness and coverage by states in Brazil, using Chao 1 (24). Number of different refers to the number of non-redundant sequences. The states of RR, PI, RN and MS could not be analyzed due to low sampling.

| State | Number of different | Coverage | Richness |
|---|---|---|---|
| RO | 27 | 0,2899 | 93,13 |
| AC | 10 | 0,198 | 50,50 |
| AM | 203 | 0,2699 | 752,13 |
| RR | 12 | – | – |
| PA | 112 | 0,1765 | 634,56 |
| AP | 59 | 0,072 37 | 815,25 |
| TO | 14 | 0,1421 | 98,52 |
| MA | 99 | 0,0447 | 2214,76 |
| PI | 11 | – | – |
| CE | 32 | 0,1245 | 257,03 |
| RN | 54 | – | – |
| PB | 138 | 0,2022 | 682,49 |
| PE | 63 | 0,097 95 | 643,18 |
| AL | 121 | 0,072 84 | 1661,17 |
| SE | 99 | 0,1833 | 540,10 |
| BA | 162 | 0,1879 | 862,16 |
| MG | 105 | 0,078 94 | 1330,12 |
| ES | 52 | 0,115 | 452,17 |
| RJ | 842 | 0,2067 | 4073,54 |
| SP | 3095 | 0,3028 | 10 221,27 |
| PR | 109 | 0,1396 | 780,80 |
| SC | 102 | 0,0897 | 1137,12 |
| RS | 433 | 0,1438 | 3011,13 |
| MS | 62 | – | – |
| MT | 31 | 0,1454 | 213,20 |
| GO | 382 | 0,1726 | 2213,21 |
| DF | 15 | 0,1508 | 99,47 |

States: Acre (AC), Alagoas (AL), Amapá (AP), Amazonas (AM), Bahia (BA), Ceará (CE), Distrito Federal (DF), Espírito Santo (ES), Goiás (GO), Maranhão (MA), Mato Grosso (MT), Mato Grosso do Sul (MS), Minas Gerais (MG), Pará (PA), Paraíba (PB), Paraná (PR), Pernambuco (PE), Piauí (PI), Rio de Janeiro (RJ), Rio Grande do Norte (RN), Rio Grande do Sul (RS), Rondônia (RO), Roraima (RR), Santa Catarina (SC), São Paulo (SP), Sergipe (SE), Tocantins (TO).

**Table S11:** Coverage and Richness estimated for each cluster using Chao 1 (24). Number of different refers to the number of non-redundant sequences.

| Cluster | number of different | Coverage | Richness |
|---|---|---|---|
| CL15-1 | 376 | 0,1501 | 2504,0 |
| CL15-2 | 115 | 0,2713 | 423,9 |
| CL15-3 | 1950 | 0,2785 | 7001,0 |
| CL15-4 | 736 | 0,1870 | 3936,0 |
| CL15-5 | 56 | 0,1686 | 332,1 |
| CL15-6 | 386 | 0,1850 | 2086,0 |
| CL15-7 | 575 | 0,1981 | 2903,0 |
| CL15-8 | 861 | 0,1903 | 4525,0 |
| CL15-9 | 42 | 0,2915 | 144,1 |
| CL15-10 | 470 | 0,3006 | 1563,0 |
| CL15-11 | 179 | 0,3367 | 531,7 |
| CL15-12 | 68 | 0,2124 | 320,1 |
| CL15-13 | 47 | 0,2556 | 183,9 |
| CL15-14 | 47 | 0,4085 | 115,1 |
| CL15-15 | 238 | 0,2073 | 1148,0 |
| Total | 6146 | | 27 716,9 |

**Table S12:** Total samples taken by state in the years 2020 and 2021 (GISAID release 609), compared to the total number of quality samples. The second column is the total population of the state in millions, according to the IBGE – Instituto Brasileiro de Geografia e Estatística https://www.ibge.gov.br/cidades-e-estados. There is a marked imbalance in the sampling between Brazilian states.

| | population (millions) | 2020 | 2021 | with quality (2020/21) |
|---|---|---|---|---|
| AC | 0,894 | 26 | 2 | 11 |
| AL | 3,351 | 23 | 80 | 100 |
| AP | 0,861 | 39 | 45 | 72 |
| AM | 4,207 | 417 | 132 | 309 |
| BA | 14,930 | 126 | 152 | 211 |
| CE | 9,187 | 50 | 29 | 34 |
| ES | 4,064 | 32 | 38 | 55 |
| GO | 7,113 | 86 | 757 | 489 |
| MA | 7,114 | 43 | 121 | 131 |
| MT | 3,526 | 19 | 4 | 2 |
| MS | 2,809 | 62 | 31 | 71 |
| MG | 21,292 | 161 | 152 | 114 |
| PA | 8,690 | 110 | 66 | 147 |
| PB | 4,039 | 125 | 69 | 171 |
| PR | 11,516 | 162 | 79 | 142 |
| PE | 9,616 | 190 | 17 | 70 |
| PI | 3,281 | 9 | 10 | 11 |
| RJ | 17,366 | 704 | 1354 | 1170 |
| RN | 3,534 | 29 | 69 | 68 |
| RS | 11,422 | 691 | 263 | 556 |
| RO | 1,796 | 8 | 26 | 31 |
| RR | 0,631 | 6 | 23 | 14 |
| SC | 7,252 | 26 | 161 | 151 |
| SP | 46,289 | 1176 | 5143 | 4386 |
| SE | 2,318 | 28 | 142 | 157 |
| TO | 1,590 | 15 | 12 | 15 |
| DF | 3,055 | 27 | 4 | 21 |

**Table S13:** Comparison of clusters composition between the PANGO nomenclature v3.0.5 from 2021-06-04 and v3.1.11 from 2021-08-09.

| | v3.0.5 (2021-06-04) | | | | | v3.1.11 (2021-08-09) | | | | |
|---|---|---|---|---|---|---|---|---|---|---|
| cluster 1 | B.1.1 | B.1.1.28 | B.1.1.332 | | | B.1.1 | B.1.1.28 | B.1.1.332 | B.1.1.448 | B.1.1.464 |
| | 2 | 467 | 8 | | | 6 | 453 | 8 | 1 | 1 |
| | | | | | | P.1 | P.2 | P.7 | | |
| | | | | | | 1 | 3 | 4 | | |
| cluster 2 | A.1 | A.2 | B | B.1 | B.1.111 | A.1 | A.2 | B | B.1 | B.1.111 |
| | 1 | 3 | 3 | 59 | 5 | 1 | 3 | 3 | 58 | 5 |
| | B.1.160.25 | B.1.177.32 | B.1.177.52 | B.1.195 | B.1.212 | B.1.160.25 | B.1.177.32 | B.1.177.52 | B.1.195 | B.1.212 |
| | 1 | 1 | 1 | 54 | 15 | 1 | 1 | 1 | 54 | 15 |
| | B.1.234 | B.1.240 | B.1.319 | B.1.351 | B.1.547 | B.1.234 | B.1.240 | B.1.351 | B.1.479 | B.1.547 |
| | 2 | 1 | 1 | 5 | 1 | 3 | 1 | 5 | 1 | 1 |
| | B.1.575 | B.1.617.2 | B.1.9 | B.1.91 | B.3 | B.1.617.2 | B.1.623 | B.1.9 | B.1.91 | B.3 |
| | 1 | 1 | 1 | 15 | 2 | 1 | 1 | 1 | 15 | 2 |
| | B.39 | B.6 | | | | B.39 | B.6 | | | |
| | 5 | 1 | | | | 5 | 1 | | | |
| cluster 3 | B.1 | B.1.1 | B.1.617.1 | P.1 | P.1.1 | P.1 | P.1.1 | P.1.2 | P.1.3 | P.1.7 |
| | 1 | 1 | 2 | 2966 | 18 | 3013 | 19 | 12 | 1 | 17 |
| | P.1.2 | P.2 | P.4 | | | | | | | |
| | 26 | 27 | 21 | | | | | | | |
| cluster 4 | B.1 | B.1.1 | B.1.1.161 | B.1.1.33 | N.1 | B.1 | B.1.1 | B.1.1.161 | B.1.1.277 | B.1.1.33 |
| | 4 | 11 | 7 | 909 | 2 | 5 | 11 | 7 | 1 | 907 |
| | N.10 | N.4 | N.9 | | | N.1 | N.10 | N.4 | N.9 | |
| | 19 | 4 | 81 | | | 1 | 24 | 4 | 77 | |
| cluster 5 | B.1.1.28 | | | | | P.7 | | | | |
| | 77 | | | | | 77 | | | | |
| cluster 6 | B.1 | B.1.1 | B.1.1.192 | B.1.1.277 | B.1.1.28 | B.1 | B.1.1 | B.1.1.192 | B.1.1.277 | B.1.1.28 |
| | 1 | 171 | 1 | 3 | 317 | 1 | 158 | 1 | 3 | 300 |
| | B.1.1.372 | B.1.1.378 | B.1.1.462 | B.1.1.519 | B.1.1.74 | B.1.1.371 | B.1.1.372 | B.1.1.378 | B.1.1.462 | B.1.1.515 |
| | 1 | 29 | 1 | 2 | 1 | 3 | 1 | 29 | 1 | 1 |
| | B.59 | C.14 | C.37 | | | B.1.1.519 | B.1.1.74 | B.59 | B.60 | C.14 |
| | 1 | 1 | 2 | | | 2 | 1 | 1 | 7 | 1 |
| | | | | | | C.37 | P.5 | | | |
| | | | | | | 2 | 19 | | | |
| cluster 7 | P.1 | P.1.1 | P.1.2 | P.2 | P.4 | P.1 | P.1.1 | P.1.2 | P.1.7 | |
| | 4 | 4 | 8 | 6 | 732 | 742 | 4 | 3 | 5 | |
| cluster 8 | B.1.1.28 | P.2 | | | | B.1.1 | B.1.1.28 | P.2 | | |
| | 1 | 1089 | | | | 2 | 14 | 1074 | | |
| cluster 9 | P.1 | P.1.2 | P.4 | | | P.1 | P.1.2 | P.1.7 | | |
| | 31 | 4 | 15 | | | 46 | 1 | 3 | | |
| cluster 10 | P.1 | P.1.1 | P.1.2 | P.2 | P.4 | P.1 | P.1.1 | P.1.2 | P.1.3 | |
| | 491 | 2 | 3 | 14 | 155 | 659 | 2 | 3 | 1 | |
| cluster 11 | B.1.1 | B.1.1.7 | P.1 | | | B.1.1 | B.1.1.7 | None | | |
| | 1 | 248 | 1 | | | 2 | 242 | 6 | | |
| cluster 12 | P.1 | P.1.2 | | | | P.1 | P.1.2 | | | |
| | 4 | 100 | | | | 4 | 100 | | | |
| cluster 13 | P.4 | | | | | P.4 | | | | |
| | 79 | | | | | 79 | | | | |
| cluster 14 | B.1.1.28 | P.1 | P.4 | | | P.1 | | | | |
| | 2 | 74 | 5 | | | 81 | | | | |
| cluster 15 | P.1 | P.1.1 | P.1.2 | P.4 | | P.1 | P.1.1 | P.1.7 | | |
| | 1 | 4 | 12 | 267 | | 268 | 4 | 12 | | |
