## Supplementary3_tSNE_by_week for "Genomic landscape of SARS-CoV-2 pandemic in Brazil suggests an external P.1 variant origin"

November 10, 2021

1. Laboratory of Artificial Intelligence Applied to Bioinformatics - SEPT, Federal University of Paraná, Curtiba, Paraná, Brazil
2. Graduate Program in Bioinformatics - SEPT, Federal University of Paraná, Curtiba, Paraná, Brazil
3. Department of Biochemistry and Molecular Biology, Federal University of Paraná, Curtiba, Paraná, Brazil

**Corresponding Author:**
Roberto T. Raittz:

Lineage evolution over time through visualization with t-SNE graph. Each slide adds the samples of a new week. The colors represent each cluster obtained for Brazilian sequences. Here was adopted the nomenclature PANGO v3.0.5. The key shows the main lineages that each cluster represents. The emergence, growth and extinction of the groups is visible, as well as the gradual replacement of the T0 group variants by the TP1 group.

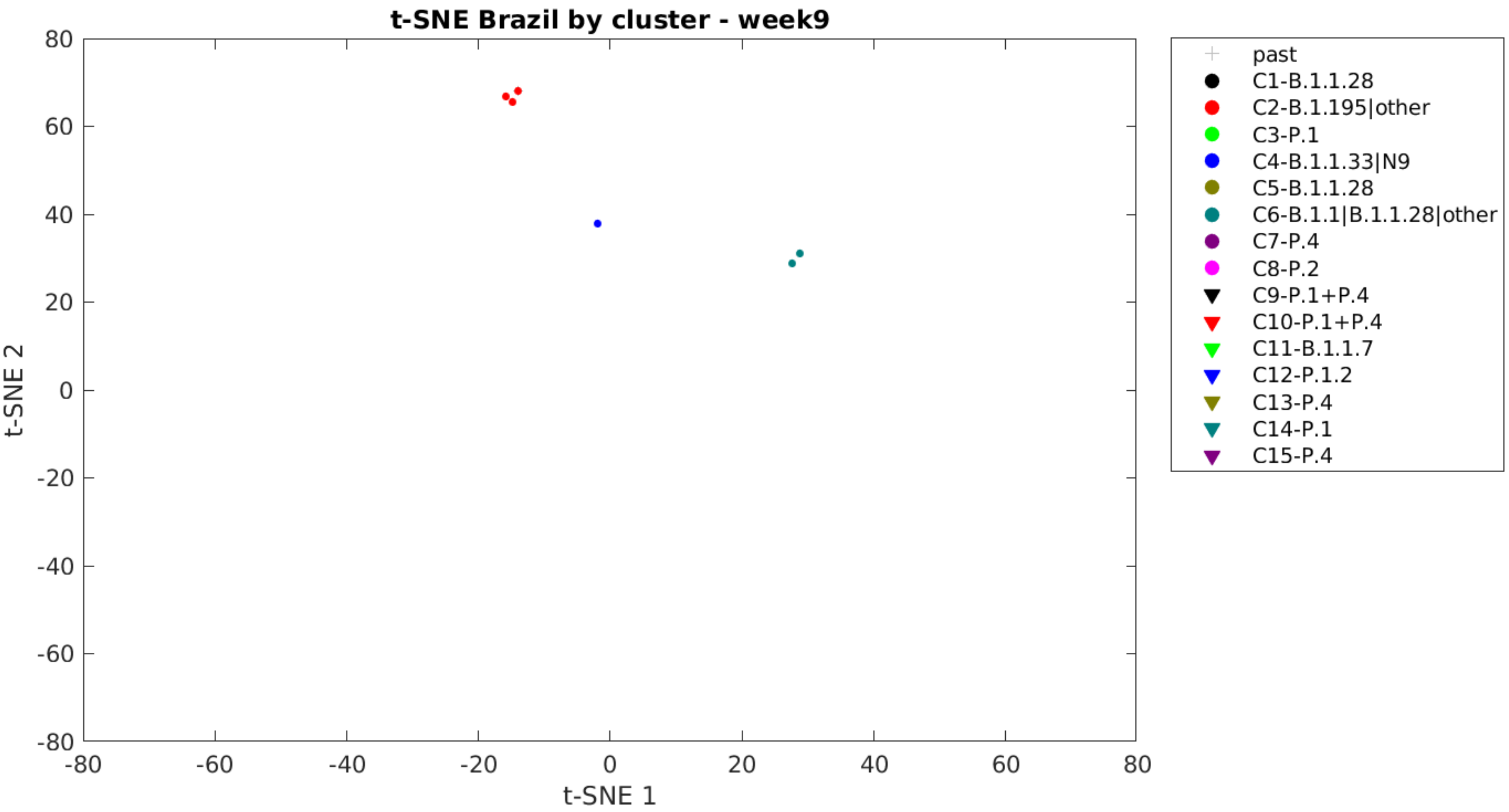
t-SNE Brazil by cluster - week9
t-SNE 2
t-SNE 1
80
60
40
20
0
-20
-40
-60
-80
-80
-60
-40
-20
0
20
40
60
80
past
C1-B.1.1.28
C2-B.1.195|other
C3-P.1
C4-B.1.1.33|N9
C5-B.1.1.28
C6-B.1.1|B.1.1.28|other
C7-P.4
C8-P.2
C9-P.1+P.4
C10-P.1+P.4
C11-B.1.1.7
C12-P.1.2
C13-P.4
C14-P.1
C15-P.4

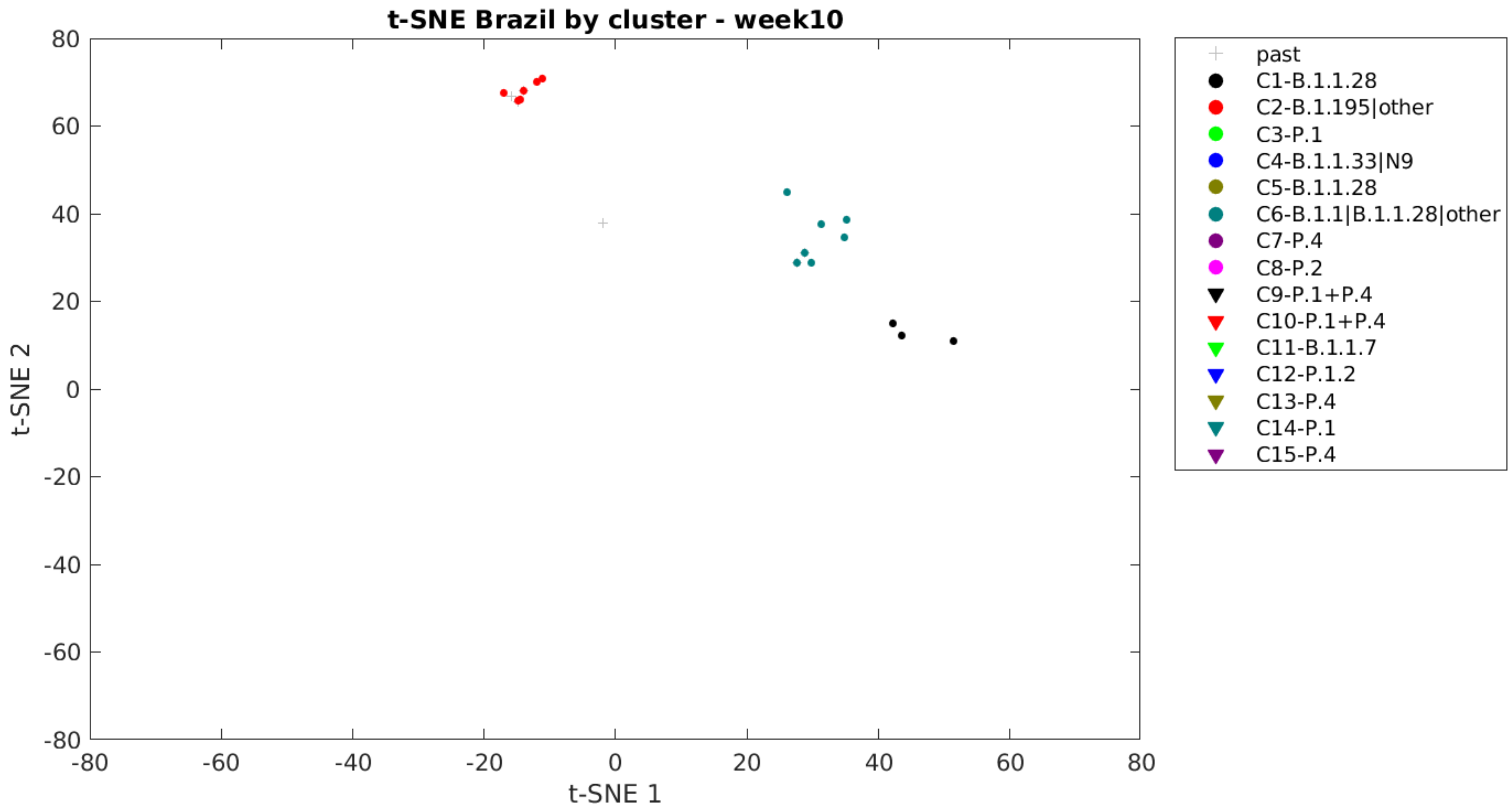
t-SNE Brazil by cluster - week10
t-SNE 1
t-SNE 2
80
60
40
20
0
-20
-40
-60
-80
-80
-60
-40
-20
0
20
40
60
80
past
C1-B.1.1.28
C2-B.1.195|other
C3-P.1
C4-B.1.1.33|N9
C5-B.1.1.28
C6-B.1.1|B.1.1.28|other
C7-P.4
C8-P.2
C9-P.1+P.4
C10-P.1+P.4
C11-B.1.1.7
C12-P.1.2
C13-P.4
C14-P.1
C15-P.4

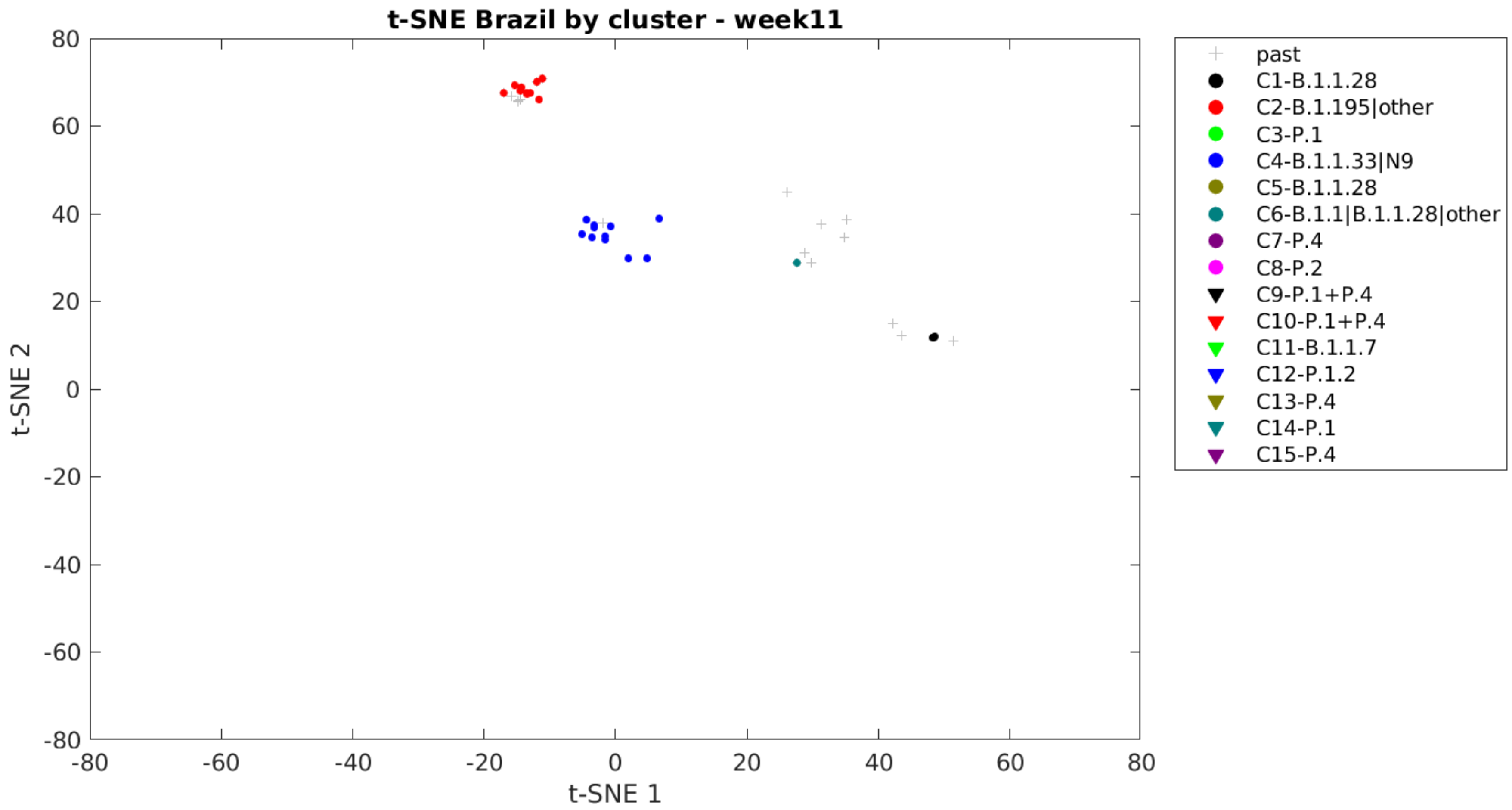
t-SNE Brazil by cluster - week11
t-SNE 2
t-SNE 1
80
60
40
20
0
-20
-40
-60
-80
-80
-60
-40
-20
0
20
40
60
80
past
C1-B.1.1.28
C2-B.1.195|other
C3-P.1
C4-B.1.1.33|N9
C5-B.1.1.28
C6-B.1.1|B.1.1.28|other
C7-P.4
C8-P.2
C9-P.1+P.4
C10-P.1+P.4
C11-B.1.1.7
C12-P.1.2
C13-P.4
C14-P.1
C15-P.4

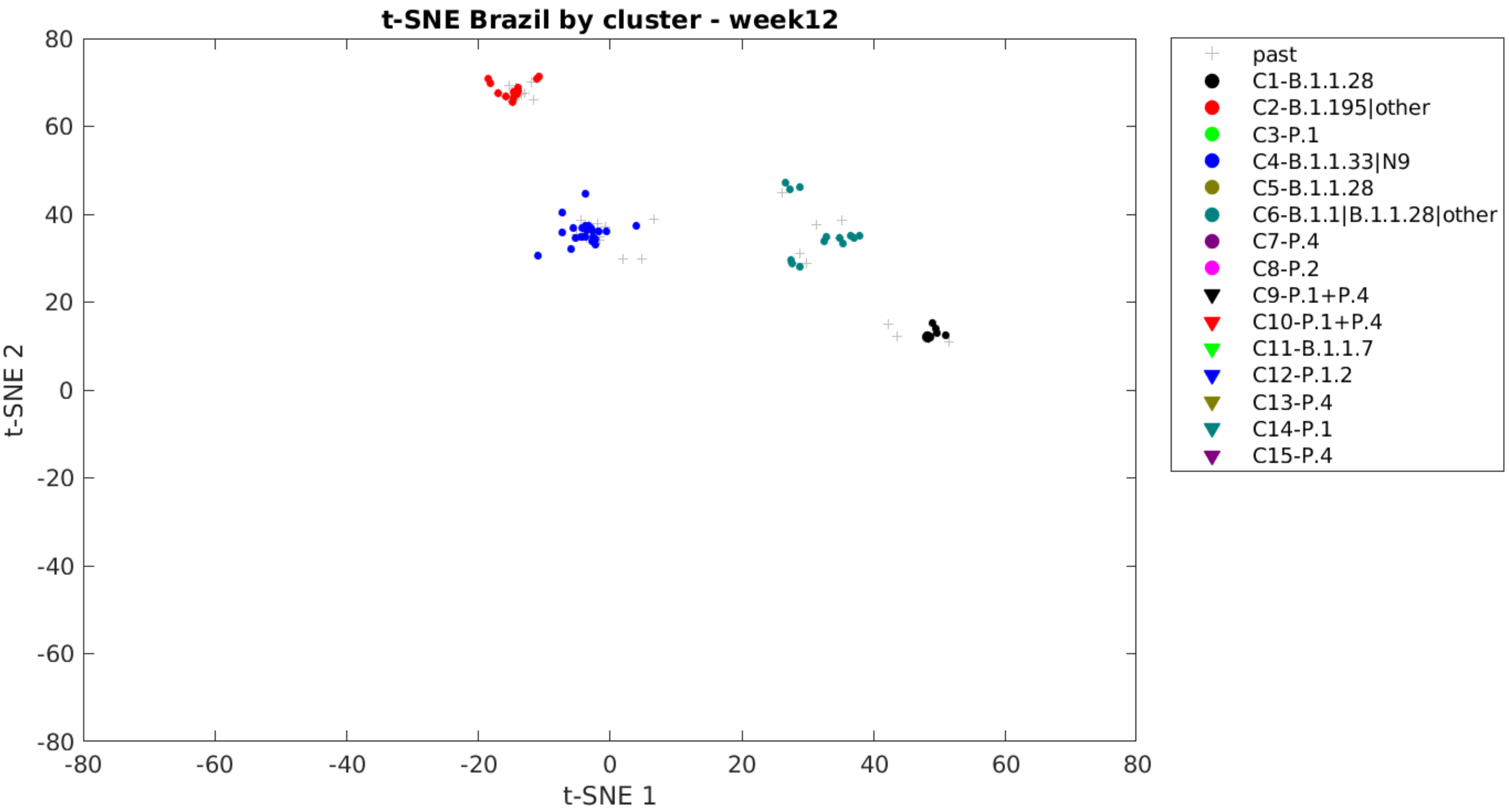
t-SNE Brazil by cluster - week12
t-SNE 2
t-SNE 1
80
60
40
20
0
-20
-40
-60
-80
-80
-60
-40
-20
0
20
40
60
80
past
C1-B.1.1.28
C2-B.1.195|other
C3-P.1
C4-B.1.1.33|N9
C5-B.1.1.28
C6-B.1.1|B.1.1.28|other
C7-P.4
C8-P.2
C9-P.1+P.4
C10-P.1+P.4
C11-B.1.1.7
C12-P.1.2
C13-P.4
C14-P.1
C15-P.4

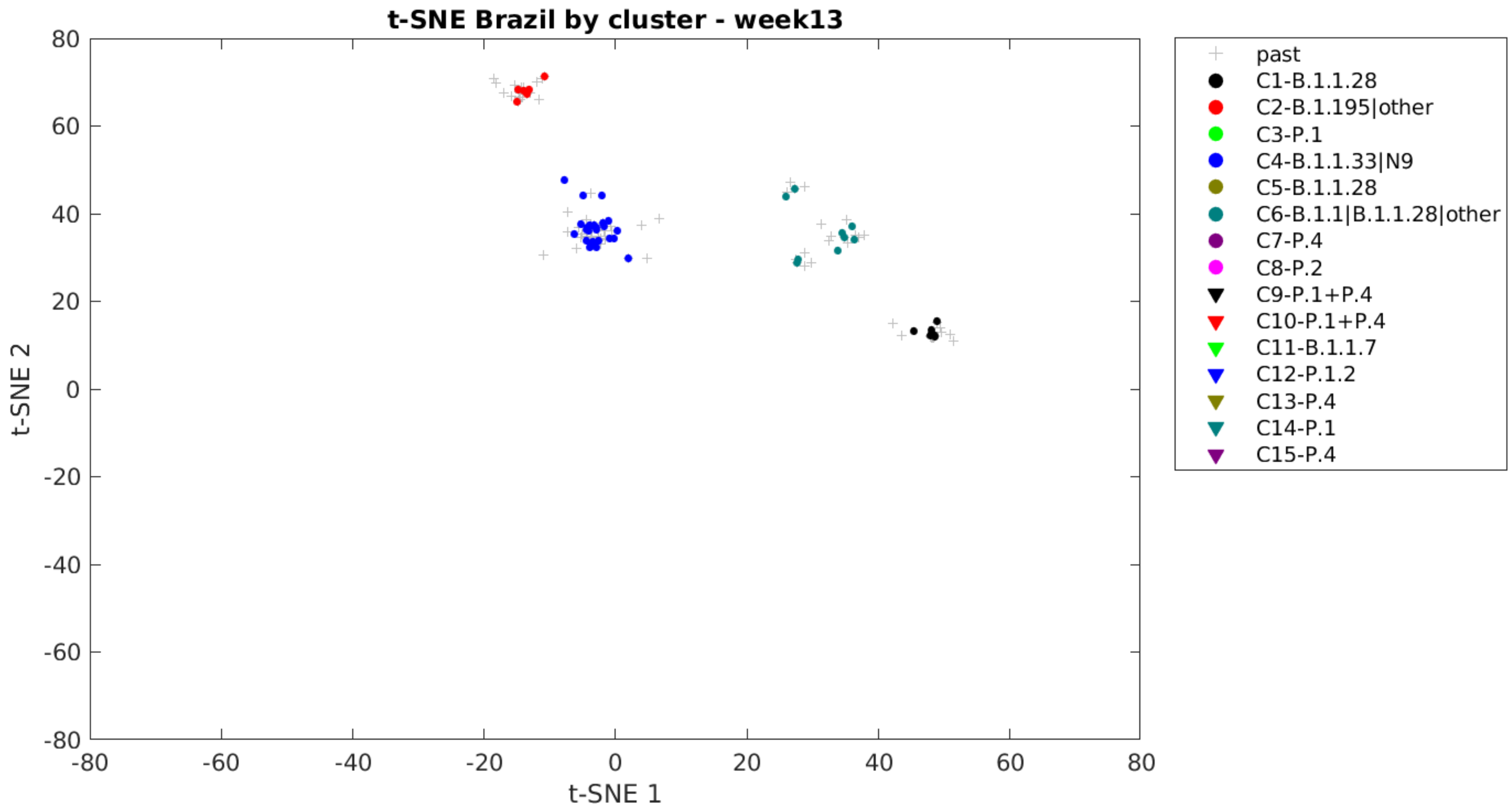

t-SNE Brazil by cluster - week13
t-SNE 1
t-SNE 2
80
60
40
20
0
-20
-40
-60
-80
-80
-60
-40
-20
0
20
40
60
80
past
C1-B.1.1.28
C2-B.1.195|other
C3-P.1
C4-B.1.1.33|N9
C5-B.1.1.28
C6-B.1.1|B.1.1.28|other
C7-P.4
C8-P.2
C9-P.1+P.4
C10-P.1+P.4
C11-B.1.1.7
C12-P.1.2
C13-P.4
C14-P.1
C15-P.4

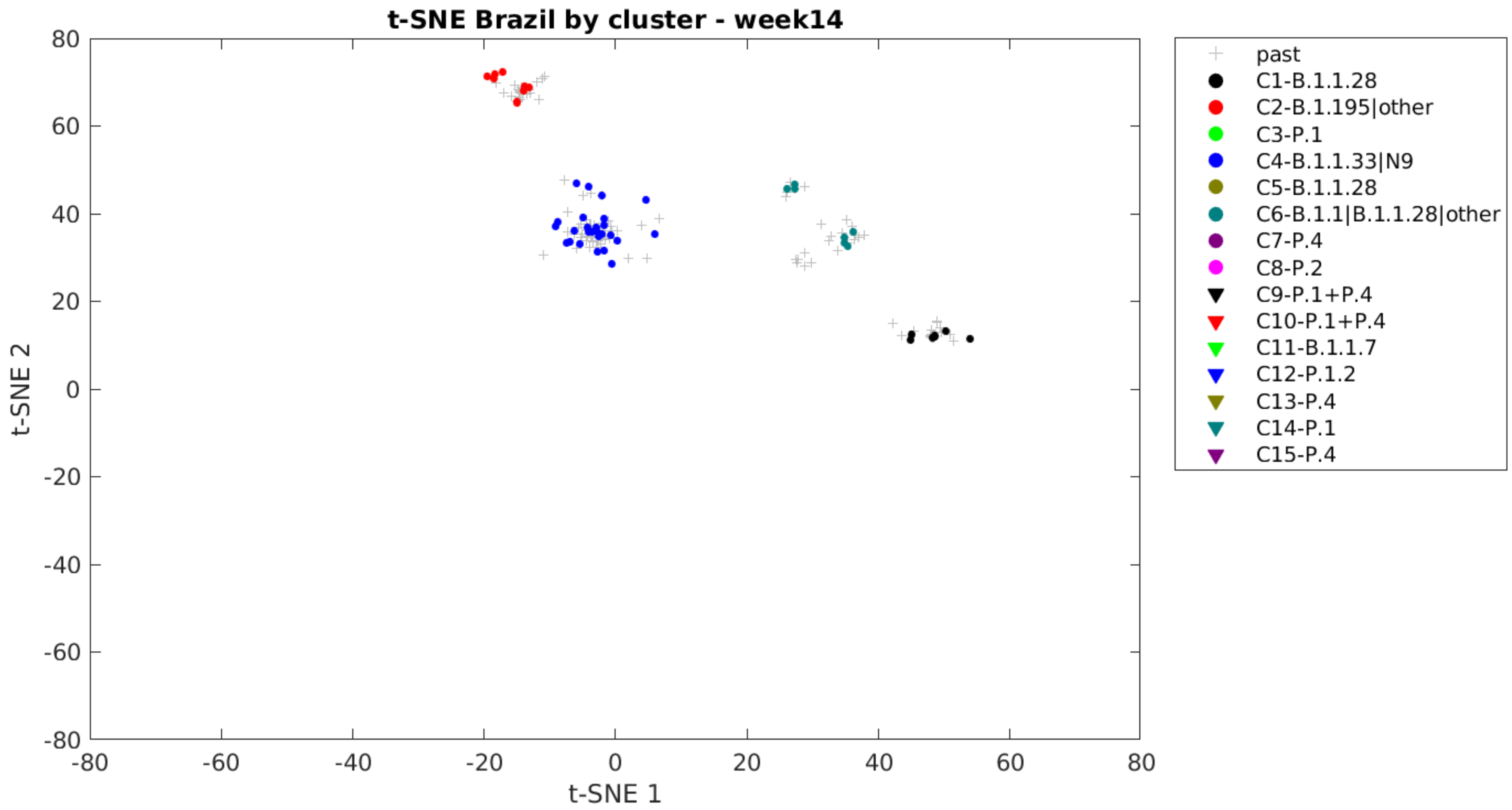

t-SNE Brazil by cluster - week14
t-SNE 2
t-SNE 1
80
60
40
20
0
-20
-40
-60
-80
-80
-60
-40
-20
0
20
40
60
80
past
C1-B.1.1.28
C2-B.1.195|other
C3-P.1
C4-B.1.1.33|N9
C5-B.1.1.28
C6-B.1.1|B.1.1.28|other
C7-P.4
C8-P.2
C9-P.1+P.4
C10-P.1+P.4
C11-B.1.1.7
C12-P.1.2
C13-P.4
C14-P.1
C15-P.4

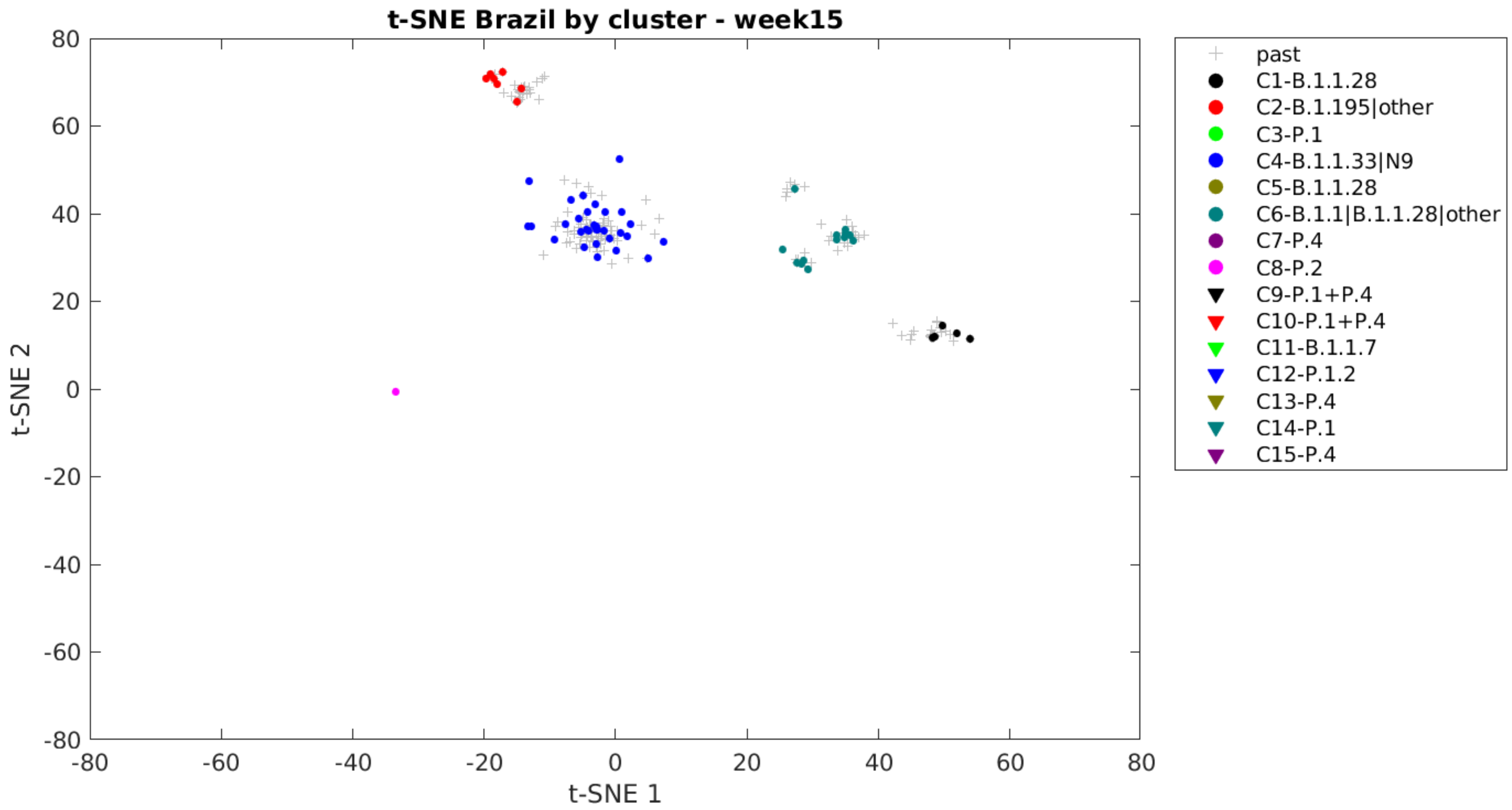
t-SNE Brazil by cluster - week15
80
60
40
20
0
-20
-40
-60
-80
-80
-60
-40
-20
0
20
40
60
80
t-SNE 1
t-SNE 2
past
C1-B.1.1.28
C2-B.1.195|other
C3-P.1
C4-B.1.1.33|N9
C5-B.1.1.28
C6-B.1.1|B.1.1.28|other
C7-P.4
C8-P.2
C9-P.1+P.4
C10-P.1+P.4
C11-B.1.1.7
C12-P.1.2
C13-P.4
C14-P.1
C15-P.4

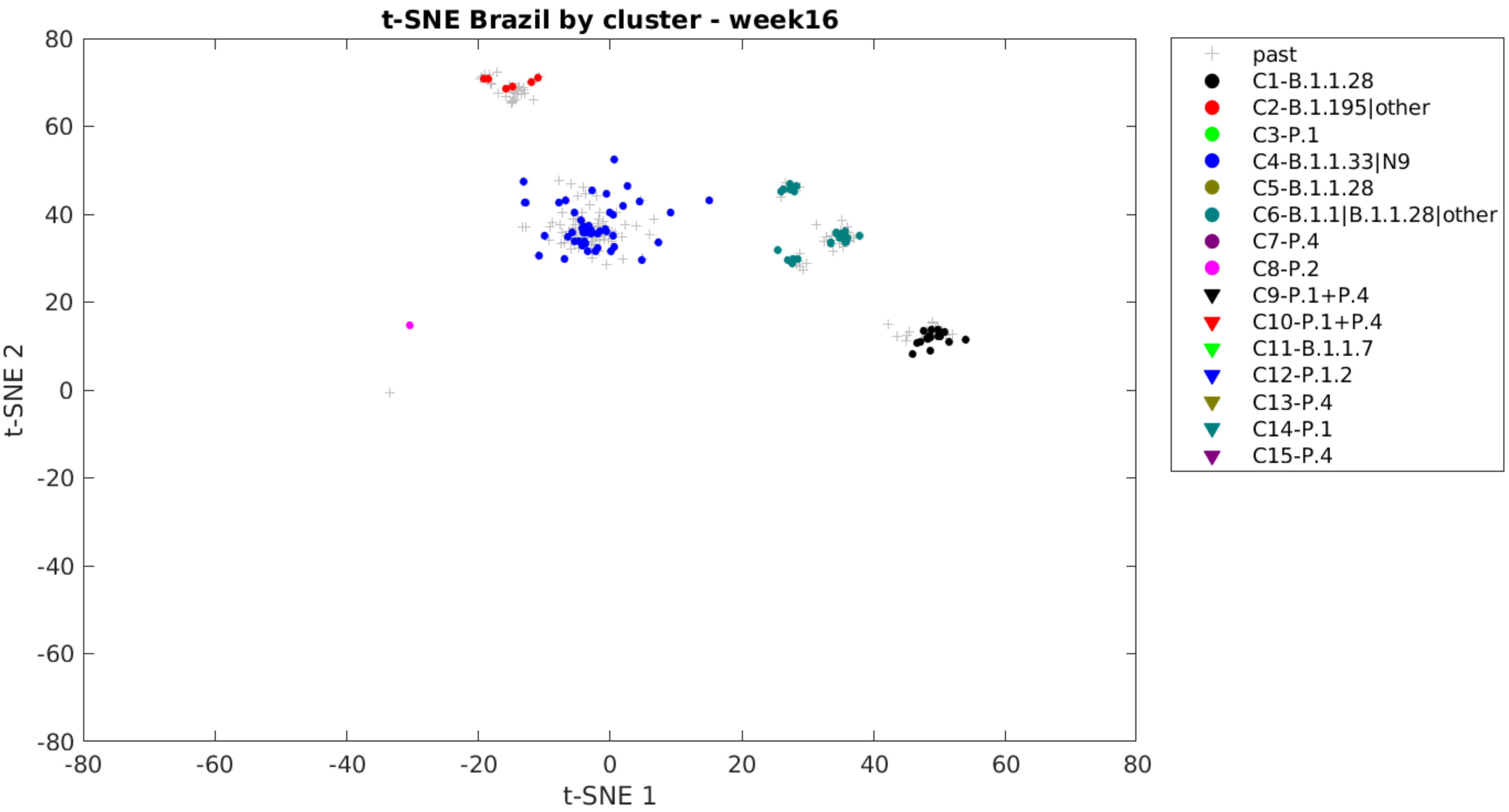

t-SNE Brazil by cluster - week16
80
60
40
20
0
-20
-40
-60
-80
-80
-60
-40
-20
0
20
40
60
80
t-SNE 1
t-SNE 2
past
C1-B.1.1.28
C2-B.1.195|other
C3-P.1
C4-B.1.1.33|N9
C5-B.1.1.28
C6-B.1.1|B.1.1.28|other
C7-P.4
C8-P.2
C9-P.1+P.4
C10-P.1+P.4
C11-B.1.1.7
C12-P.1.2
C13-P.4
C14-P.1
C15-P.4

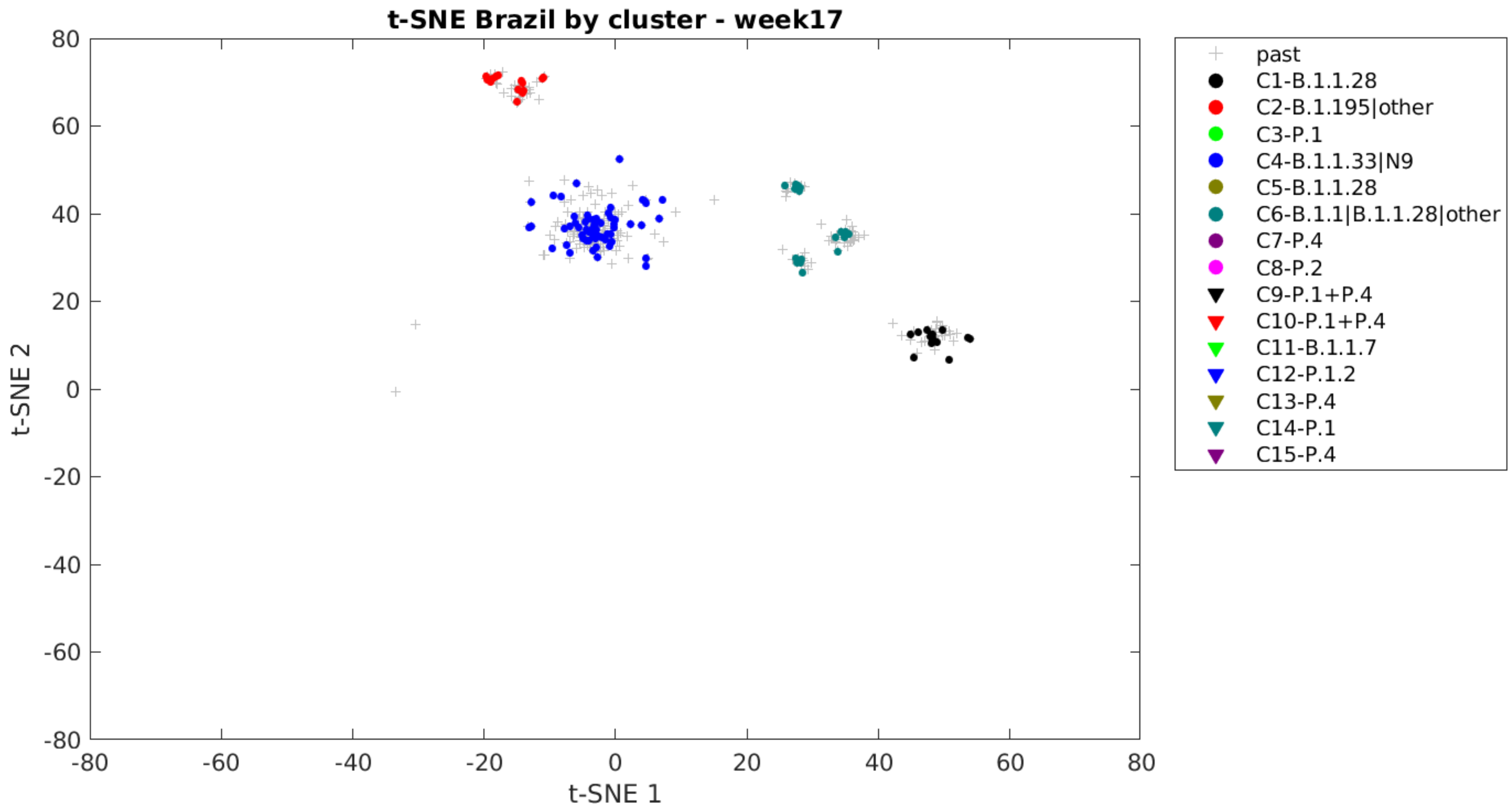
t-SNE Brazil by cluster - week17
80
60
40
20
0
-20
-40
-60
-80
-80
-60
-40
-20
0
20
40
60
80
t-SNE 1
t-SNE 2
past
C1-B.1.1.28
C2-B.1.195|other
C3-P.1
C4-B.1.1.33|N9
C5-B.1.1.28
C6-B.1.1|B.1.1.28|other
C7-P.4
C8-P.2
C9-P.1+P.4
C10-P.1+P.4
C11-B.1.1.7
C12-P.1.2
C13-P.4
C14-P.1
C15-P.4

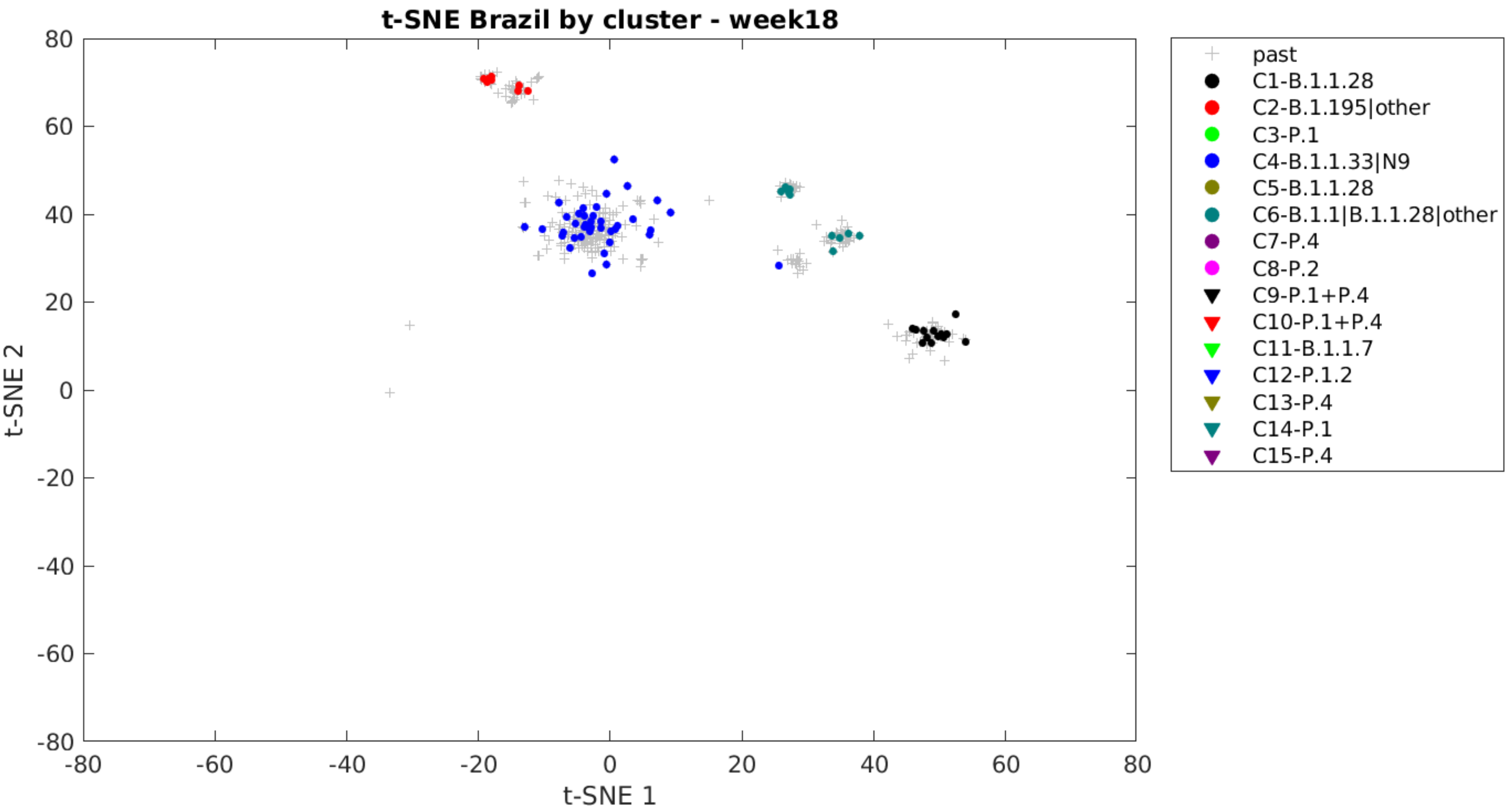

t-SNE Brazil by cluster - week18
80
60
40
20
0
-20
-40
-60
-80
t-SNE 2
-80
-60
-40
-20
0
20
40
60
80
t-SNE 1
past
C1-B.1.1.28
C2-B.1.195|other
C3-P.1
C4-B.1.1.33|N9
C5-B.1.1.28
C6-B.1.1|B.1.1.28|other
C7-P.4
C8-P.2
C9-P.1+P.4
C10-P.1+P.4
C11-B.1.1.7
C12-P.1.2
C13-P.4
C14-P.1
C15-P.4

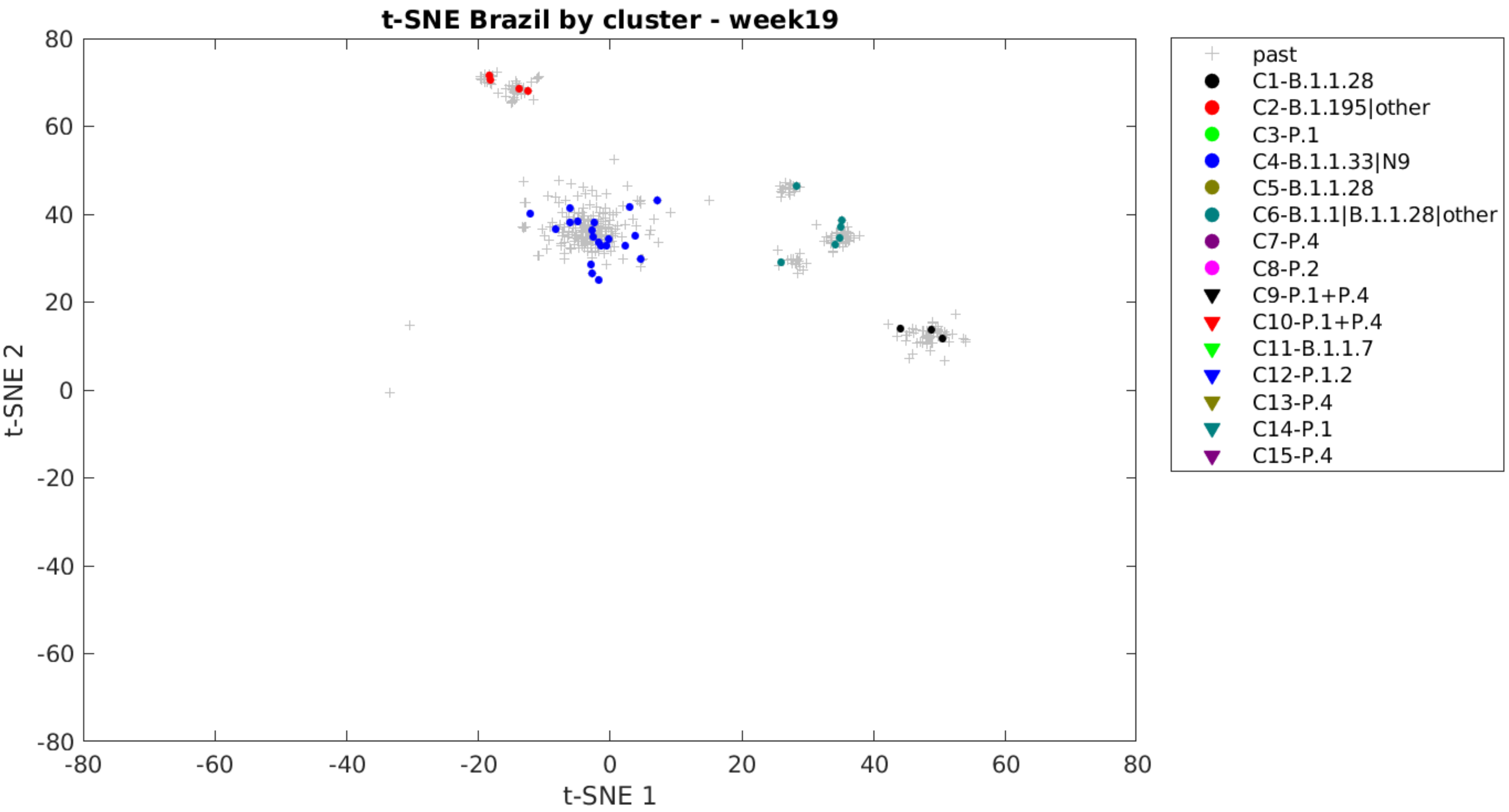

t-SNE Brazil by cluster - week19
t-SNE 2
t-SNE 1
80
60
40
20
0
-20
-40
-60
-80
-80
-60
-40
-20
0
20
40
60
80
past
C1-B.1.1.28
C2-B.1.195|other
C3-P.1
C4-B.1.1.33|N9
C5-B.1.1.28
C6-B.1.1|B.1.1.28|other
C7-P.4
C8-P.2
C9-P.1+P.4
C10-P.1+P.4
C11-B.1.1.7
C12-P.1.2
C13-P.4
C14-P.1
C15-P.4

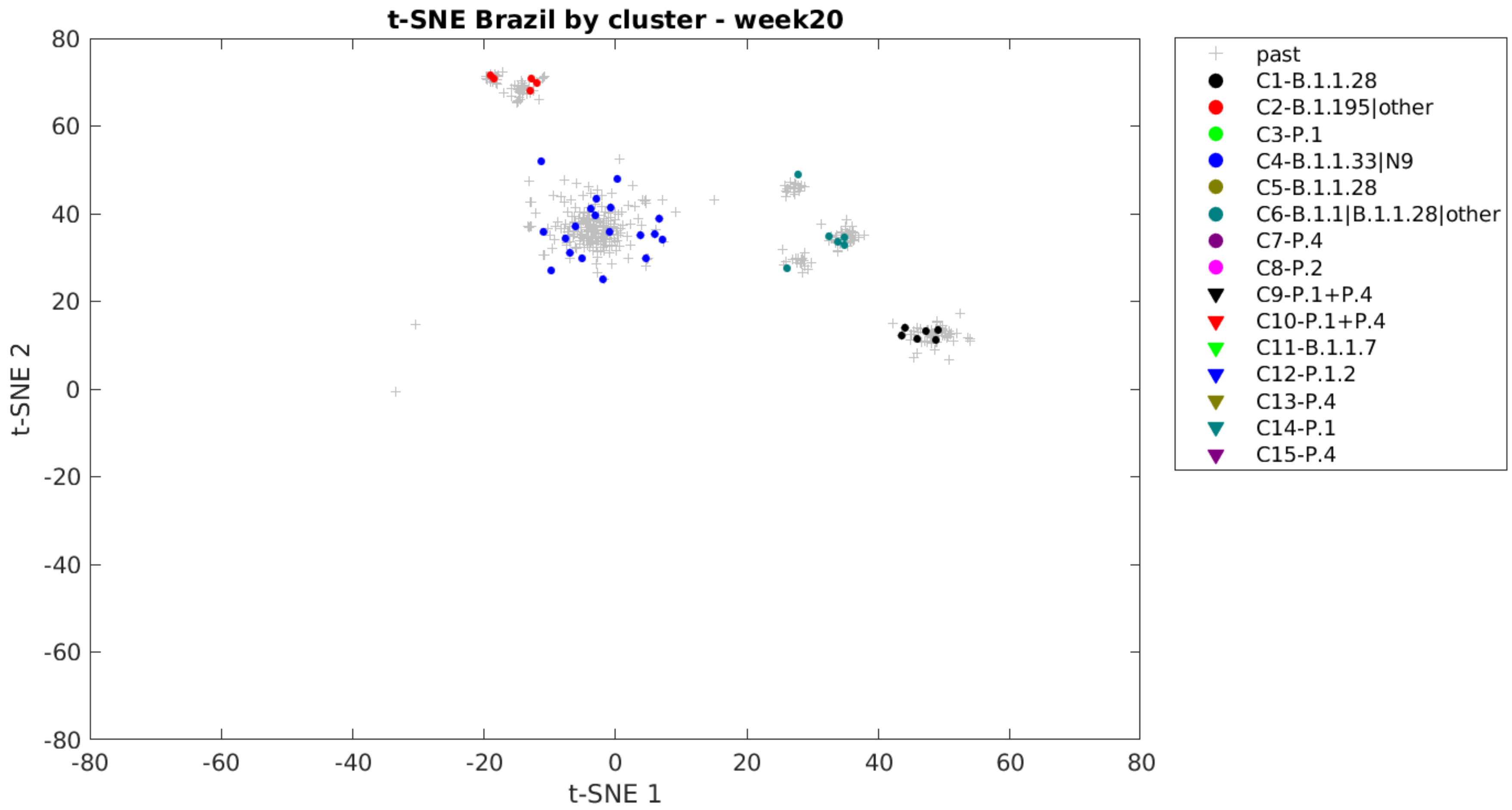

t-SNE Brazil by cluster - week20
t-SNE 2
t-SNE 1
80
60
40
20
0
-20
-40
-60
-80
-80
-60
-40
-20
0
20
40
60
80
past
C1-B.1.1.28
C2-B.1.195|other
C3-P.1
C4-B.1.1.33|N9
C5-B.1.1.28
C6-B.1.1|B.1.1.28|other
C7-P.4
C8-P.2
C9-P.1+P.4
C10-P.1+P.4
C11-B.1.1.7
C12-P.1.2
C13-P.4
C14-P.1
C15-P.4

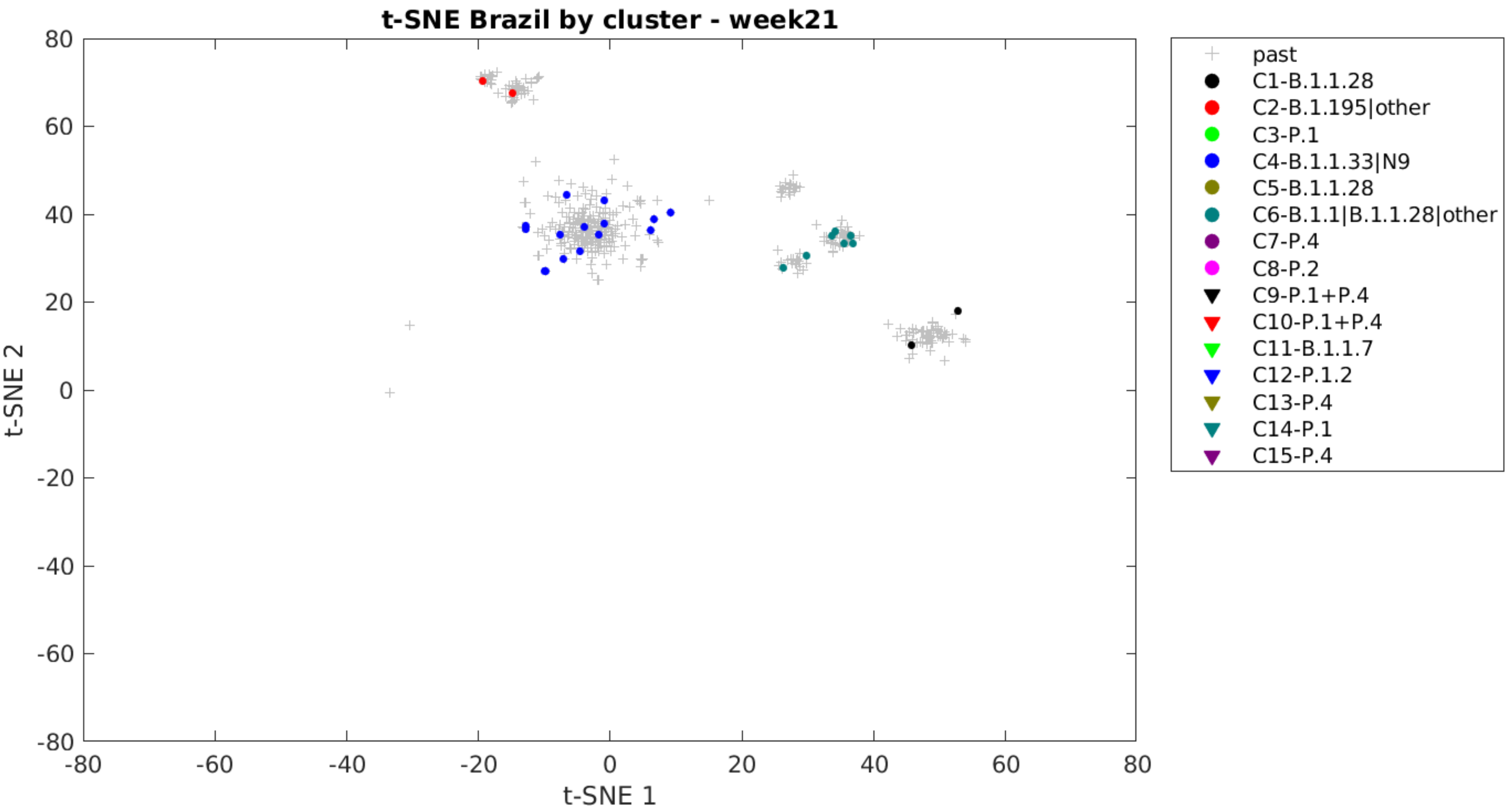
t-SNE Brazil by cluster - week21
t-SNE 1
t-SNE 2
80
60
40
20
0
-20
-40
-60
-80
-80
-60
-40
-20
0
20
40
60
80
past
C1-B.1.1.28
C2-B.1.195|other
C3-P.1
C4-B.1.1.33|N9
C5-B.1.1.28
C6-B.1.1|B.1.1.28|other
C7-P.4
C8-P.2
C9-P.1+P.4
C10-P.1+P.4
C11-B.1.1.7
C12-P.1.2
C13-P.4
C14-P.1
C15-P.4

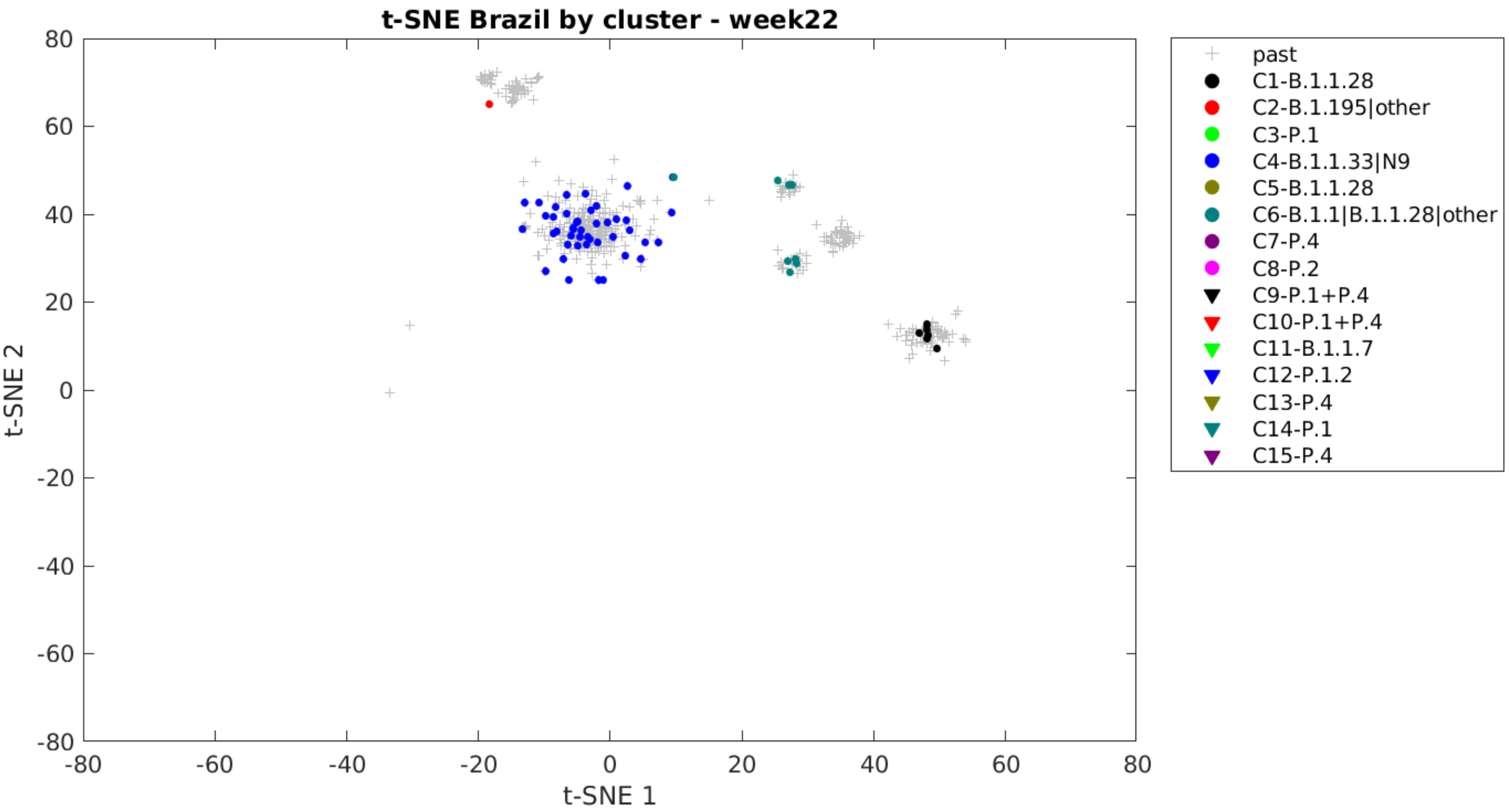

t-SNE Brazil by cluster - week22
t-SNE 1
t-SNE 2
80
60
40
20
0
-20
-40
-60
-80
past
C1-B.1.1.28
C2-B.1.195|other
C3-P.1
C4-B.1.1.33|N9
C5-B.1.1.28
C6-B.1.1|B.1.1.28|other
C7-P.4
C8-P.2
C9-P.1+P.4
C10-P.1+P.4
C11-B.1.1.7
C12-P.1.2
C13-P.4
C14-P.1
C15-P.4

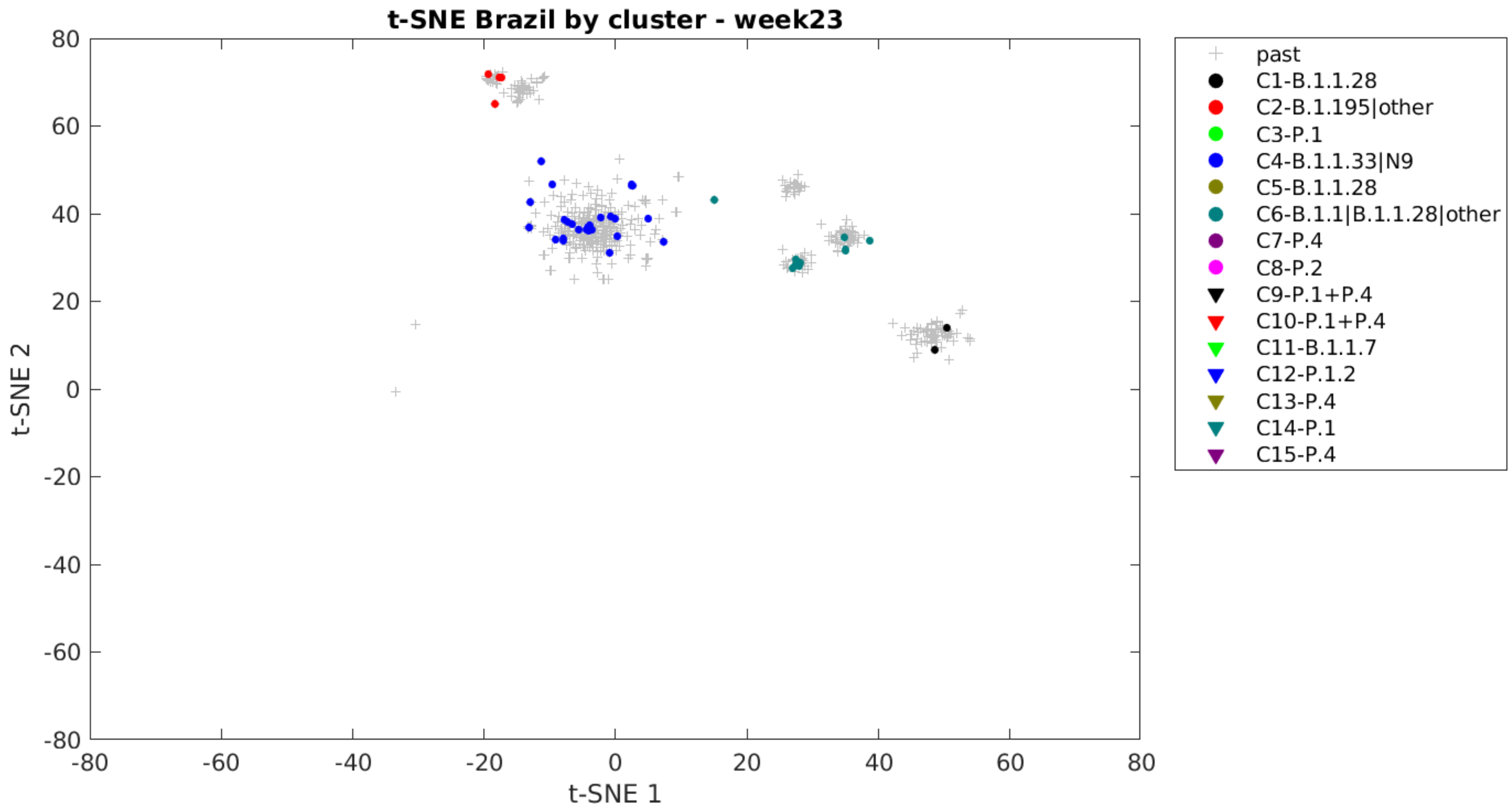
t-SNE Brazil by cluster - week23
t-SNE 1
t-SNE 2
80
60
40
20
0
-20
-40
-60
-80
past
C1-B.1.1.28
C2-B.1.195|other
C3-P.1
C4-B.1.1.33|N9
C5-B.1.1.28
C6-B.1.1|B.1.1.28|other
C7-P.4
C8-P.2
C9-P.1+P.4
C10-P.1+P.4
C11-B.1.1.7
C12-P.1.2
C13-P.4
C14-P.1
C15-P.4

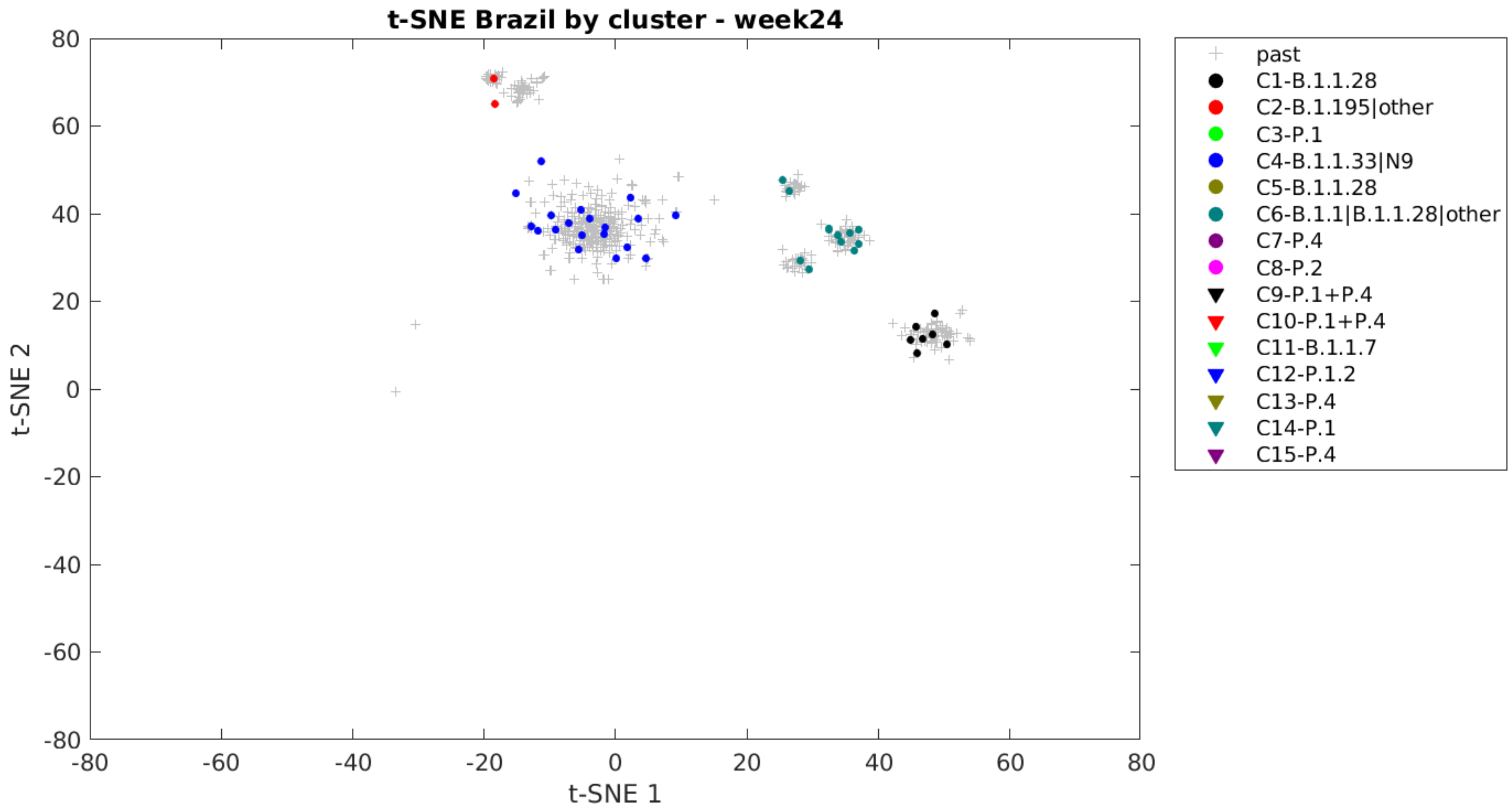
t-SNE Brazil by cluster - week24
t-SNE 1
t-SNE 2
80
60
40
20
0
-20
-40
-60
-80
-80
-60
-40
-20
0
20
40
60
80
past
C1-B.1.1.28
C2-B.1.195|other
C3-P.1
C4-B.1.1.33|N9
C5-B.1.1.28
C6-B.1.1|B.1.1.28|other
C7-P.4
C8-P.2
C9-P.1+P.4
C10-P.1+P.4
C11-B.1.1.7
C12-P.1.2
C13-P.4
C14-P.1
C15-P.4

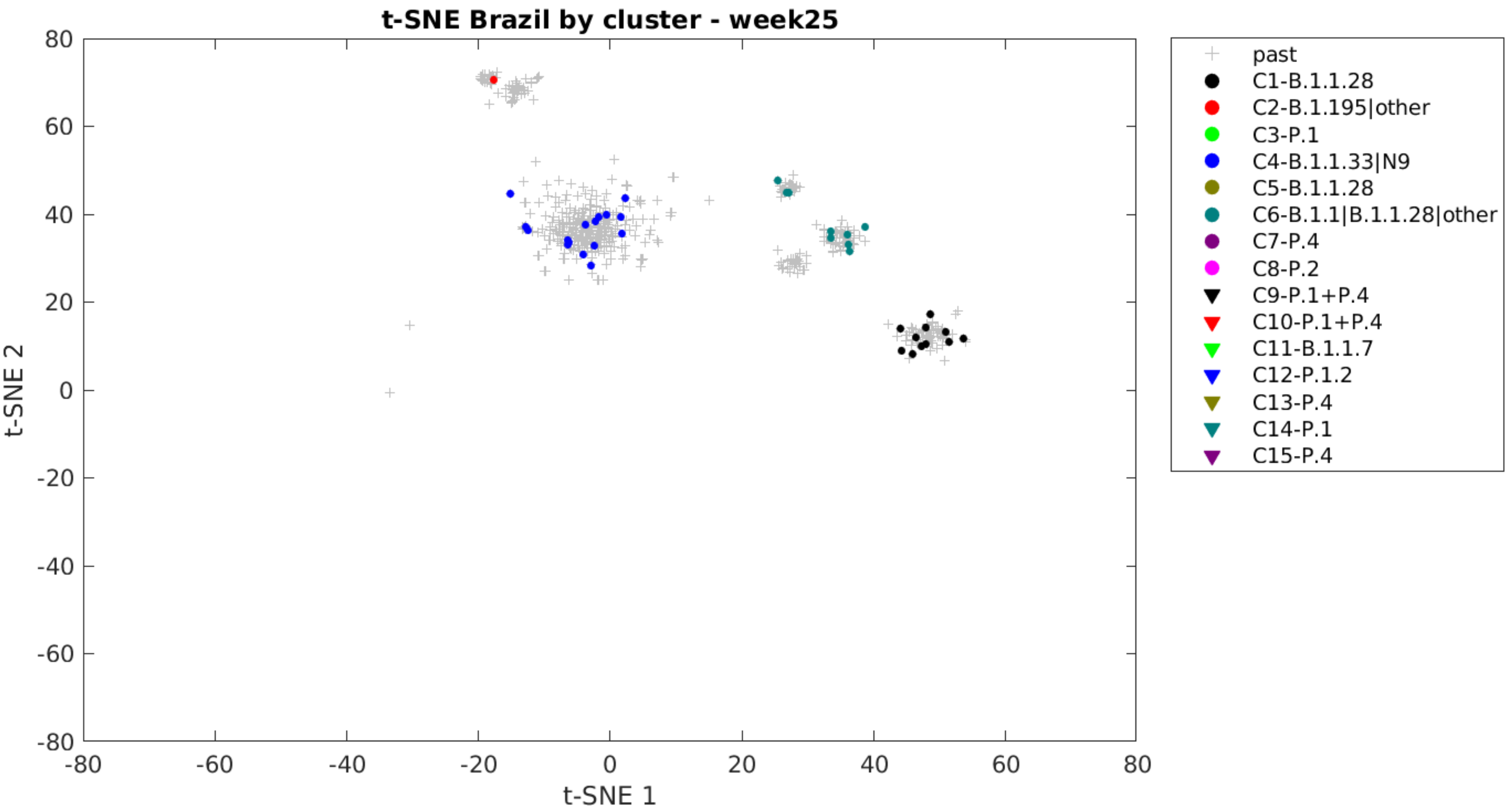
t-SNE Brazil by cluster - week25
t-SNE 1
t-SNE 2
past
C1-B.1.1.28
C2-B.1.195|other
C3-P.1
C4-B.1.1.33|N9
C5-B.1.1.28
C6-B.1.1|B.1.1.28|other
C7-P.4
C8-P.2
C9-P.1+P.4
C10-P.1+P.4
C11-B.1.1.7
C12-P.1.2
C13-P.4
C14-P.1
C15-P.4

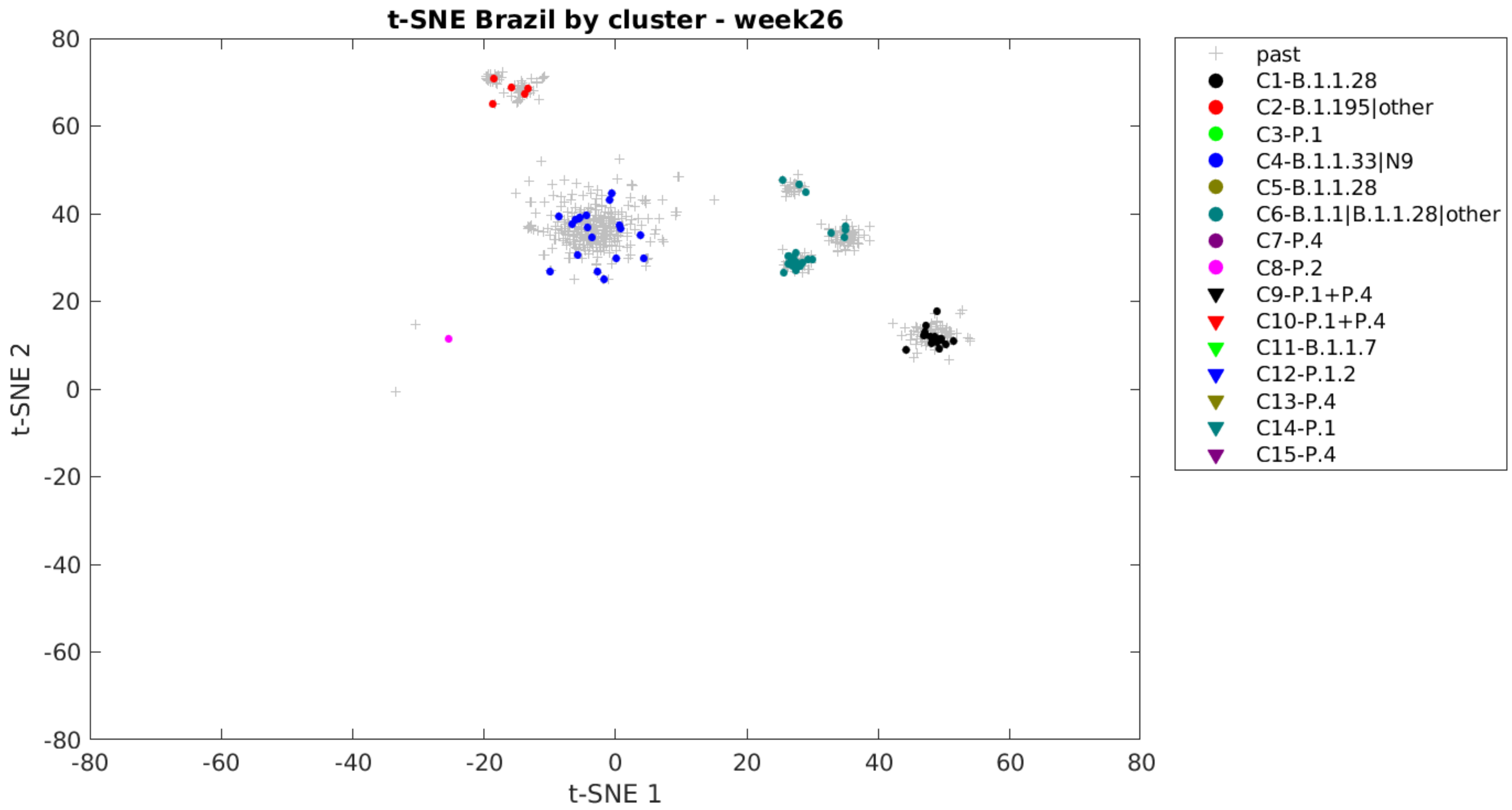

t-SNE Brazil by cluster - week26
t-SNE 1
t-SNE 2
80
60
40
20
0
-20
-40
-60
-80
-80
-60
-40
-20
0
20
40
60
80
past
C1-B.1.1.28
C2-B.1.195|other
C3-P.1
C4-B.1.1.33|N9
C5-B.1.1.28
C6-B.1.1|B.1.1.28|other
C7-P.4
C8-P.2
C9-P.1+P.4
C10-P.1+P.4
C11-B.1.1.7
C12-P.1.2
C13-P.4
C14-P.1
C15-P.4

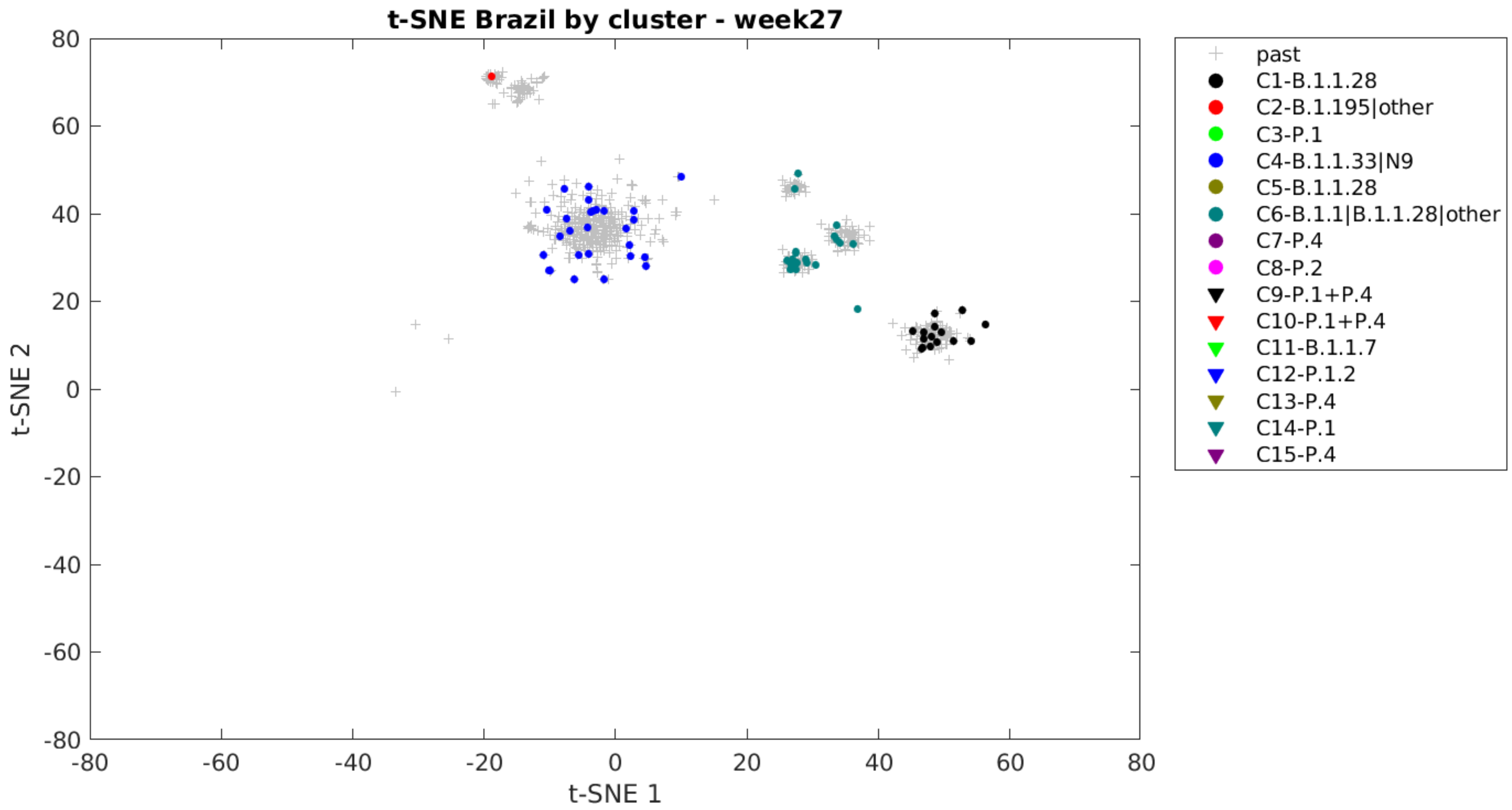

t-SNE Brazil by cluster - week27
80
60
40
20
0
-20
-40
-60
-80
t-SNE 2
-80
-60
-40
-20
0
20
40
60
80
t-SNE 1
past
C1-B.1.1.28
C2-B.1.195|other
C3-P.1
C4-B.1.1.33|N9
C5-B.1.1.28
C6-B.1.1|B.1.1.28|other
C7-P.4
C8-P.2
C9-P.1+P.4
C10-P.1+P.4
C11-B.1.1.7
C12-P.1.2
C13-P.4
C14-P.1
C15-P.4

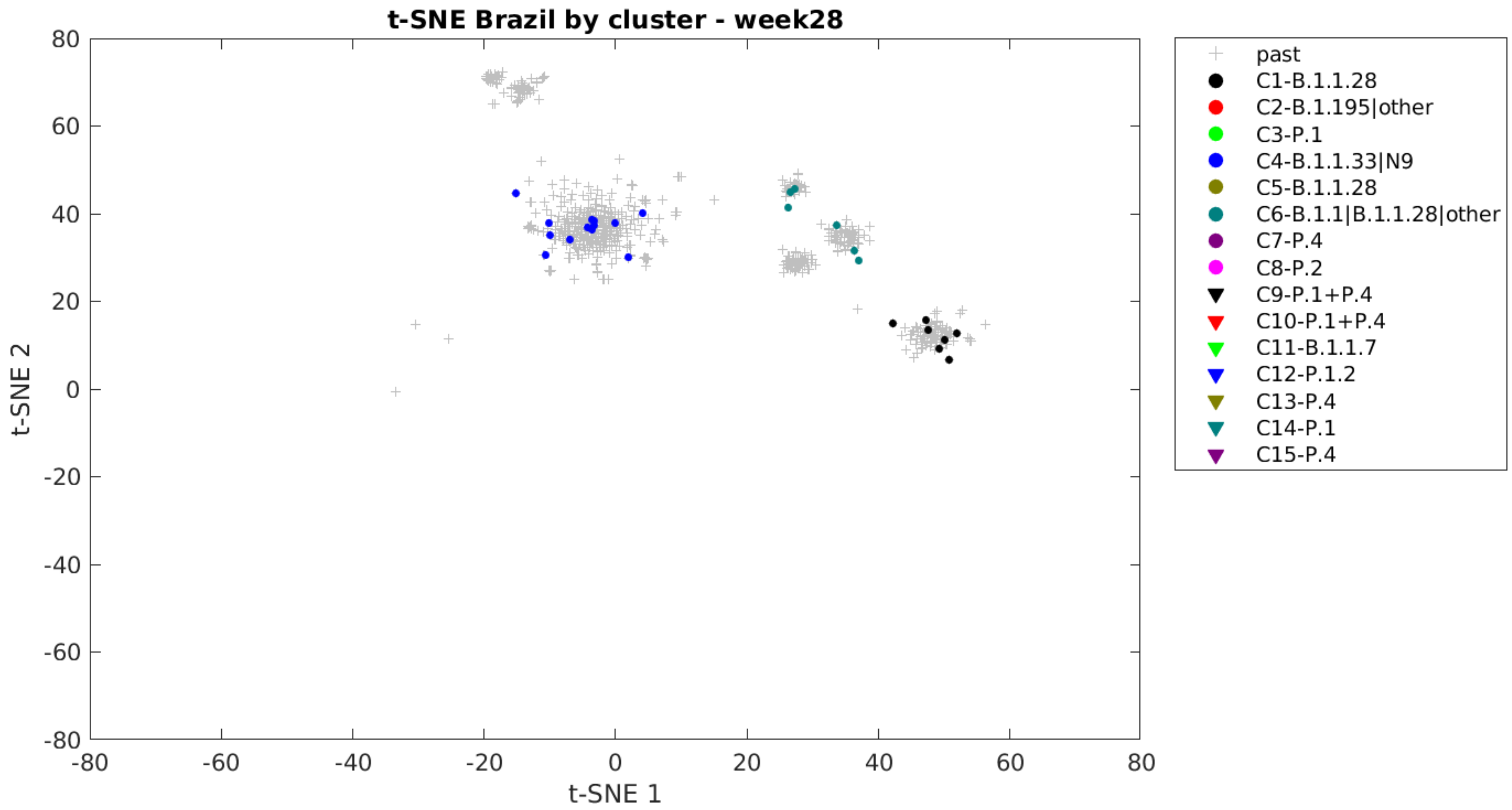
t-SNE Brazil by cluster - week28
t-SNE 1
t-SNE 2
80
60
40
20
0
-20
-40
-60
-80
-80
-60
-40
-20
0
20
40
60
80
past
C1-B.1.1.28
C2-B.1.195|other
C3-P.1
C4-B.1.1.33|N9
C5-B.1.1.28
C6-B.1.1|B.1.1.28|other
C7-P.4
C8-P.2
C9-P.1+P.4
C10-P.1+P.4
C11-B.1.1.7
C12-P.1.2
C13-P.4
C14-P.1
C15-P.4

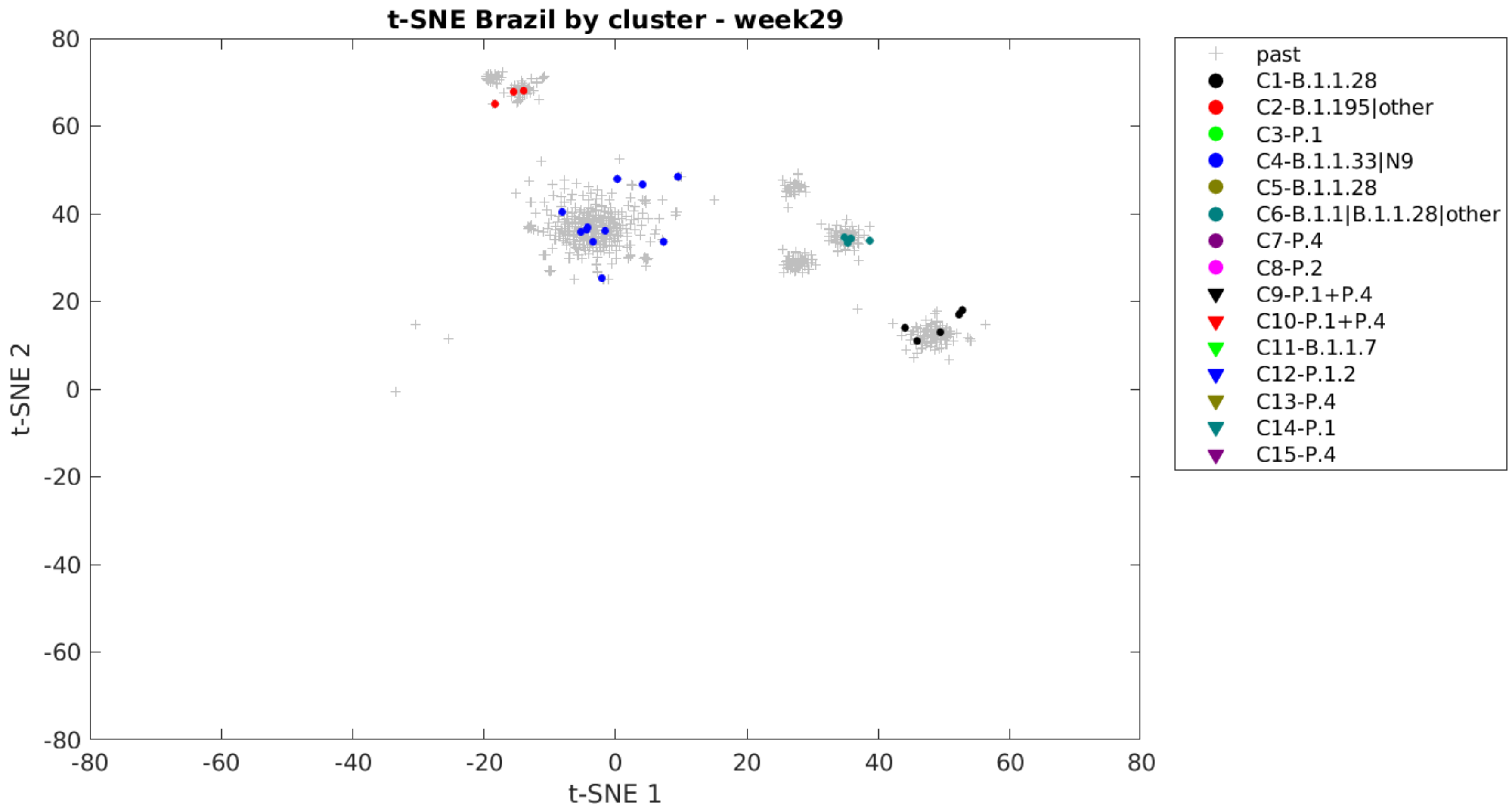
t-SNE Brazil by cluster - week29
t-SNE 1
t-SNE 2
past
C1-B.1.1.28
C2-B.1.195|other
C3-P.1
C4-B.1.1.33|N9
C5-B.1.1.28
C6-B.1.1|B.1.1.28|other
C7-P.4
C8-P.2
C9-P.1+P.4
C10-P.1+P.4
C11-B.1.1.7
C12-P.1.2
C13-P.4
C14-P.1
C15-P.4

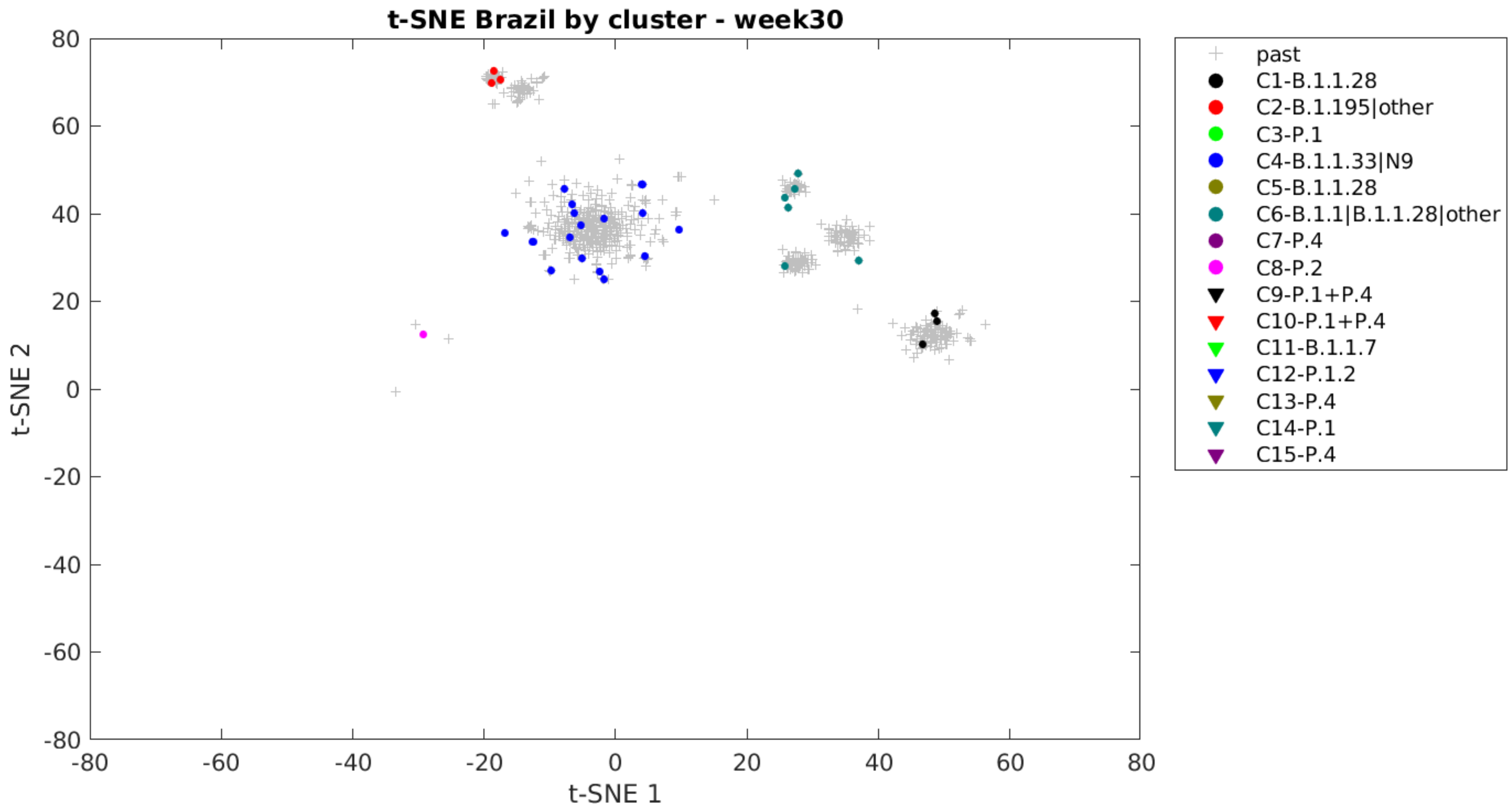

t-SNE Brazil by cluster - week30
80
60
40
20
0
-20
-40
-60
-80
t-SNE 2
-80
-60
-40
-20
0
20
40
60
80
t-SNE 1
past
C1-B.1.1.28
C2-B.1.195|other
C3-P.1
C4-B.1.1.33|N9
C5-B.1.1.28
C6-B.1.1|B.1.1.28|other
C7-P.4
C8-P.2
C9-P.1+P.4
C10-P.1+P.4
C11-B.1.1.7
C12-P.1.2
C13-P.4
C14-P.1
C15-P.4

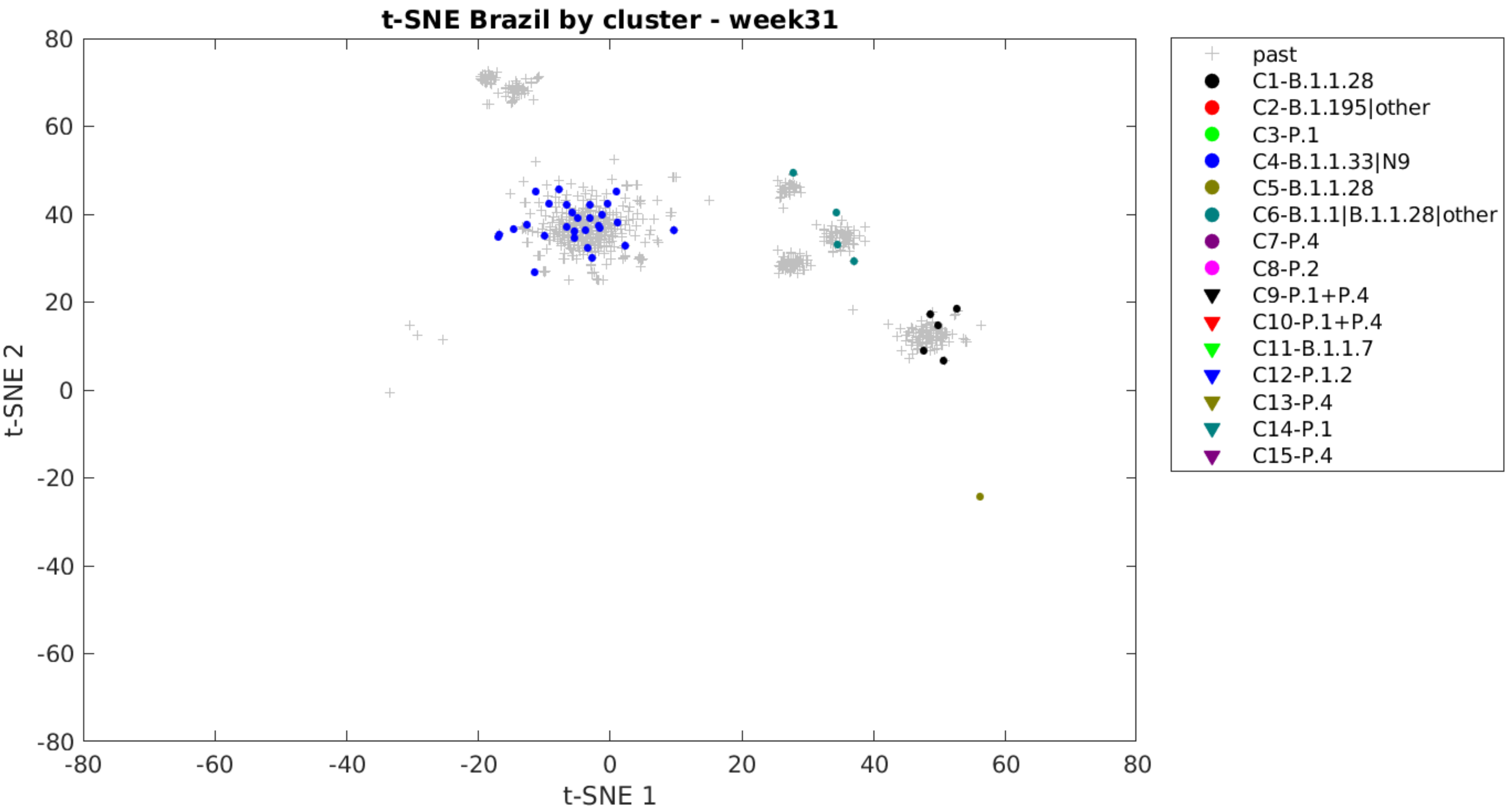

t-SNE Brazil by cluster - week31
t-SNE 1
t-SNE 2
80
60
40
20
0
-20
-40
-60
-80
-80
-60
-40
-20
0
20
40
60
80
past
C1-B.1.1.28
C2-B.1.195|other
C3-P.1
C4-B.1.1.33|N9
C5-B.1.1.28
C6-B.1.1|B.1.1.28|other
C7-P.4
C8-P.2
C9-P.1+P.4
C10-P.1+P.4
C11-B.1.1.7
C12-P.1.2
C13-P.4
C14-P.1
C15-P.4

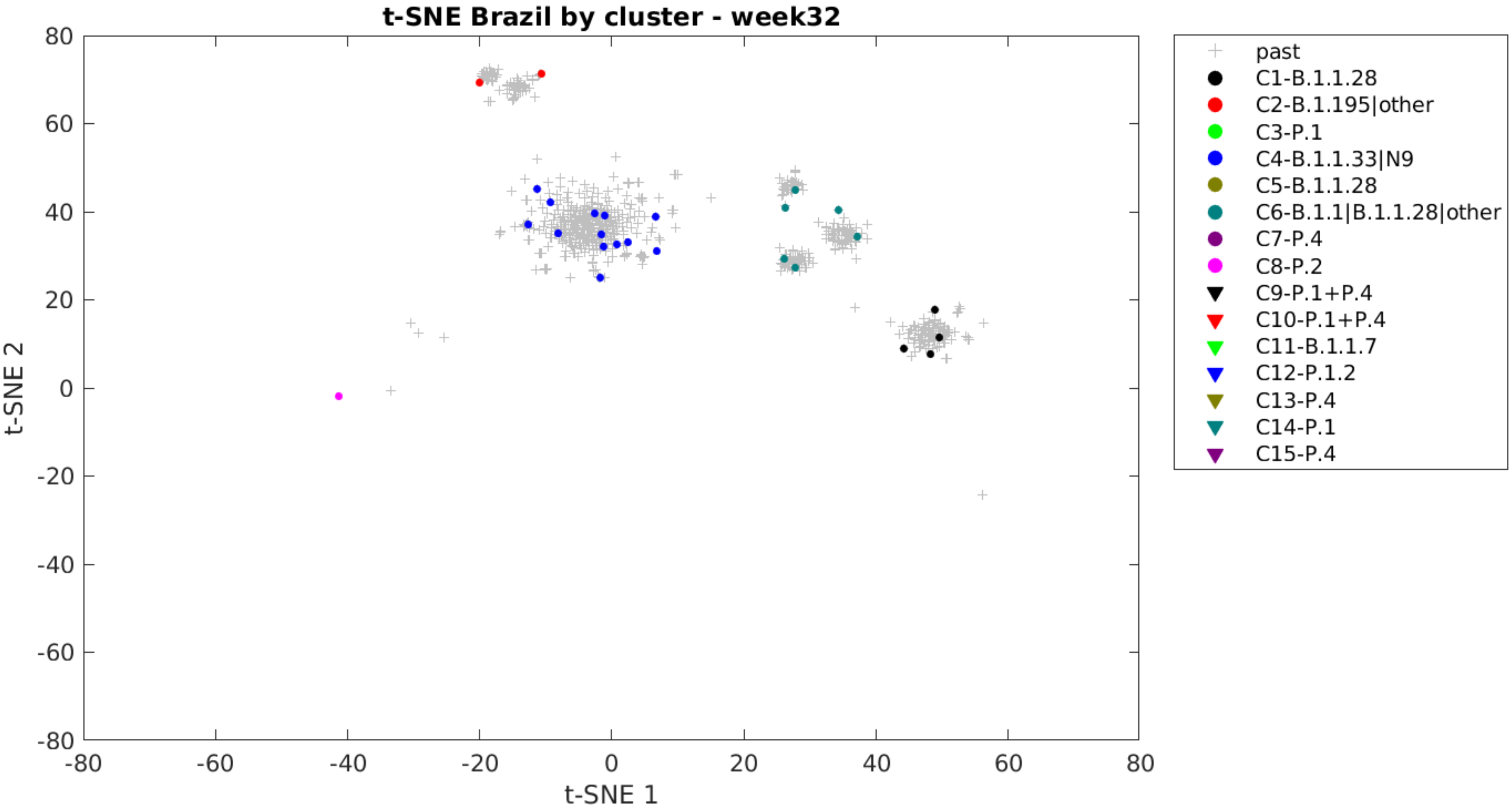
t-SNE Brazil by cluster - week32
t-SNE 1
t-SNE 2
80
60
40
20
0
-20
-40
-60
-80
-80
-60
-40
-20
0
20
40
60
80
past
C1-B.1.1.28
C2-B.1.195|other
C3-P.1
C4-B.1.1.33|N9
C5-B.1.1.28
C6-B.1.1|B.1.1.28|other
C7-P.4
C8-P.2
C9-P.1+P.4
C10-P.1+P.4
C11-B.1.1.7
C12-P.1.2
C13-P.4
C14-P.1
C15-P.4

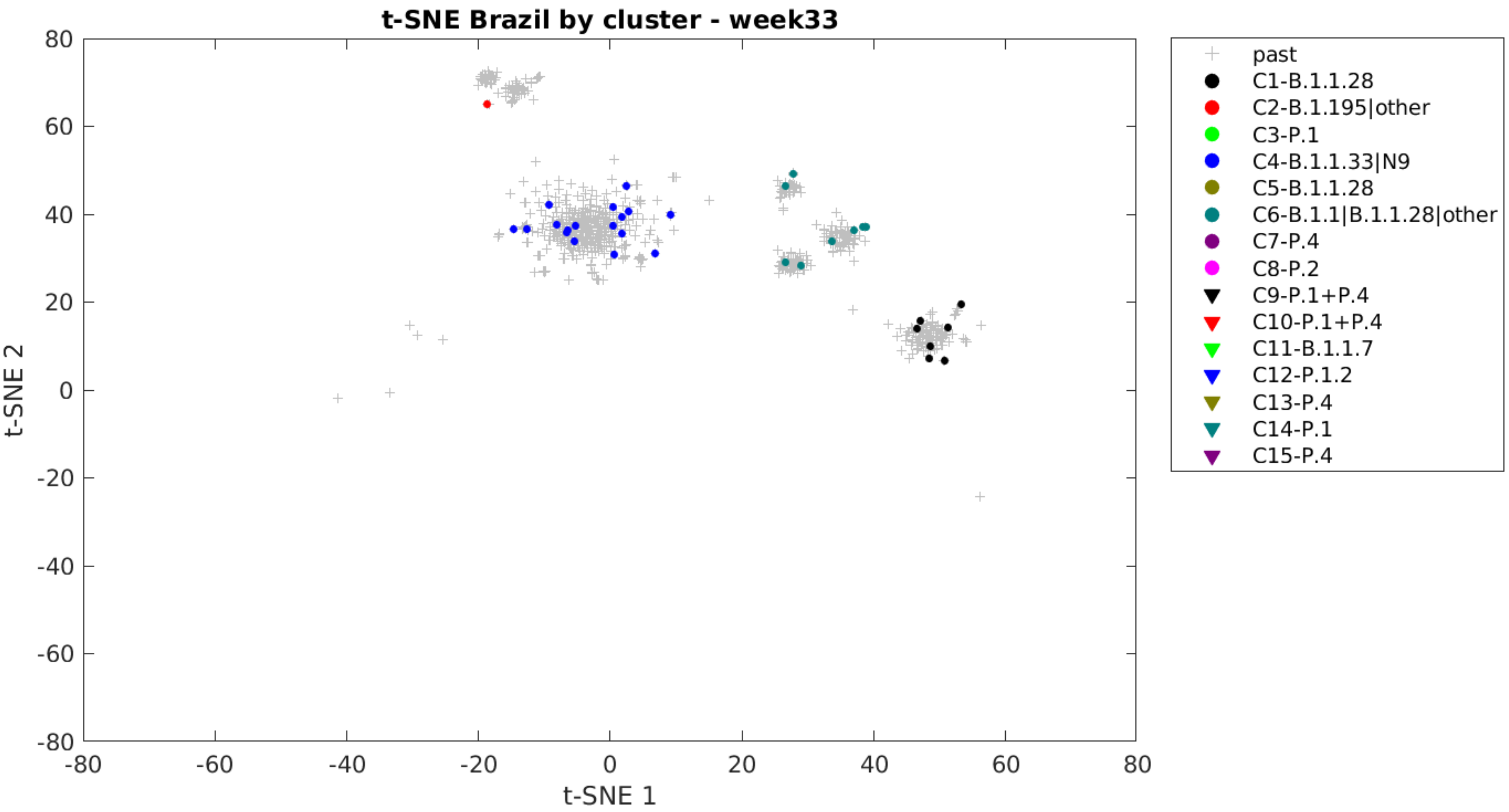

t-SNE Brazil by cluster - week33
80
60
40
20
0
-20
-40
-60
-80
-80
-60
-40
-20
0
20
40
60
80
t-SNE 2
t-SNE 1
past
C1-B.1.1.28
C2-B.1.195|other
C3-P.1
C4-B.1.1.33|N9
C5-B.1.1.28
C6-B.1.1|B.1.1.28|other
C7-P.4
C8-P.2
C9-P.1+P.4
C10-P.1+P.4
C11-B.1.1.7
C12-P.1.2
C13-P.4
C14-P.1
C15-P.4

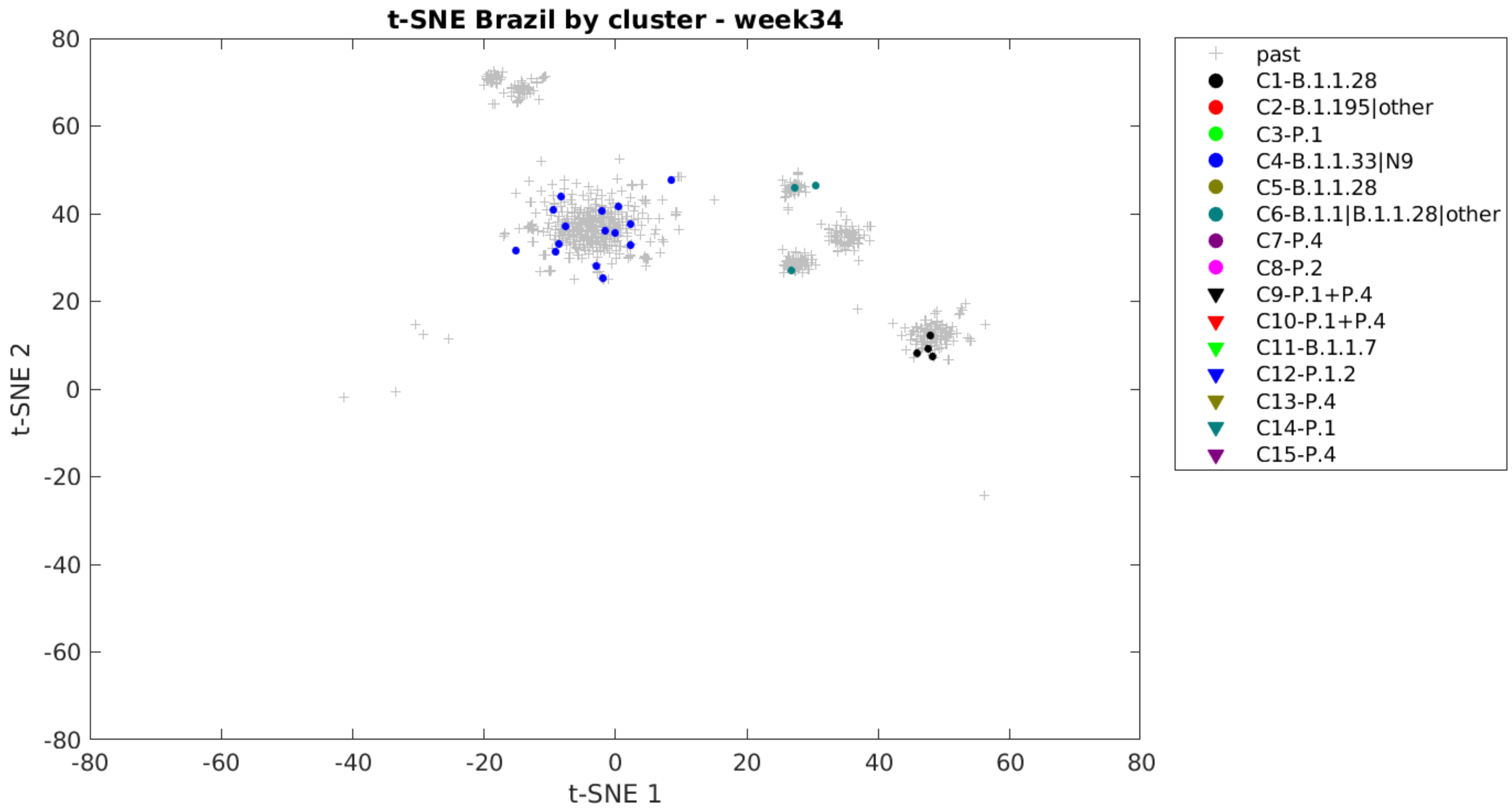
t-SNE Brazil by cluster - week34
t-SNE 1
t-SNE 2
past
C1-B.1.1.28
C2-B.1.195|other
C3-P.1
C4-B.1.1.33|N9
C5-B.1.1.28
C6-B.1.1|B.1.1.28|other
C7-P.4
C8-P.2
C9-P.1+P.4
C10-P.1+P.4
C11-B.1.1.7
C12-P.1.2
C13-P.4
C14-P.1
C15-P.4

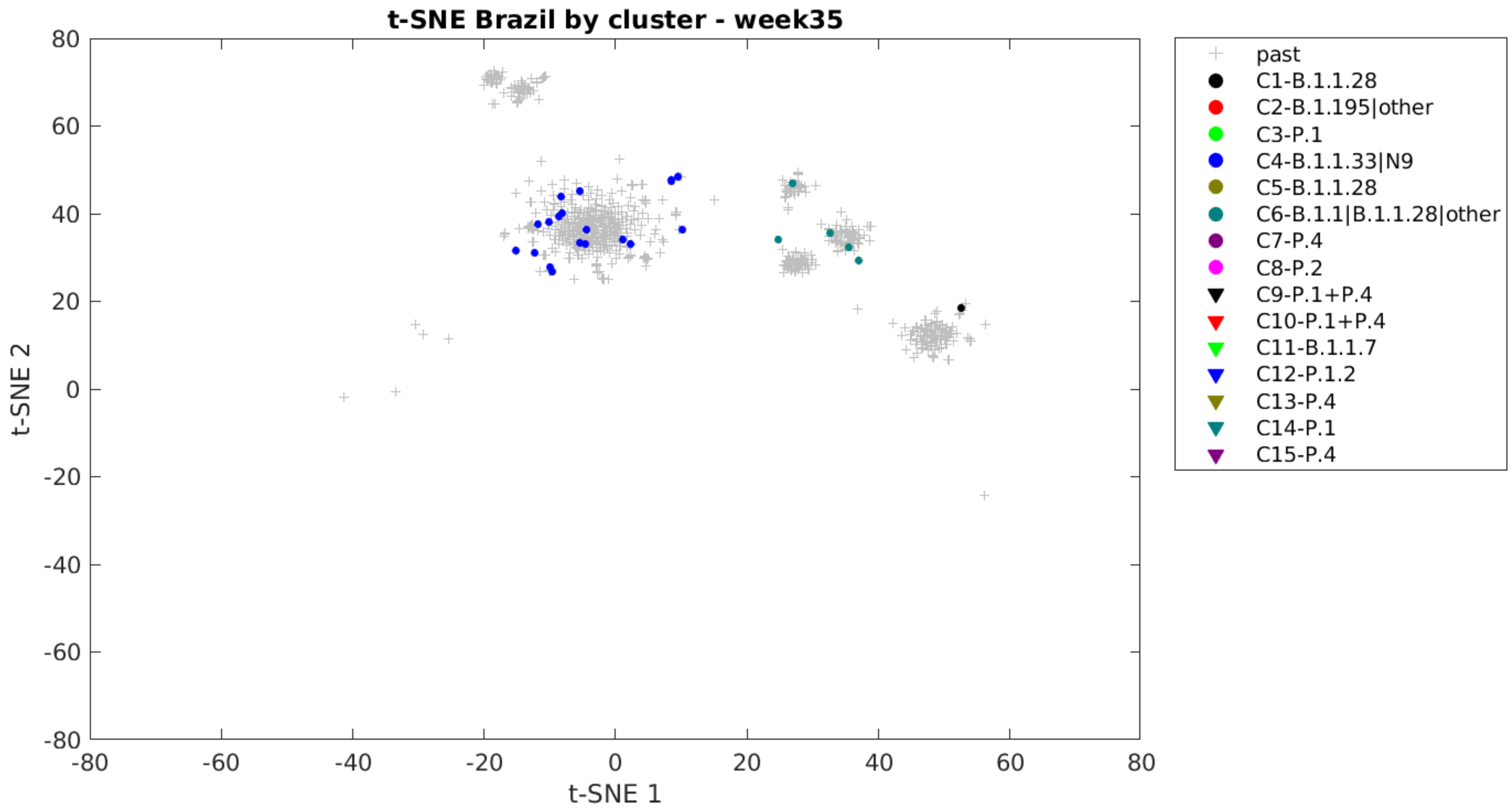

t-SNE Brazil by cluster - week35
80
60
40
20
0
-20
-40
-60
-80
t-SNE 2
-80
-60
-40
-20
0
20
40
60
80
t-SNE 1
past
C1-B.1.1.28
C2-B.1.195|other
C3-P.1
C4-B.1.1.33|N9
C5-B.1.1.28
C6-B.1.1|B.1.1.28|other
C7-P.4
C8-P.2
C9-P.1+P.4
C10-P.1+P.4
C11-B.1.1.7
C12-P.1.2
C13-P.4
C14-P.1
C15-P.4

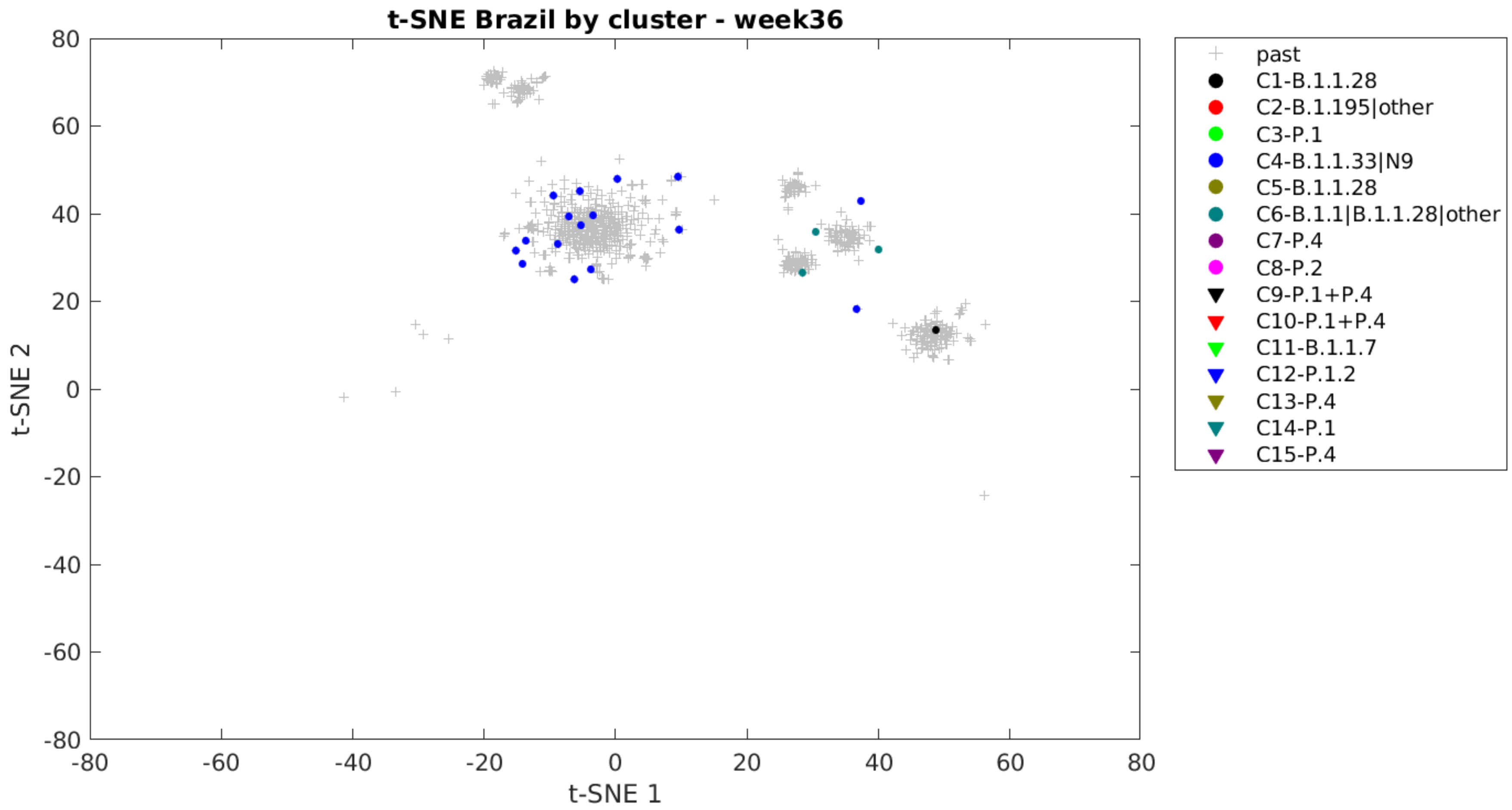

t-SNE Brazil by cluster - week36
t-SNE 2
t-SNE 1
80
60
40
20
0
-20
-40
-60
-80
-80
-60
-40
-20
0
20
40
60
80
past
C1-B.1.1.28
C2-B.1.195|other
C3-P.1
C4-B.1.1.33|N9
C5-B.1.1.28
C6-B.1.1|B.1.1.28|other
C7-P.4
C8-P.2
C9-P.1+P.4
C10-P.1+P.4
C11-B.1.1.7
C12-P.1.2
C13-P.4
C14-P.1
C15-P.4

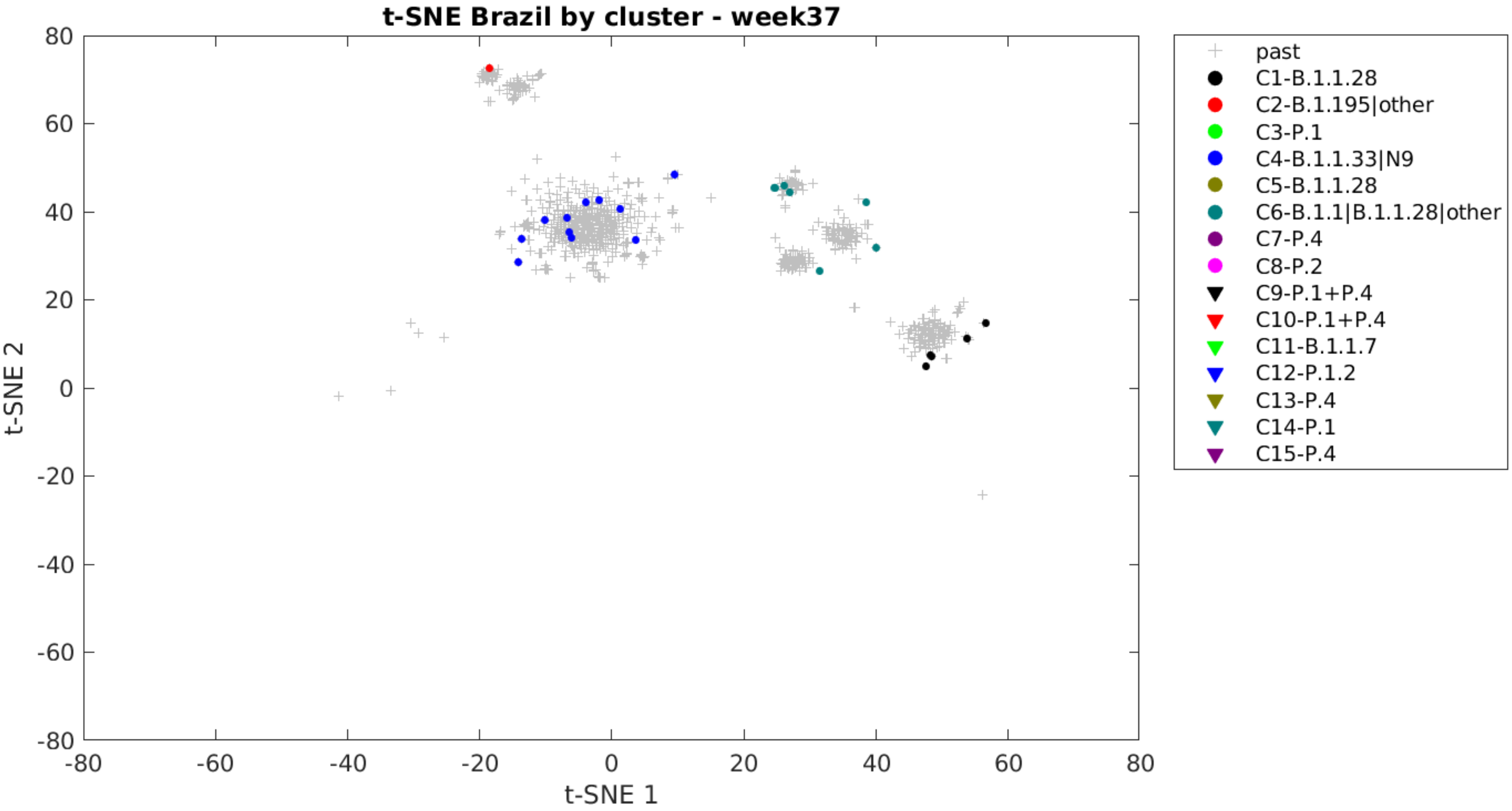

t-SNE Brazil by cluster - week37
t-SNE 2
t-SNE 1
80
60
40
20
0
-20
-40
-60
-80
-80
-60
-40
-20
0
20
40
60
80
past
C1-B.1.1.28
C2-B.1.195|other
C3-P.1
C4-B.1.1.33|N9
C5-B.1.1.28
C6-B.1.1|B.1.1.28|other
C7-P.4
C8-P.2
C9-P.1+P.4
C10-P.1+P.4
C11-B.1.1.7
C12-P.1.2
C13-P.4
C14-P.1
C15-P.4

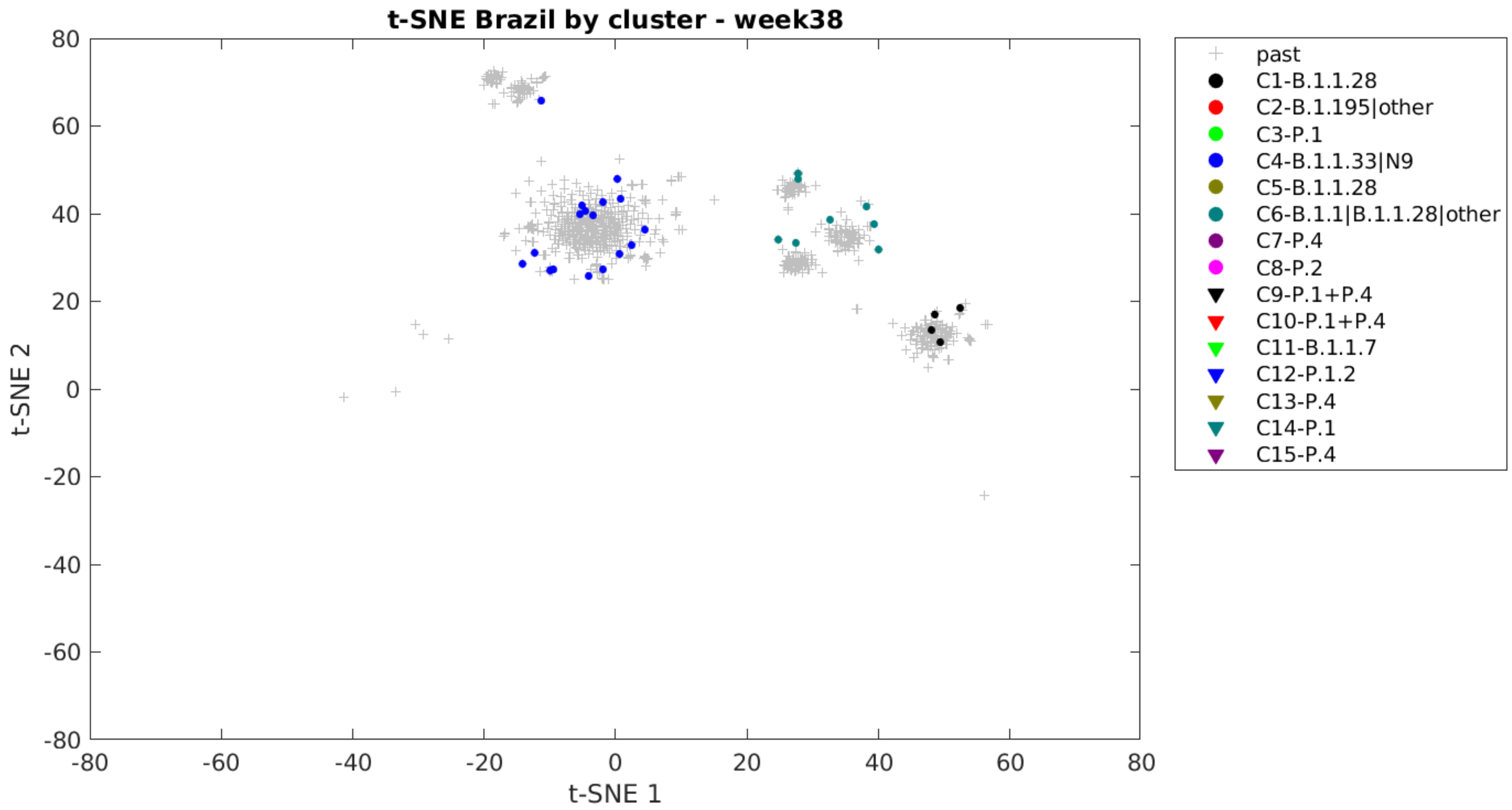
t-SNE Brazil by cluster - week38
t-SNE 1
t-SNE 2
80
60
40
20
0
-20
-40
-60
-80
past
C1-B.1.1.28
C2-B.1.195|other
C3-P.1
C4-B.1.1.33|N9
C5-B.1.1.28
C6-B.1.1|B.1.1.28|other
C7-P.4
C8-P.2
C9-P.1+P.4
C10-P.1+P.4
C11-B.1.1.7
C12-P.1.2
C13-P.4
C14-P.1
C15-P.4

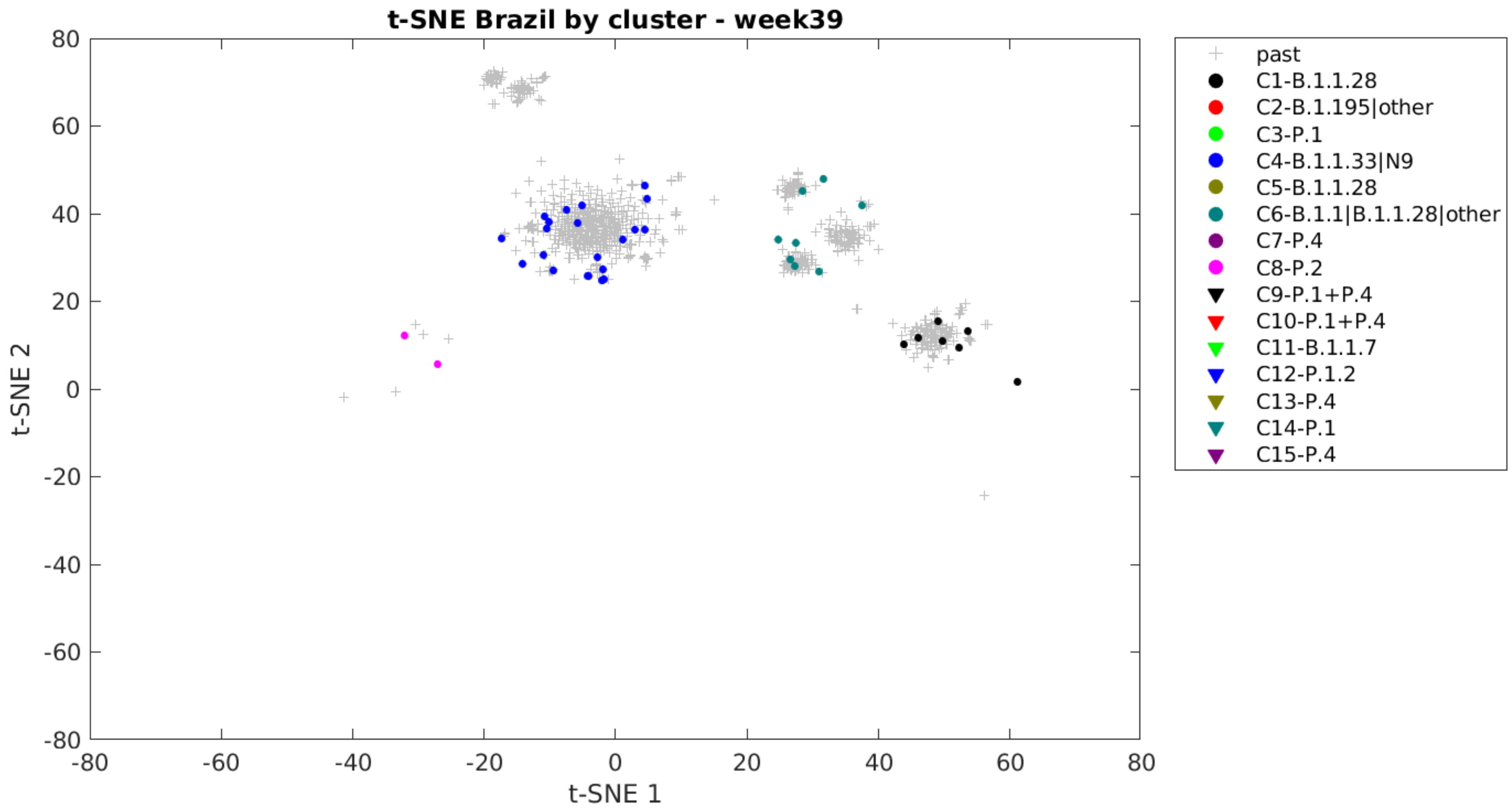
t-SNE Brazil by cluster - week39
80
60
40
20
0
-20
-40
-60
-80
t-SNE 2
-80
-60
-40
-20
0
20
40
60
80
t-SNE 1
past
C1-B.1.1.28
C2-B.1.195|other
C3-P.1
C4-B.1.1.33|N9
C5-B.1.1.28
C6-B.1.1|B.1.1.28|other
C7-P.4
C8-P.2
C9-P.1+P.4
C10-P.1+P.4
C11-B.1.1.7
C12-P.1.2
C13-P.4
C14-P.1
C15-P.4

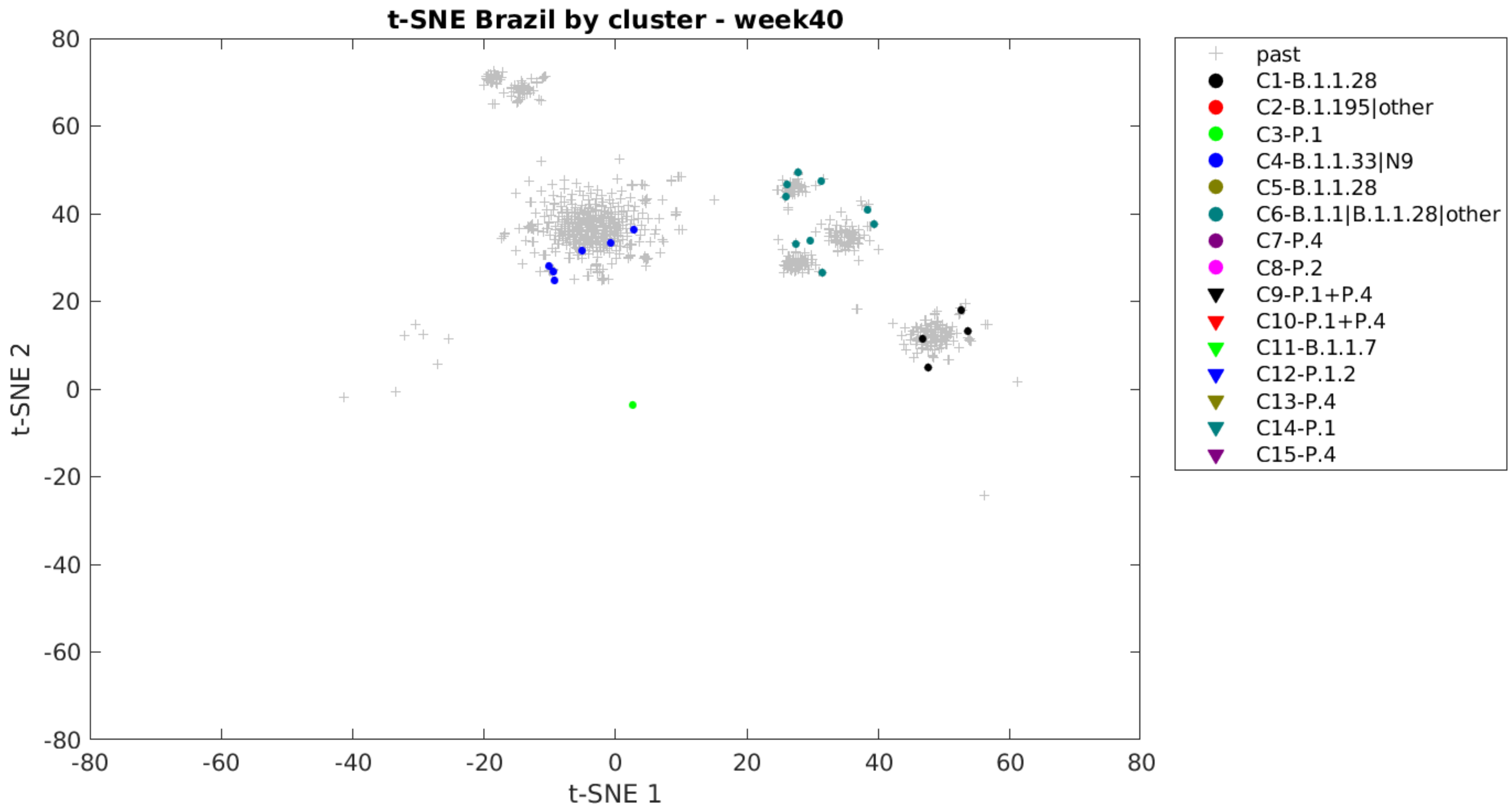
t-SNE Brazil by cluster - week40
t-SNE 1
t-SNE 2
past
C1-B.1.1.28
C2-B.1.195|other
C3-P.1
C4-B.1.1.33|N9
C5-B.1.1.28
C6-B.1.1|B.1.1.28|other
C7-P.4
C8-P.2
C9-P.1+P.4
C10-P.1+P.4
C11-B.1.1.7
C12-P.1.2
C13-P.4
C14-P.1
C15-P.4

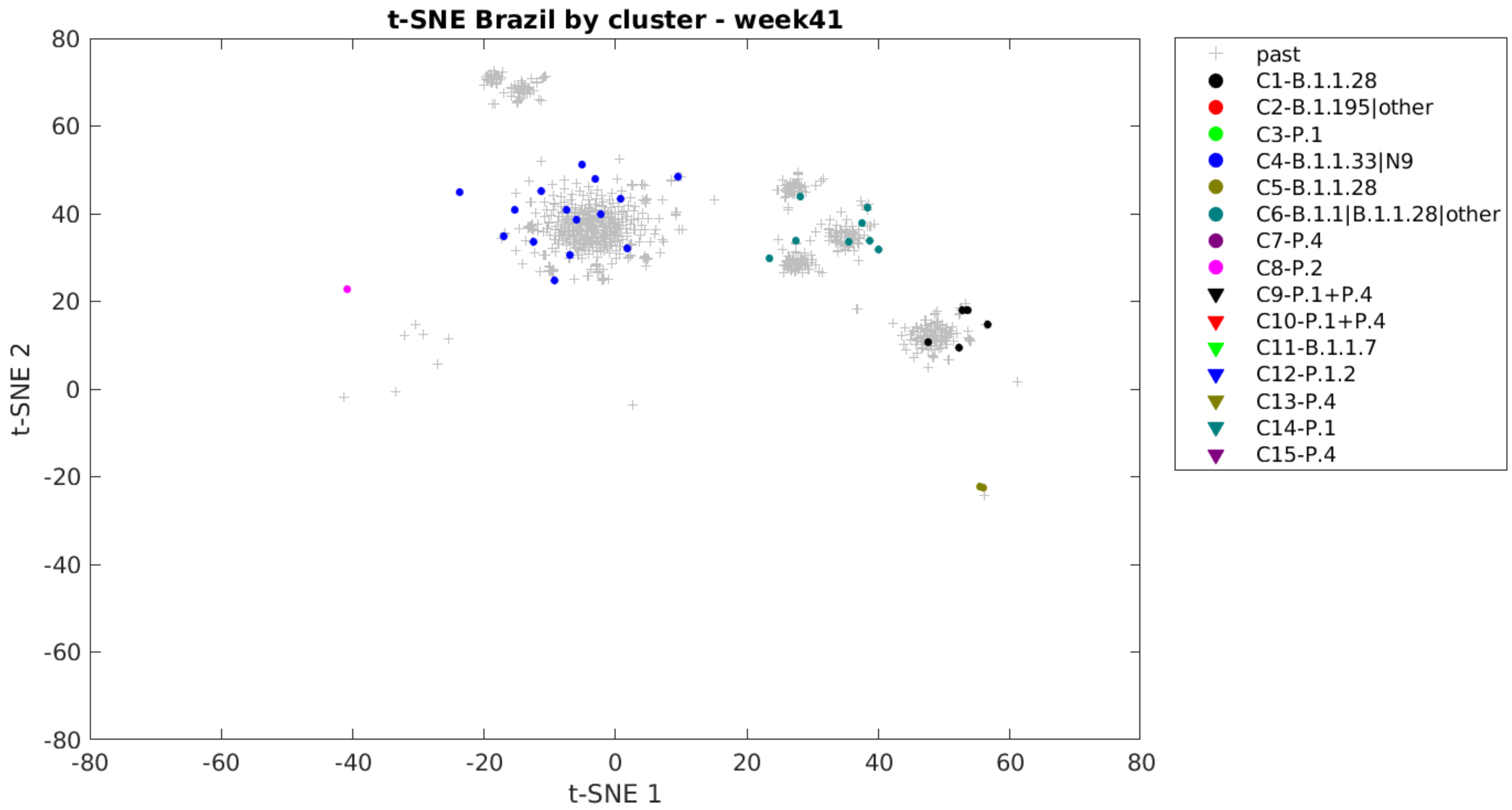

t-SNE Brazil by cluster - week41
t-SNE 1
t-SNE 2
80
60
40
20
0
-20
-40
-60
-80
-80
-60
-40
-20
0
20
40
60
80
past
C1-B.1.1.28
C2-B.1.195|other
C3-P.1
C4-B.1.1.33|N9
C5-B.1.1.28
C6-B.1.1|B.1.1.28|other
C7-P.4
C8-P.2
C9-P.1+P.4
C10-P.1+P.4
C11-B.1.1.7
C12-P.1.2
C13-P.4
C14-P.1
C15-P.4

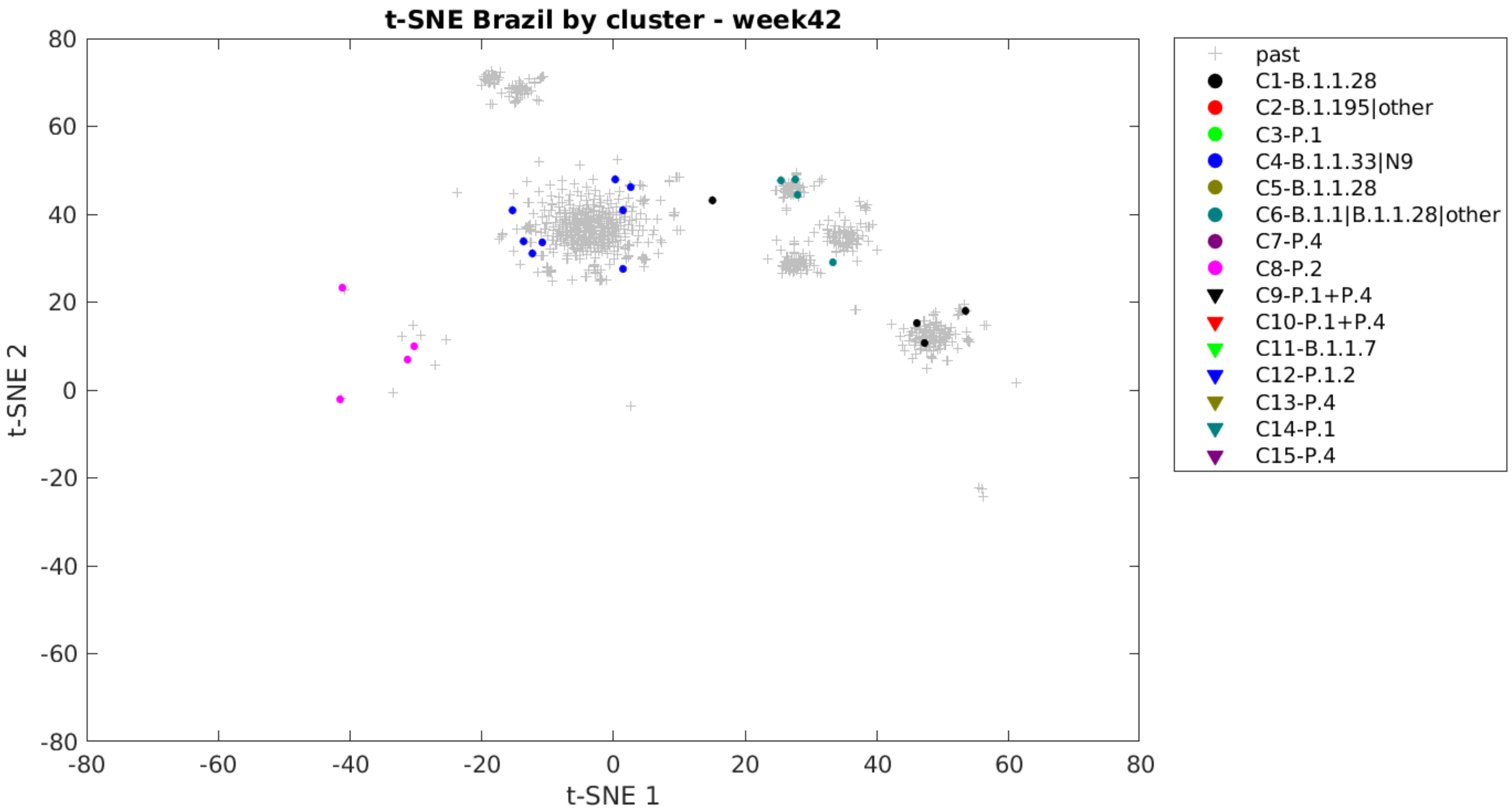
t-SNE Brazil by cluster - week42
t-SNE 1
t-SNE 2
past
C1-B.1.1.28
C2-B.1.195|other
C3-P.1
C4-B.1.1.33|N9
C5-B.1.1.28
C6-B.1.1|B.1.1.28|other
C7-P.4
C8-P.2
C9-P.1+P.4
C10-P.1+P.4
C11-B.1.1.7
C12-P.1.2
C13-P.4
C14-P.1
C15-P.4

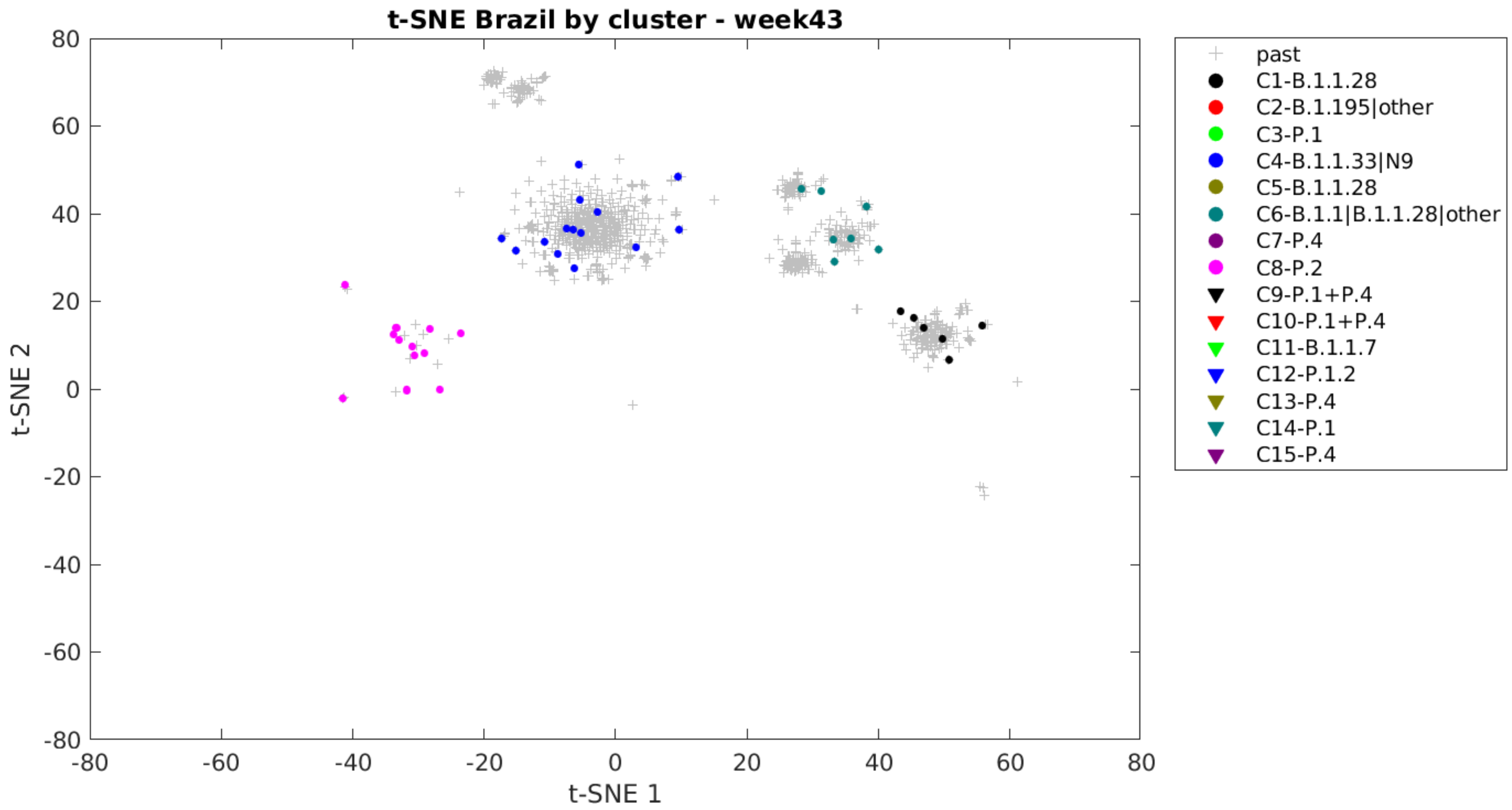

t-SNE Brazil by cluster - week43
t-SNE 1
t-SNE 2
past
C1-B.1.1.28
C2-B.1.195|other
C3-P.1
C4-B.1.1.33|N9
C5-B.1.1.28
C6-B.1.1|B.1.1.28|other
C7-P.4
C8-P.2
C9-P.1+P.4
C10-P.1+P.4
C11-B.1.1.7
C12-P.1.2
C13-P.4
C14-P.1
C15-P.4

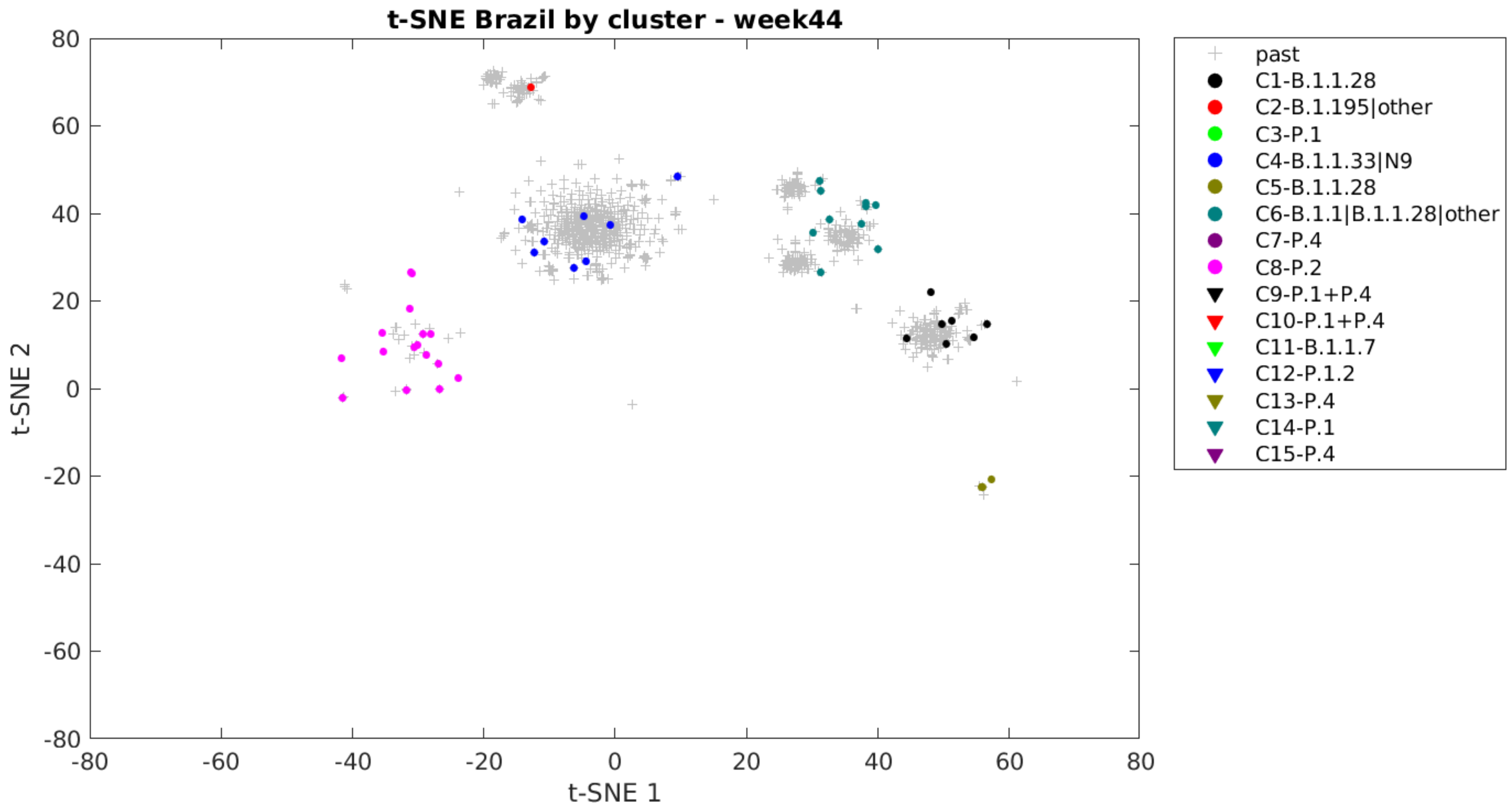

t-SNE Brazil by cluster - week44
t-SNE 1
t-SNE 2
80
60
40
20
0
-20
-40
-60
-80
-80
-60
-40
-20
0
20
40
60
80
past
C1-B.1.1.28
C2-B.1.195|other
C3-P.1
C4-B.1.1.33|N9
C5-B.1.1.28
C6-B.1.1|B.1.1.28|other
C7-P.4
C8-P.2
C9-P.1+P.4
C10-P.1+P.4
C11-B.1.1.7
C12-P.1.2
C13-P.4
C14-P.1
C15-P.4

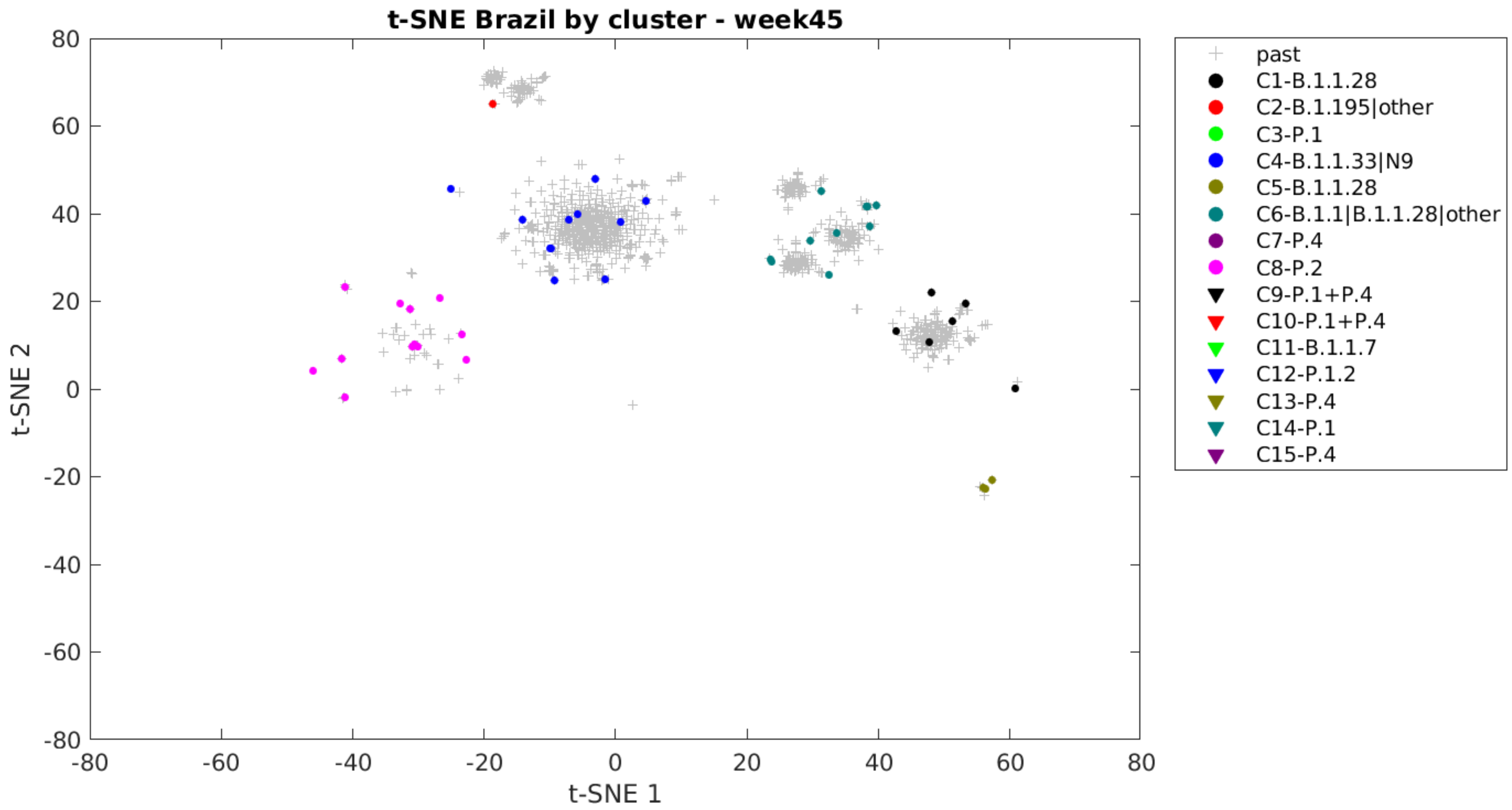

t-SNE Brazil by cluster - week45
t-SNE 2
t-SNE 1
80
60
40
20
0
-20
-40
-60
-80
-80
-60
-40
-20
0
20
40
60
80
past
C1-B.1.1.28
C2-B.1.195|other
C3-P.1
C4-B.1.1.33|N9
C5-B.1.1.28
C6-B.1.1|B.1.1.28|other
C7-P.4
C8-P.2
C9-P.1+P.4
C10-P.1+P.4
C11-B.1.1.7
C12-P.1.2
C13-P.4
C14-P.1
C15-P.4

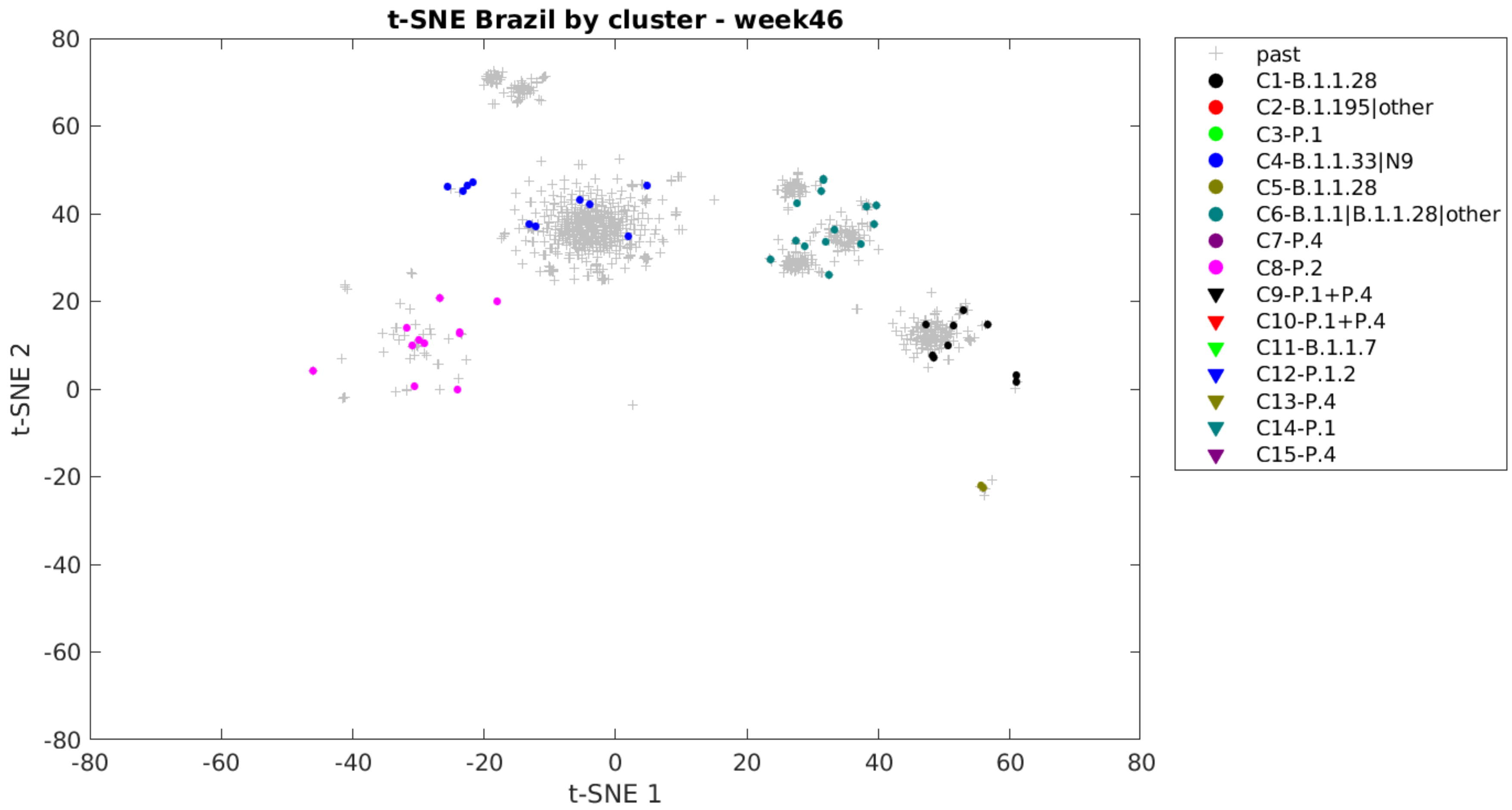

t-SNE Brazil by cluster - week46
t-SNE 1
t-SNE 2
80
60
40
20
0
-20
-40
-60
-80
past
C1-B.1.1.28
C2-B.1.195|other
C3-P.1
C4-B.1.1.33|N9
C5-B.1.1.28
C6-B.1.1|B.1.1.28|other
C7-P.4
C8-P.2
C9-P.1+P.4
C10-P.1+P.4
C11-B.1.1.7
C12-P.1.2
C13-P.4
C14-P.1
C15-P.4

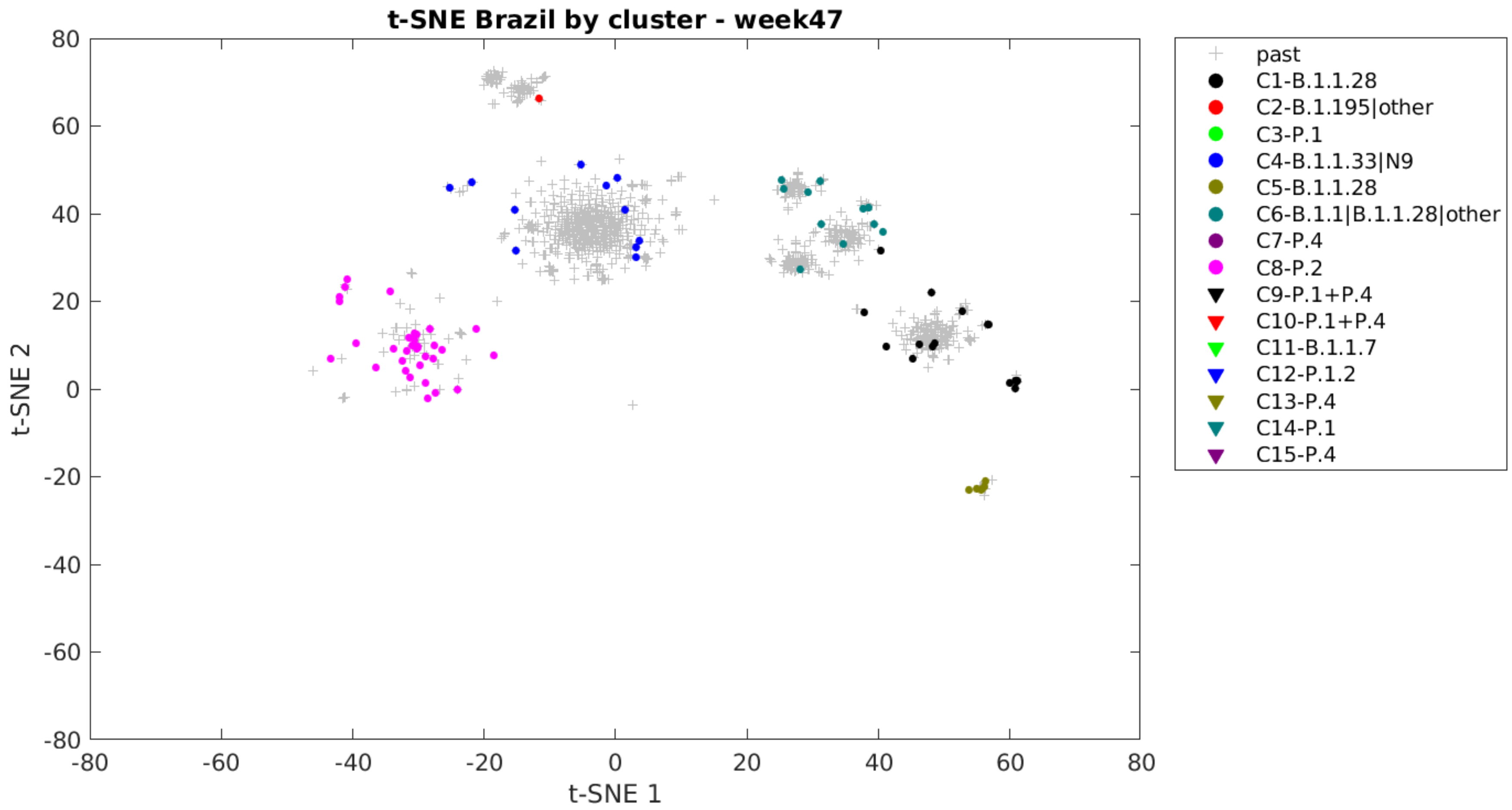
t-SNE Brazil by cluster - week47
t-SNE 1
t-SNE 2
80
60
40
20
0
-20
-40
-60
-80
-80
-60
-40
-20
0
20
40
60
80
past
C1-B.1.1.28
C2-B.1.195|other
C3-P.1
C4-B.1.1.33|N9
C5-B.1.1.28
C6-B.1.1|B.1.1.28|other
C7-P.4
C8-P.2
C9-P.1+P.4
C10-P.1+P.4
C11-B.1.1.7
C12-P.1.2
C13-P.4
C14-P.1
C15-P.4

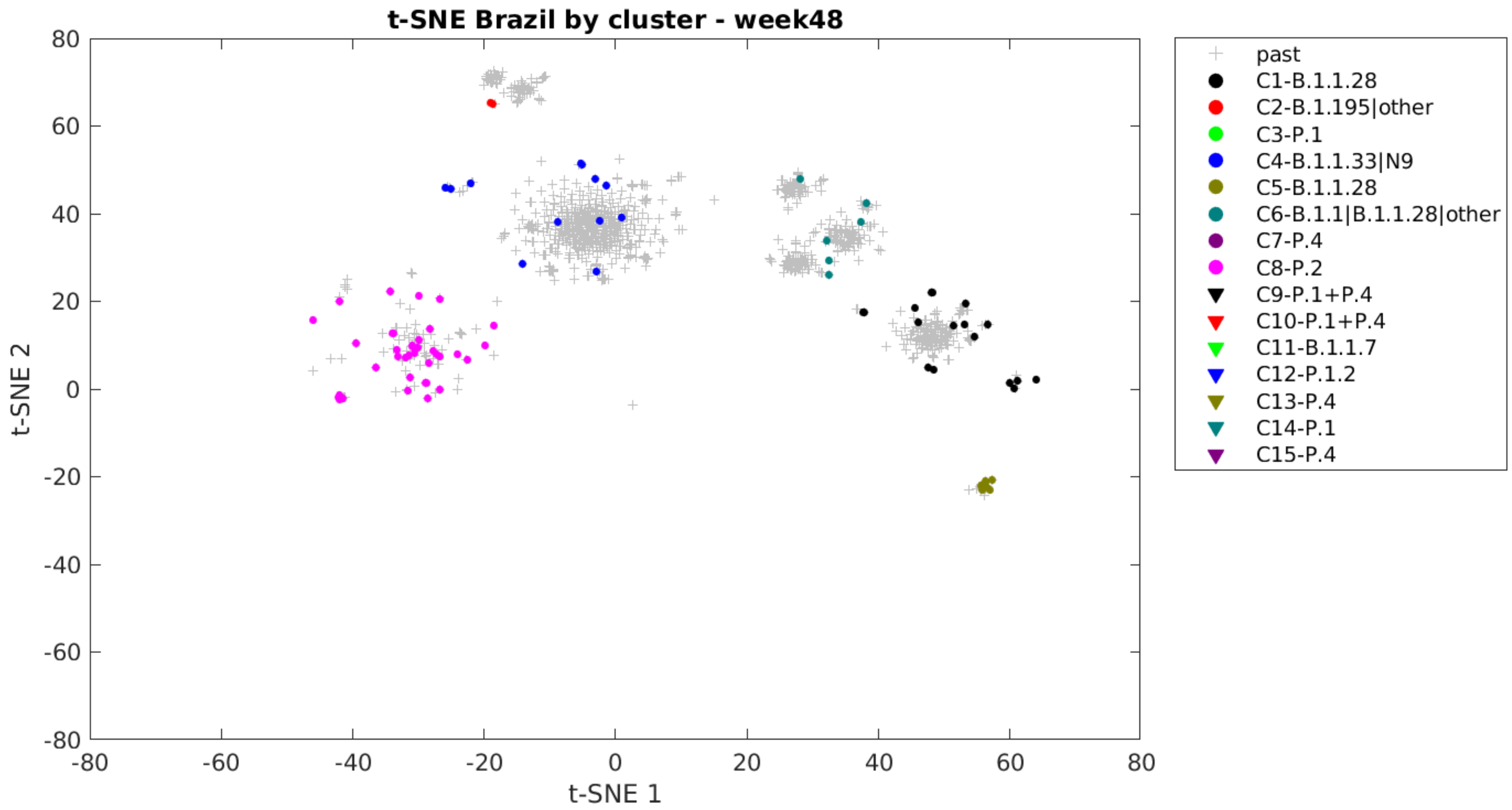
t-SNE Brazil by cluster - week48
t-SNE 1
t-SNE 2
80
60
40
20
0
-20
-40
-60
-80
past
C1-B.1.1.28
C2-B.1.195|other
C3-P.1
C4-B.1.1.33|N9
C5-B.1.1.28
C6-B.1.1|B.1.1.28|other
C7-P.4
C8-P.2
C9-P.1+P.4
C10-P.1+P.4
C11-B.1.1.7
C12-P.1.2
C13-P.4
C14-P.1
C15-P.4

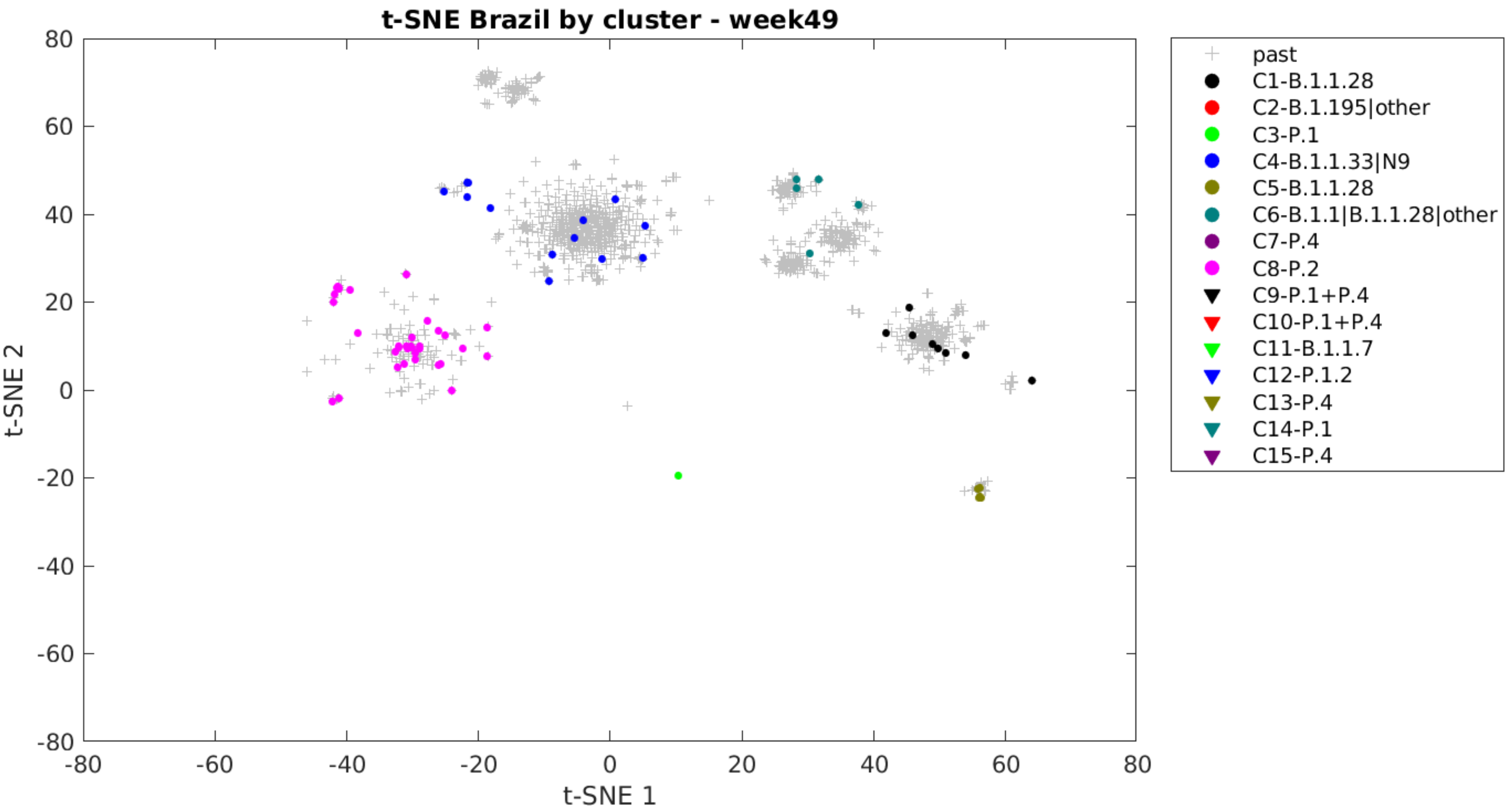
t-SNE Brazil by cluster - week49
t-SNE 1
t-SNE 2
-80
-60
-40
-20
0
20
40
60
80
past
C1-B.1.1.28
C2-B.1.195|other
C3-P.1
C4-B.1.1.33|N9
C5-B.1.1.28
C6-B.1.1|B.1.1.28|other
C7-P.4
C8-P.2
C9-P.1+P.4
C10-P.1+P.4
C11-B.1.1.7
C12-P.1.2
C13-P.4
C14-P.1
C15-P.4

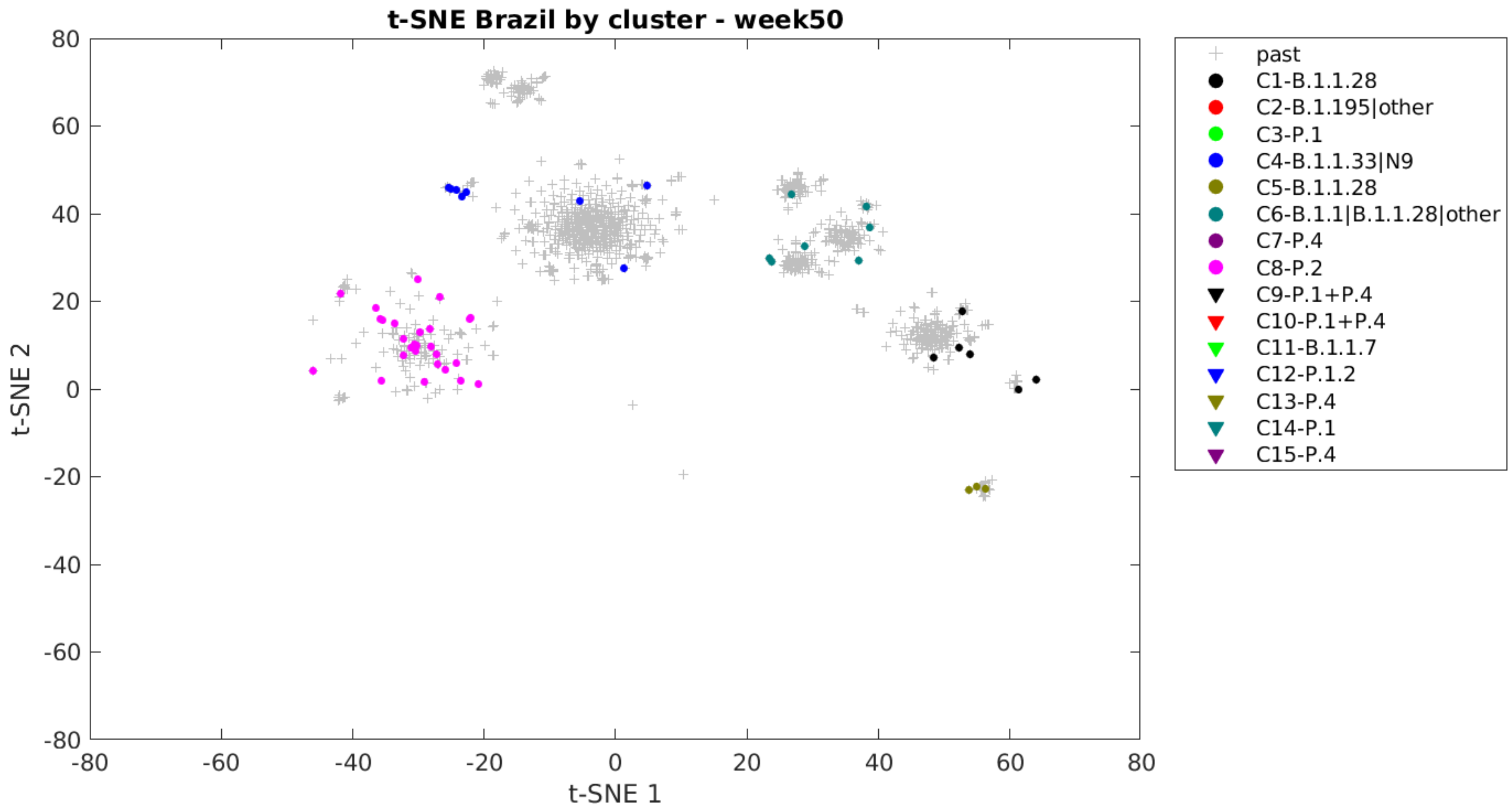
t-SNE Brazil by cluster - week50
t-SNE 1
t-SNE 2
80
60
40
20
0
-20
-40
-60
-80
past
C1-B.1.1.28
C2-B.1.195|other
C3-P.1
C4-B.1.1.33|N9
C5-B.1.1.28
C6-B.1.1|B.1.1.28|other
C7-P.4
C8-P.2
C9-P.1+P.4
C10-P.1+P.4
C11-B.1.1.7
C12-P.1.2
C13-P.4
C14-P.1
C15-P.4

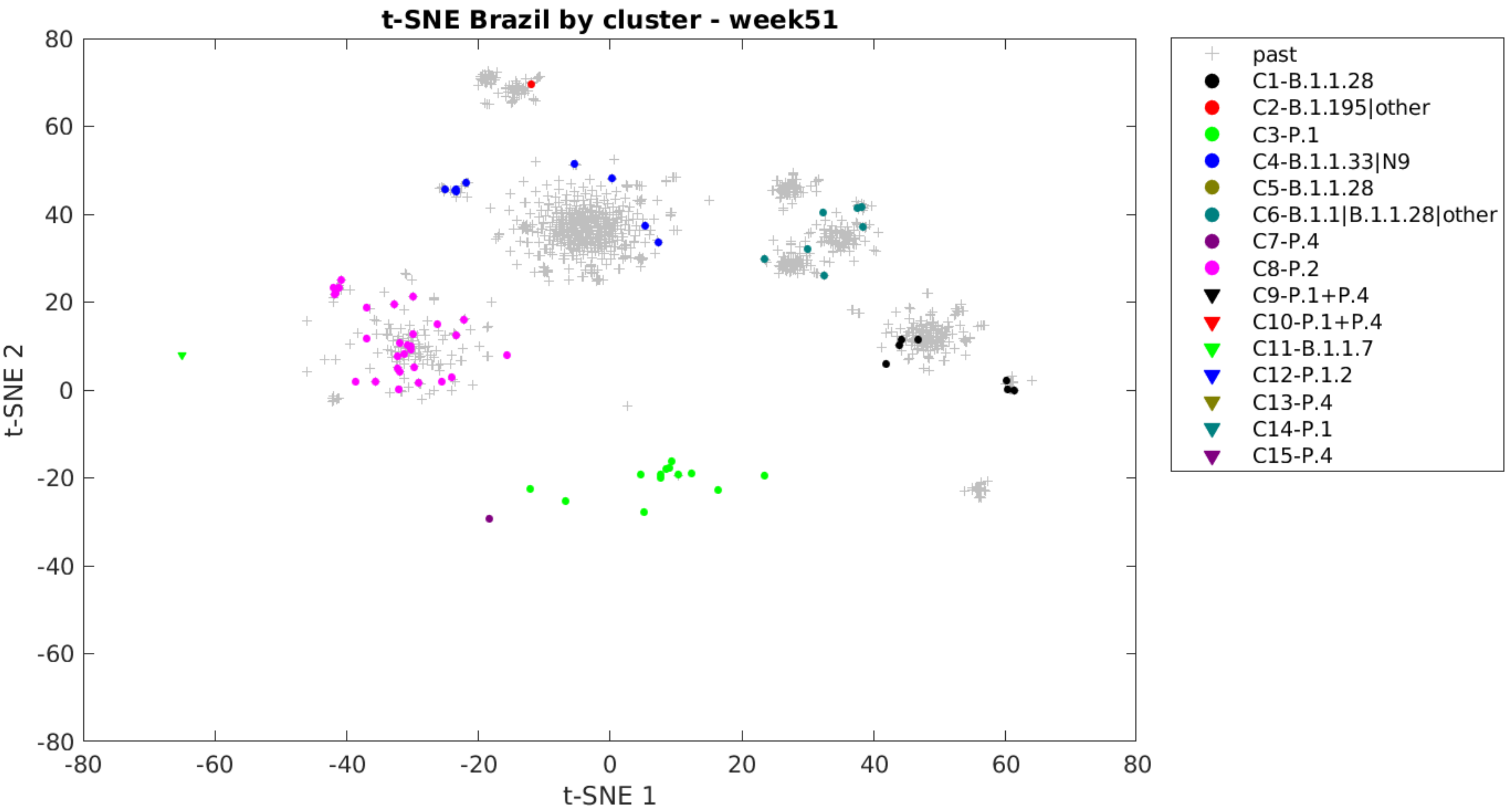
t-SNE Brazil by cluster - week51
t-SNE 1
t-SNE 2
-80
-60
-40
-20
0
20
40
60
80
past
C1-B.1.1.28
C2-B.1.195|other
C3-P.1
C4-B.1.1.33|N9
C5-B.1.1.28
C6-B.1.1|B.1.1.28|other
C7-P.4
C8-P.2
C9-P.1+P.4
C10-P.1+P.4
C11-B.1.1.7
C12-P.1.2
C13-P.4
C14-P.1
C15-P.4

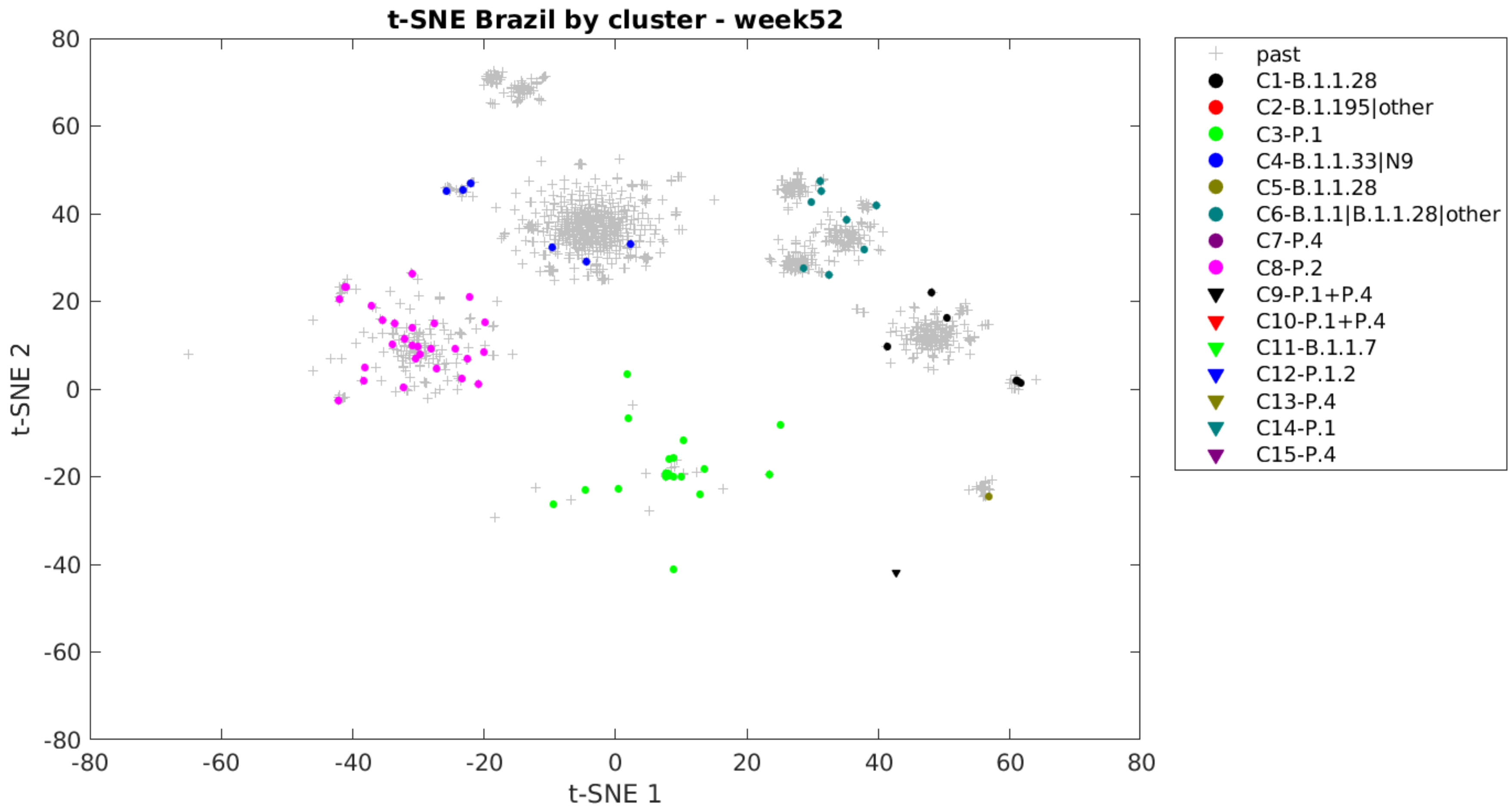
t-SNE Brazil by cluster - week52
t-SNE 1
t-SNE 2
80
60
40
20
0
-20
-40
-60
-80
-80
-60
-40
-20
0
20
40
60
80
past
C1-B.1.1.28
C2-B.1.195|other
C3-P.1
C4-B.1.1.33|N9
C5-B.1.1.28
C6-B.1.1|B.1.1.28|other
C7-P.4
C8-P.2
C9-P.1+P.4
C10-P.1+P.4
C11-B.1.1.7
C12-P.1.2
C13-P.4
C14-P.1
C15-P.4

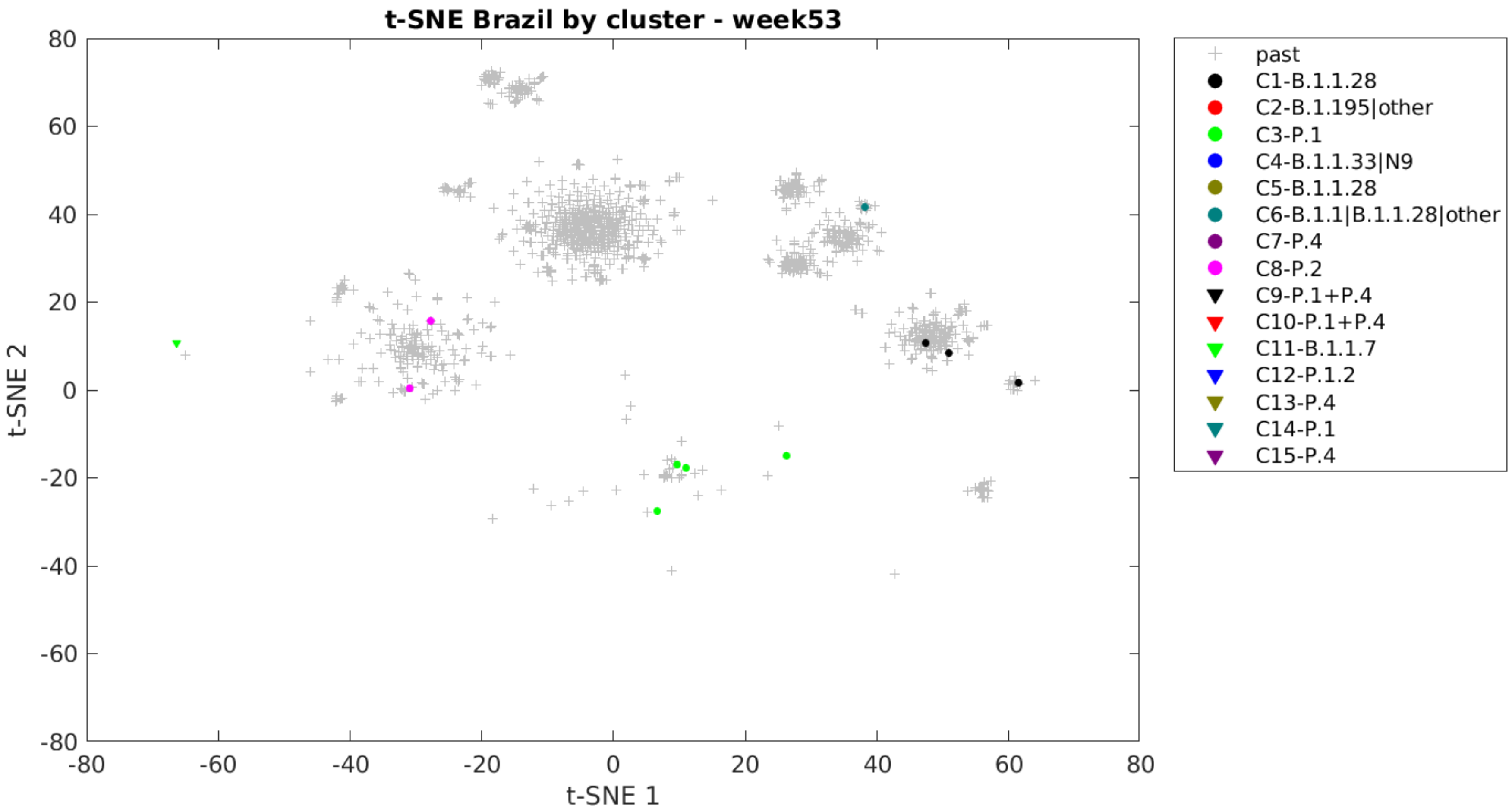

t-SNE Brazil by cluster - week53
t-SNE 1
t-SNE 2
80
60
40
20
0
-20
-40
-60
-80
-80
-60
-40
-20
0
20
40
60
80
past
C1-B.1.1.28
C2-B.1.195|other
C3-P.1
C4-B.1.1.33|N9
C5-B.1.1.28
C6-B.1.1|B.1.1.28|other
C7-P.4
C8-P.2
C9-P.1+P.4
C10-P.1+P.4
C11-B.1.1.7
C12-P.1.2
C13-P.4
C14-P.1
C15-P.4

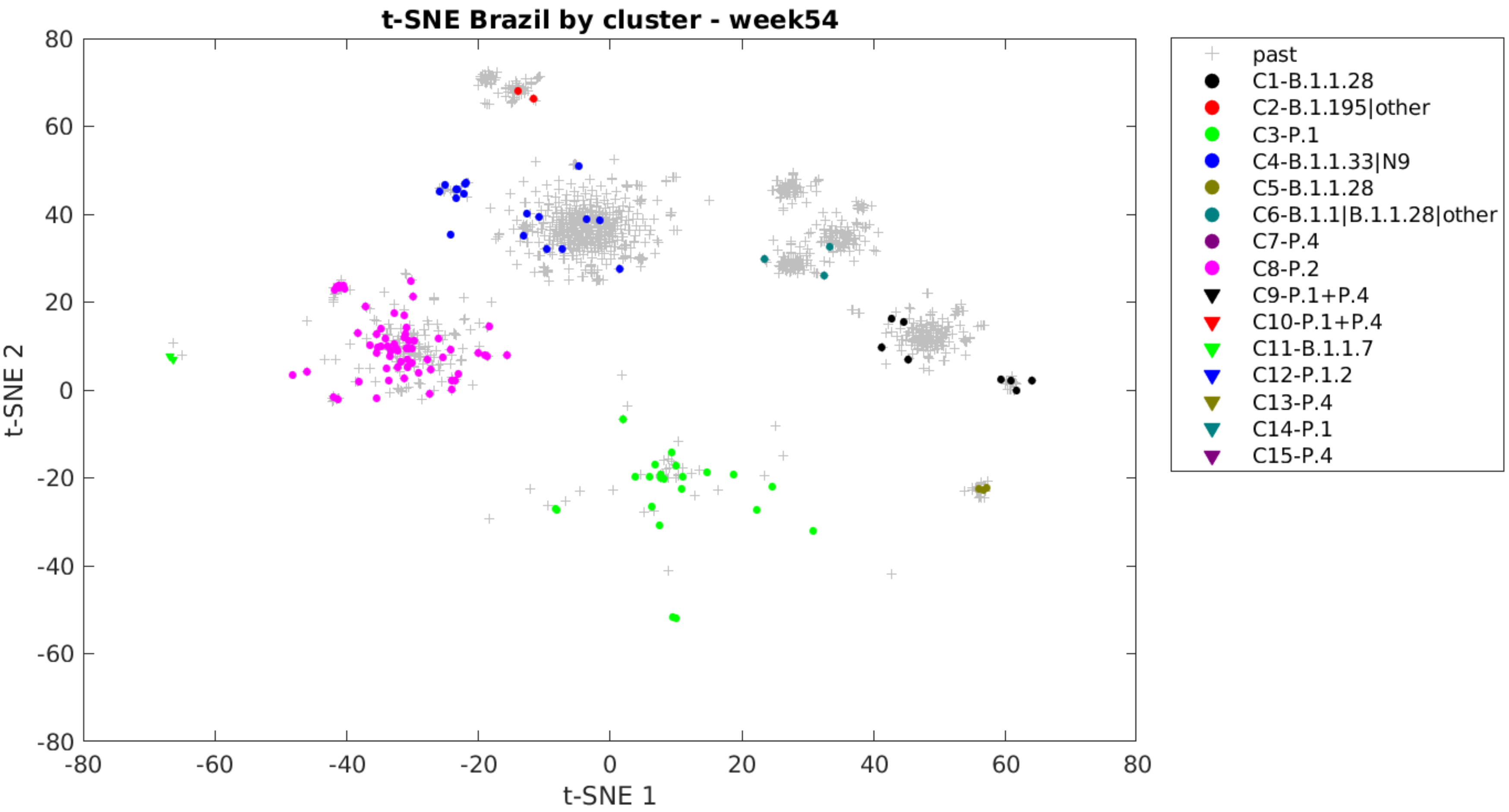
t-SNE Brazil by cluster - week54
t-SNE 1
t-SNE 2
80
60
40
20
0
-20
-40
-60
-80
-80
-60
-40
-20
0
20
40
60
80
past
C1-B.1.1.28
C2-B.1.195|other
C3-P.1
C4-B.1.1.33|N9
C5-B.1.1.28
C6-B.1.1|B.1.1.28|other
C7-P.4
C8-P.2
C9-P.1+P.4
C10-P.1+P.4
C11-B.1.1.7
C12-P.1.2
C13-P.4
C14-P.1
C15-P.4

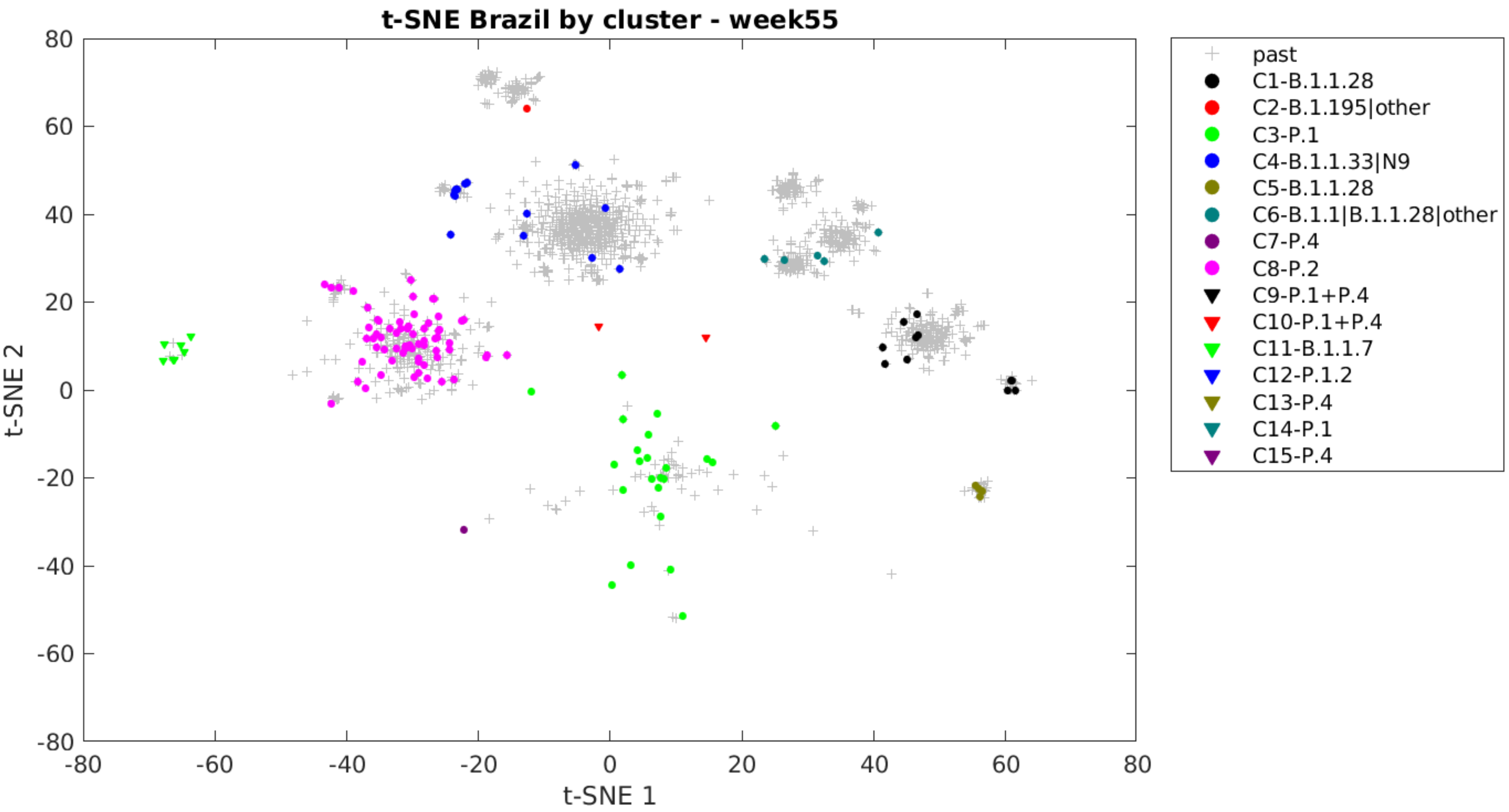
t-SNE Brazil by cluster - week55
t-SNE 2
t-SNE 1
80
60
40
20
0
-20
-40
-60
-80
-80
-60
-40
-20
0
20
40
60
80
past
C1-B.1.1.28
C2-B.1.195|other
C3-P.1
C4-B.1.1.33|N9
C5-B.1.1.28
C6-B.1.1|B.1.1.28|other
C7-P.4
C8-P.2
C9-P.1+P.4
C10-P.1+P.4
C11-B.1.1.7
C12-P.1.2
C13-P.4
C14-P.1
C15-P.4

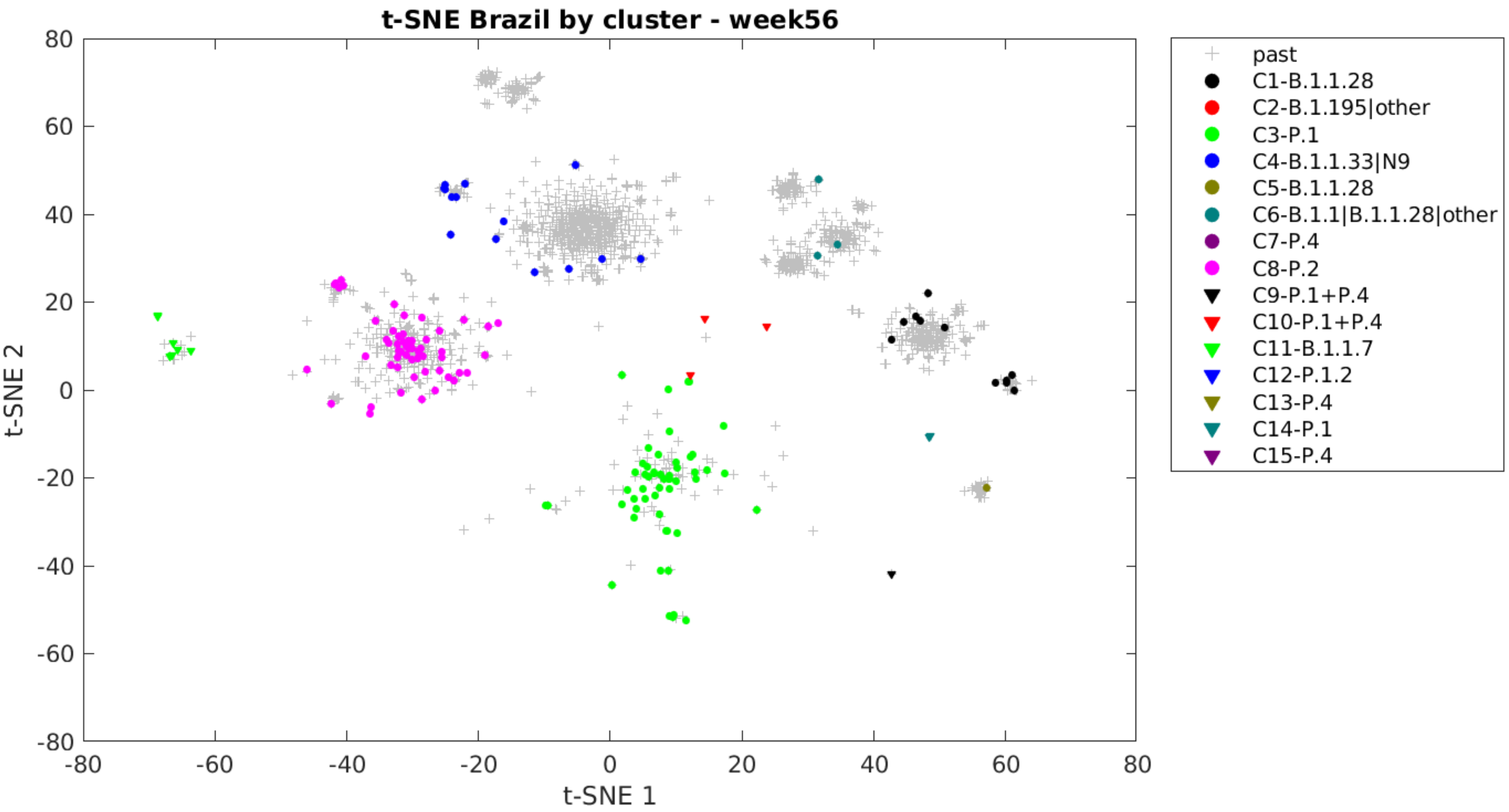

t-SNE Brazil by cluster - week56
t-SNE 1
t-SNE 2
past
C1-B.1.1.28
C2-B.1.195|other
C3-P.1
C4-B.1.1.33|N9
C5-B.1.1.28
C6-B.1.1|B.1.1.28|other
C7-P.4
C8-P.2
C9-P.1+P.4
C10-P.1+P.4
C11-B.1.1.7
C12-P.1.2
C13-P.4
C14-P.1
C15-P.4

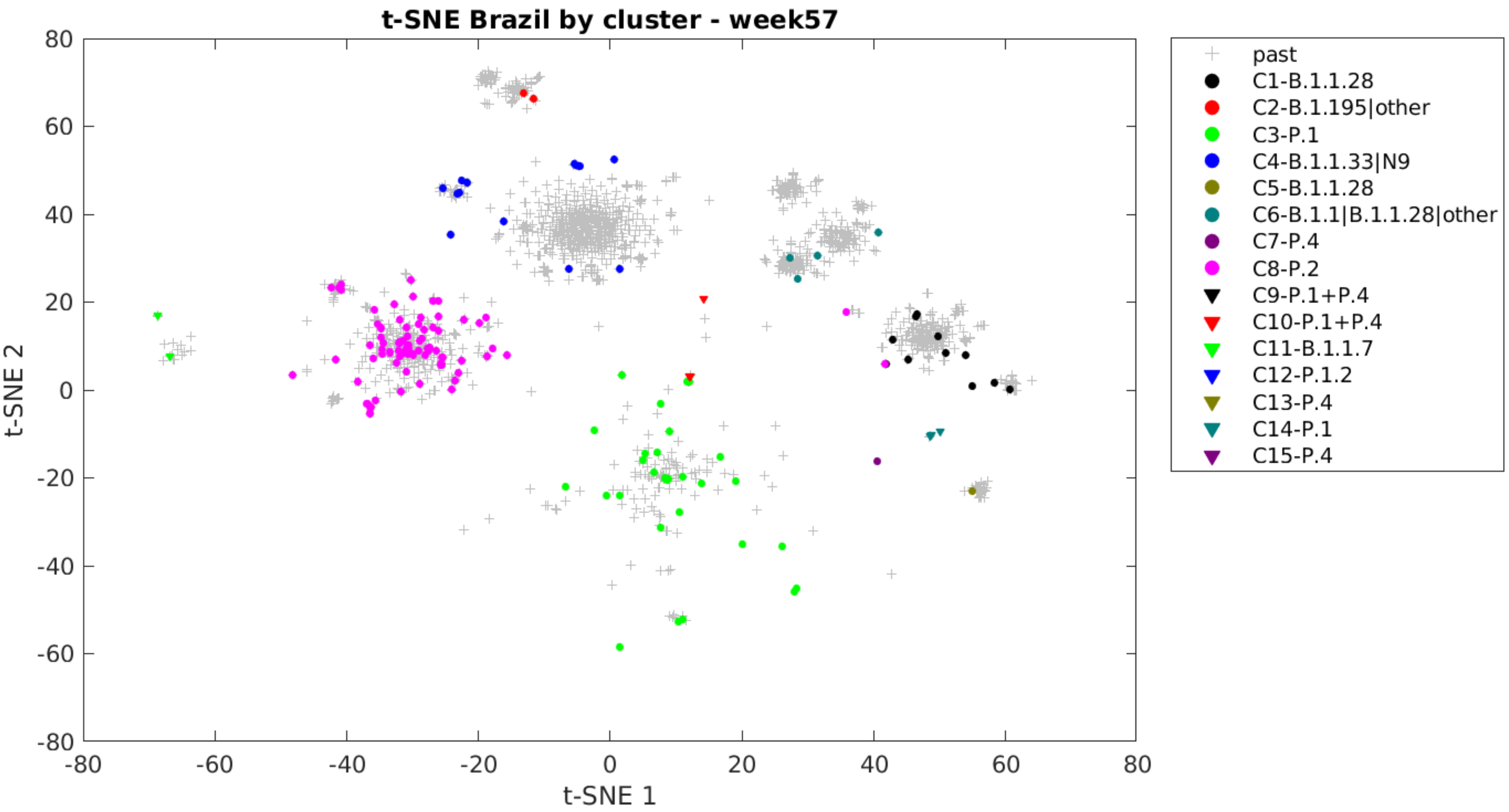

t-SNE Brazil by cluster - week57
t-SNE 1
t-SNE 2
80
60
40
20
0
-20
-40
-60
-80
past
C1-B.1.1.28
C2-B.1.195|other
C3-P.1
C4-B.1.1.33|N9
C5-B.1.1.28
C6-B.1.1|B.1.1.28|other
C7-P.4
C8-P.2
C9-P.1+P.4
C10-P.1+P.4
C11-B.1.1.7
C12-P.1.2
C13-P.4
C14-P.1
C15-P.4

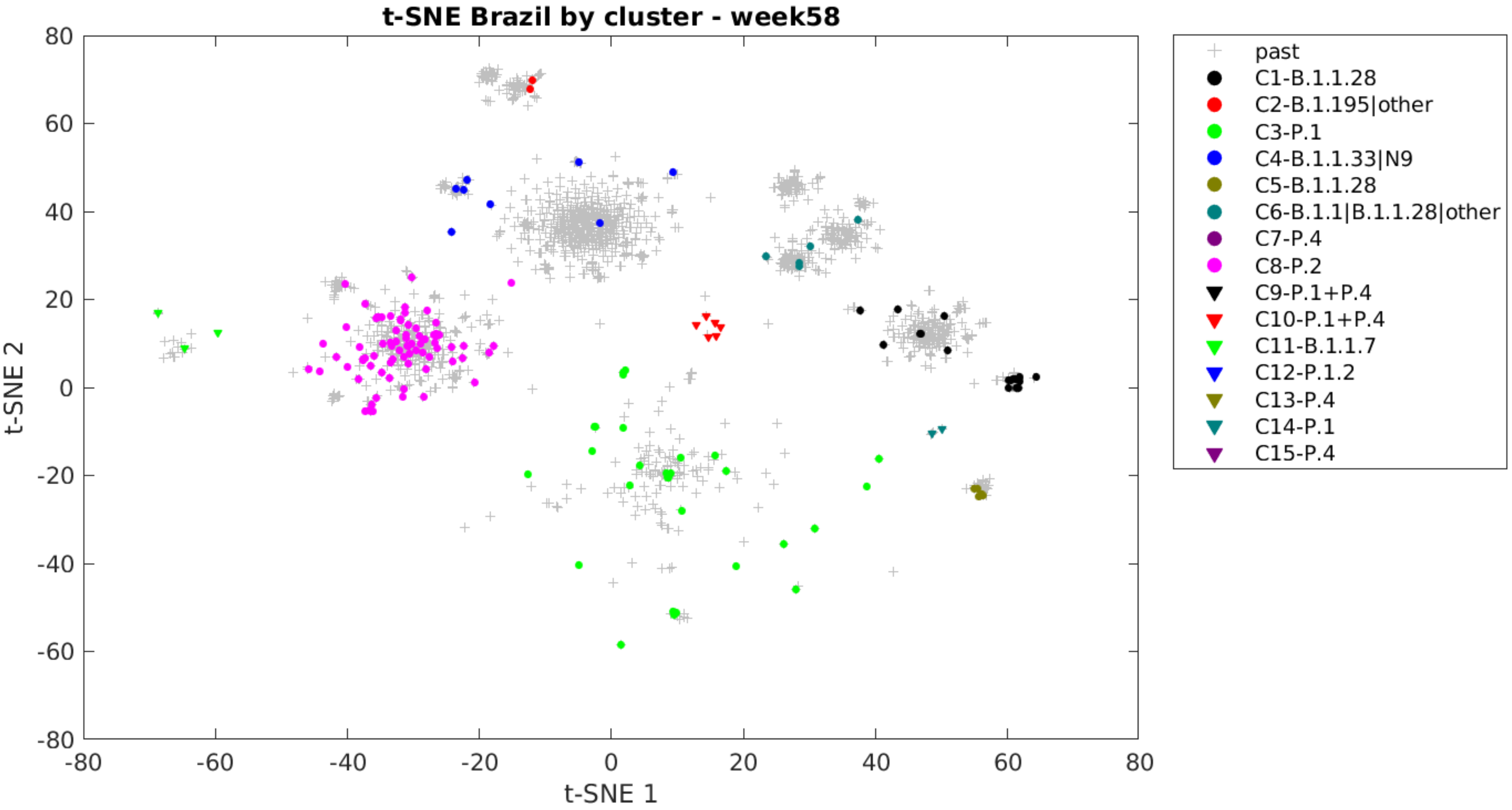

t-SNE Brazil by cluster - week58
t-SNE 2
t-SNE 1
80
60
40
20
0
-20
-40
-60
-80
-80
-60
-40
-20
0
20
40
60
80
past
C1-B.1.1.28
C2-B.1.195|other
C3-P.1
C4-B.1.1.33|N9
C5-B.1.1.28
C6-B.1.1|B.1.1.28|other
C7-P.4
C8-P.2
C9-P.1+P.4
C10-P.1+P.4
C11-B.1.1.7
C12-P.1.2
C13-P.4
C14-P.1
C15-P.4

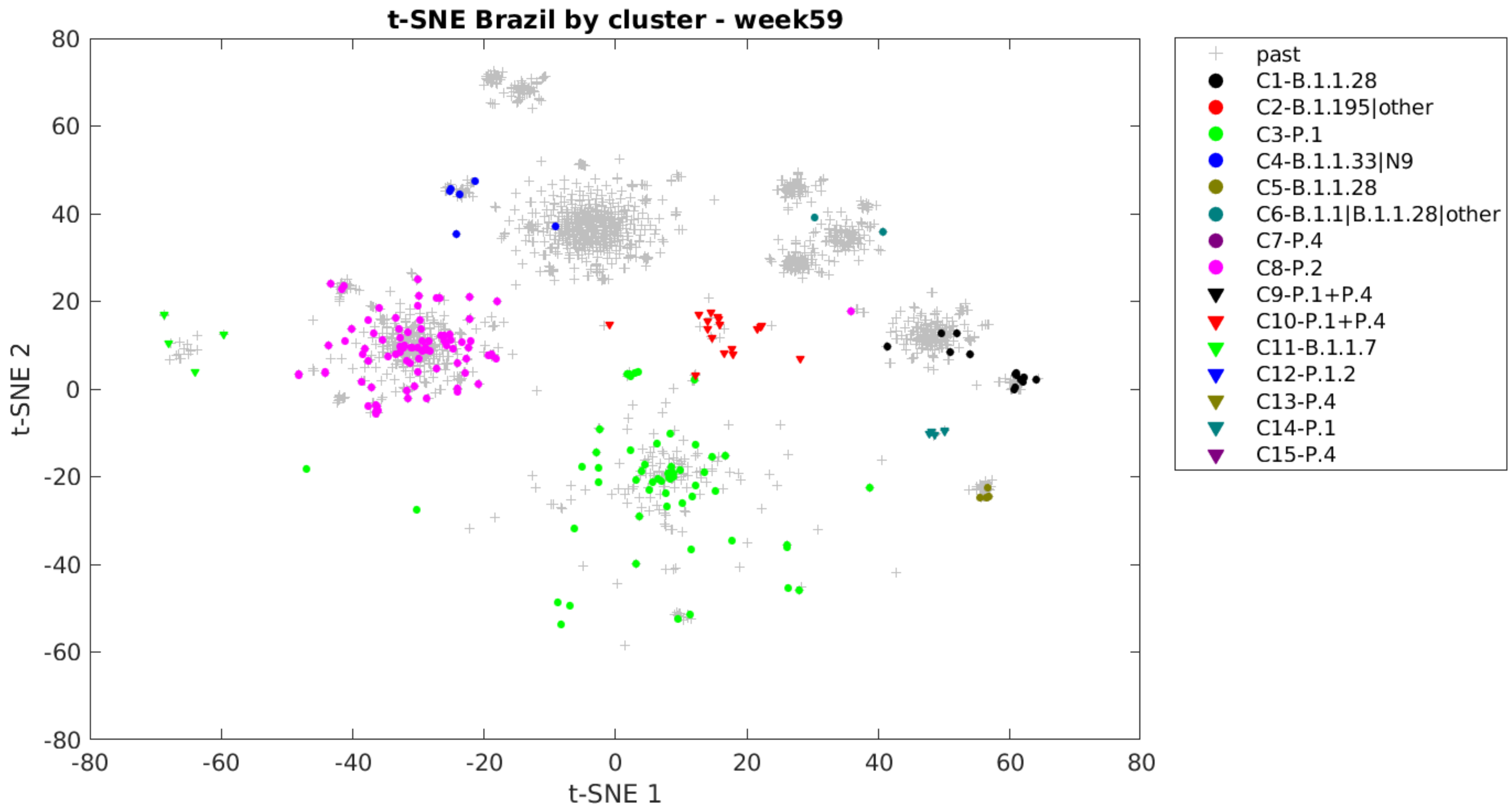
t-SNE Brazil by cluster - week59
t-SNE 1
t-SNE 2
past
C1-B.1.1.28
C2-B.1.195|other
C3-P.1
C4-B.1.1.33|N9
C5-B.1.1.28
C6-B.1.1|B.1.1.28|other
C7-P.4
C8-P.2
C9-P.1+P.4
C10-P.1+P.4
C11-B.1.1.7
C12-P.1.2
C13-P.4
C14-P.1
C15-P.4

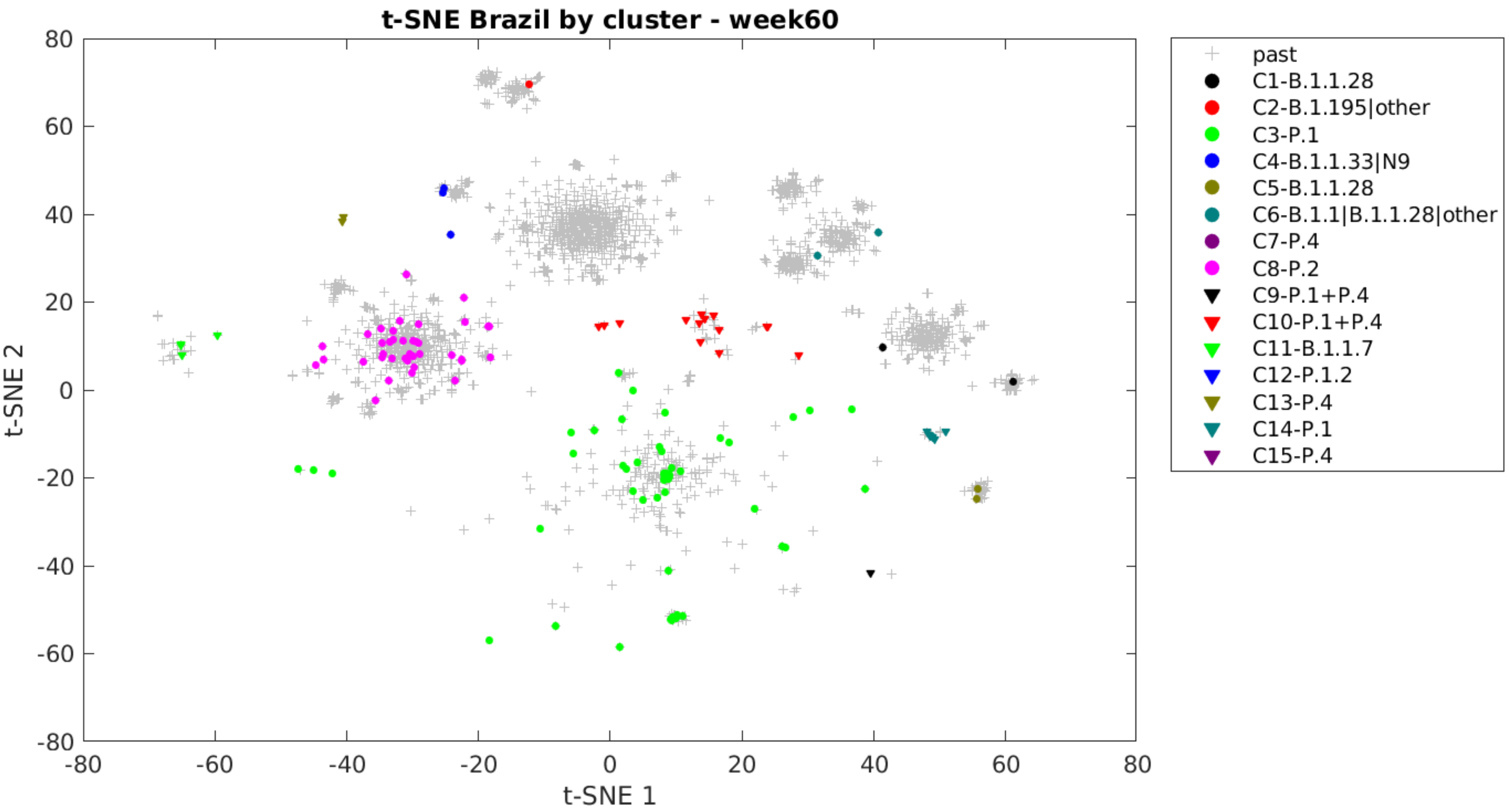
t-SNE Brazil by cluster - week60
t-SNE 2
t-SNE 1
80
60
40
20
0
-20
-40
-60
-80
-80
-60
-40
-20
0
20
40
60
80
past
C1-B.1.1.28
C2-B.1.195|other
C3-P.1
C4-B.1.1.33|N9
C5-B.1.1.28
C6-B.1.1|B.1.1.28|other
C7-P.4
C8-P.2
C9-P.1+P.4
C10-P.1+P.4
C11-B.1.1.7
C12-P.1.2
C13-P.4
C14-P.1
C15-P.4

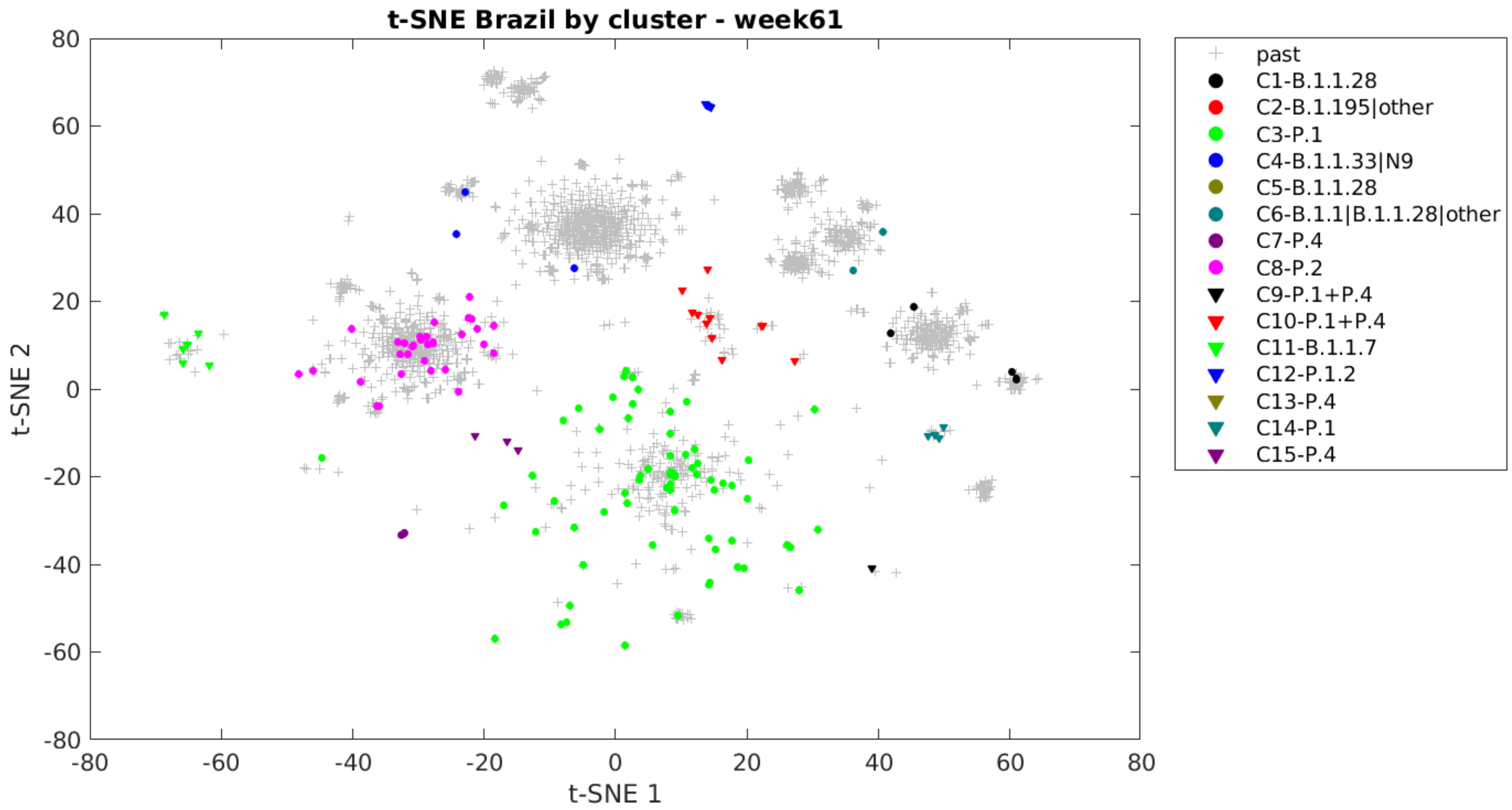

t-SNE Brazil by cluster - week61
t-SNE 1
t-SNE 2
past
C1-B.1.1.28
C2-B.1.195|other
C3-P.1
C4-B.1.1.33|N9
C5-B.1.1.28
C6-B.1.1|B.1.1.28|other
C7-P.4
C8-P.2
C9-P.1+P.4
C10-P.1+P.4
C11-B.1.1.7
C12-P.1.2
C13-P.4
C14-P.1
C15-P.4

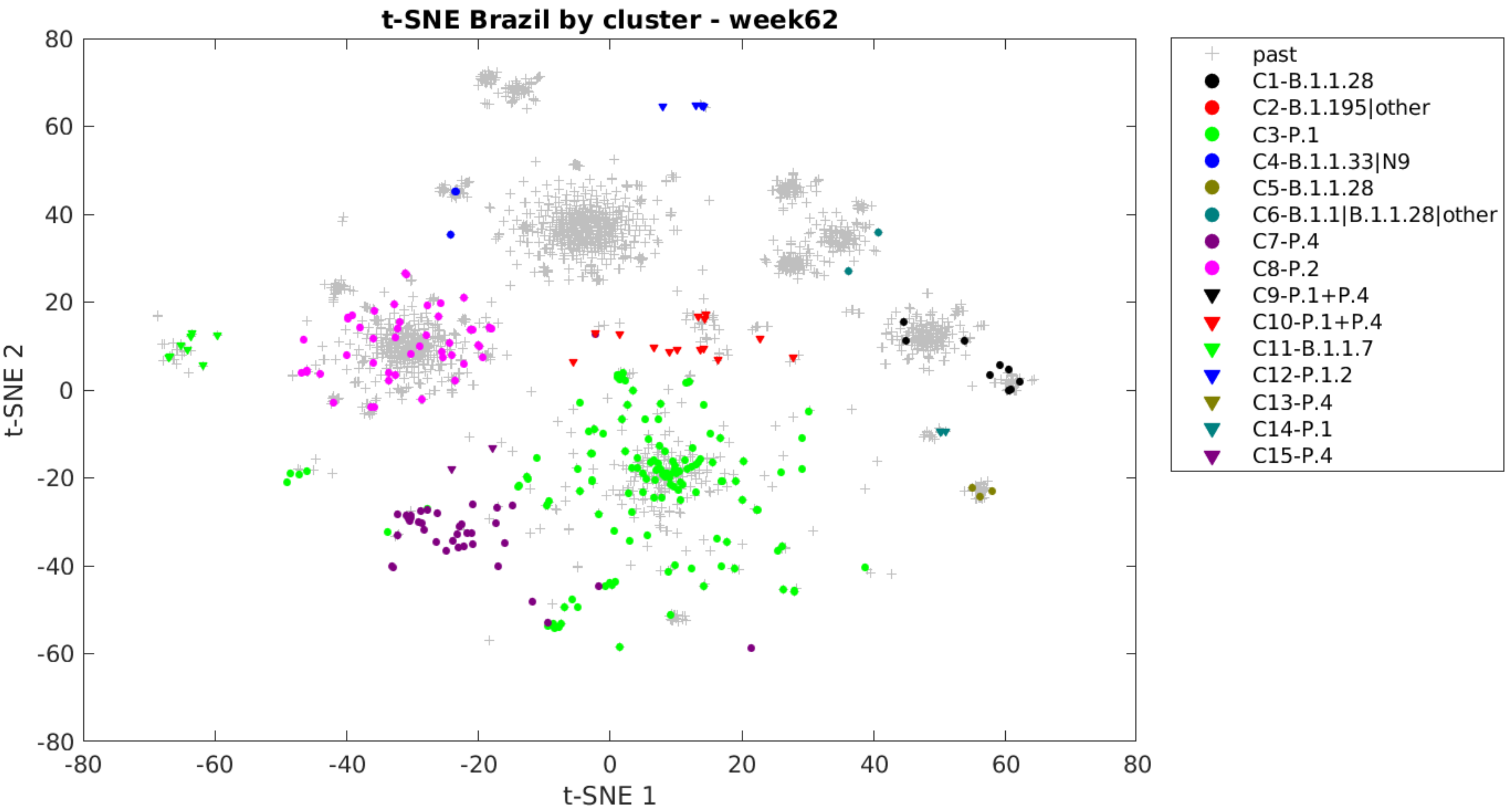

t-SNE Brazil by cluster - week62
t-SNE 1
t-SNE 2
80
60
40
20
0
-20
-40
-60
-80
past
C1-B.1.1.28
C2-B.1.195|other
C3-P.1
C4-B.1.1.33|N9
C5-B.1.1.28
C6-B.1.1|B.1.1.28|other
C7-P.4
C8-P.2
C9-P.1+P.4
C10-P.1+P.4
C11-B.1.1.7
C12-P.1.2
C13-P.4
C14-P.1
C15-P.4

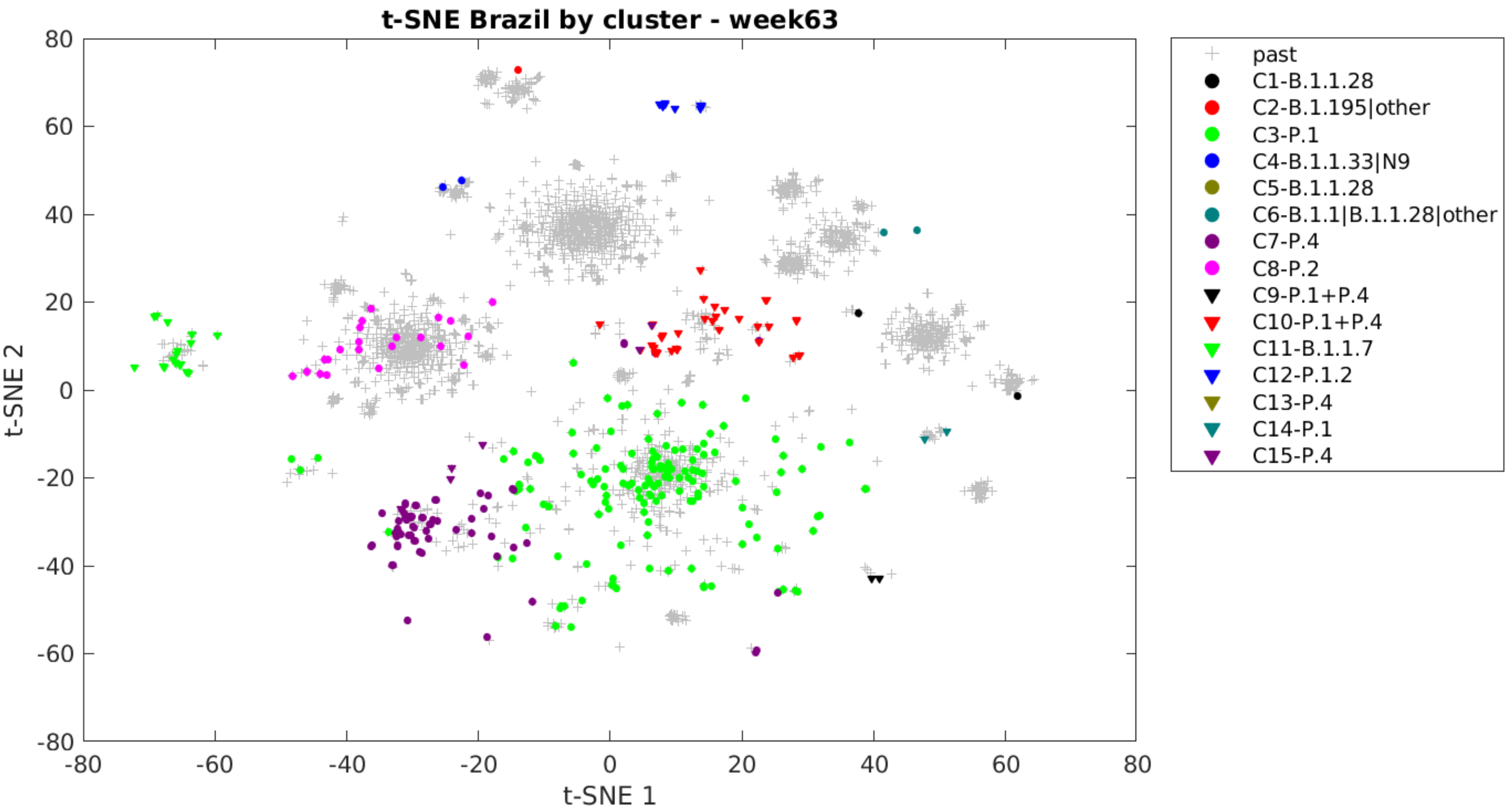

t-SNE Brazil by cluster - week63
t-SNE 1
t-SNE 2
-80
-60
-40
-20
0
20
40
60
80
past
C1-B.1.1.28
C2-B.1.195|other
C3-P.1
C4-B.1.1.33|N9
C5-B.1.1.28
C6-B.1.1|B.1.1.28|other
C7-P.4
C8-P.2
C9-P.1+P.4
C10-P.1+P.4
C11-B.1.1.7
C12-P.1.2
C13-P.4
C14-P.1
C15-P.4

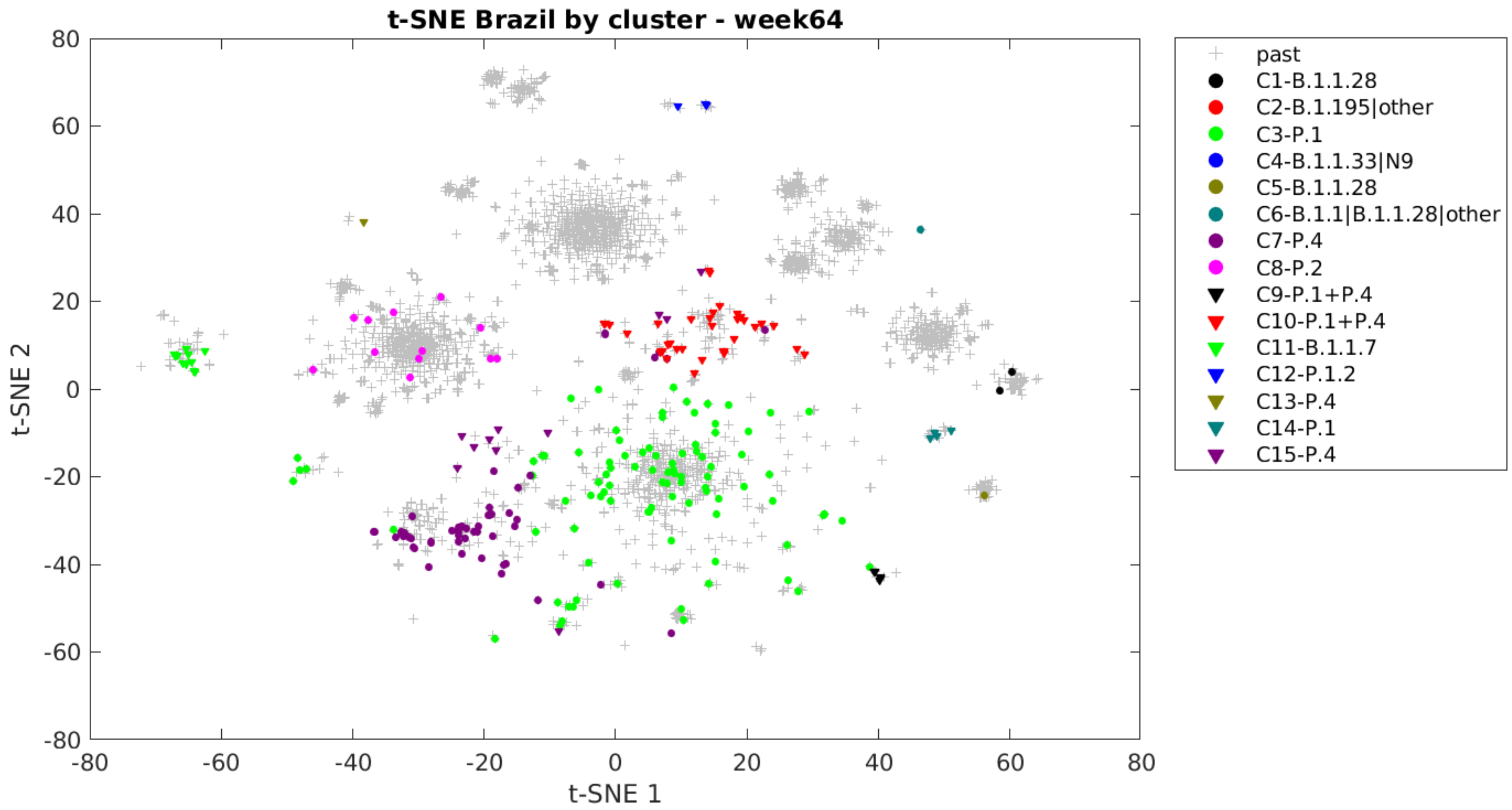

t-SNE Brazil by cluster - week64
t-SNE 2
t-SNE 1
80
60
40
20
0
-20
-40
-60
-80
-80
-60
-40
-20
0
20
40
60
80
past
C1-B.1.1.28
C2-B.1.195|other
C3-P.1
C4-B.1.1.33|N9
C5-B.1.1.28
C6-B.1.1|B.1.1.28|other
C7-P.4
C8-P.2
C9-P.1+P.4
C10-P.1+P.4
C11-B.1.1.7
C12-P.1.2
C13-P.4
C14-P.1
C15-P.4

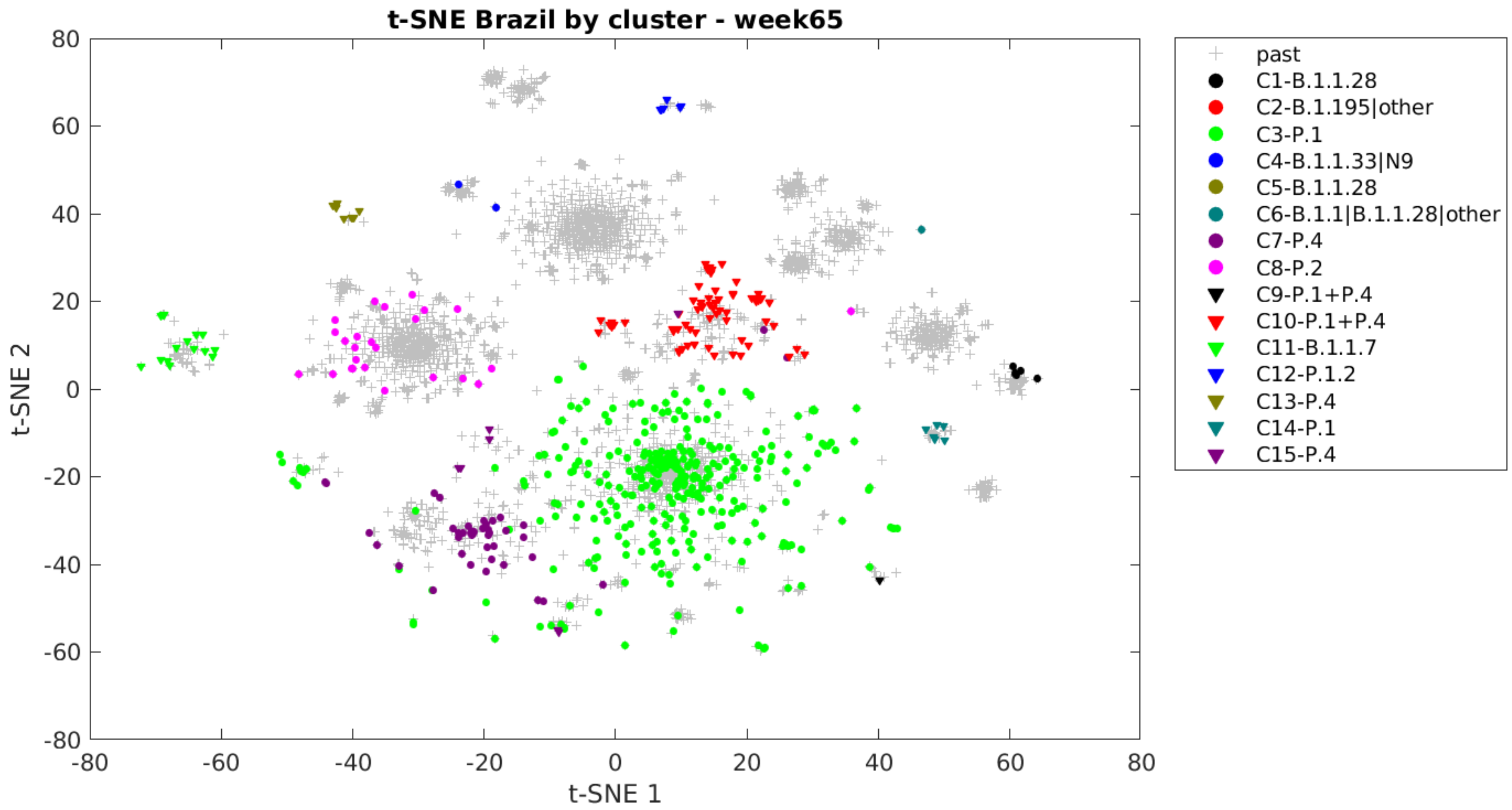

t-SNE Brazil by cluster - week65
80
60
40
20
0
-20
-40
-60
-80
-80
-60
-40
-20
0
20
40
60
80
t-SNE 1
t-SNE 2
past
C1-B.1.1.28
C2-B.1.195|other
C3-P.1
C4-B.1.1.33|N9
C5-B.1.1.28
C6-B.1.1|B.1.1.28|other
C7-P.4
C8-P.2
C9-P.1+P.4
C10-P.1+P.4
C11-B.1.1.7
C12-P.1.2
C13-P.4
C14-P.1
C15-P.4

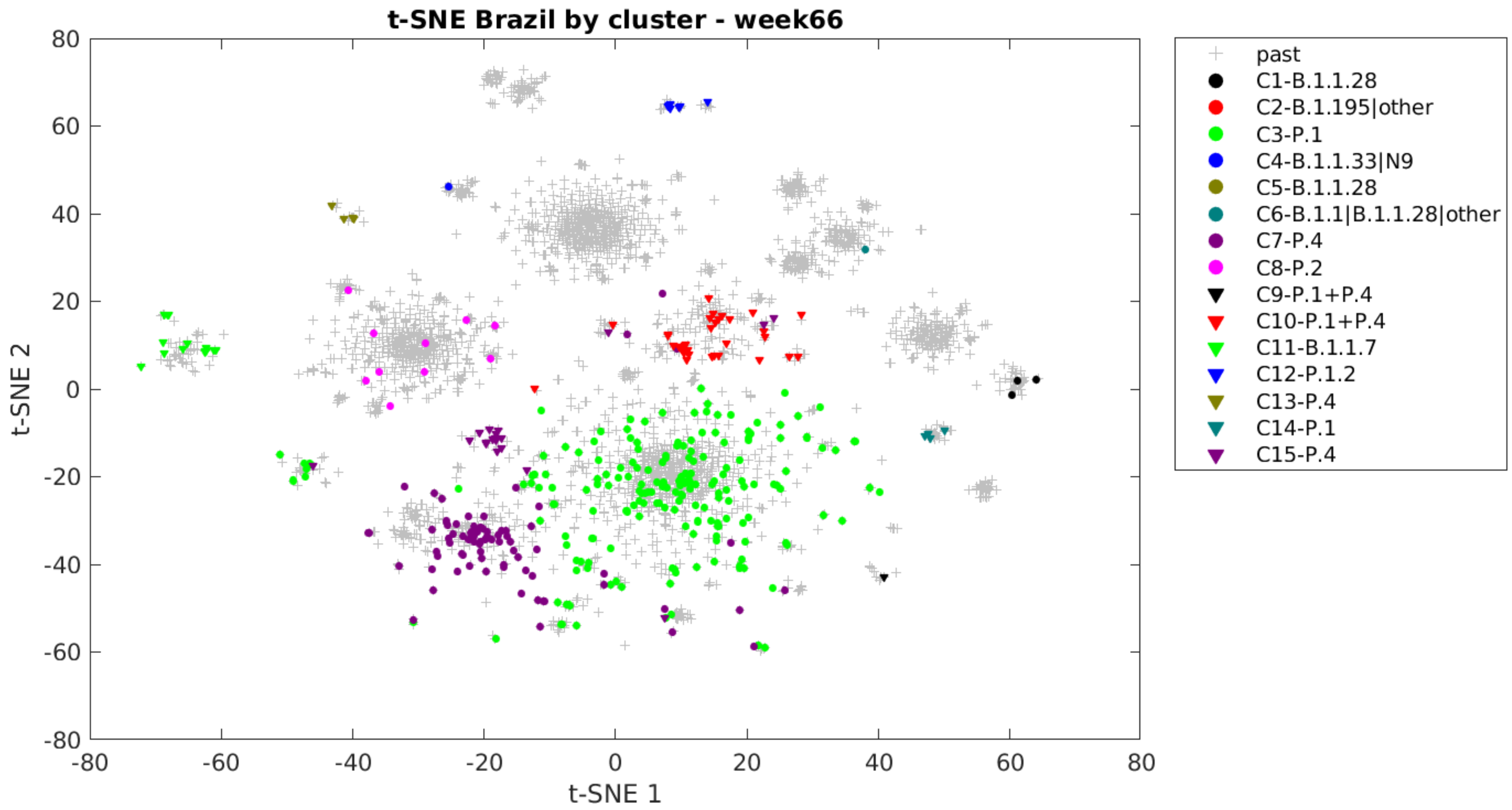

t-SNE Brazil by cluster - week66
t-SNE 1
t-SNE 2
80
60
40
20
0
-20
-40
-60
-80
past
C1-B.1.1.28
C2-B.1.195|other
C3-P.1
C4-B.1.1.33|N9
C5-B.1.1.28
C6-B.1.1|B.1.1.28|other
C7-P.4
C8-P.2
C9-P.1+P.4
C10-P.1+P.4
C11-B.1.1.7
C12-P.1.2
C13-P.4
C14-P.1
C15-P.4

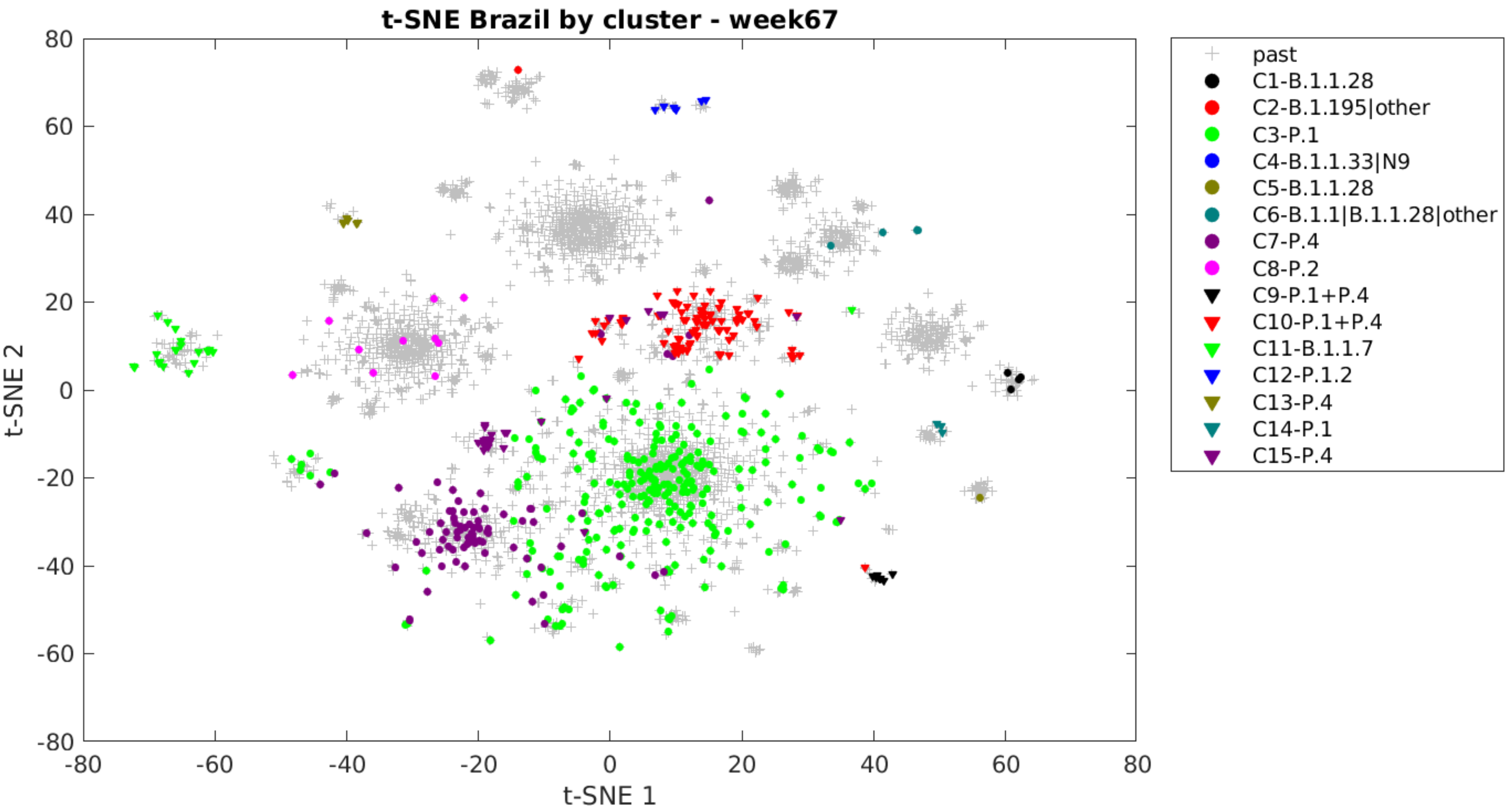
t-SNE Brazil by cluster - week67
t-SNE 1
t-SNE 2
80
60
40
20
0
-20
-40
-60
-80
-80
-60
-40
-20
0
20
40
60
80
past
C1-B.1.1.28
C2-B.1.195|other
C3-P.1
C4-B.1.1.33|N9
C5-B.1.1.28
C6-B.1.1|B.1.1.28|other
C7-P.4
C8-P.2
C9-P.1+P.4
C10-P.1+P.4
C11-B.1.1.7
C12-P.1.2
C13-P.4
C14-P.1
C15-P.4

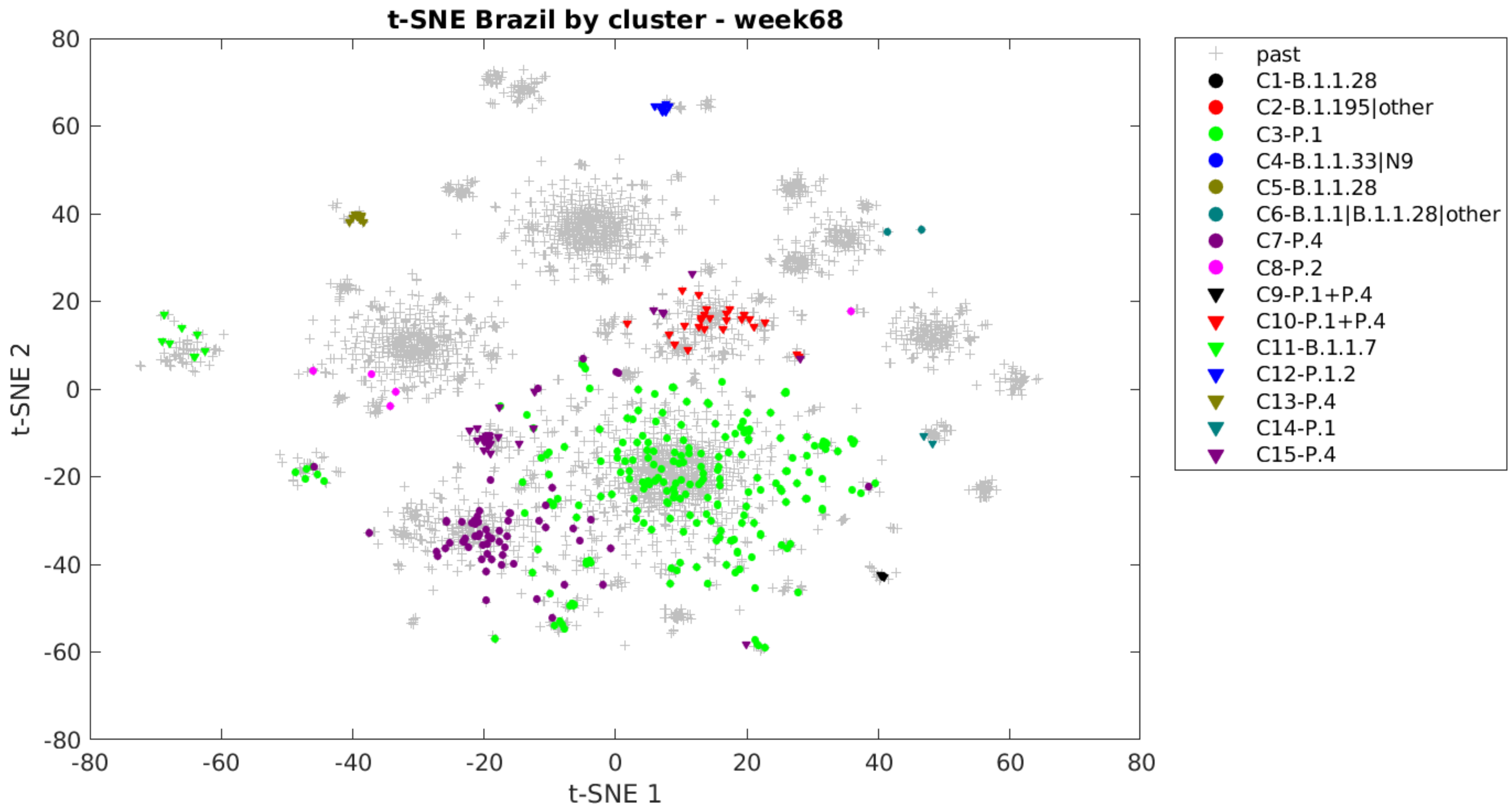

t-SNE Brazil by cluster - week68
t-SNE 2
t-SNE 1
80
60
40
20
0
-20
-40
-60
-80
-80
-60
-40
-20
0
20
40
60
80
past
C1-B.1.1.28
C2-B.1.195|other
C3-P.1
C4-B.1.1.33|N9
C5-B.1.1.28
C6-B.1.1|B.1.1.28|other
C7-P.4
C8-P.2
C9-P.1+P.4
C10-P.1+P.4
C11-B.1.1.7
C12-P.1.2
C13-P.4
C14-P.1
C15-P.4

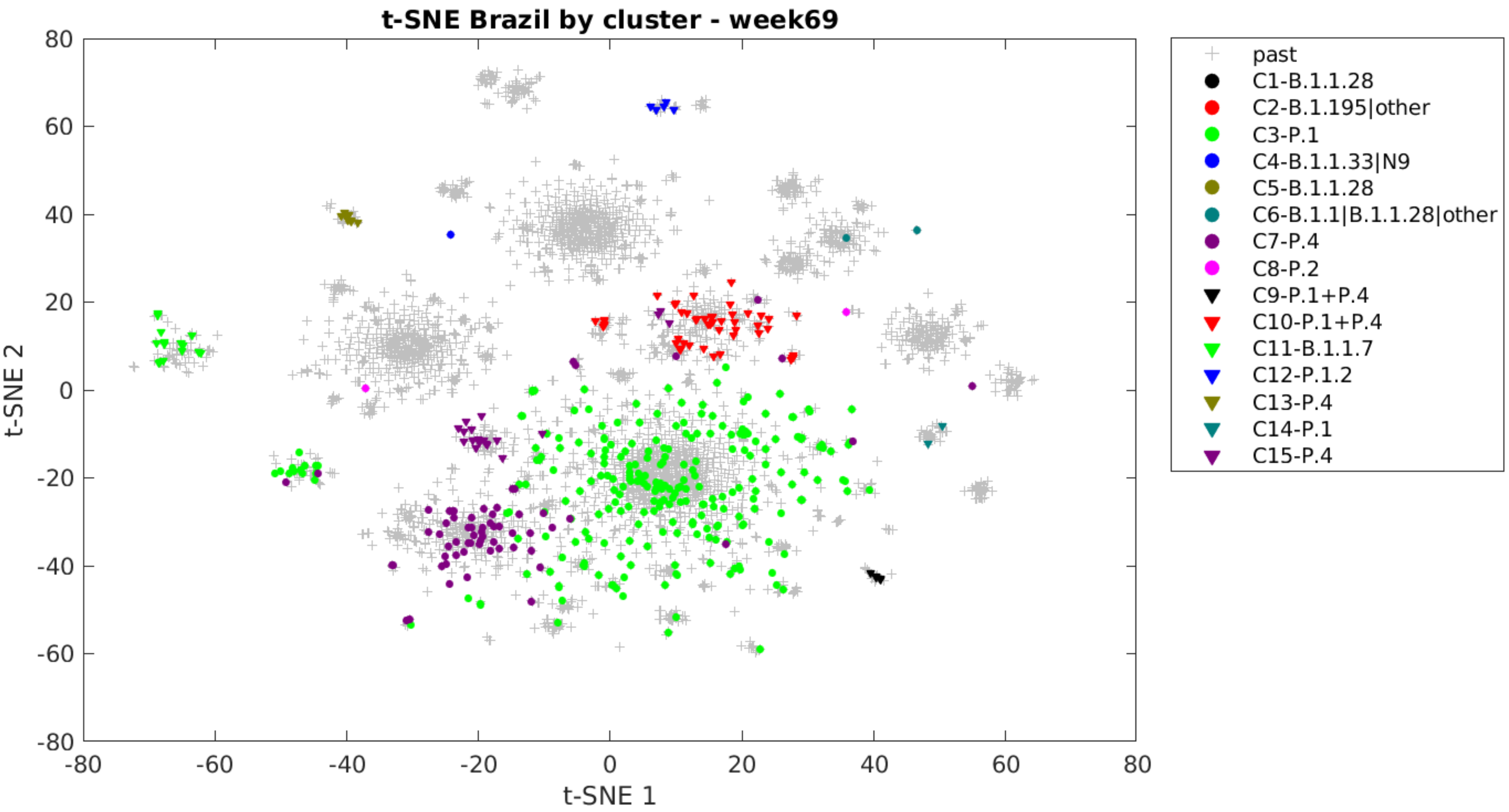

t-SNE Brazil by cluster - week69
t-SNE 1
t-SNE 2
-80
-60
-40
-20
0
20
40
60
80
past
C1-B.1.1.28
C2-B.1.195|other
C3-P.1
C4-B.1.1.33|N9
C5-B.1.1.28
C6-B.1.1|B.1.1.28|other
C7-P.4
C8-P.2
C9-P.1+P.4
C10-P.1+P.4
C11-B.1.1.7
C12-P.1.2
C13-P.4
C14-P.1
C15-P.4

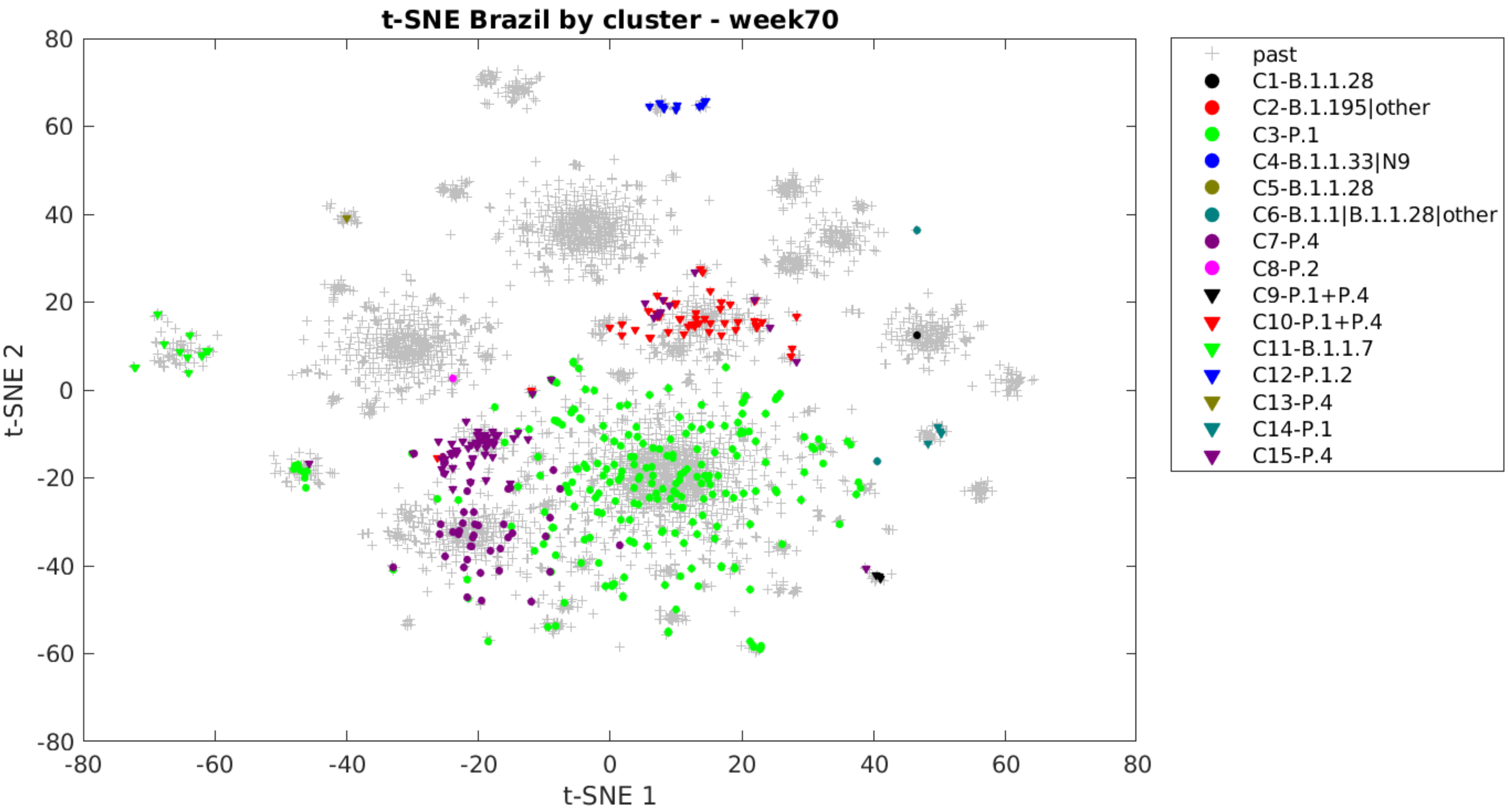
t-SNE Brazil by cluster - week70
t-SNE 1
t-SNE 2
80
60
40
20
0
-20
-40
-60
-80
past
C1-B.1.1.28
C2-B.1.195|other
C3-P.1
C4-B.1.1.33|N9
C5-B.1.1.28
C6-B.1.1|B.1.1.28|other
C7-P.4
C8-P.2
C9-P.1+P.4
C10-P.1+P.4
C11-B.1.1.7
C12-P.1.2
C13-P.4
C14-P.1
C15-P.4

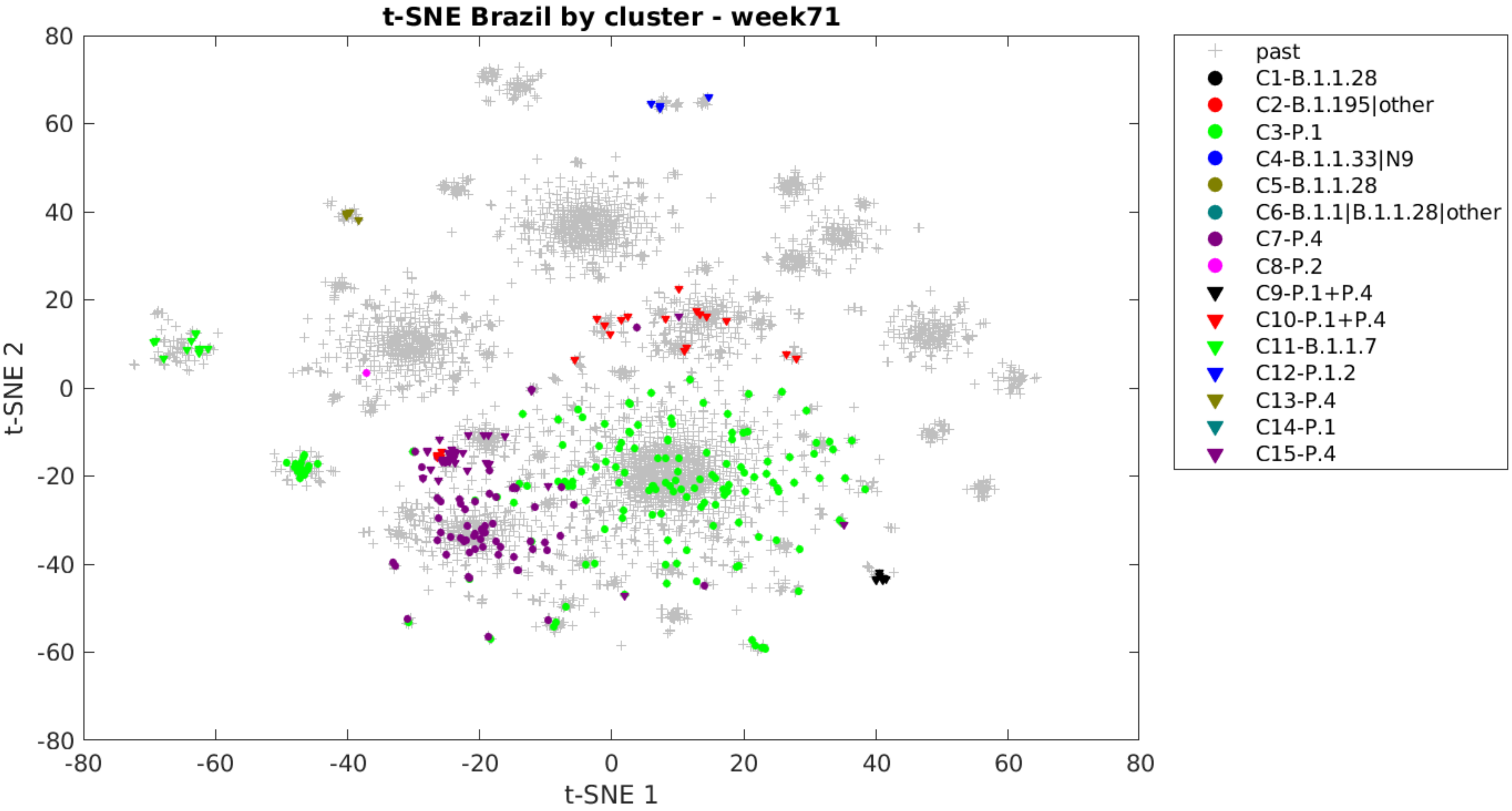
t-SNE Brazil by cluster - week71
t-SNE 1
t-SNE 2
past
C1-B.1.1.28
C2-B.1.195|other
C3-P.1
C4-B.1.1.33|N9
C5-B.1.1.28
C6-B.1.1|B.1.1.28|other
C7-P.4
C8-P.2
C9-P.1+P.4
C10-P.1+P.4
C11-B.1.1.7
C12-P.1.2
C13-P.4
C14-P.1
C15-P.4
