## Supplementary4_PCA_by_week for "Genomic landscape of SARS-CoV-2 pandemic in Brazil suggests an external P.1 variant origin"

November 10, 2021

1. Laboratory of Artificial Intelligence Applied to Bioinformatics - SEPT, Federal University of Paraná, Curtiba, Paraná, Brazil

2. Graduate Program in Bioinformatics - SEPT, Federal University of Paraná, Curtiba, Paraná, Brazil

3. Department of Biochemistry and Molecular Biology, Federal University of Paraná, Curtiba, Paraná, Brazil

**Corresponding Author:**
Roberto T. Raittz:

Lineage evolution over time through visualization with PCA graph. Each slide adds the samples of a new week. The colors represent each cluster obtained for Brazilian sequences. Here was adopted the nomenclature PANGO v3.0.5. The key shows the main lineages that each cluster represents. The emergence, growth and extinction of the groups is visible, as well as the gradual replacement of the T0 group variants by the TP1 group. Note that the second wave was bigger and started with the emergence of P.1 lineage. The information on the number of COVID-19 cases in Brazil was obtained at the official website https://covid.saude.gov.br/.

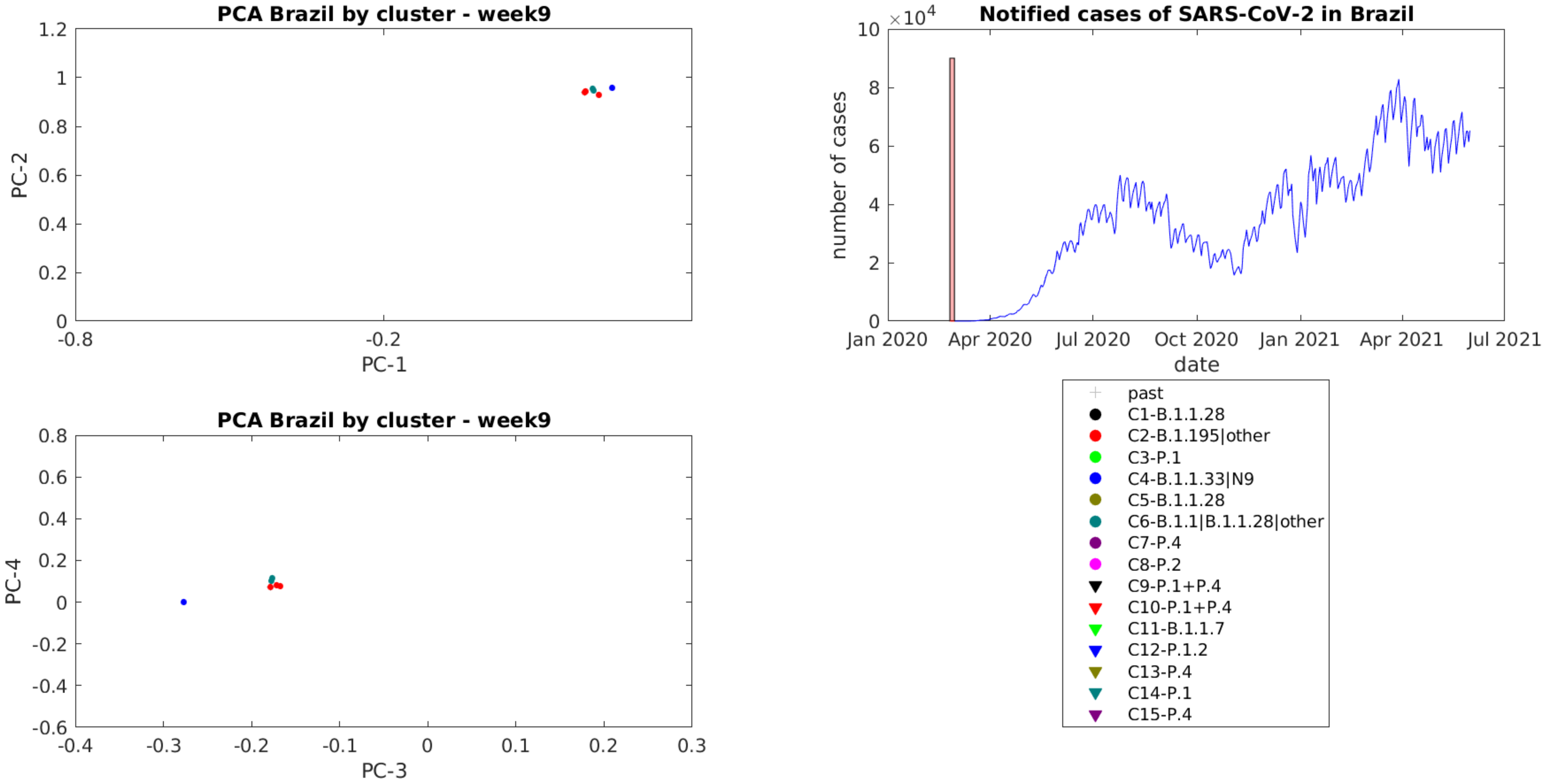
PCA Brazil by cluster - week9
PC-2
PC-1
1.2
1
0.8
0.6
0.4
0.2
0
-0.8
-0.2
PCA Brazil by cluster - week9
PC-4
PC-3
0.8
0.6
0.4
0.2
0
-0.2
-0.4
-0.6
-0.4
-0.3
-0.2
-0.1
0
0.1
0.2
0.3
Notified cases of SARS-CoV-2 in Brazil
×10^4
number of cases
10
8
6
4
2
0
Jan 2020
Apr 2020
Jul 2020
Oct 2020
Jan 2021
Apr 2021
Jul 2021
date
past
C1-B.1.1.28
C2-B.1.195|other
C3-P.1
C4-B.1.1.33|N9
C5-B.1.1.28
C6-B.1.1|B.1.1.28|other
C7-P.4
C8-P.2
C9-P.1+P.4
C10-P.1+P.4
C11-B.1.1.7
C12-P.1.2
C13-P.4
C14-P.1
C15-P.4

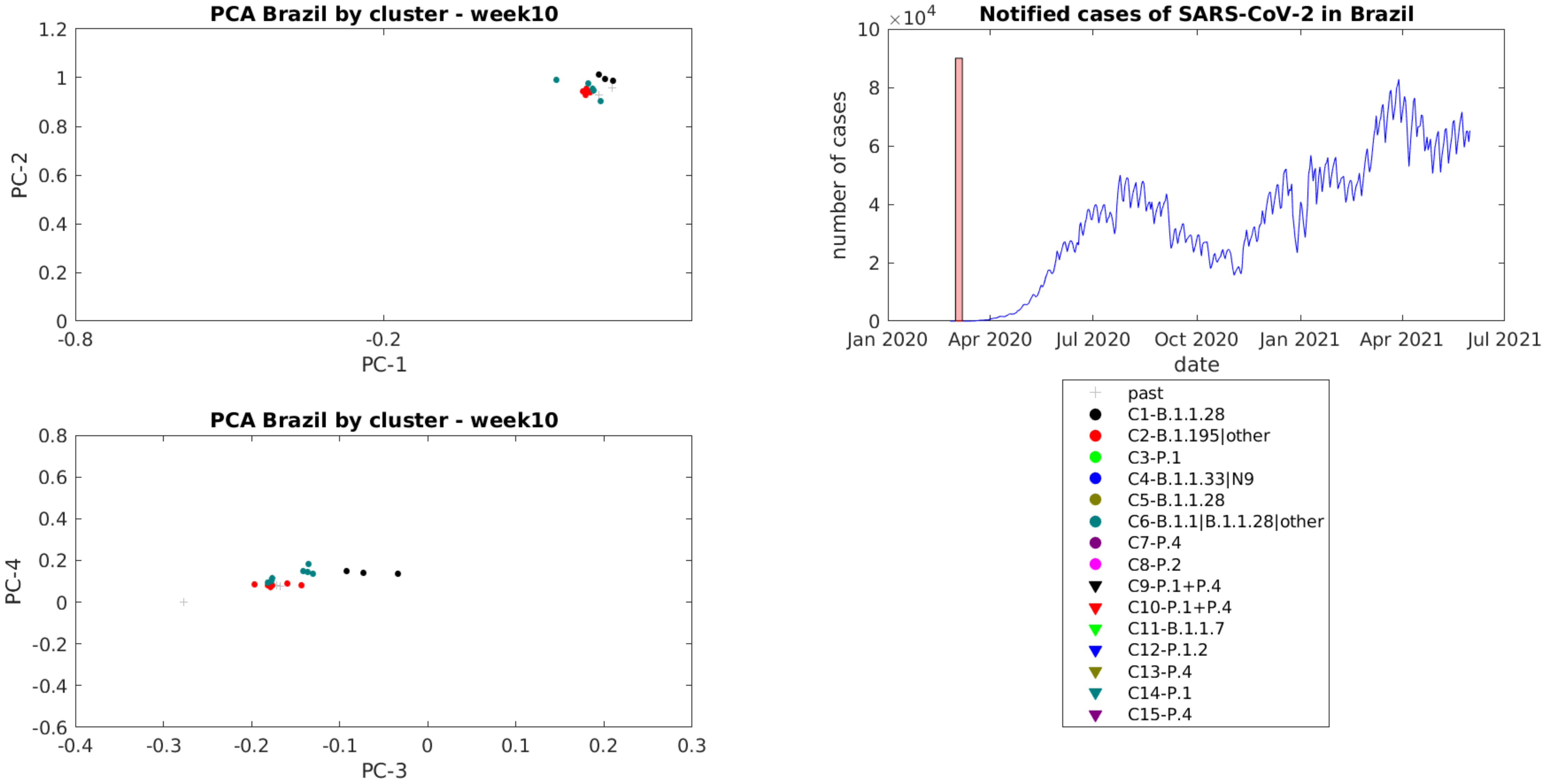
PCA Brazil by cluster - week10
PC-1
PC-2
1.2
1
0.8
0.6
0.4
0.2
0
-0.8
-0.2
PCA Brazil by cluster - week10
PC-3
PC-4
0.8
0.6
0.4
0.2
0
-0.2
-0.4
-0.6
-0.4
-0.3
-0.2
-0.1
0
0.1
0.2
0.3
Notified cases of SARS-CoV-2 in Brazil
×10^4
10
8
6
4
2
0
number of cases
date
Jan 2020
Apr 2020
Jul 2020
Oct 2020
Jan 2021
Apr 2021
Jul 2021
past
C1-B.1.1.28
C2-B.1.195|other
C3-P.1
C4-B.1.1.33|N9
C5-B.1.1.28
C6-B.1.1|B.1.1.28|other
C7-P.4
C8-P.2
C9-P.1+P.4
C10-P.1+P.4
C11-B.1.1.7
C12-P.1.2
C13-P.4
C14-P.1
C15-P.4

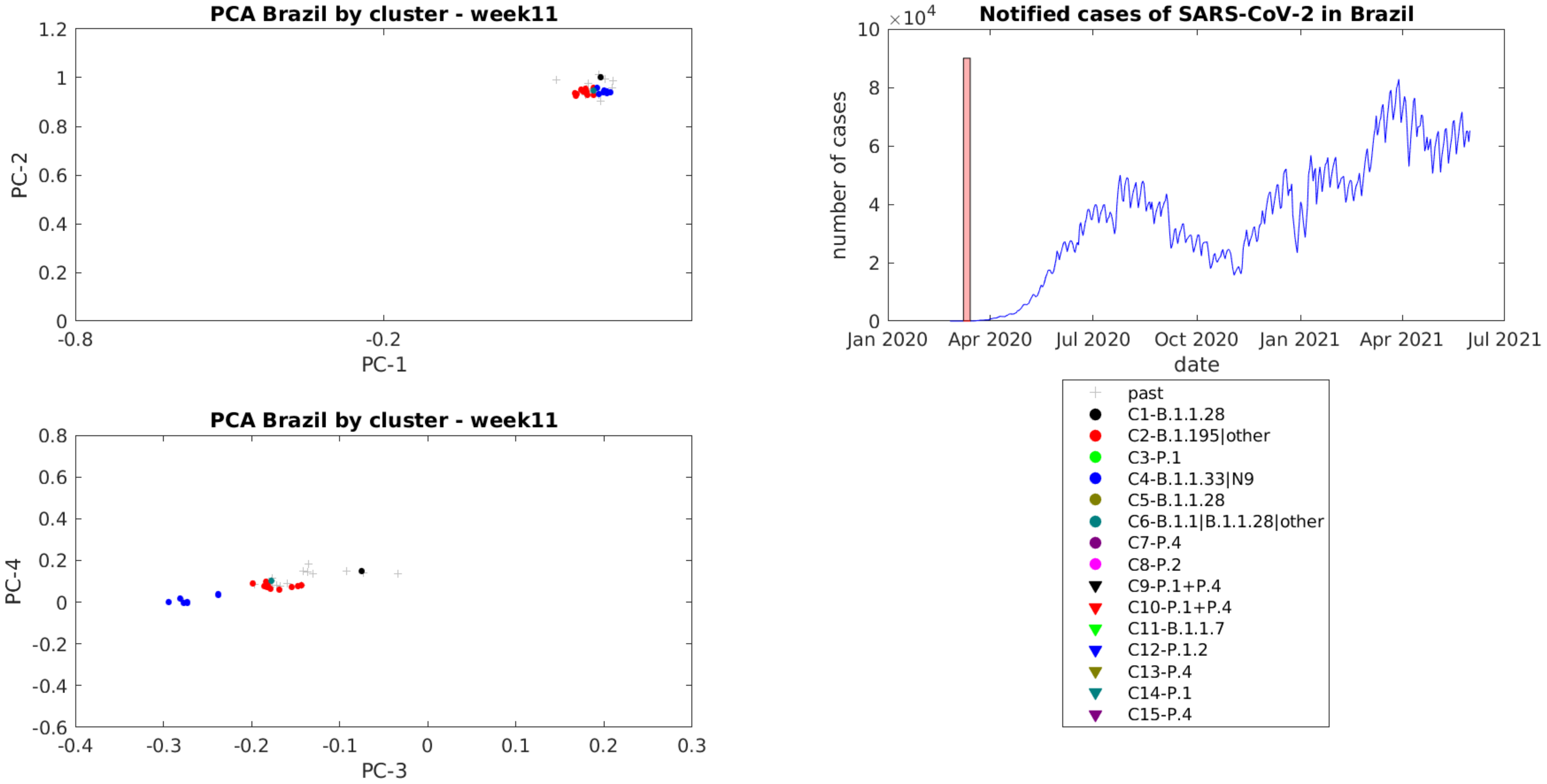
PCA Brazil by cluster - week11
PC-2
PC-1
1.2
1
0.8
0.6
0.4
0.2
0
-0.8
-0.2
PCA Brazil by cluster - week11
PC-4
PC-3
0.8
0.6
0.4
0.2
0
-0.2
-0.4
-0.6
-0.4
-0.3
-0.2
-0.1
0
0.1
0.2
0.3
Notified cases of SARS-CoV-2 in Brazil
×10^4
10
8
6
4
2
0
number of cases
date
Jan 2020
Apr 2020
Jul 2020
Oct 2020
Jan 2021
Apr 2021
Jul 2021
past
C1-B.1.1.28
C2-B.1.195|other
C3-P.1
C4-B.1.1.33|N9
C5-B.1.1.28
C6-B.1.1|B.1.1.28|other
C7-P.4
C8-P.2
C9-P.1+P.4
C10-P.1+P.4
C11-B.1.1.7
C12-P.1.2
C13-P.4
C14-P.1
C15-P.4

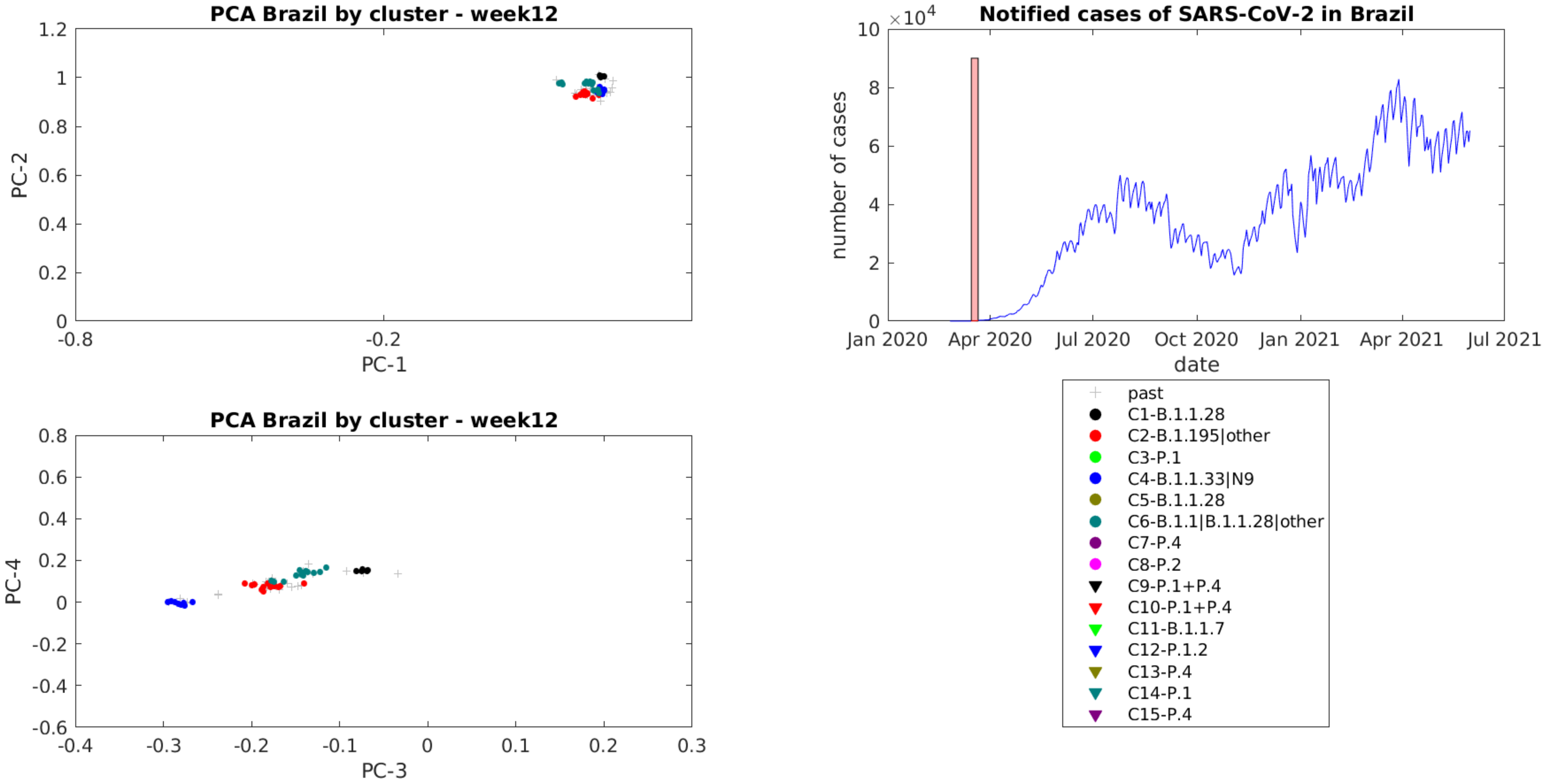
PCA Brazil by cluster - week12
PC-1
PC-2
PCA Brazil by cluster - week12
PC-3
PC-4
Notified cases of SARS-CoV-2 in Brazil
×10^4
number of cases
date
Jan 2020
Apr 2020
Jul 2020
Oct 2020
Jan 2021
Apr 2021
Jul 2021
past
C1-B.1.1.28
C2-B.1.195|other
C3-P.1
C4-B.1.1.33|N9
C5-B.1.1.28
C6-B.1.1|B.1.1.28|other
C7-P.4
C8-P.2
C9-P.1+P.4
C10-P.1+P.4
C11-B.1.1.7
C12-P.1.2
C13-P.4
C14-P.1
C15-P.4

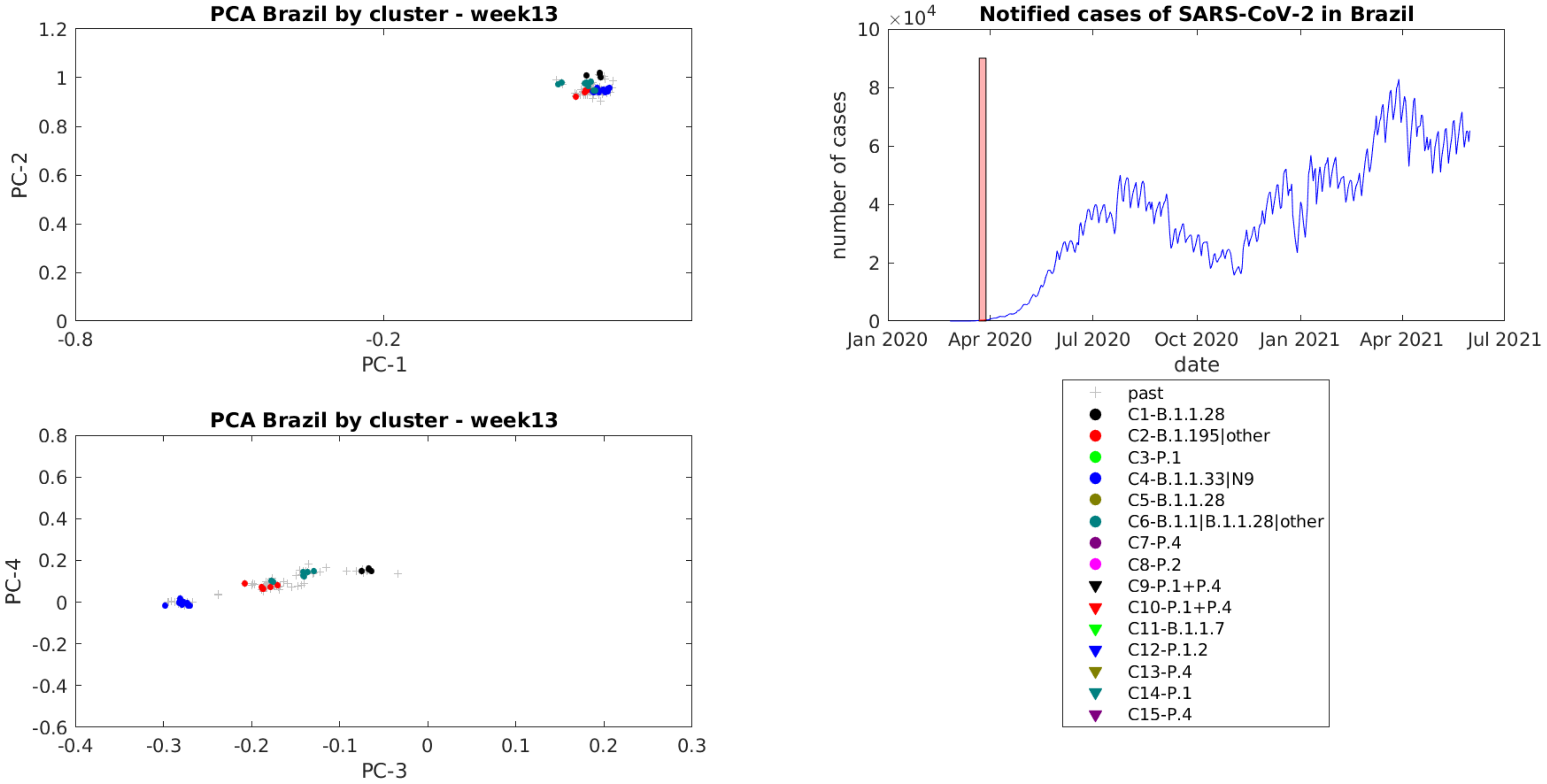

PCA Brazil by cluster - week13
PC-2
PC-1
PCA Brazil by cluster - week13
PC-4
PC-3
Notified cases of SARS-CoV-2 in Brazil
number of cases
date
Jan 2020
Apr 2020
Jul 2020
Oct 2020
Jan 2021
Apr 2021
Jul 2021
past
C1-B.1.1.28
C2-B.1.195|other
C3-P.1
C4-B.1.1.33|N9
C5-B.1.1.28
C6-B.1.1|B.1.1.28|other
C7-P.4
C8-P.2
C9-P.1+P.4
C10-P.1+P.4
C11-B.1.1.7
C12-P.1.2
C13-P.4
C14-P.1
C15-P.4

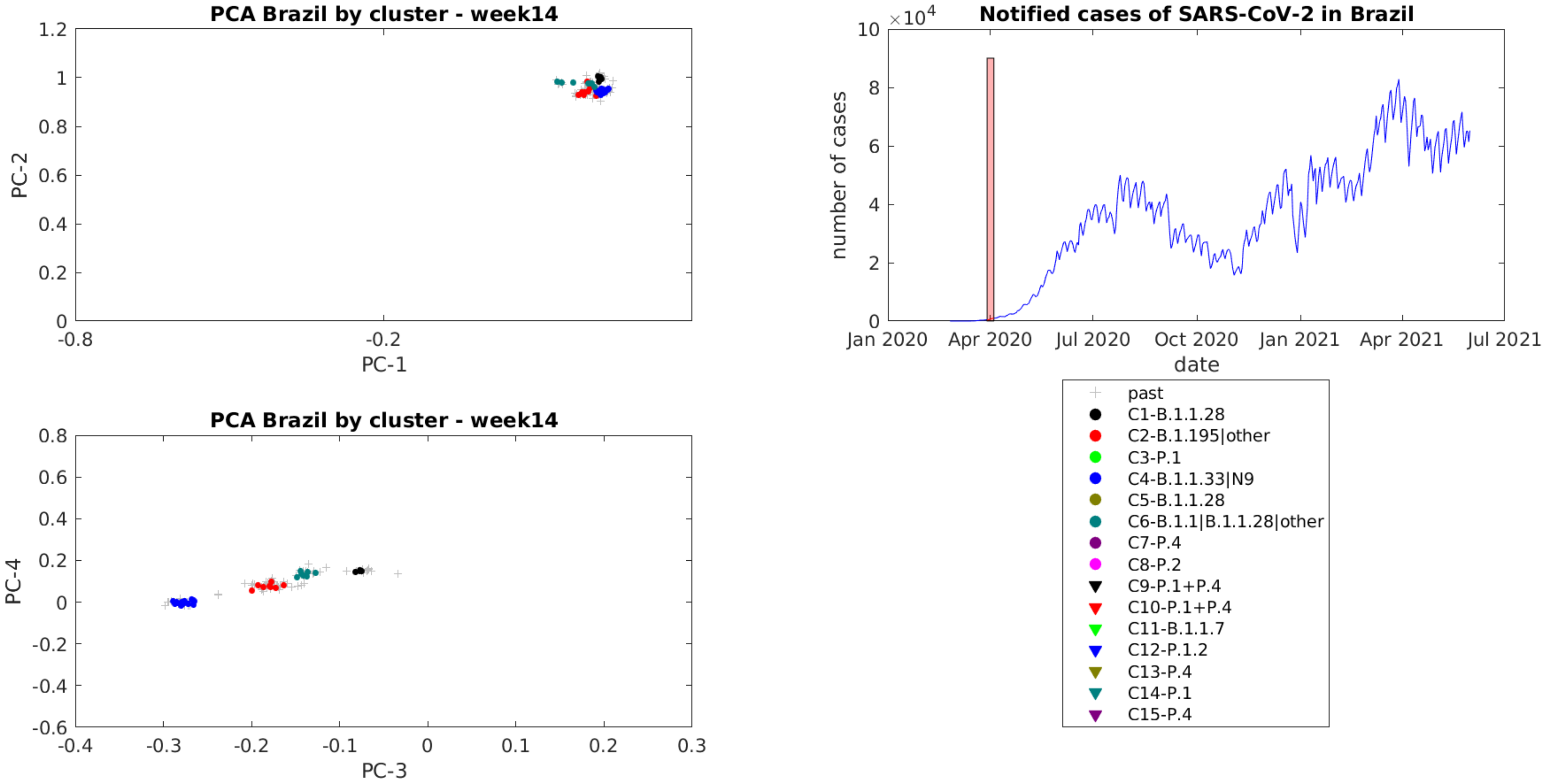

PCA Brazil by cluster - week14
PC-1
PC-2
PCA Brazil by cluster - week14
PC-3
PC-4
Notified cases of SARS-CoV-2 in Brazil
×10^4
number of cases
date
Jan 2020
Apr 2020
Jul 2020
Oct 2020
Jan 2021
Apr 2021
Jul 2021
past
C1-B.1.1.28
C2-B.1.195|other
C3-P.1
C4-B.1.1.33|N9
C5-B.1.1.28
C6-B.1.1|B.1.1.28|other
C7-P.4
C8-P.2
C9-P.1+P.4
C10-P.1+P.4
C11-B.1.1.7
C12-P.1.2
C13-P.4
C14-P.1
C15-P.4

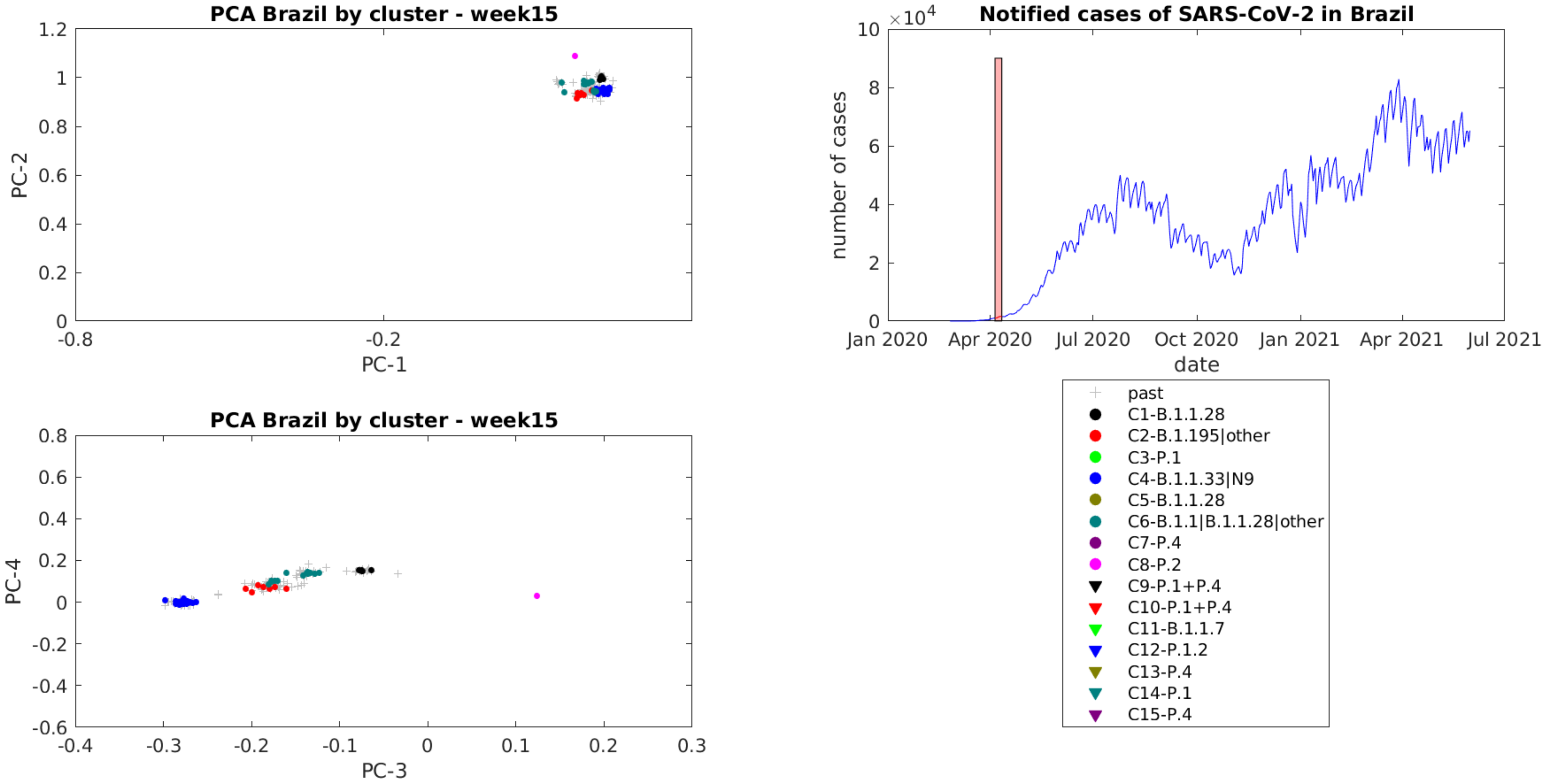

PCA Brazil by cluster - week15
PC-2
PC-1
1.2
1
0.8
0.6
0.4
0.2
0
-0.8
-0.2
Notified cases of SARS-CoV-2 in Brazil
×10^4
number of cases
10
8
6
4
2
0
date
Jan 2020
Apr 2020
Jul 2020
Oct 2020
Jan 2021
Apr 2021
Jul 2021
PCA Brazil by cluster - week15
PC-4
PC-3
0.8
0.6
0.4
0.2
0
-0.2
-0.4
-0.6
-0.4
-0.3
-0.2
-0.1
0
0.1
0.2
0.3
past
C1-B.1.1.28
C2-B.1.195|other
C3-P.1
C4-B.1.1.33|N9
C5-B.1.1.28
C6-B.1.1|B.1.1.28|other
C7-P.4
C8-P.2
C9-P.1+P.4
C10-P.1+P.4
C11-B.1.1.7
C12-P.1.2
C13-P.4
C14-P.1
C15-P.4

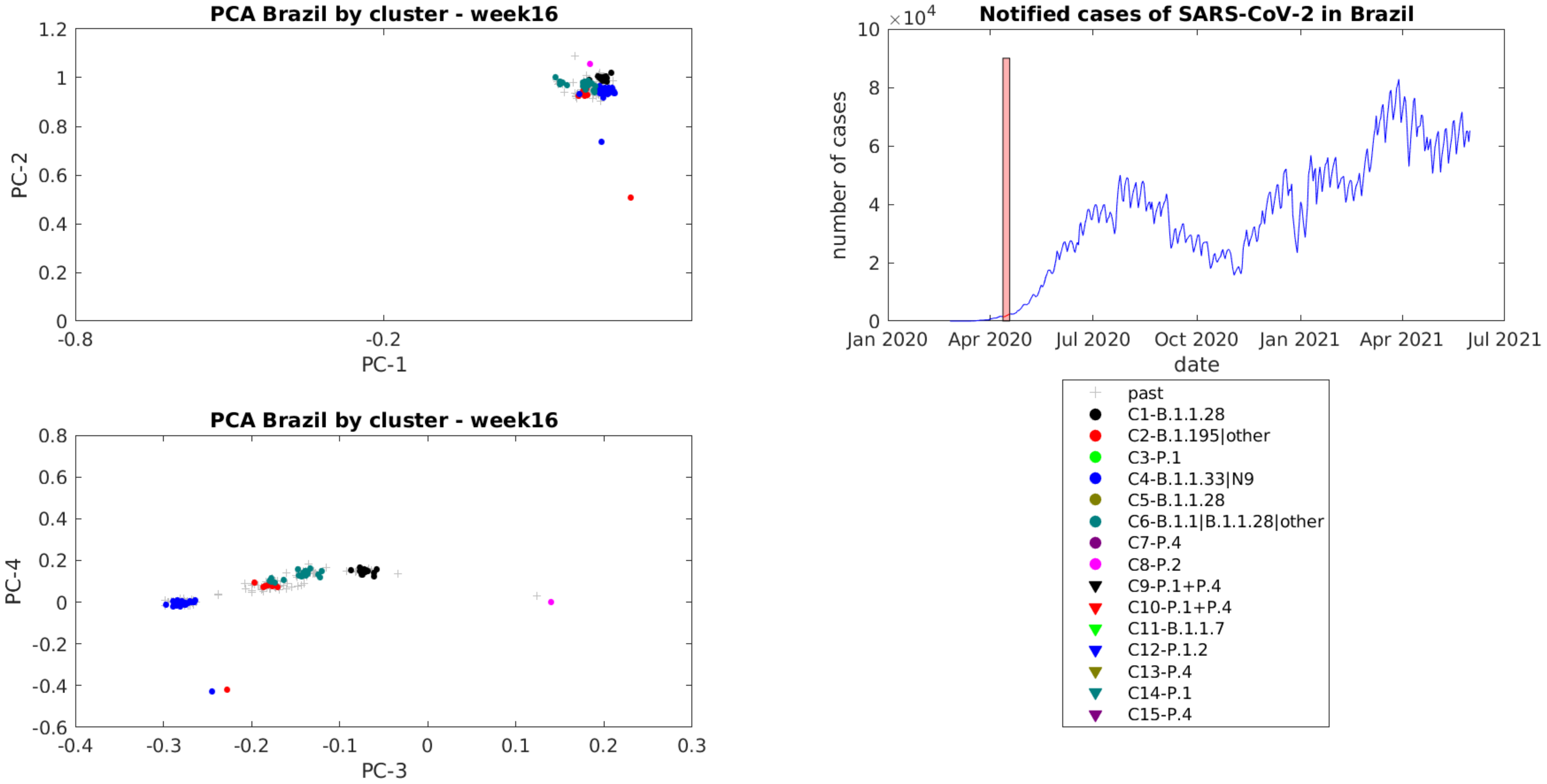
PCA Brazil by cluster - week16
PC-1
PC-2
-0.8
-0.2
0
0.2
0.4
0.6
0.8
1
1.2
Notified cases of SARS-CoV-2 in Brazil
$\times 10^4$
number of cases
date
0
2
4
6
8
10
Jan 2020
Apr 2020
Jul 2020
Oct 2020
Jan 2021
Apr 2021
Jul 2021
PCA Brazil by cluster - week16
PC-3
PC-4
-0.4
-0.3
-0.2
-0.1
0
0.1
0.2
0.3
-0.6
-0.4
-0.2
0
0.2
0.4
0.6
0.8
past
C1-B.1.1.28
C2-B.1.195|other
C3-P.1
C4-B.1.1.33|N9
C5-B.1.1.28
C6-B.1.1|B.1.1.28|other
C7-P.4
C8-P.2
C9-P.1+P.4
C10-P.1+P.4
C11-B.1.1.7
C12-P.1.2
C13-P.4
C14-P.1
C15-P.4

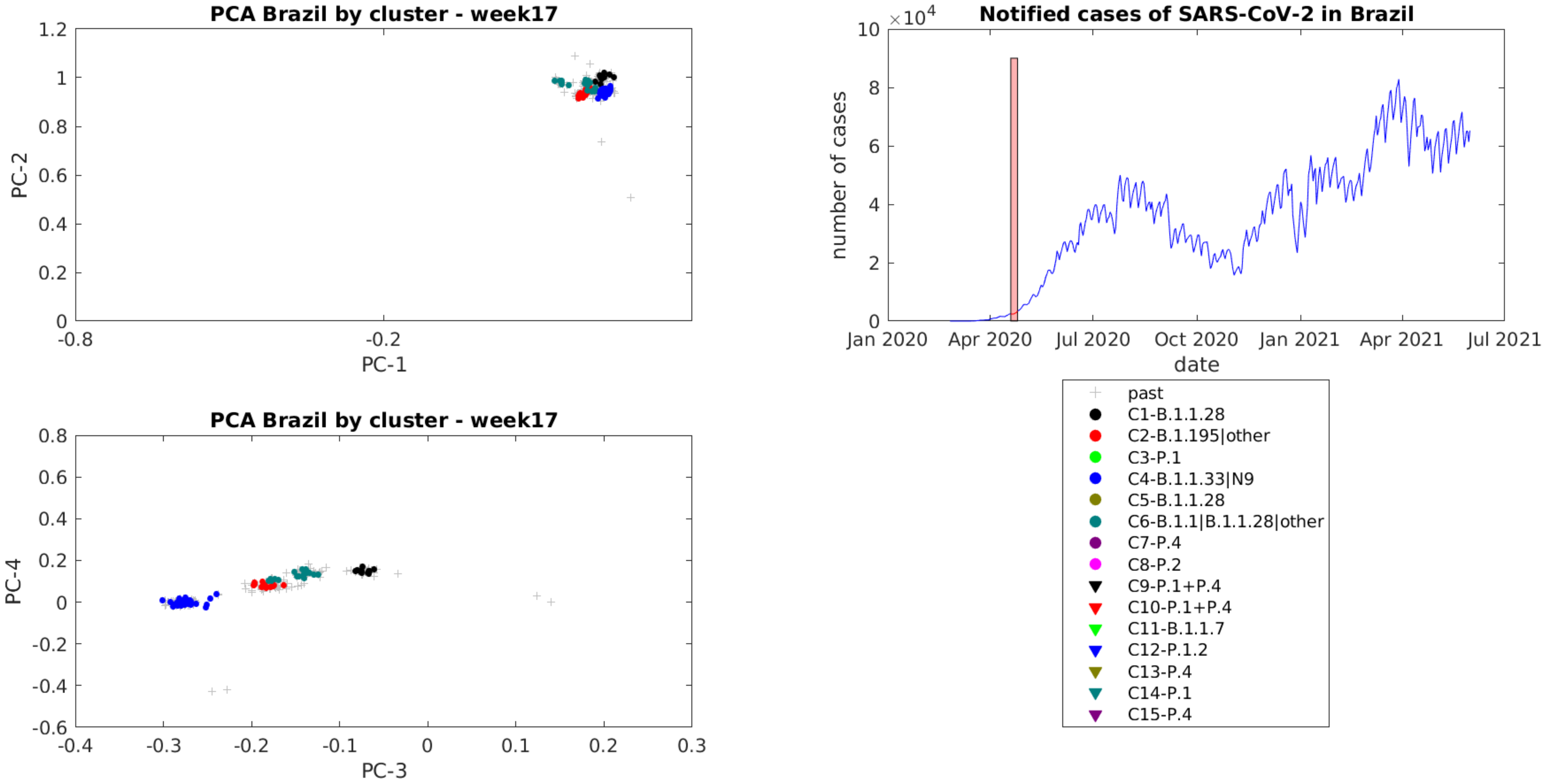
PCA Brazil by cluster - week17
PC-1
PC-2
Notified cases of SARS-CoV-2 in Brazil
number of cases
date
Jan 2020
Apr 2020
Jul 2020
Oct 2020
Jan 2021
Apr 2021
Jul 2021
PCA Brazil by cluster - week17
PC-3
PC-4
past
C1-B.1.1.28
C2-B.1.195|other
C3-P.1
C4-B.1.1.33|N9
C5-B.1.1.28
C6-B.1.1|B.1.1.28|other
C7-P.4
C8-P.2
C9-P.1+P.4
C10-P.1+P.4
C11-B.1.1.7
C12-P.1.2
C13-P.4
C14-P.1
C15-P.4

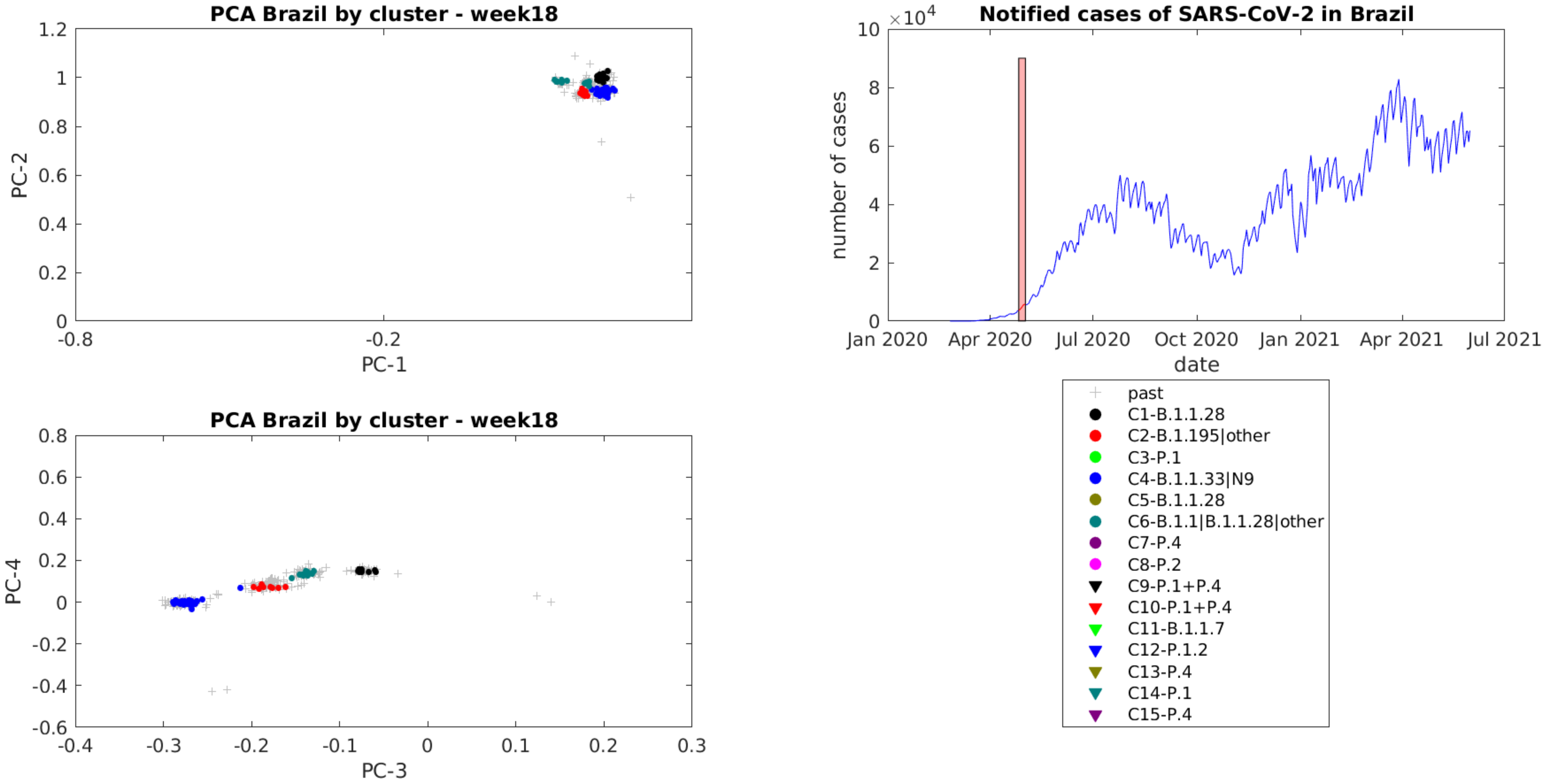
PCA Brazil by cluster - week18
PC-2
PC-1
1.2
1
0.8
0.6
0.4
0.2
0
-0.8
-0.2
PCA Brazil by cluster - week18
PC-4
PC-3
0.8
0.6
0.4
0.2
0
-0.2
-0.4
-0.6
-0.4
-0.3
-0.2
-0.1
0
0.1
0.2
0.3
Notified cases of SARS-CoV-2 in Brazil
×10^4
number of cases
date
10
8
6
4
2
0
Jan 2020
Apr 2020
Jul 2020
Oct 2020
Jan 2021
Apr 2021
Jul 2021
past
C1-B.1.1.28
C2-B.1.195|other
C3-P.1
C4-B.1.1.33|N9
C5-B.1.1.28
C6-B.1.1|B.1.1.28|other
C7-P.4
C8-P.2
C9-P.1+P.4
C10-P.1+P.4
C11-B.1.1.7
C12-P.1.2
C13-P.4
C14-P.1
C15-P.4

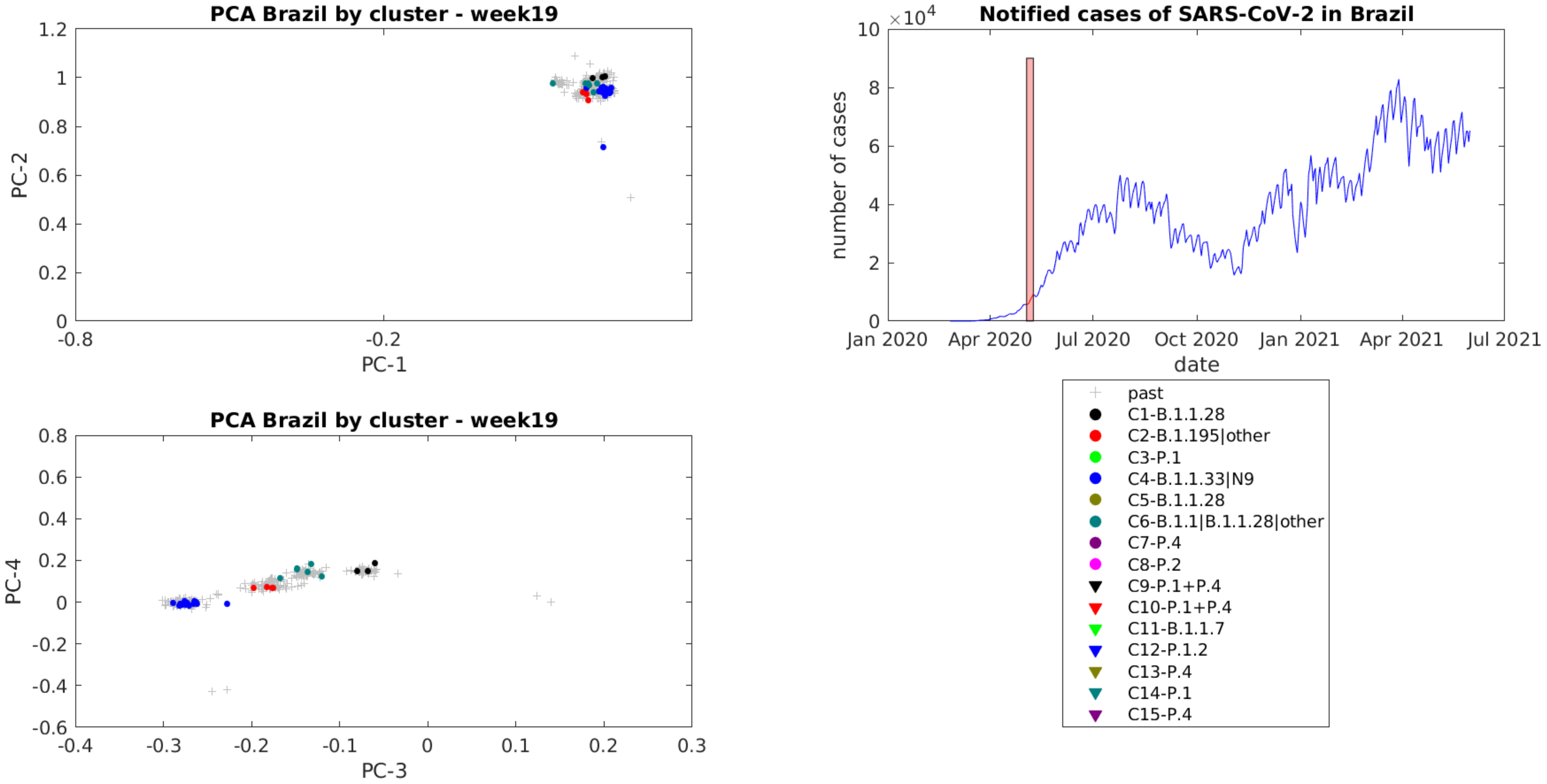
PCA Brazil by cluster - week19
PC-1
PC-2
PCA Brazil by cluster - week19
PC-3
PC-4
Notified cases of SARS-CoV-2 in Brazil
×10^4
number of cases
date
Jan 2020
Apr 2020
Jul 2020
Oct 2020
Jan 2021
Apr 2021
Jul 2021
past
C1-B.1.1.28
C2-B.1.195|other
C3-P.1
C4-B.1.1.33|N9
C5-B.1.1.28
C6-B.1.1|B.1.1.28|other
C7-P.4
C8-P.2
C9-P.1+P.4
C10-P.1+P.4
C11-B.1.1.7
C12-P.1.2
C13-P.4
C14-P.1
C15-P.4

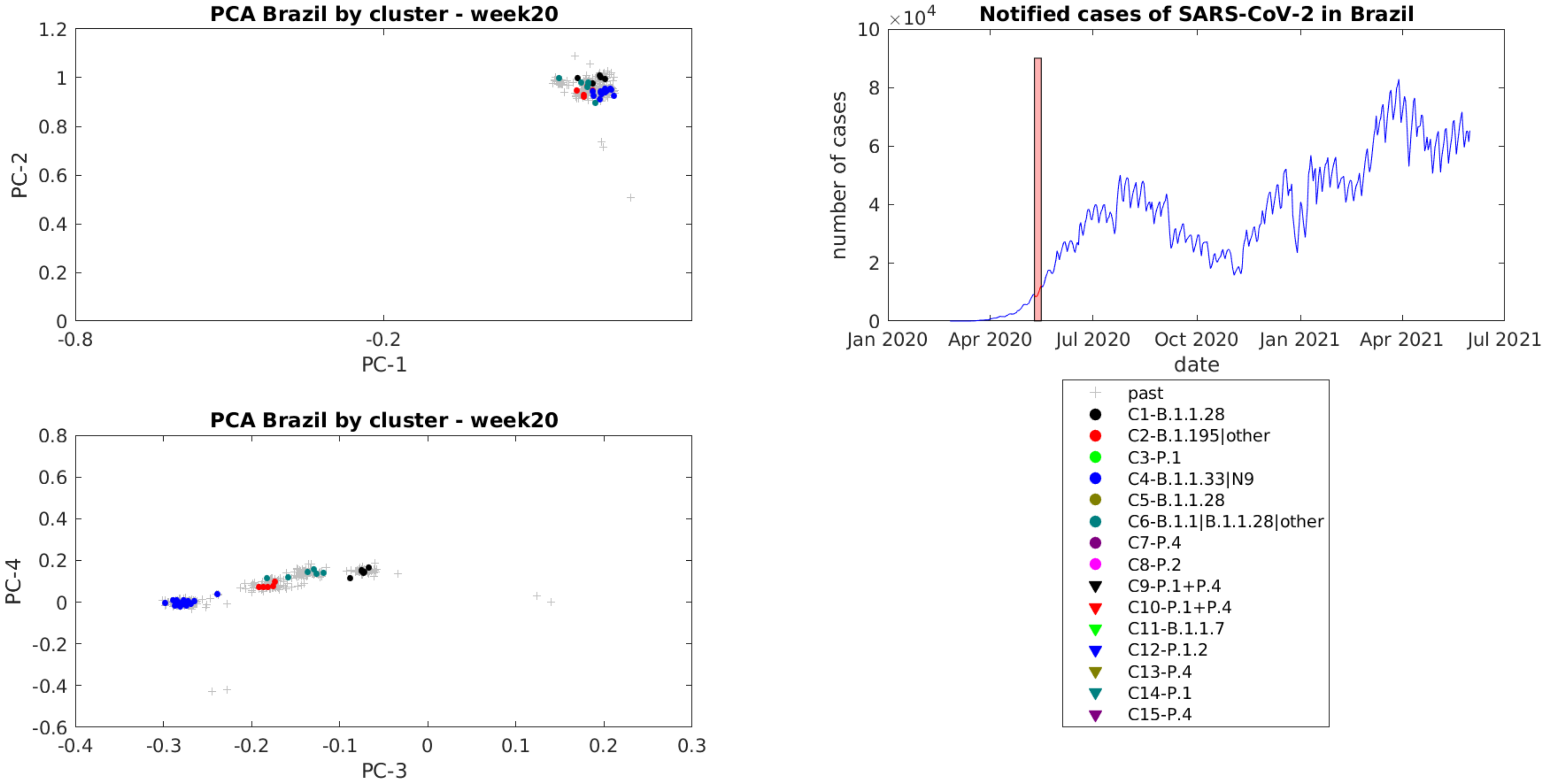

PCA Brazil by cluster - week20
PC-2
PC-1
PCA Brazil by cluster - week20
PC-4
PC-3
Notified cases of SARS-CoV-2 in Brazil
×10^4
number of cases
date
Jan 2020
Apr 2020
Jul 2020
Oct 2020
Jan 2021
Apr 2021
Jul 2021
past
C1-B.1.1.28
C2-B.1.195|other
C3-P.1
C4-B.1.1.33|N9
C5-B.1.1.28
C6-B.1.1|B.1.1.28|other
C7-P.4
C8-P.2
C9-P.1+P.4
C10-P.1+P.4
C11-B.1.1.7
C12-P.1.2
C13-P.4
C14-P.1
C15-P.4

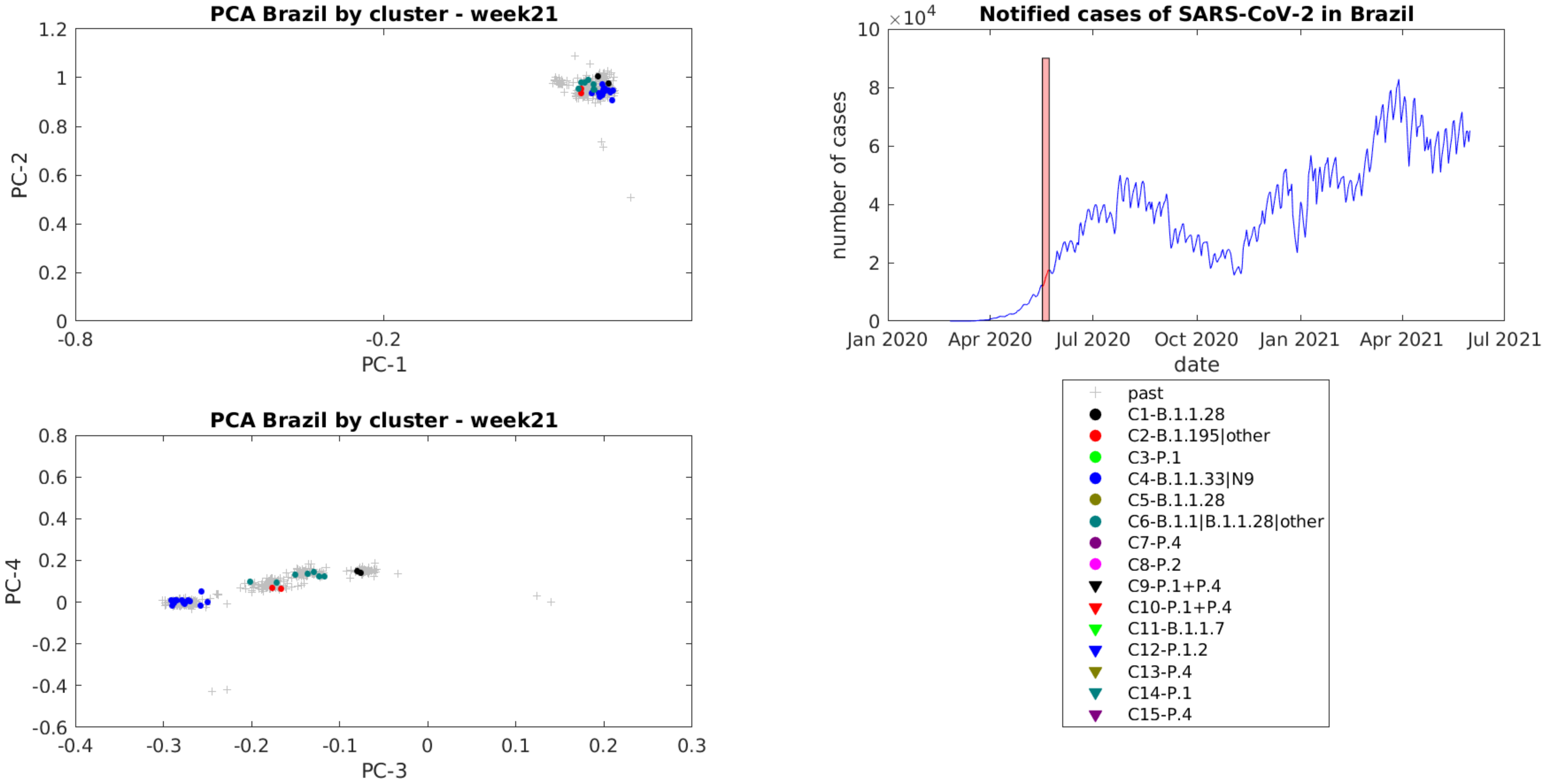
PCA Brazil by cluster - week21
PC-1
PC-2
PCA Brazil by cluster - week21
PC-3
PC-4
Notified cases of SARS-CoV-2 in Brazil
×10^4
number of cases
date
Jan 2020
Apr 2020
Jul 2020
Oct 2020
Jan 2021
Apr 2021
Jul 2021
past
C1-B.1.1.28
C2-B.1.195|other
C3-P.1
C4-B.1.1.33|N9
C5-B.1.1.28
C6-B.1.1|B.1.1.28|other
C7-P.4
C8-P.2
C9-P.1+P.4
C10-P.1+P.4
C11-B.1.1.7
C12-P.1.2
C13-P.4
C14-P.1
C15-P.4

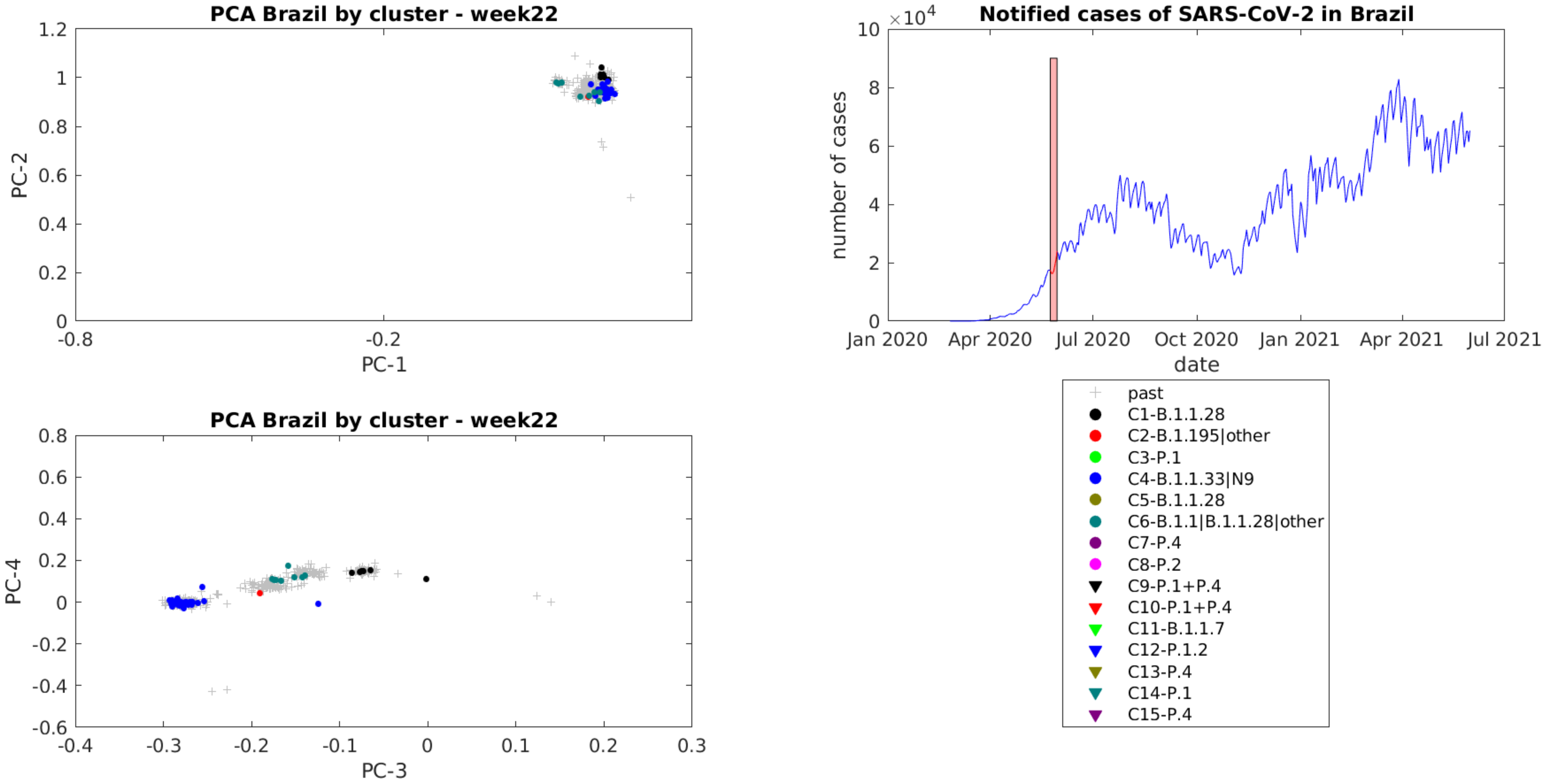
PCA Brazil by cluster - week22
PC-1
PC-2
PCA Brazil by cluster - week22
PC-3
PC-4
Notified cases of SARS-CoV-2 in Brazil
number of cases
date
Jan 2020
Apr 2020
Jul 2020
Oct 2020
Jan 2021
Apr 2021
Jul 2021
past
C1-B.1.1.28
C2-B.1.195|other
C3-P.1
C4-B.1.1.33|N9
C5-B.1.1.28
C6-B.1.1|B.1.1.28|other
C7-P.4
C8-P.2
C9-P.1+P.4
C10-P.1+P.4
C11-B.1.1.7
C12-P.1.2
C13-P.4
C14-P.1
C15-P.4

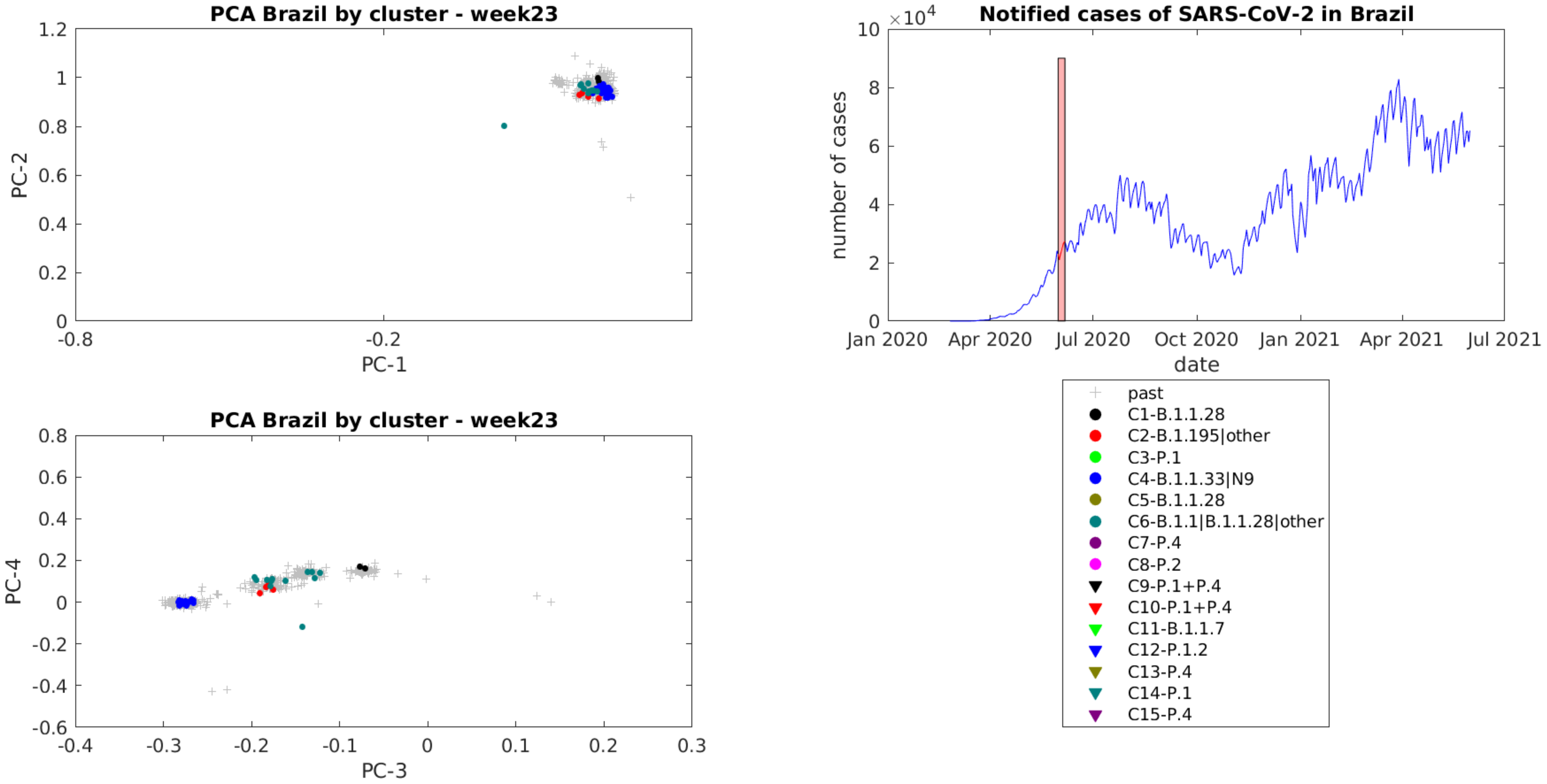
PCA Brazil by cluster - week23
PC-1
PC-2
PCA Brazil by cluster - week23
PC-3
PC-4
Notified cases of SARS-CoV-2 in Brazil
×10^4
number of cases
date
Jan 2020
Apr 2020
Jul 2020
Oct 2020
Jan 2021
Apr 2021
Jul 2021
past
C1-B.1.1.28
C2-B.1.195|other
C3-P.1
C4-B.1.1.33|N9
C5-B.1.1.28
C6-B.1.1|B.1.1.28|other
C7-P.4
C8-P.2
C9-P.1+P.4
C10-P.1+P.4
C11-B.1.1.7
C12-P.1.2
C13-P.4
C14-P.1
C15-P.4

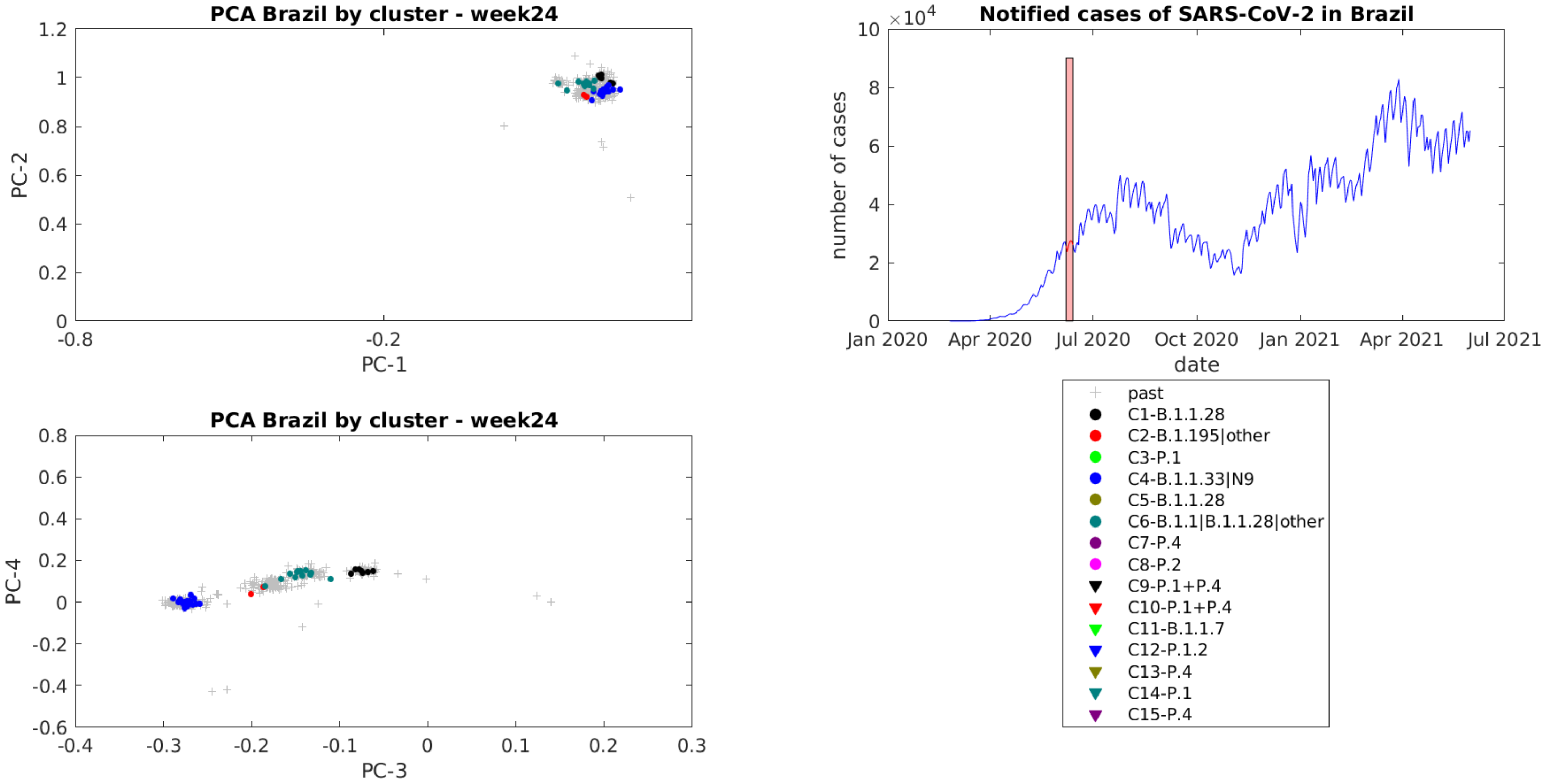
PCA Brazil by cluster - week24
PC-1
PC-2
Notified cases of SARS-CoV-2 in Brazil
×10^4
number of cases
date
Jan 2020
Apr 2020
Jul 2020
Oct 2020
Jan 2021
Apr 2021
Jul 2021
past
C1-B.1.1.28
C2-B.1.195|other
C3-P.1
C4-B.1.1.33|N9
C5-B.1.1.28
C6-B.1.1|B.1.1.28|other
C7-P.4
C8-P.2
C9-P.1+P.4
C10-P.1+P.4
C11-B.1.1.7
C12-P.1.2
C13-P.4
C14-P.1
C15-P.4
PCA Brazil by cluster - week24
PC-3
PC-4

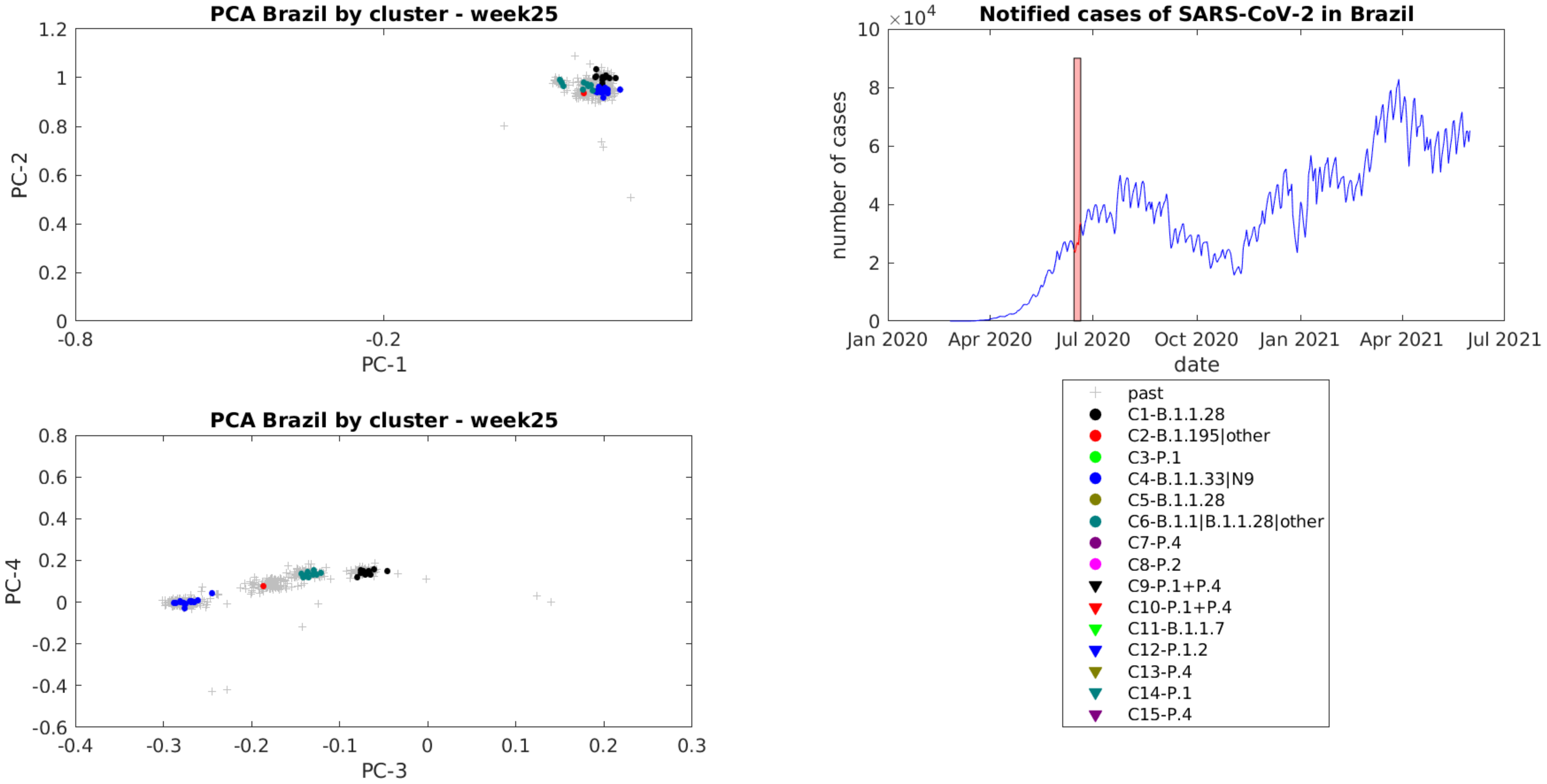

PCA Brazil by cluster - week25
PC-1
PC-2
PCA Brazil by cluster - week25
PC-3
PC-4
Notified cases of SARS-CoV-2 in Brazil
×10^4
number of cases
date
Jan 2020
Apr 2020
Jul 2020
Oct 2020
Jan 2021
Apr 2021
Jul 2021
past
C1-B.1.1.28
C2-B.1.195|other
C3-P.1
C4-B.1.1.33|N9
C5-B.1.1.28
C6-B.1.1|B.1.1.28|other
C7-P.4
C8-P.2
C9-P.1+P.4
C10-P.1+P.4
C11-B.1.1.7
C12-P.1.2
C13-P.4
C14-P.1
C15-P.4

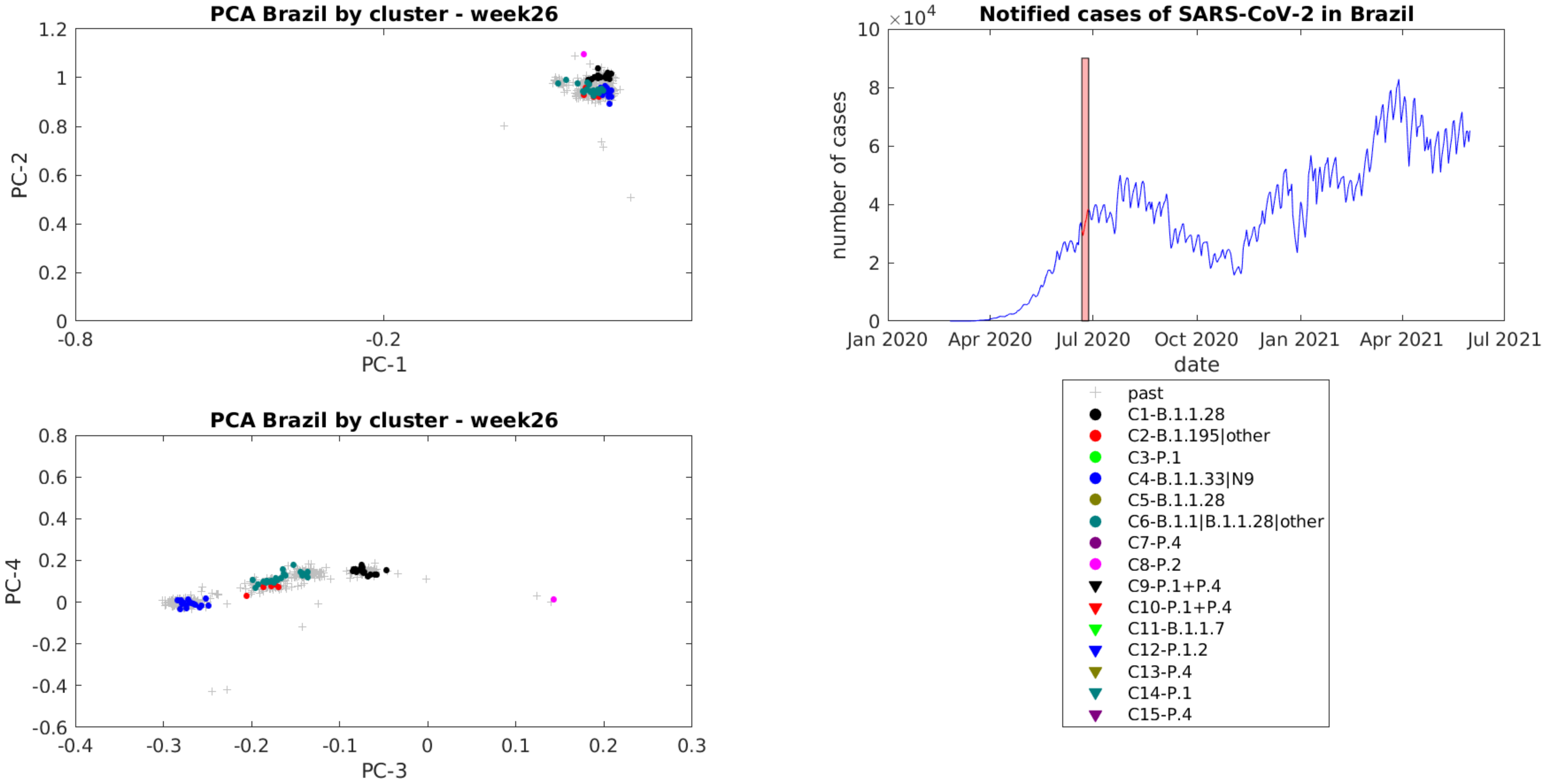
PCA Brazil by cluster - week26
PC-2
PC-1
1.2
1
0.8
0.6
0.4
0.2
0
-0.8
-0.2
PCA Brazil by cluster - week26
PC-4
PC-3
0.8
0.6
0.4
0.2
0
-0.2
-0.4
-0.6
-0.4
-0.3
-0.2
-0.1
0
0.1
0.2
0.3
Notified cases of SARS-CoV-2 in Brazil
×10^4
10
8
6
4
2
0
number of cases
date
Jan 2020
Apr 2020
Jul 2020
Oct 2020
Jan 2021
Apr 2021
Jul 2021
past
C1-B.1.1.28
C2-B.1.195|other
C3-P.1
C4-B.1.1.33|N9
C5-B.1.1.28
C6-B.1.1|B.1.1.28|other
C7-P.4
C8-P.2
C9-P.1+P.4
C10-P.1+P.4
C11-B.1.1.7
C12-P.1.2
C13-P.4
C14-P.1
C15-P.4

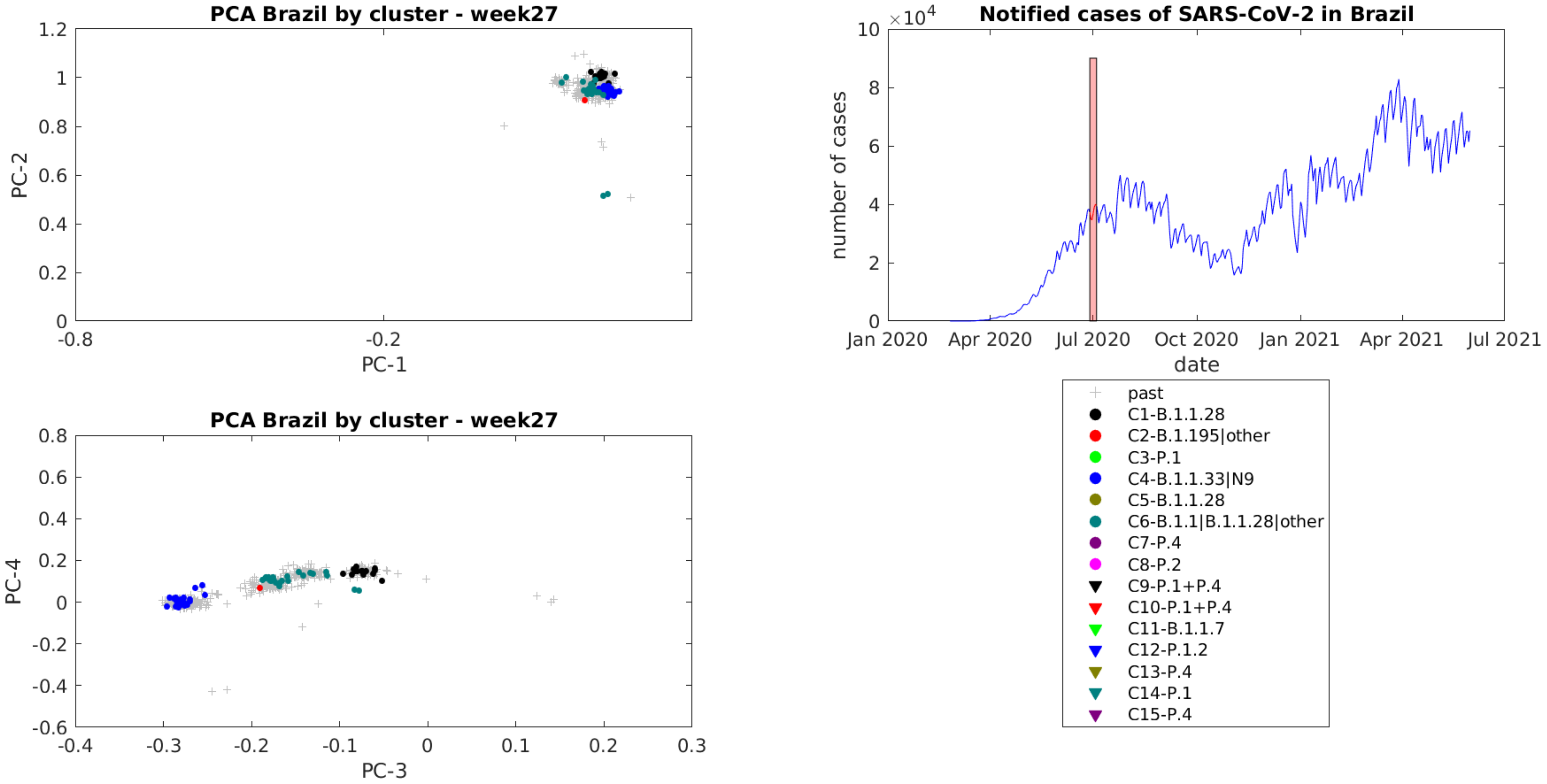
PCA Brazil by cluster - week27
PC-1
PC-2
1.2
1
0.8
0.6
0.4
0.2
0
-0.8
-0.2
PCA Brazil by cluster - week27
PC-3
PC-4
0.8
0.6
0.4
0.2
0
-0.2
-0.4
-0.6
-0.4
-0.3
-0.2
-0.1
0
0.1
0.2
0.3
Notified cases of SARS-CoV-2 in Brazil
$\times 10^4$
number of cases
10
8
6
4
2
0
Jan 2020
Apr 2020
Jul 2020
Oct 2020
Jan 2021
Apr 2021
Jul 2021
date
past
C1-B.1.1.28
C2-B.1.195|other
C3-P.1
C4-B.1.1.33|N9
C5-B.1.1.28
C6-B.1.1|B.1.1.28|other
C7-P.4
C8-P.2
C9-P.1+P.4
C10-P.1+P.4
C11-B.1.1.7
C12-P.1.2
C13-P.4
C14-P.1
C15-P.4

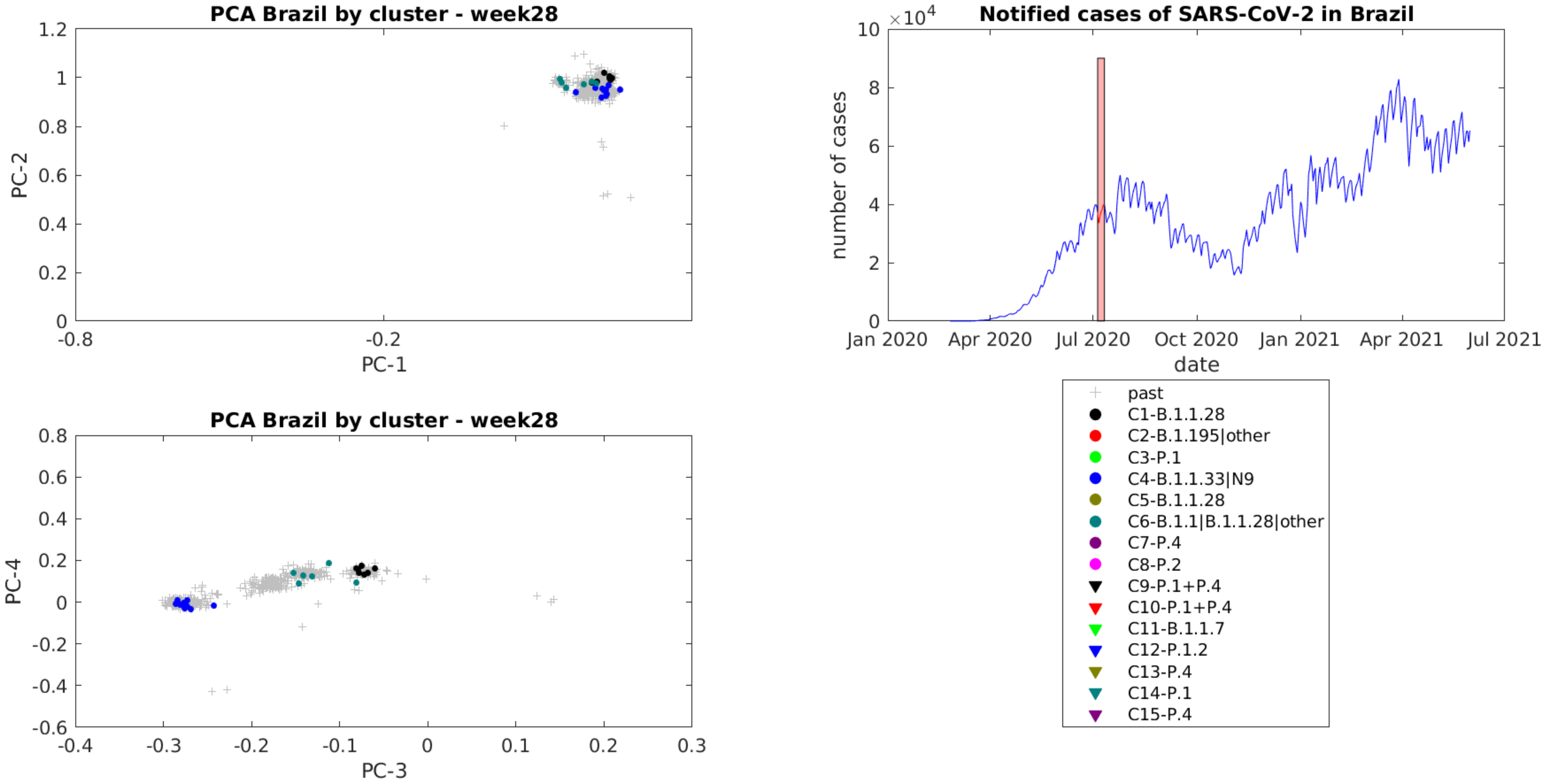

PCA Brazil by cluster - week28
PC-1
PC-2
PCA Brazil by cluster - week28
PC-3
PC-4
Notified cases of SARS-CoV-2 in Brazil
number of cases
date
Jan 2020
Apr 2020
Jul 2020
Oct 2020
Jan 2021
Apr 2021
Jul 2021
past
C1-B.1.1.28
C2-B.1.195|other
C3-P.1
C4-B.1.1.33|N9
C5-B.1.1.28
C6-B.1.1|B.1.1.28|other
C7-P.4
C8-P.2
C9-P.1+P.4
C10-P.1+P.4
C11-B.1.1.7
C12-P.1.2
C13-P.4
C14-P.1
C15-P.4

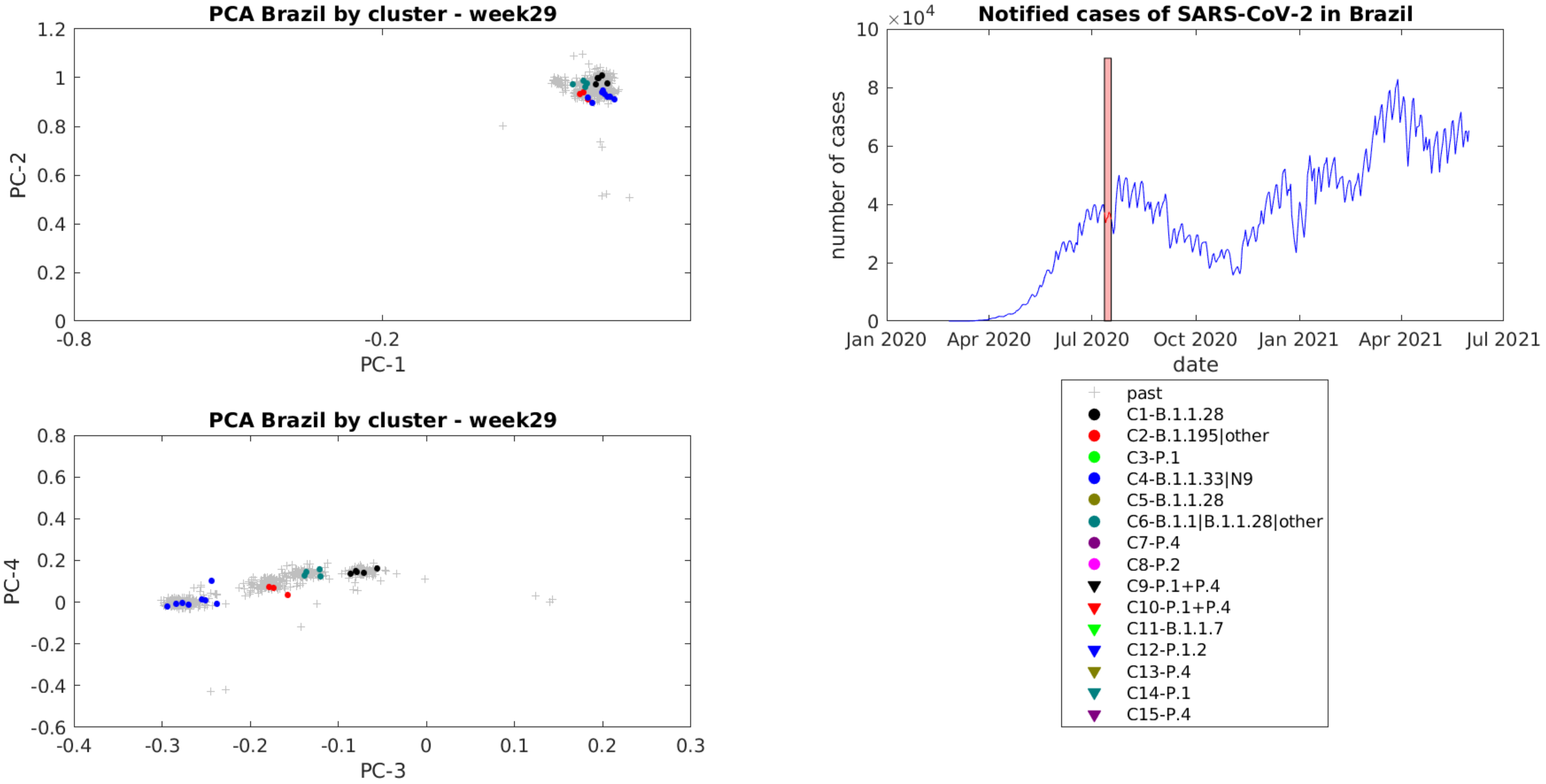
PCA Brazil by cluster - week29
PC-1
PC-2
PCA Brazil by cluster - week29
PC-3
PC-4
Notified cases of SARS-CoV-2 in Brazil
$\times 10^4$
number of cases
date
Jan 2020
Apr 2020
Jul 2020
Oct 2020
Jan 2021
Apr 2021
Jul 2021
past
C1-B.1.1.28
C2-B.1.195|other
C3-P.1
C4-B.1.1.33|N9
C5-B.1.1.28
C6-B.1.1|B.1.1.28|other
C7-P.4
C8-P.2
C9-P.1+P.4
C10-P.1+P.4
C11-B.1.1.7
C12-P.1.2
C13-P.4
C14-P.1
C15-P.4

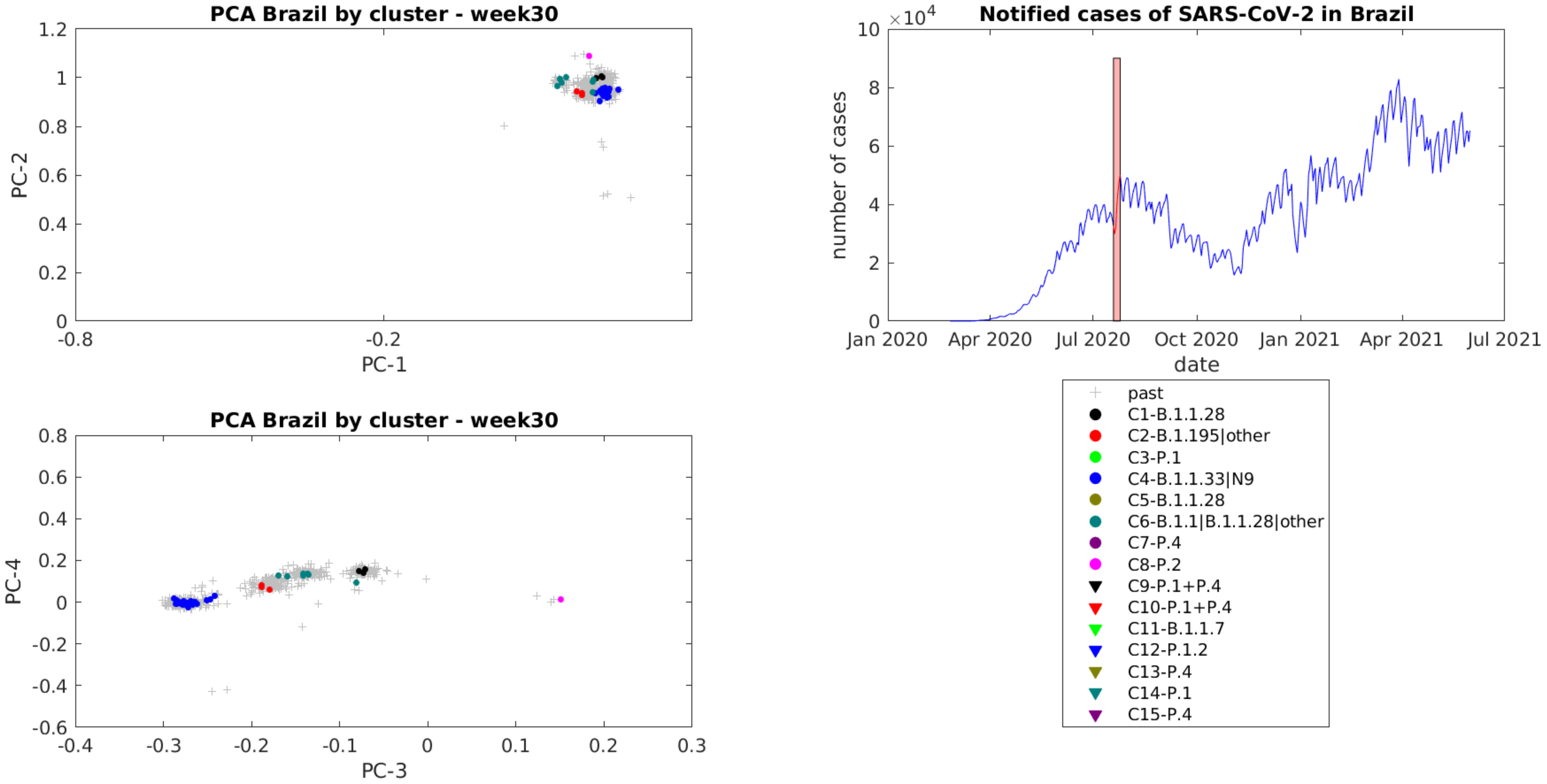

PCA Brazil by cluster - week30
PC-1
PC-2
PC-3
PC-4
Notified cases of SARS-CoV-2 in Brazil
number of cases
date
Jan 2020
Apr 2020
Jul 2020
Oct 2020
Jan 2021
Apr 2021
Jul 2021
past
C1-B.1.1.28
C2-B.1.195|other
C3-P.1
C4-B.1.1.33|N9
C5-B.1.1.28
C6-B.1.1|B.1.1.28|other
C7-P.4
C8-P.2
C9-P.1+P.4
C10-P.1+P.4
C11-B.1.1.7
C12-P.1.2
C13-P.4
C14-P.1
C15-P.4

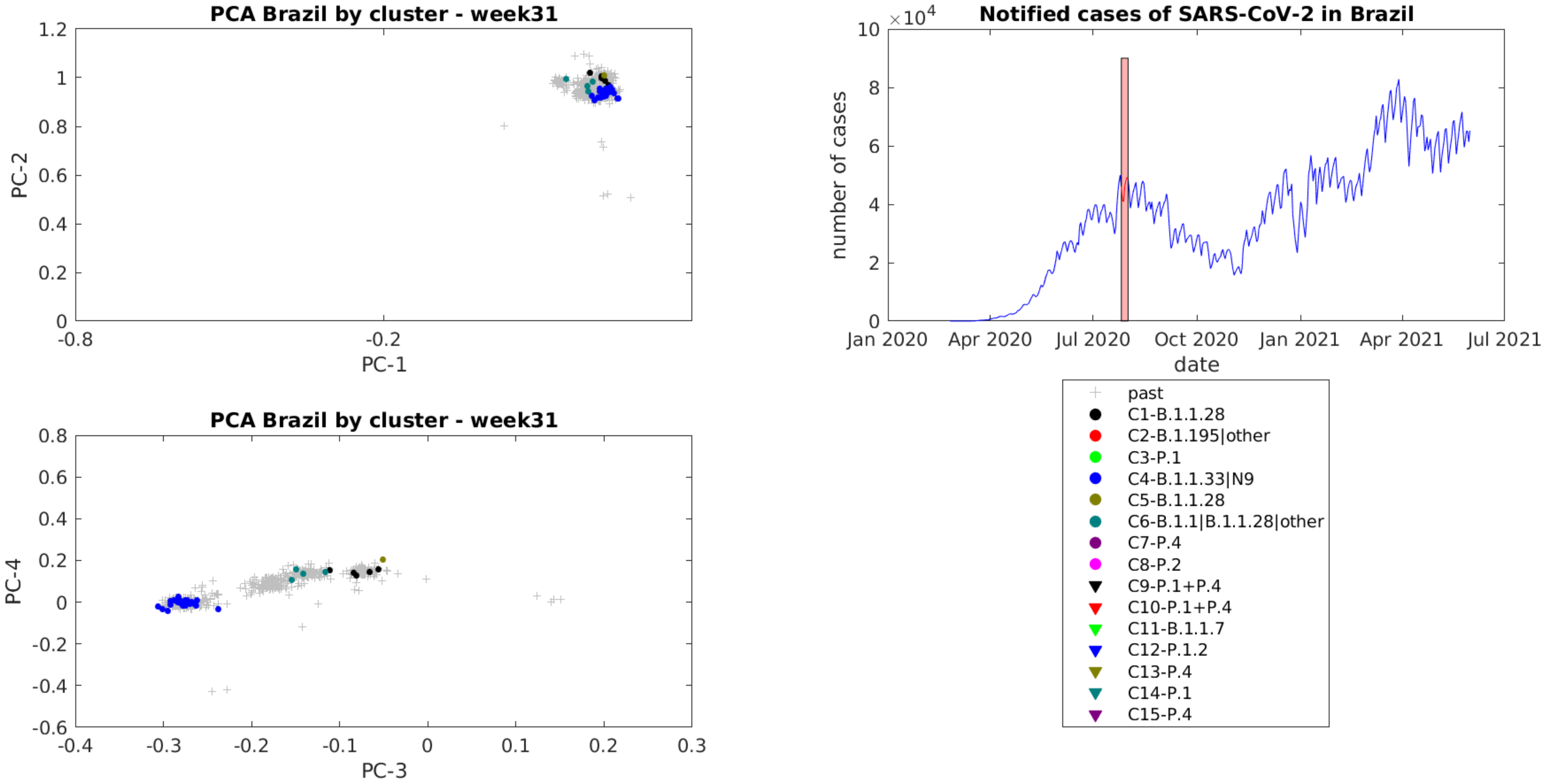

PCA Brazil by cluster - week31
PC-1
PC-2
PCA Brazil by cluster - week31
PC-3
PC-4
Notified cases of SARS-CoV-2 in Brazil
×10^4
number of cases
date
Jan 2020
Apr 2020
Jul 2020
Oct 2020
Jan 2021
Apr 2021
Jul 2021
past
C1-B.1.1.28
C2-B.1.195|other
C3-P.1
C4-B.1.1.33|N9
C5-B.1.1.28
C6-B.1.1|B.1.1.28|other
C7-P.4
C8-P.2
C9-P.1+P.4
C10-P.1+P.4
C11-B.1.1.7
C12-P.1.2
C13-P.4
C14-P.1
C15-P.4

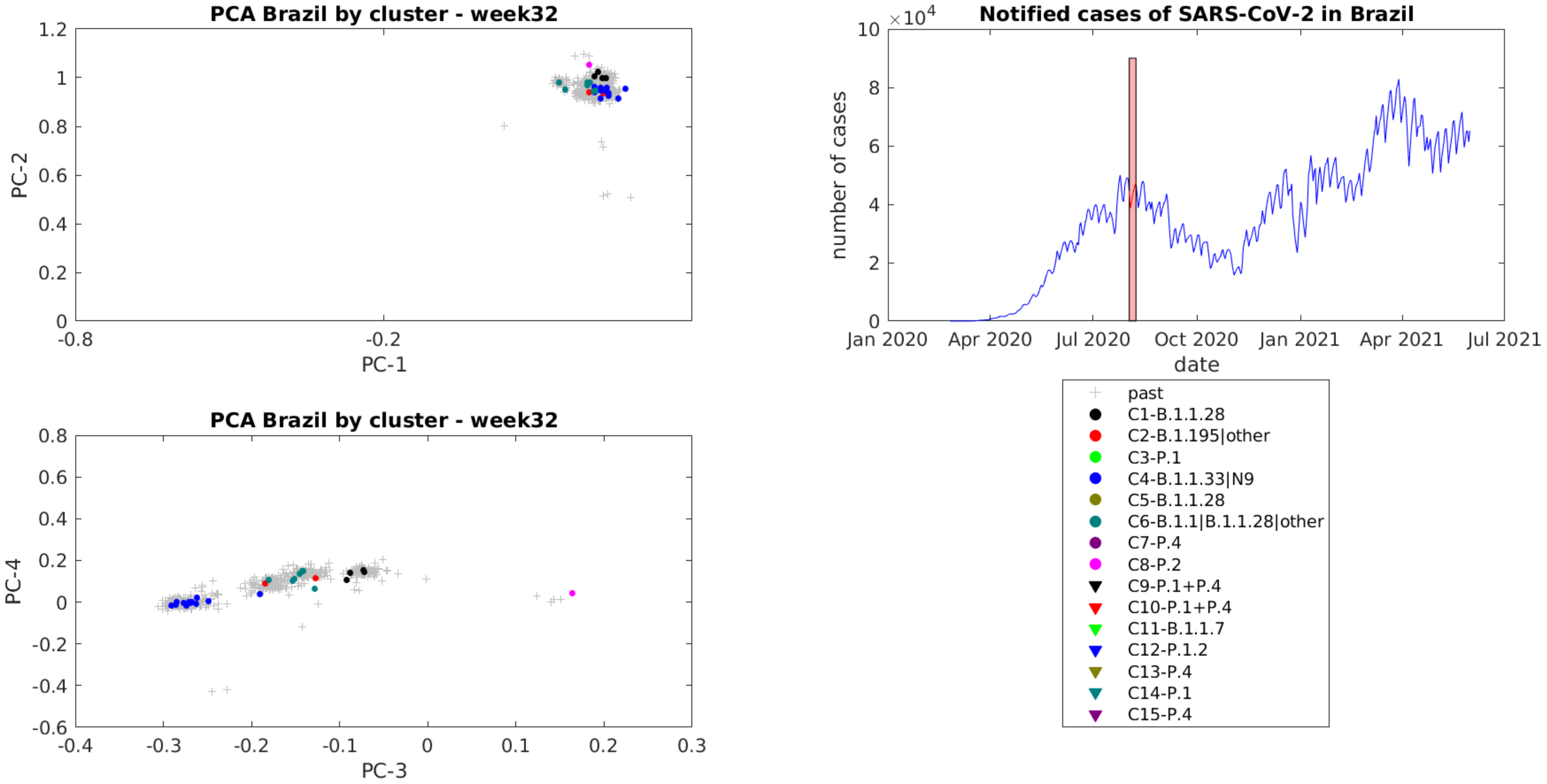
PCA Brazil by cluster - week32
PC-1
PC-2
PCA Brazil by cluster - week32
PC-3
PC-4
Notified cases of SARS-CoV-2 in Brazil
$\times 10^4$
number of cases
date
Jan 2020
Apr 2020
Jul 2020
Oct 2020
Jan 2021
Apr 2021
Jul 2021
past
C1-B.1.1.28
C2-B.1.195|other
C3-P.1
C4-B.1.1.33|N9
C5-B.1.1.28
C6-B.1.1|B.1.1.28|other
C7-P.4
C8-P.2
C9-P.1+P.4
C10-P.1+P.4
C11-B.1.1.7
C12-P.1.2
C13-P.4
C14-P.1
C15-P.4

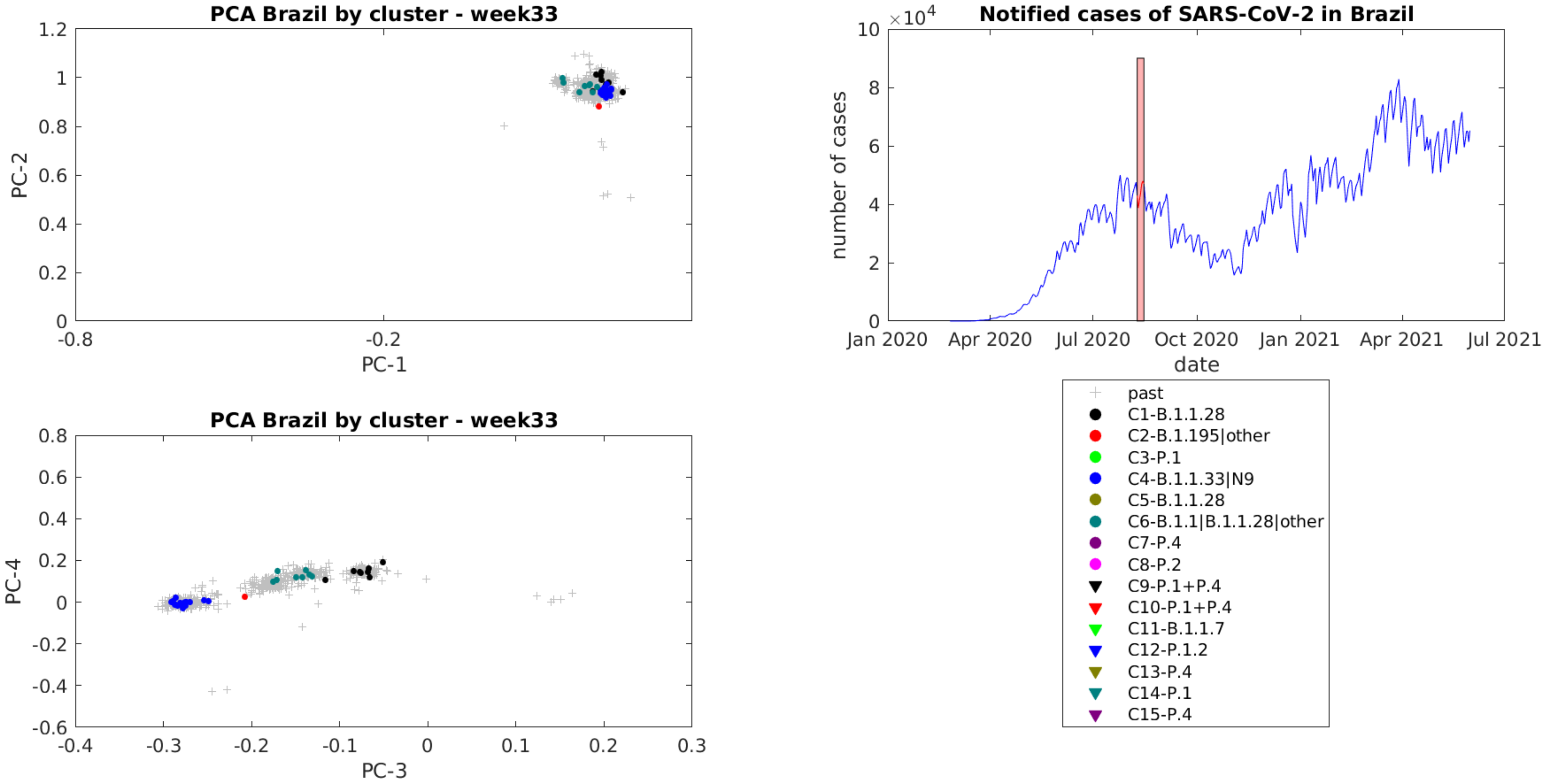

PCA Brazil by cluster - week33
PC-2
PC-1
PCA Brazil by cluster - week33
PC-4
PC-3
Notified cases of SARS-CoV-2 in Brazil
number of cases
date
Jan 2020
Apr 2020
Jul 2020
Oct 2020
Jan 2021
Apr 2021
Jul 2021
past
C1-B.1.1.28
C2-B.1.195|other
C3-P.1
C4-B.1.1.33|N9
C5-B.1.1.28
C6-B.1.1|B.1.1.28|other
C7-P.4
C8-P.2
C9-P.1+P.4
C10-P.1+P.4
C11-B.1.1.7
C12-P.1.2
C13-P.4
C14-P.1
C15-P.4

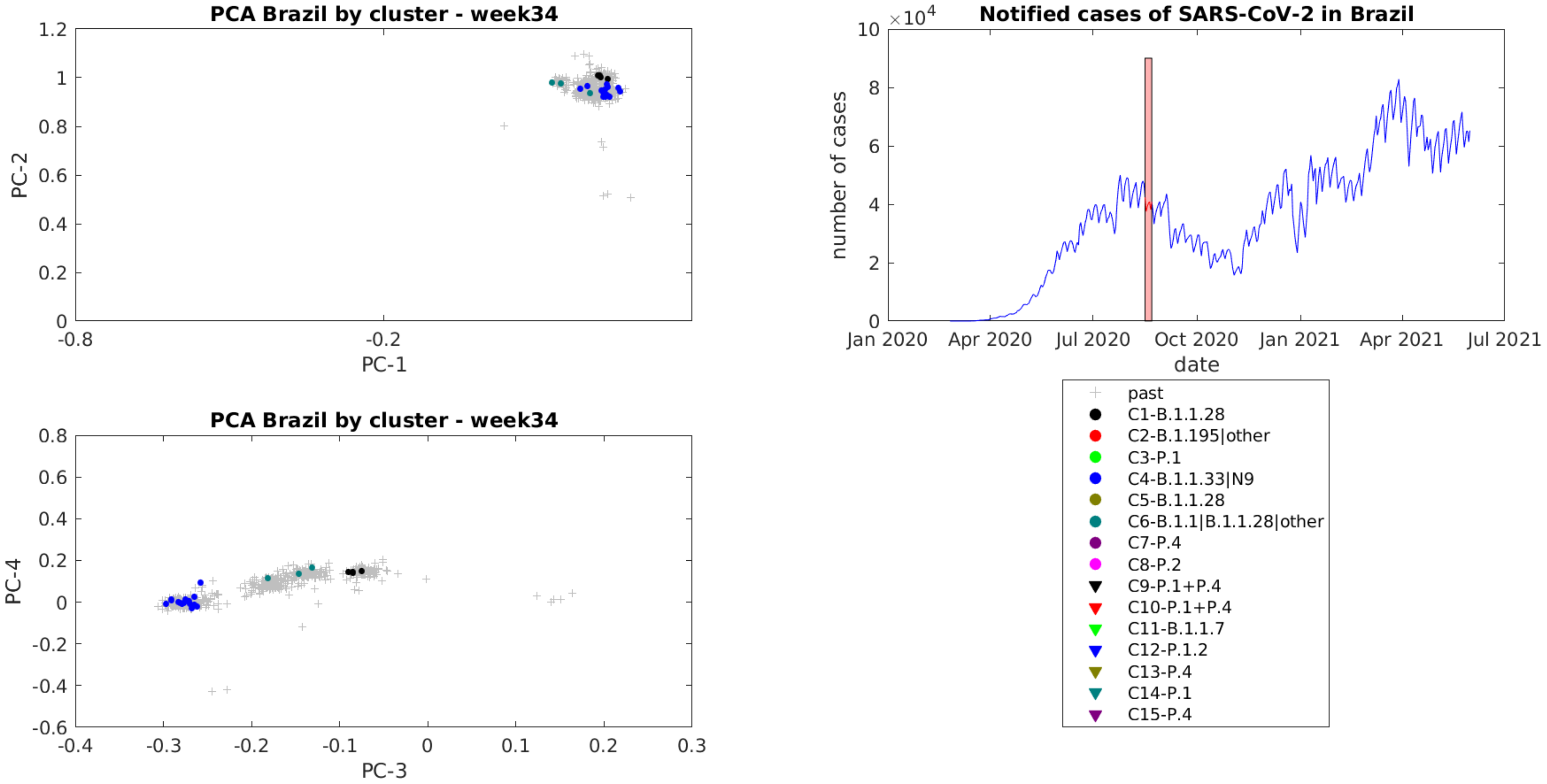
PCA Brazil by cluster - week34
1.2
1
0.8
0.6
0.4
0.2
0
PC-2
-0.8
-0.2
PC-1
Notified cases of SARS-CoV-2 in Brazil
×10^4
10
8
6
4
2
0
number of cases
Jan 2020
Apr 2020
Jul 2020
Oct 2020
Jan 2021
Apr 2021
Jul 2021
date
PCA Brazil by cluster - week34
0.8
0.6
0.4
0.2
0
-0.2
-0.4
-0.6
PC-4
-0.4
-0.3
-0.2
-0.1
0
0.1
0.2
0.3
PC-3
past
C1-B.1.1.28
C2-B.1.195|other
C3-P.1
C4-B.1.1.33|N9
C5-B.1.1.28
C6-B.1.1|B.1.1.28|other
C7-P.4
C8-P.2
C9-P.1+P.4
C10-P.1+P.4
C11-B.1.1.7
C12-P.1.2
C13-P.4
C14-P.1
C15-P.4

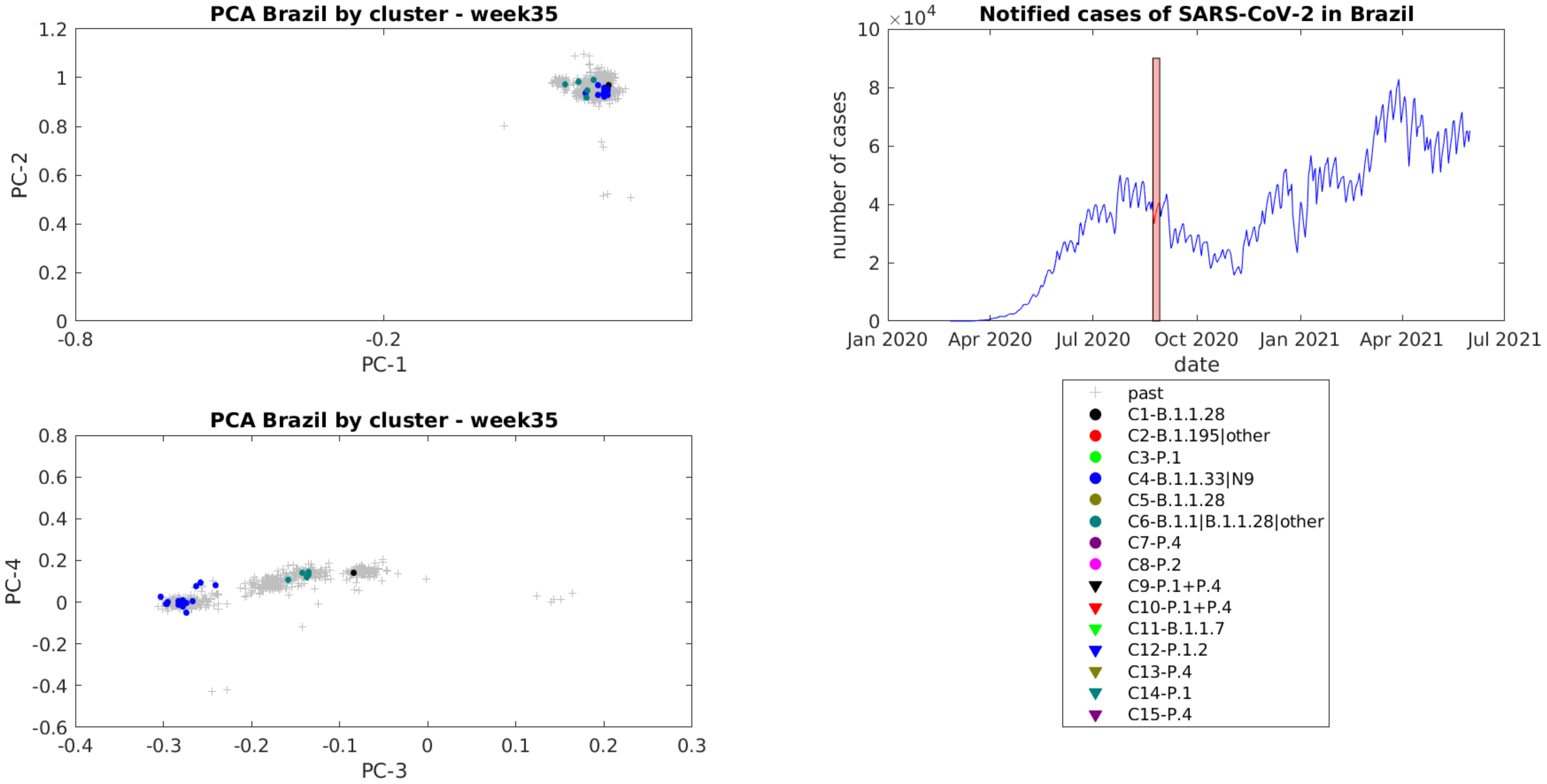

PCA Brazil by cluster - week35
PC-1
PC-2
Notified cases of SARS-CoV-2 in Brazil
×10^4
number of cases
date
Jan 2020
Apr 2020
Jul 2020
Oct 2020
Jan 2021
Apr 2021
Jul 2021
PCA Brazil by cluster - week35
PC-3
PC-4
past
C1-B.1.1.28
C2-B.1.195|other
C3-P.1
C4-B.1.1.33|N9
C5-B.1.1.28
C6-B.1.1|B.1.1.28|other
C7-P.4
C8-P.2
C9-P.1+P.4
C10-P.1+P.4
C11-B.1.1.7
C12-P.1.2
C13-P.4
C14-P.1
C15-P.4

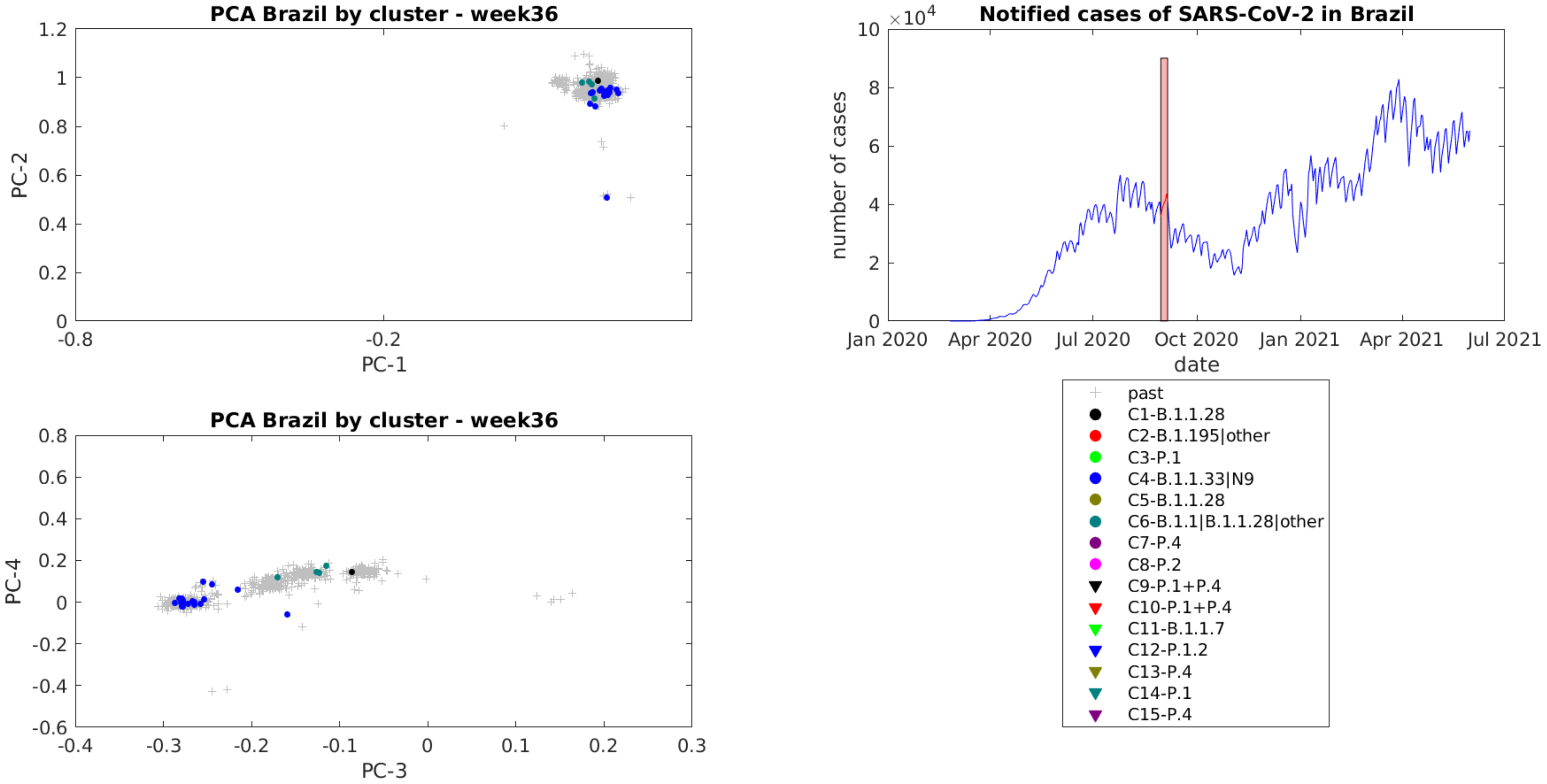

PCA Brazil by cluster - week36
PC-2
PC-1
PCA Brazil by cluster - week36
PC-4
PC-3
Notified cases of SARS-CoV-2 in Brazil
×10^4
number of cases
date
Jan 2020
Apr 2020
Jul 2020
Oct 2020
Jan 2021
Apr 2021
Jul 2021
past
C1-B.1.1.28
C2-B.1.195|other
C3-P.1
C4-B.1.1.33|N9
C5-B.1.1.28
C6-B.1.1|B.1.1.28|other
C7-P.4
C8-P.2
C9-P.1+P.4
C10-P.1+P.4
C11-B.1.1.7
C12-P.1.2
C13-P.4
C14-P.1
C15-P.4

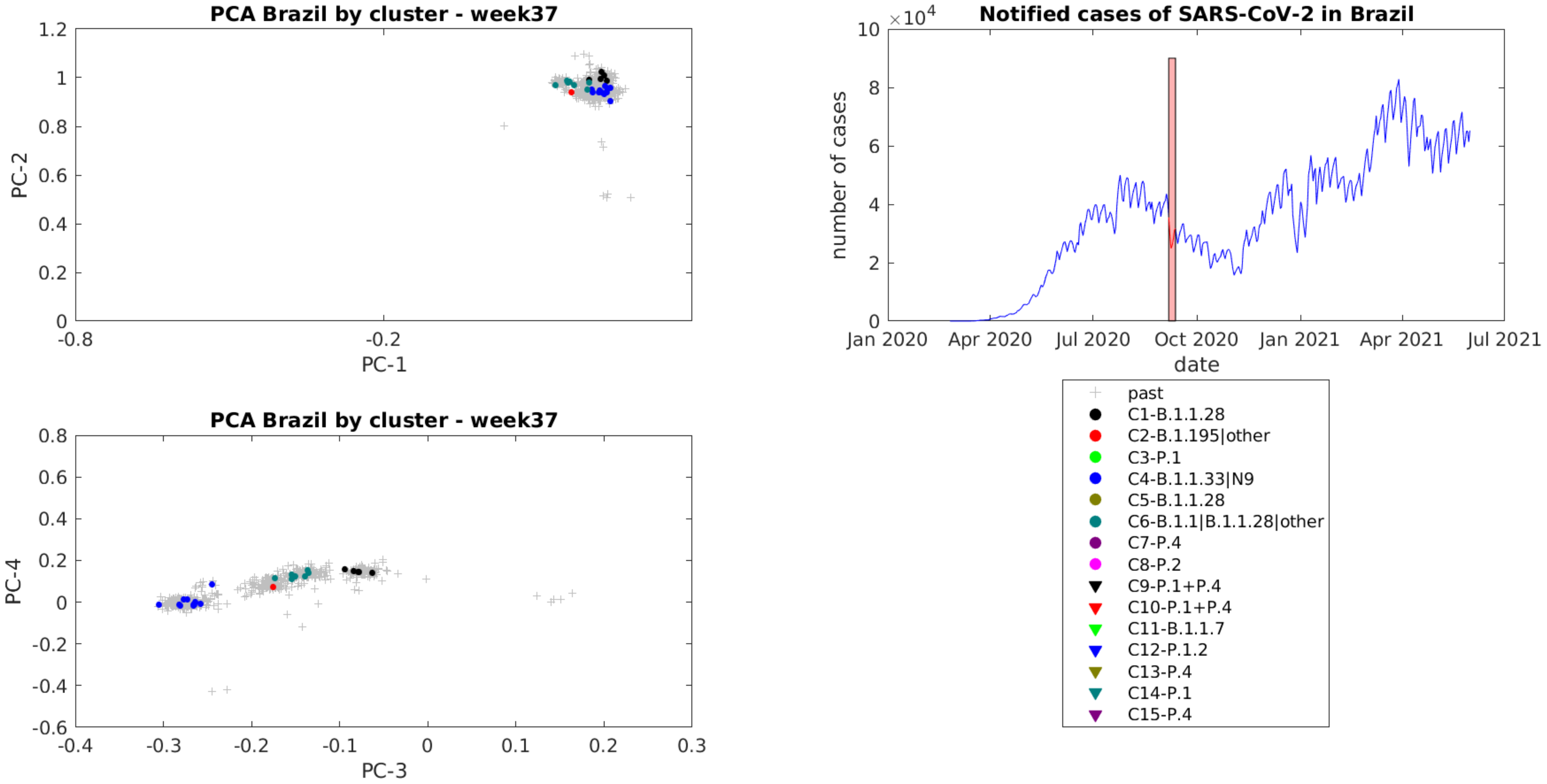
PCA Brazil by cluster - week37
PC-2
PC-1
1.2
1
0.8
0.6
0.4
0.2
0
-0.8
-0.2
Notified cases of SARS-CoV-2 in Brazil
×10^4
10
8
6
4
2
0
number of cases
Jan 2020
Apr 2020
Jul 2020
Oct 2020
Jan 2021
Apr 2021
Jul 2021
date
PCA Brazil by cluster - week37
PC-4
PC-3
0.8
0.6
0.4
0.2
0
-0.2
-0.4
-0.6
-0.4
-0.3
-0.2
-0.1
0
0.1
0.2
0.3
past
C1-B.1.1.28
C2-B.1.195|other
C3-P.1
C4-B.1.1.33|N9
C5-B.1.1.28
C6-B.1.1|B.1.1.28|other
C7-P.4
C8-P.2
C9-P.1+P.4
C10-P.1+P.4
C11-B.1.1.7
C12-P.1.2
C13-P.4
C14-P.1
C15-P.4

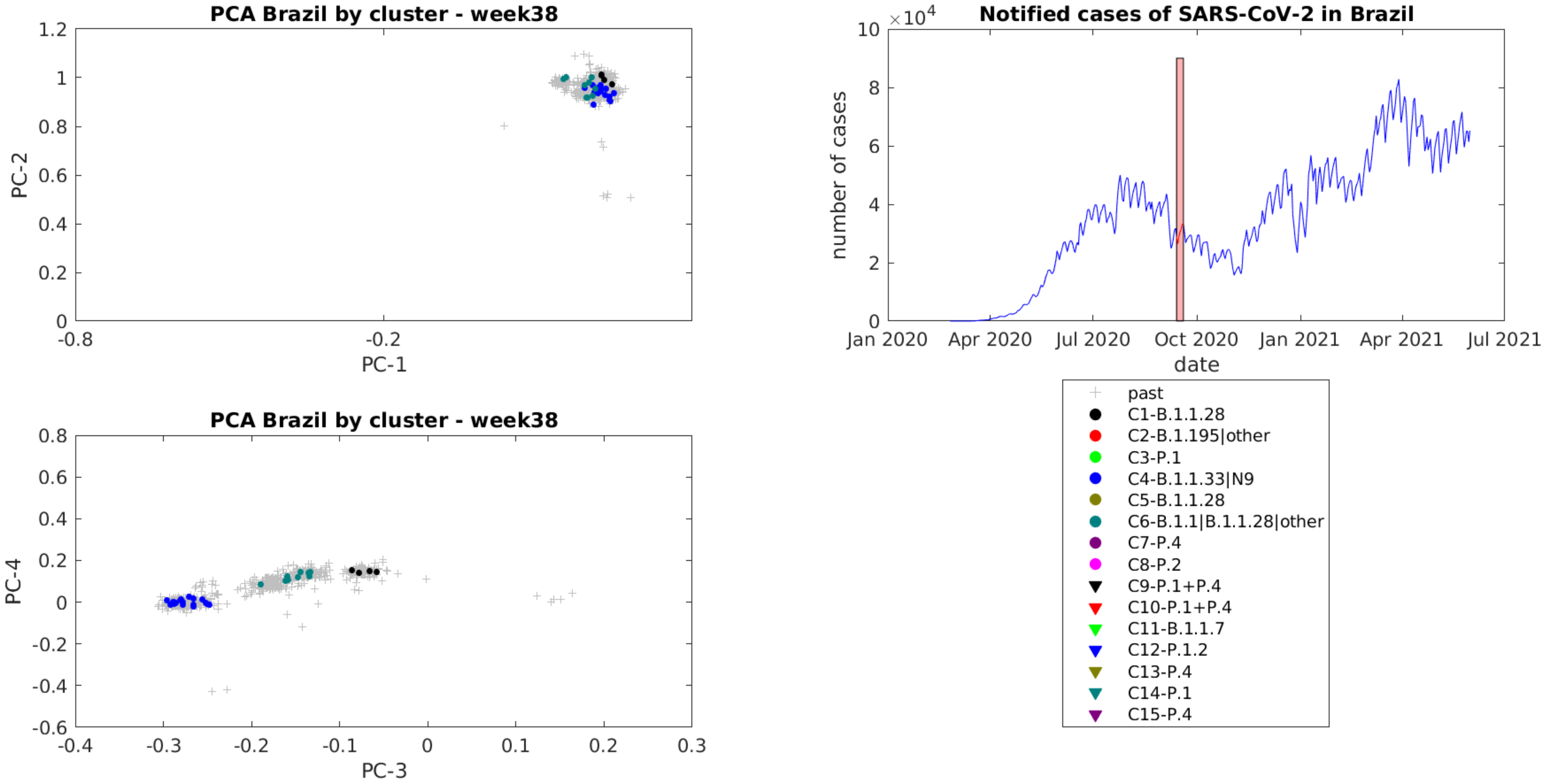
PCA Brazil by cluster - week38
PC-1
PC-2
PCA Brazil by cluster - week38
PC-3
PC-4
Notified cases of SARS-CoV-2 in Brazil
×10^4
number of cases
date
Jan 2020
Apr 2020
Jul 2020
Oct 2020
Jan 2021
Apr 2021
Jul 2021
past
C1-B.1.1.28
C2-B.1.195|other
C3-P.1
C4-B.1.1.33|N9
C5-B.1.1.28
C6-B.1.1|B.1.1.28|other
C7-P.4
C8-P.2
C9-P.1+P.4
C10-P.1+P.4
C11-B.1.1.7
C12-P.1.2
C13-P.4
C14-P.1
C15-P.4

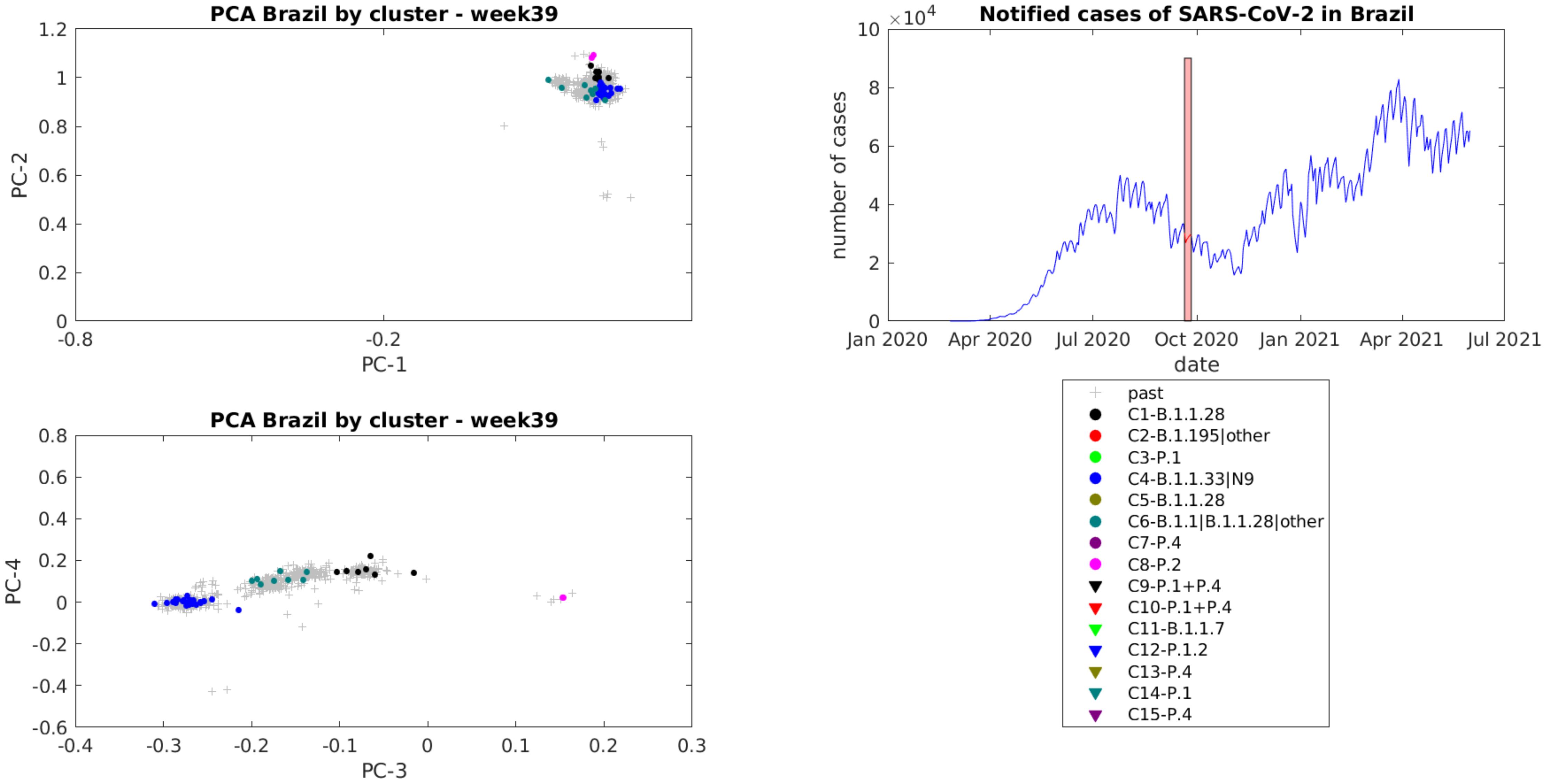
PCA Brazil by cluster - week39
PC-2
PC-1
1.2
1
0.8
0.6
0.4
0.2
0
-0.8
-0.2
PCA Brazil by cluster - week39
PC-4
PC-3
0.8
0.6
0.4
0.2
0
-0.2
-0.4
-0.6
-0.4
-0.3
-0.2
-0.1
0
0.1
0.2
0.3
Notified cases of SARS-CoV-2 in Brazil
×10^4
10
8
6
4
2
0
number of cases
date
Jan 2020
Apr 2020
Jul 2020
Oct 2020
Jan 2021
Apr 2021
Jul 2021
past
C1-B.1.1.28
C2-B.1.195|other
C3-P.1
C4-B.1.1.33|N9
C5-B.1.1.28
C6-B.1.1|B.1.1.28|other
C7-P.4
C8-P.2
C9-P.1+P.4
C10-P.1+P.4
C11-B.1.1.7
C12-P.1.2
C13-P.4
C14-P.1
C15-P.4

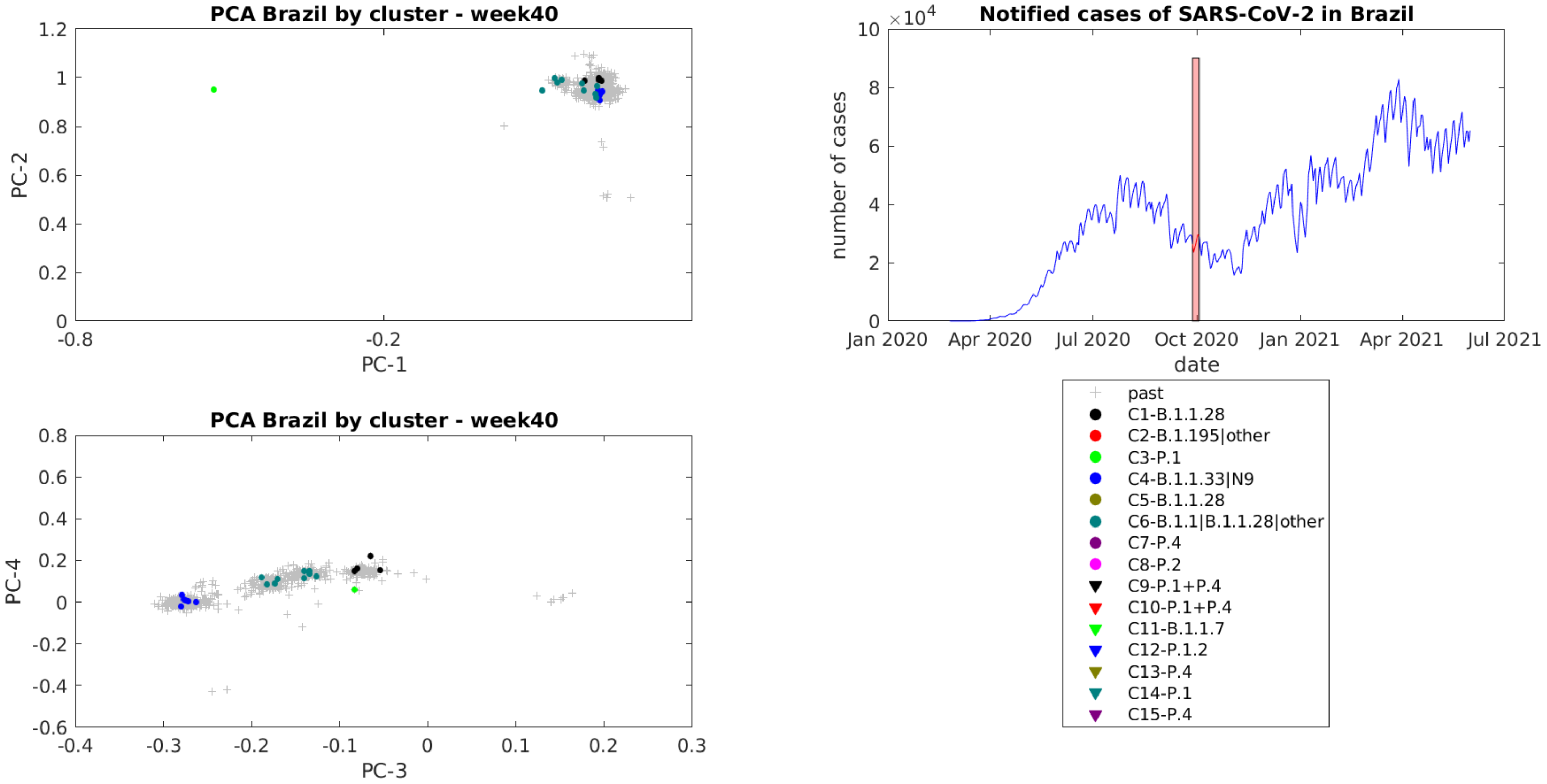
PCA Brazil by cluster - week40
PC-1
PC-2
PCA Brazil by cluster - week40
PC-3
PC-4
Notified cases of SARS-CoV-2 in Brazil
×10^4
number of cases
date
Jan 2020
Apr 2020
Jul 2020
Oct 2020
Jan 2021
Apr 2021
Jul 2021
past
C1-B.1.1.28
C2-B.1.195|other
C3-P.1
C4-B.1.1.33|N9
C5-B.1.1.28
C6-B.1.1|B.1.1.28|other
C7-P.4
C8-P.2
C9-P.1+P.4
C10-P.1+P.4
C11-B.1.1.7
C12-P.1.2
C13-P.4
C14-P.1
C15-P.4

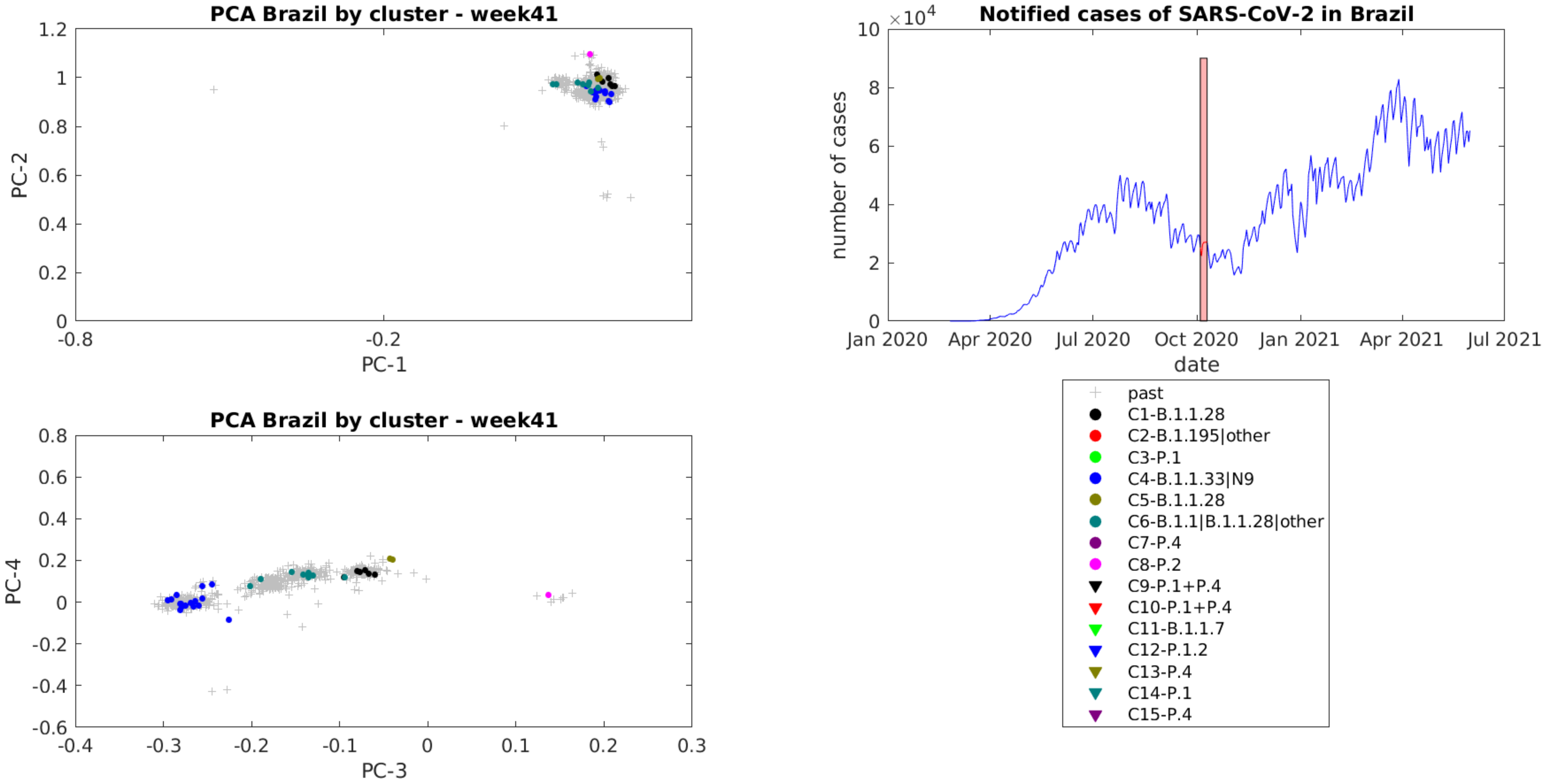
PCA Brazil by cluster - week41
PC-2
PC-1
1.2
1
0.8
0.6
0.4
0.2
0
-0.8
-0.2
Notified cases of SARS-CoV-2 in Brazil
×10^4
10
8
6
4
2
0
number of cases
date
Jan 2020
Apr 2020
Jul 2020
Oct 2020
Jan 2021
Apr 2021
Jul 2021
PCA Brazil by cluster - week41
PC-4
PC-3
0.8
0.6
0.4
0.2
0
-0.2
-0.4
-0.6
-0.4
-0.3
-0.2
-0.1
0
0.1
0.2
0.3
past
C1-B.1.1.28
C2-B.1.195|other
C3-P.1
C4-B.1.1.33|N9
C5-B.1.1.28
C6-B.1.1|B.1.1.28|other
C7-P.4
C8-P.2
C9-P.1+P.4
C10-P.1+P.4
C11-B.1.1.7
C12-P.1.2
C13-P.4
C14-P.1
C15-P.4

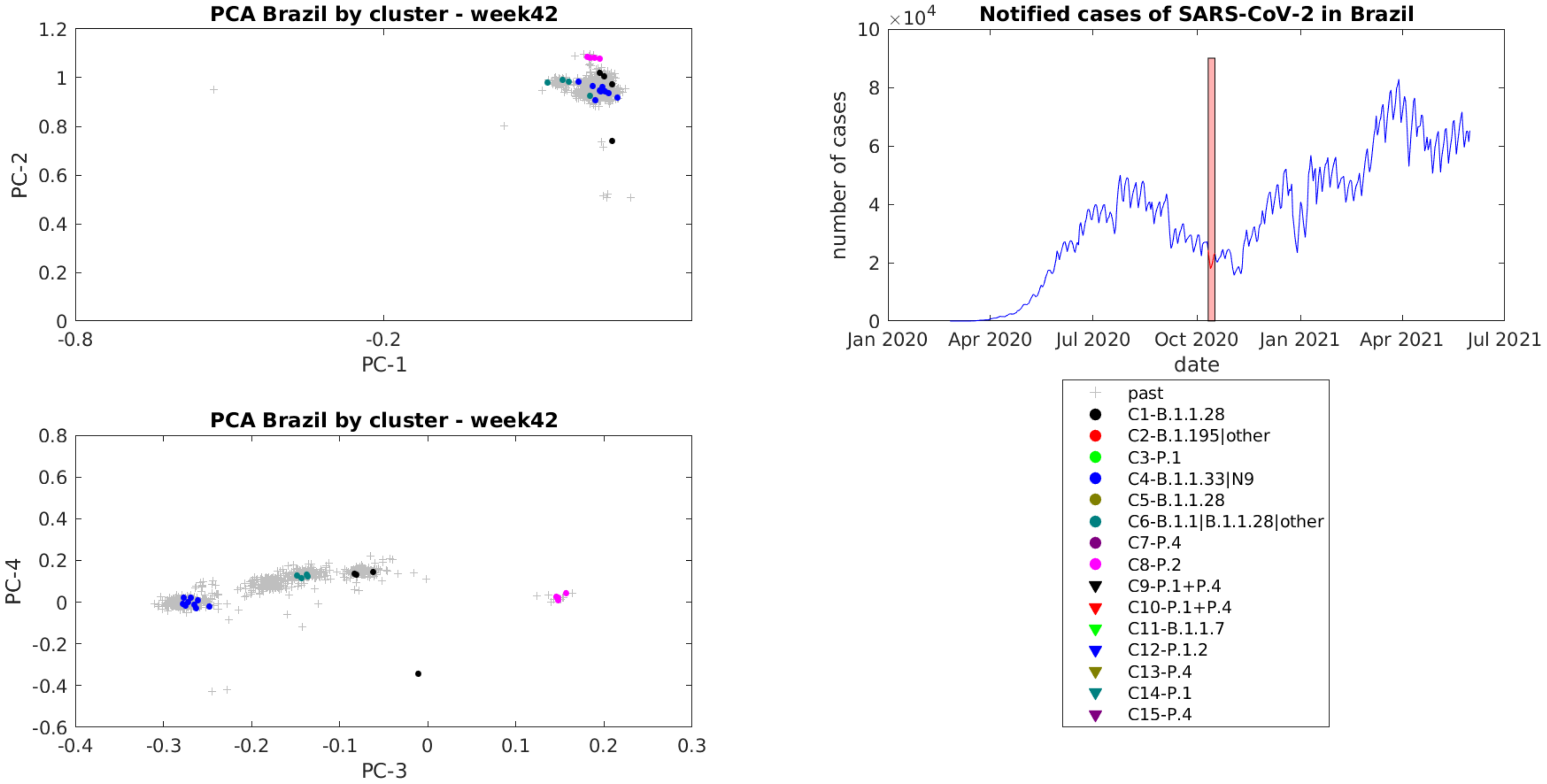

PCA Brazil by cluster - week42
PC-1
PC-2
Notified cases of SARS-CoV-2 in Brazil
×10^4
number of cases
date
Jan 2020
Apr 2020
Jul 2020
Oct 2020
Jan 2021
Apr 2021
Jul 2021
PCA Brazil by cluster - week42
PC-3
PC-4
past
C1-B.1.1.28
C2-B.1.195|other
C3-P.1
C4-B.1.1.33|N9
C5-B.1.1.28
C6-B.1.1|B.1.1.28|other
C7-P.4
C8-P.2
C9-P.1+P.4
C10-P.1+P.4
C11-B.1.1.7
C12-P.1.2
C13-P.4
C14-P.1
C15-P.4

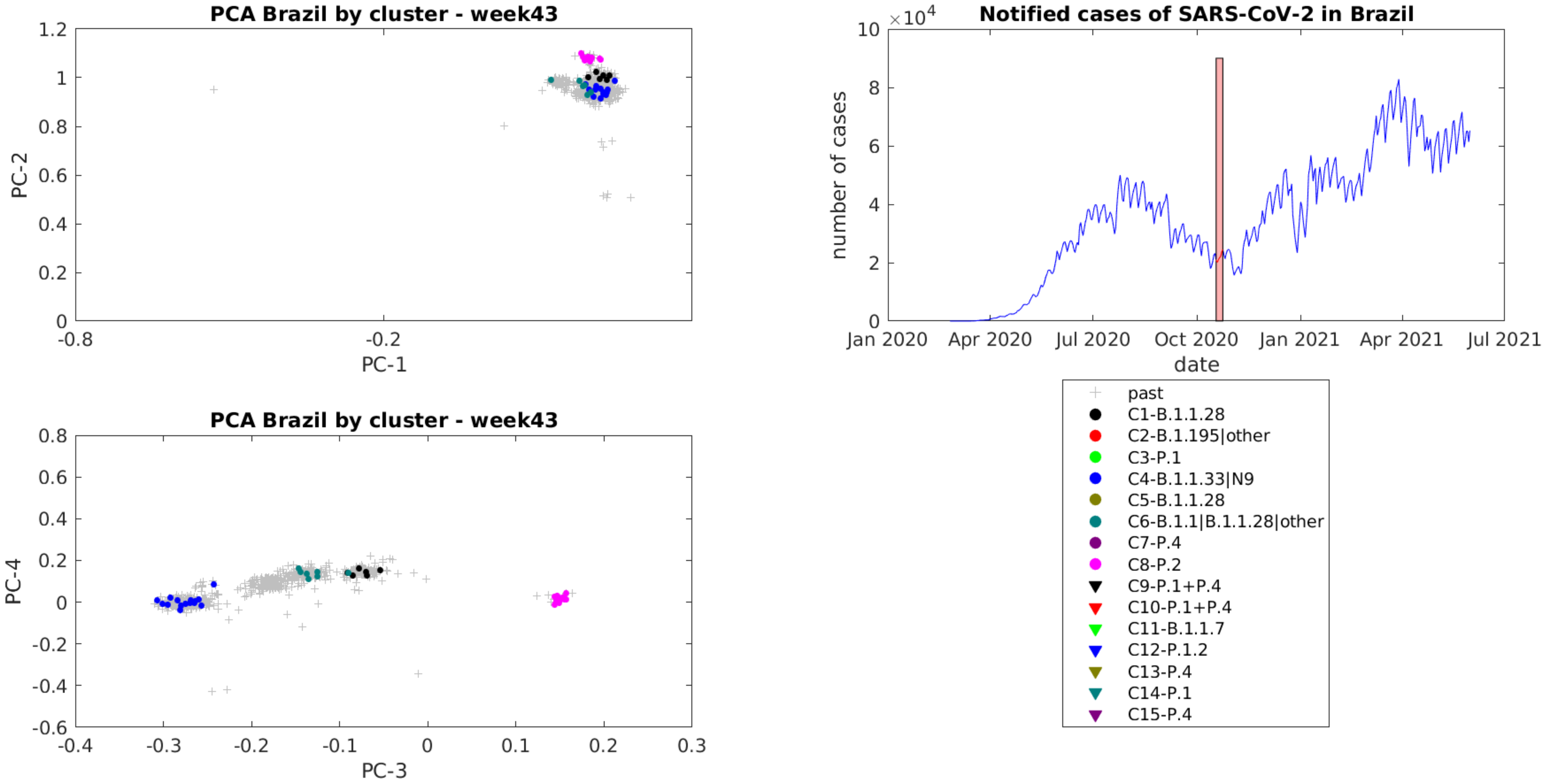
PCA Brazil by cluster - week43
PC-1
PC-2
1.2
1
0.8
0.6
0.4
0.2
0
-0.8
-0.2
Notified cases of SARS-CoV-2 in Brazil
×10^4
10
8
6
4
2
0
number of cases
date
Jan 2020
Apr 2020
Jul 2020
Oct 2020
Jan 2021
Apr 2021
Jul 2021
PCA Brazil by cluster - week43
PC-3
PC-4
0.8
0.6
0.4
0.2
0
-0.2
-0.4
-0.6
-0.4
-0.3
-0.2
-0.1
0
0.1
0.2
0.3
past
C1-B.1.1.28
C2-B.1.195|other
C3-P.1
C4-B.1.1.33|N9
C5-B.1.1.28
C6-B.1.1|B.1.1.28|other
C7-P.4
C8-P.2
C9-P.1+P.4
C10-P.1+P.4
C11-B.1.1.7
C12-P.1.2
C13-P.4
C14-P.1
C15-P.4

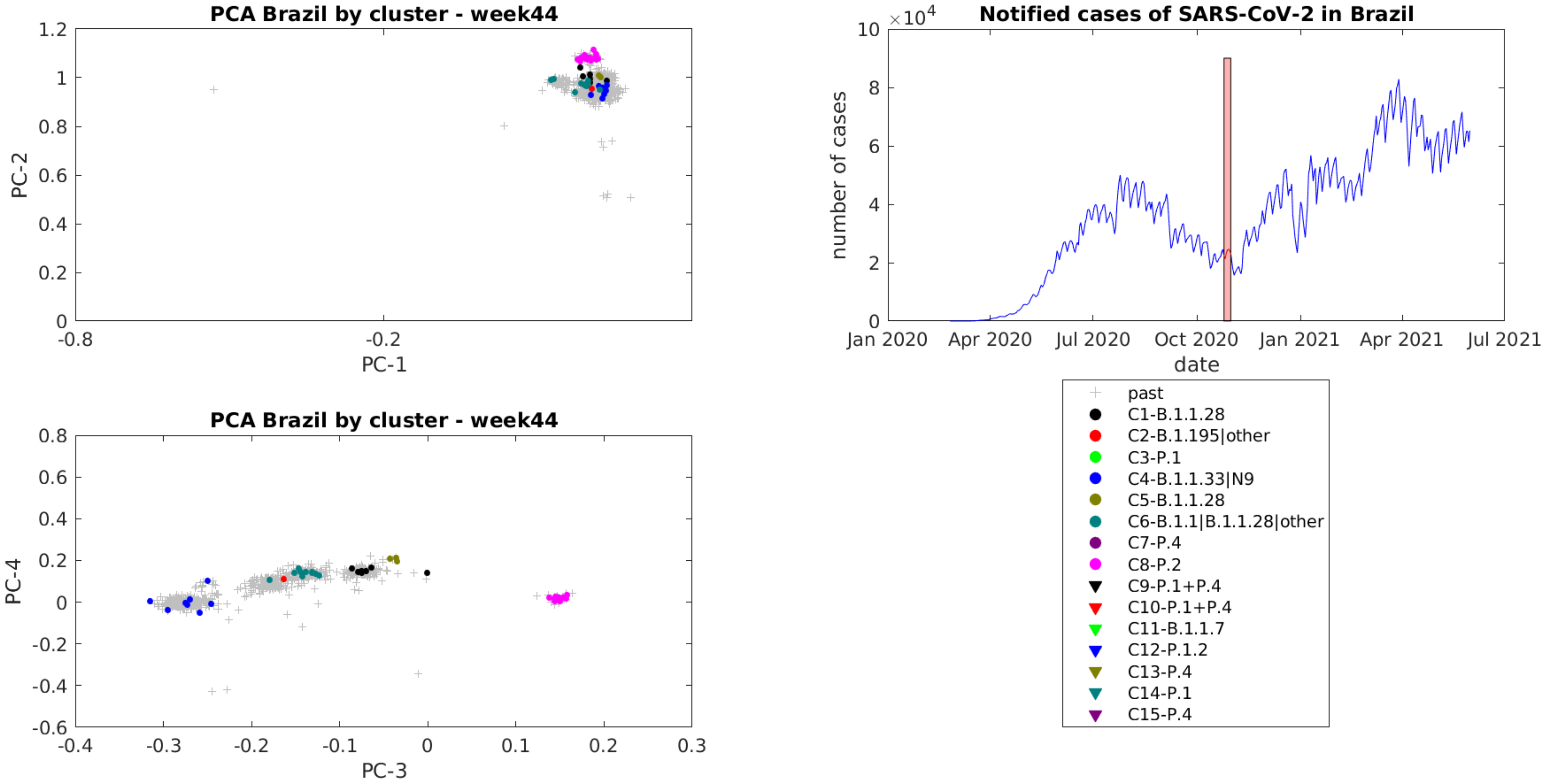

PCA Brazil by cluster - week44
PC-1
PC-2
PCA Brazil by cluster - week44
PC-3
PC-4
Notified cases of SARS-CoV-2 in Brazil
number of cases
date
Jan 2020
Apr 2020
Jul 2020
Oct 2020
Jan 2021
Apr 2021
Jul 2021
past
C1-B.1.1.28
C2-B.1.195|other
C3-P.1
C4-B.1.1.33|N9
C5-B.1.1.28
C6-B.1.1|B.1.1.28|other
C7-P.4
C8-P.2
C9-P.1+P.4
C10-P.1+P.4
C11-B.1.1.7
C12-P.1.2
C13-P.4
C14-P.1
C15-P.4

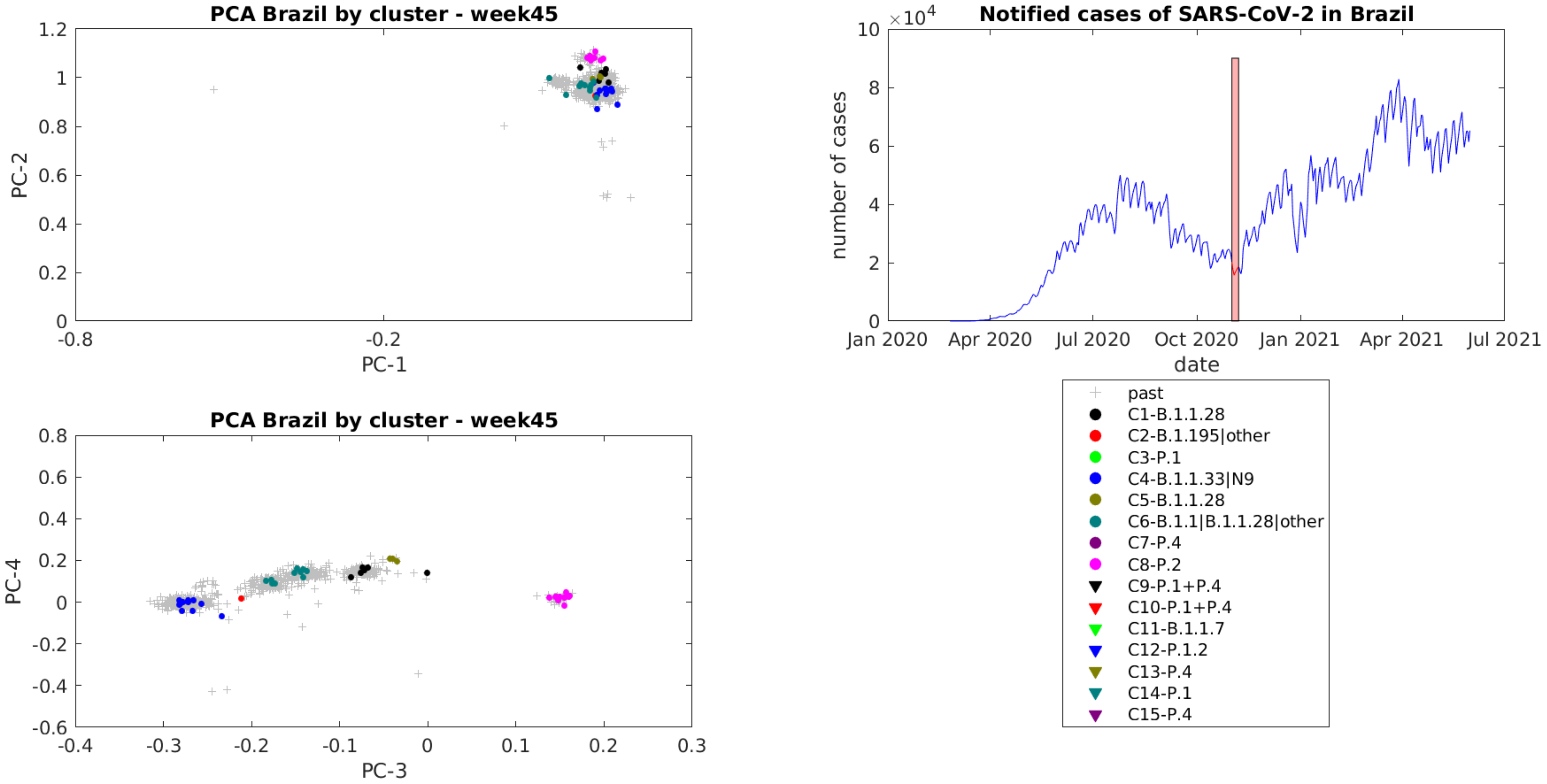
PCA Brazil by cluster - week45
PC-2
PC-1
PCA Brazil by cluster - week45
PC-4
PC-3
Notified cases of SARS-CoV-2 in Brazil
×10^4
number of cases
date
Jan 2020
Apr 2020
Jul 2020
Oct 2020
Jan 2021
Apr 2021
Jul 2021
past
C1-B.1.1.28
C2-B.1.195|other
C3-P.1
C4-B.1.1.33|N9
C5-B.1.1.28
C6-B.1.1|B.1.1.28|other
C7-P.4
C8-P.2
C9-P.1+P.4
C10-P.1+P.4
C11-B.1.1.7
C12-P.1.2
C13-P.4
C14-P.1
C15-P.4

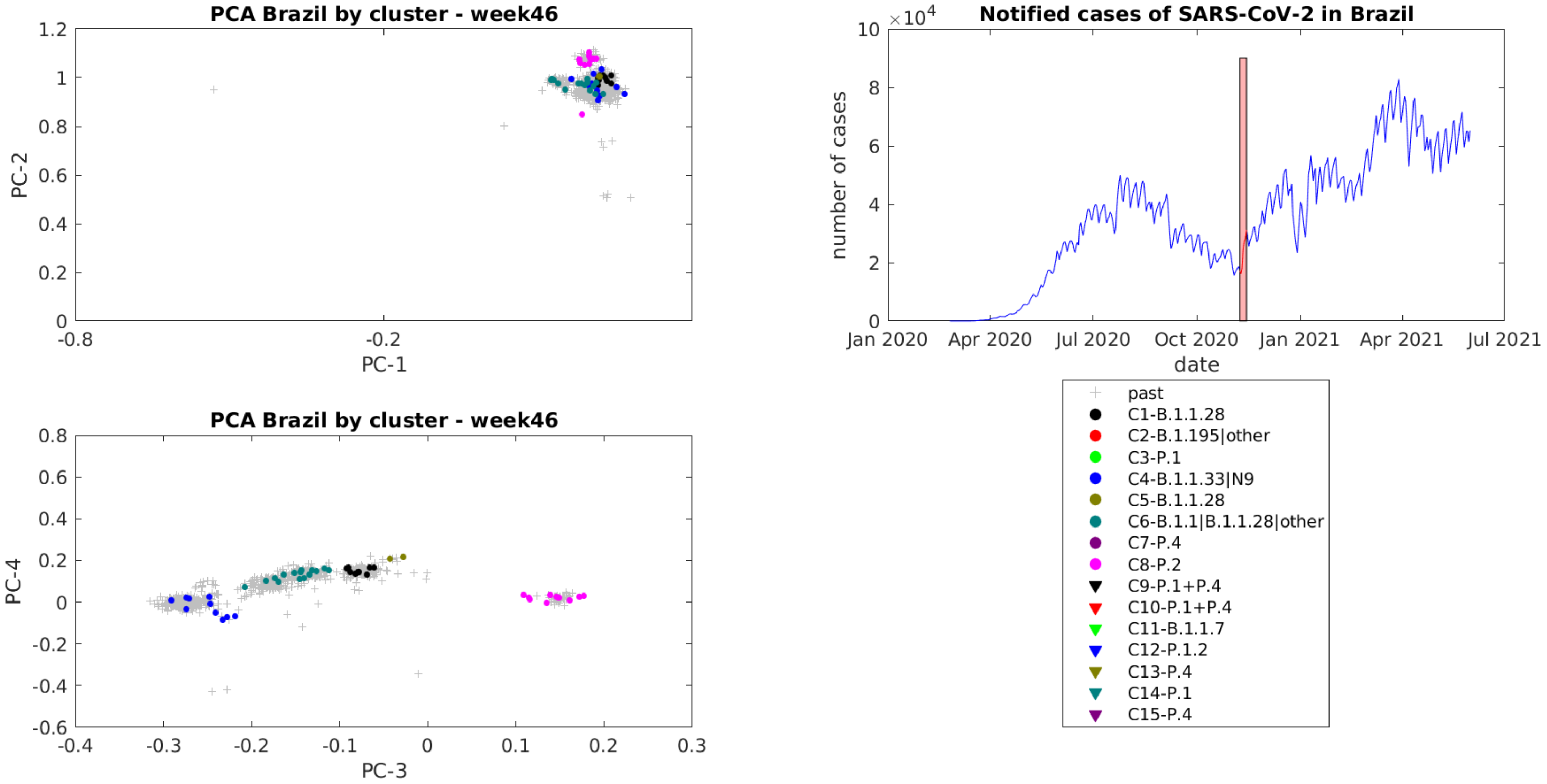
PCA Brazil by cluster - week46
PC-1
PC-2
1.2
1
0.8
0.6
0.4
0.2
0
-0.8
-0.2
Notified cases of SARS-CoV-2 in Brazil
×10^4
10
8
6
4
2
0
number of cases
Jan 2020
Apr 2020
Jul 2020
Oct 2020
Jan 2021
Apr 2021
Jul 2021
date
PCA Brazil by cluster - week46
PC-3
PC-4
0.8
0.6
0.4
0.2
0
-0.2
-0.4
-0.6
-0.4
-0.3
-0.2
-0.1
0
0.1
0.2
0.3
past
C1-B.1.1.28
C2-B.1.195|other
C3-P.1
C4-B.1.1.33|N9
C5-B.1.1.28
C6-B.1.1|B.1.1.28|other
C7-P.4
C8-P.2
C9-P.1+P.4
C10-P.1+P.4
C11-B.1.1.7
C12-P.1.2
C13-P.4
C14-P.1
C15-P.4

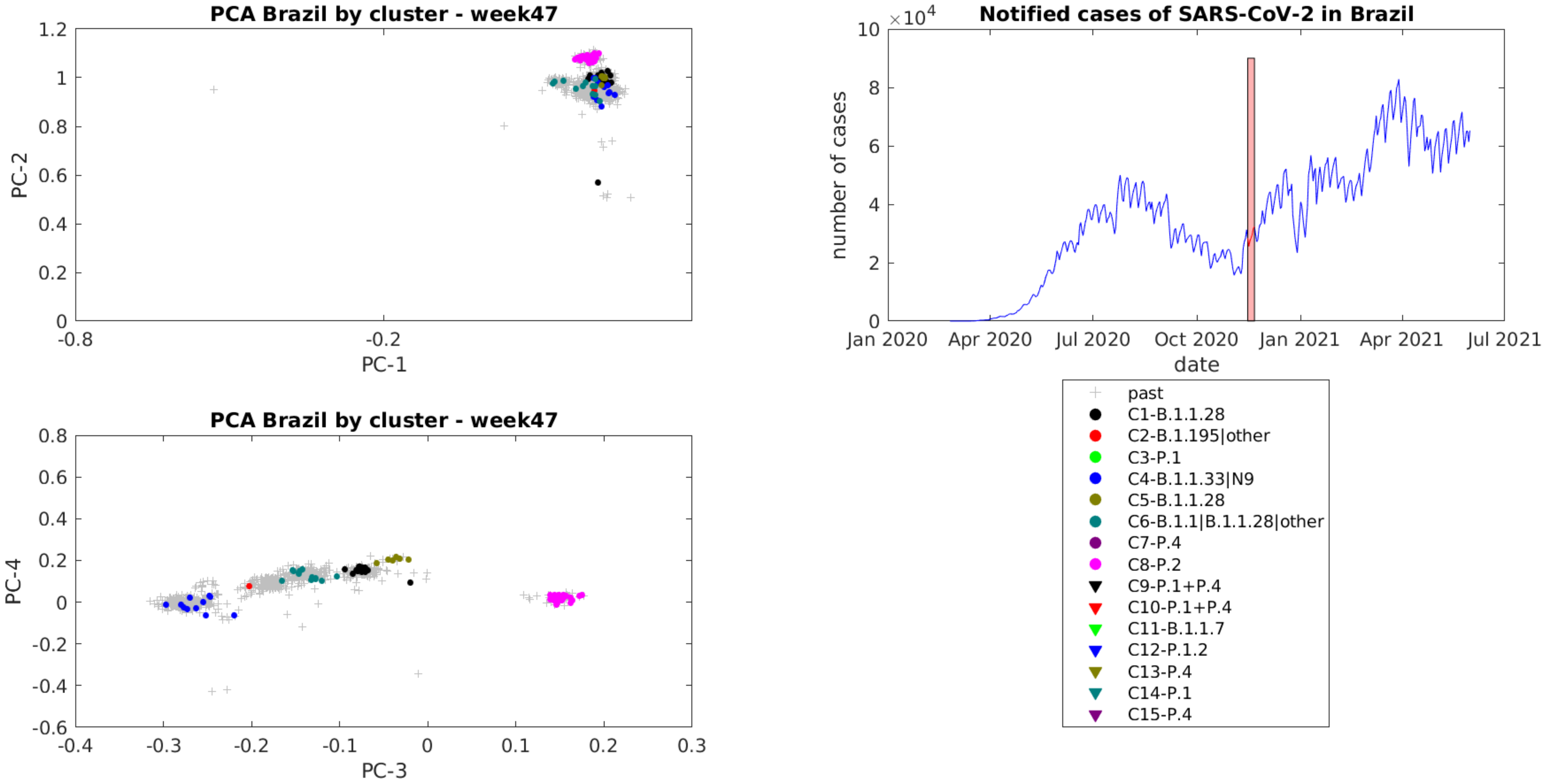
PCA Brazil by cluster - week47
PC-1
PC-2
Notified cases of SARS-CoV-2 in Brazil
×10^4
number of cases
date
Jan 2020
Apr 2020
Jul 2020
Oct 2020
Jan 2021
Apr 2021
Jul 2021
PCA Brazil by cluster - week47
PC-3
PC-4
past
C1-B.1.1.28
C2-B.1.195|other
C3-P.1
C4-B.1.1.33|N9
C5-B.1.1.28
C6-B.1.1|B.1.1.28|other
C7-P.4
C8-P.2
C9-P.1+P.4
C10-P.1+P.4
C11-B.1.1.7
C12-P.1.2
C13-P.4
C14-P.1
C15-P.4

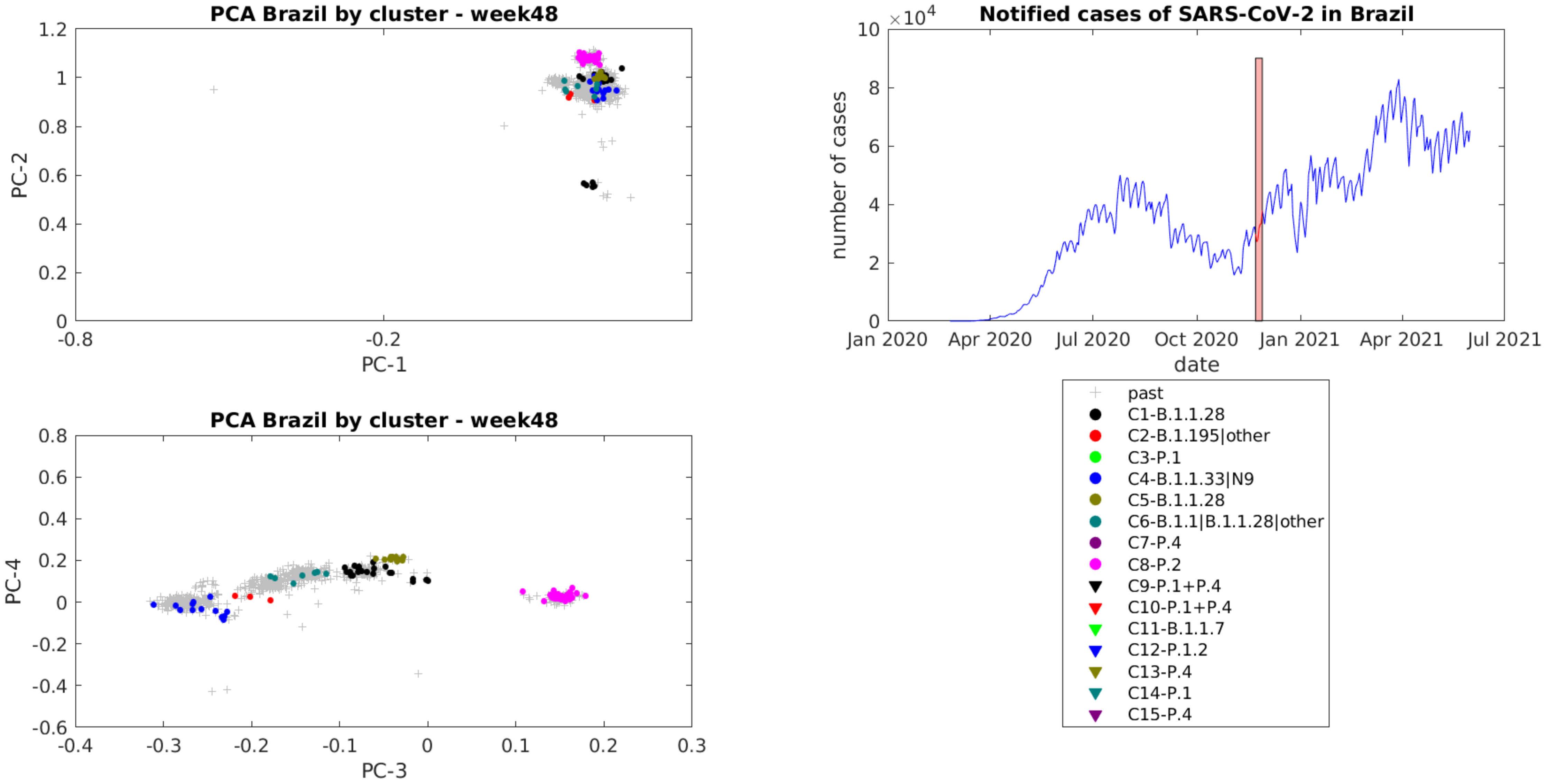
PCA Brazil by cluster - week48
PC-1
PC-2
-0.8
-0.2
0
0.2
0.4
0.6
0.8
1
1.2
Notified cases of SARS-CoV-2 in Brazil
×10^4
number of cases
date
0
2
4
6
8
10
Jan 2020
Apr 2020
Jul 2020
Oct 2020
Jan 2021
Apr 2021
Jul 2021
PCA Brazil by cluster - week48
PC-3
PC-4
-0.4
-0.3
-0.2
-0.1
0
0.1
0.2
0.3
-0.6
-0.4
-0.2
0
0.2
0.4
0.6
0.8
past
C1-B.1.1.28
C2-B.1.195|other
C3-P.1
C4-B.1.1.33|N9
C5-B.1.1.28
C6-B.1.1|B.1.1.28|other
C7-P.4
C8-P.2
C9-P.1+P.4
C10-P.1+P.4
C11-B.1.1.7
C12-P.1.2
C13-P.4
C14-P.1
C15-P.4

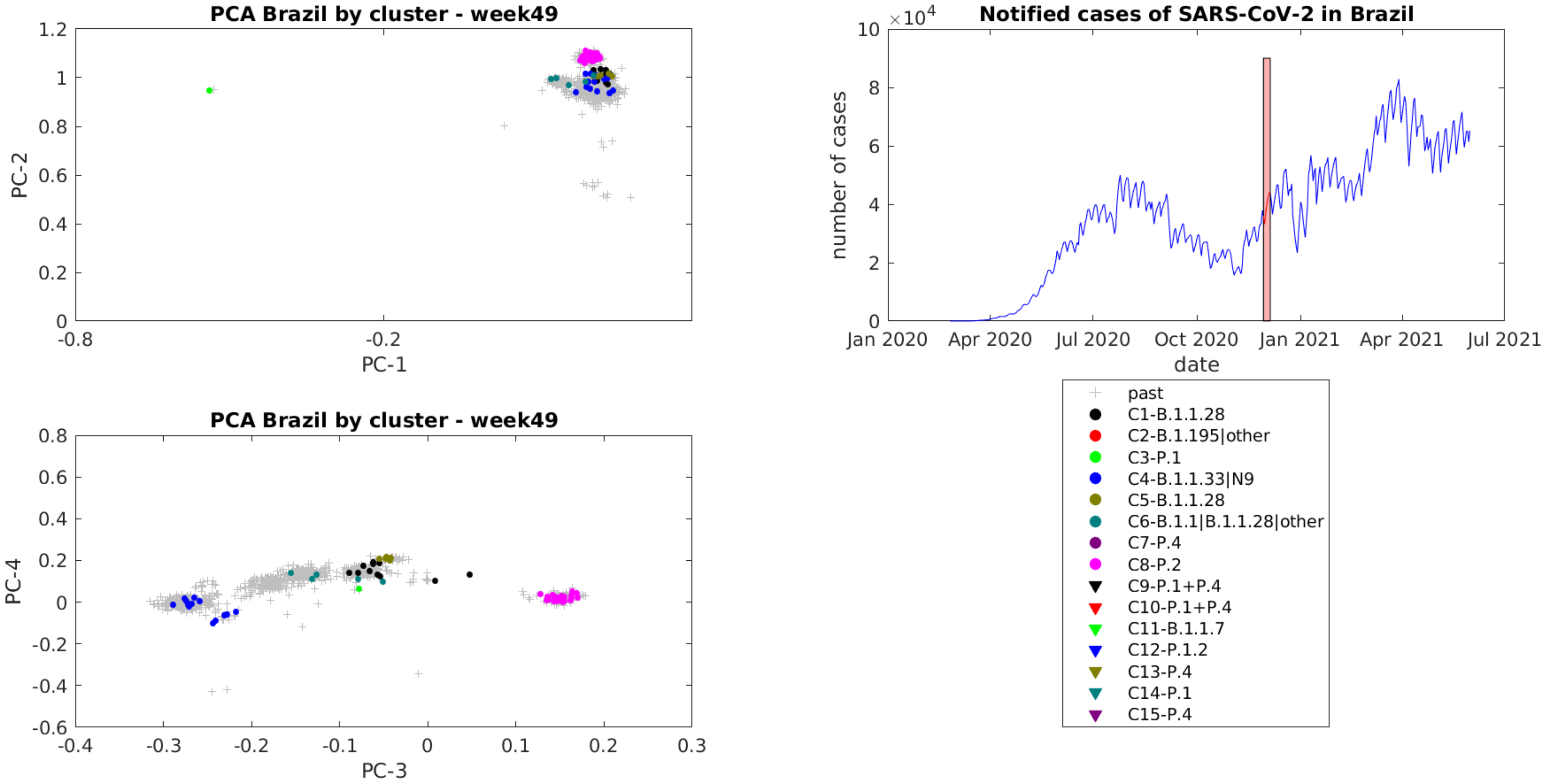
PCA Brazil by cluster - week49
PC-1
PC-2
Notified cases of SARS-CoV-2 in Brazil
×10^4
number of cases
date
Jan 2020
Apr 2020
Jul 2020
Oct 2020
Jan 2021
Apr 2021
Jul 2021
PCA Brazil by cluster - week49
PC-3
PC-4
past
C1-B.1.1.28
C2-B.1.195|other
C3-P.1
C4-B.1.1.33|N9
C5-B.1.1.28
C6-B.1.1|B.1.1.28|other
C7-P.4
C8-P.2
C9-P.1+P.4
C10-P.1+P.4
C11-B.1.1.7
C12-P.1.2
C13-P.4
C14-P.1
C15-P.4

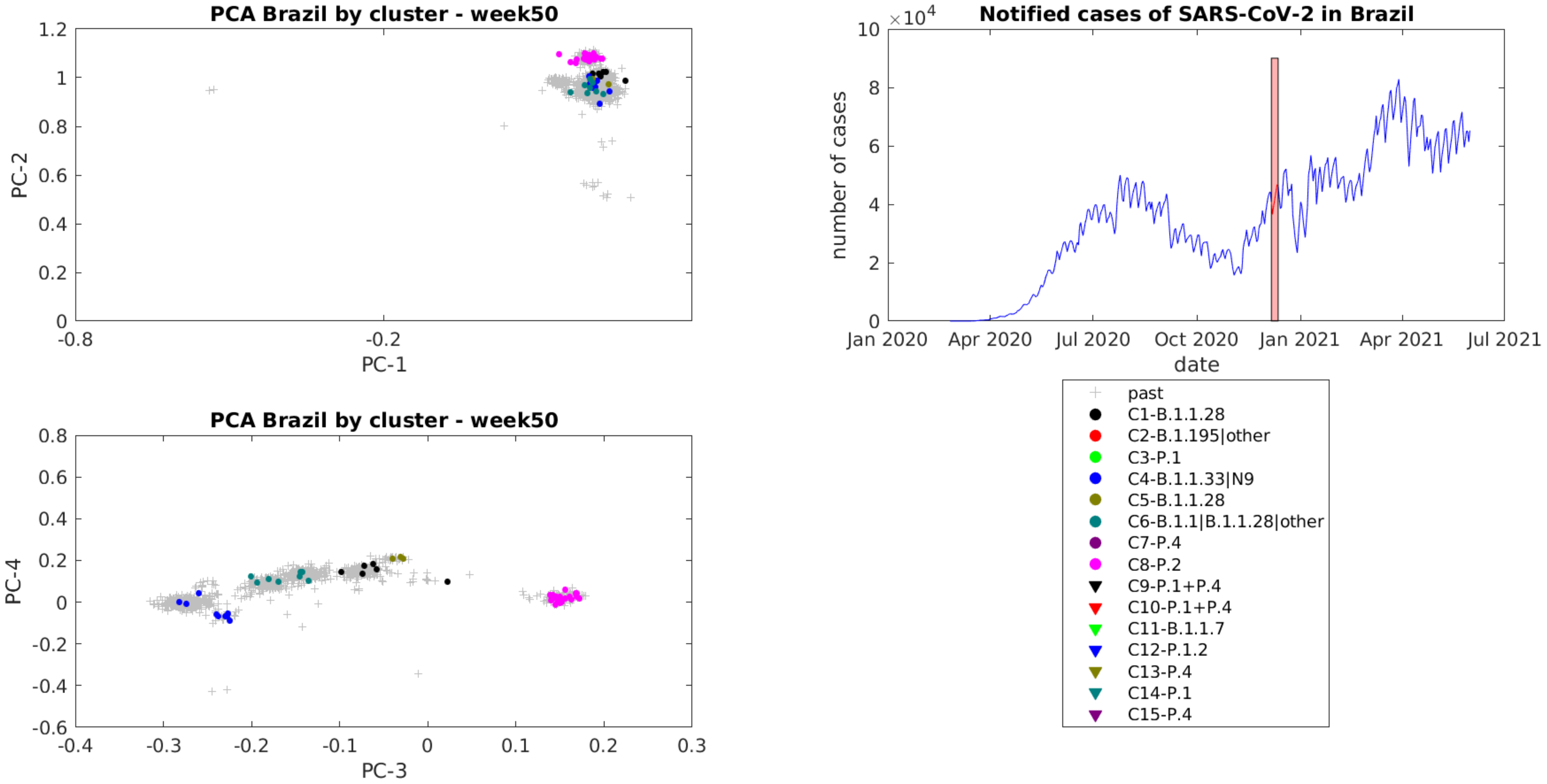
PCA Brazil by cluster - week50
PC-2
PC-1
1.2
1
0.8
0.6
0.4
0.2
0
-0.8
-0.2
Notified cases of SARS-CoV-2 in Brazil
$\times 10^4$
number of cases
date
10
8
6
4
2
0
Jan 2020
Apr 2020
Jul 2020
Oct 2020
Jan 2021
Apr 2021
Jul 2021
PCA Brazil by cluster - week50
PC-4
PC-3
0.8
0.6
0.4
0.2
0
-0.2
-0.4
-0.6
-0.4
-0.3
-0.2
-0.1
0
0.1
0.2
0.3
past
C1-B.1.1.28
C2-B.1.195|other
C3-P.1
C4-B.1.1.33|N9
C5-B.1.1.28
C6-B.1.1|B.1.1.28|other
C7-P.4
C8-P.2
C9-P.1+P.4
C10-P.1+P.4
C11-B.1.1.7
C12-P.1.2
C13-P.4
C14-P.1
C15-P.4

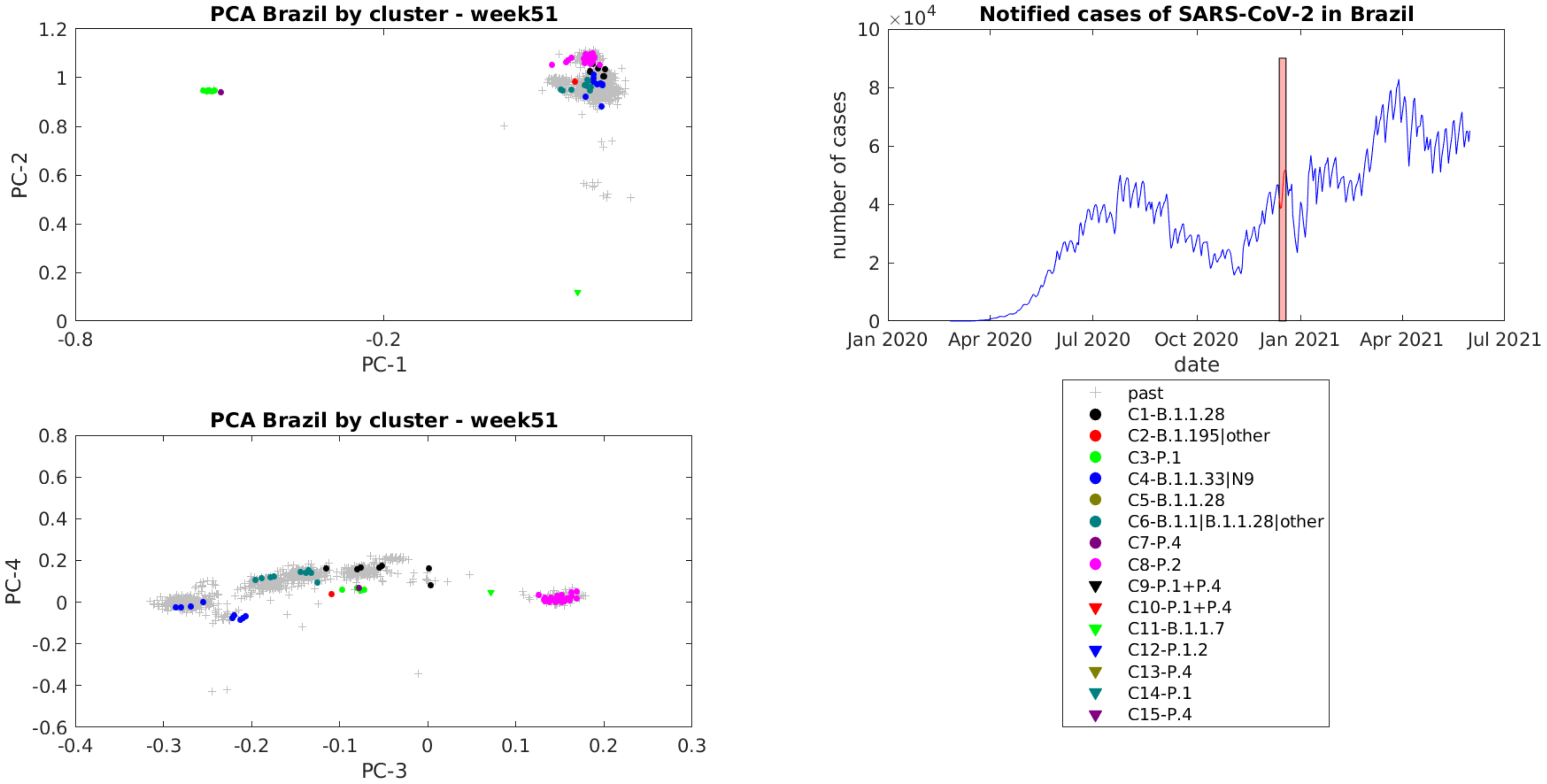
PCA Brazil by cluster - week51
PC-2
PC-1
PCA Brazil by cluster - week51
PC-4
PC-3
Notified cases of SARS-CoV-2 in Brazil
×10^4
number of cases
date
Jan 2020
Apr 2020
Jul 2020
Oct 2020
Jan 2021
Apr 2021
Jul 2021
past
C1-B.1.1.28
C2-B.1.195|other
C3-P.1
C4-B.1.1.33|N9
C5-B.1.1.28
C6-B.1.1|B.1.1.28|other
C7-P.4
C8-P.2
C9-P.1+P.4
C10-P.1+P.4
C11-B.1.1.7
C12-P.1.2
C13-P.4
C14-P.1
C15-P.4

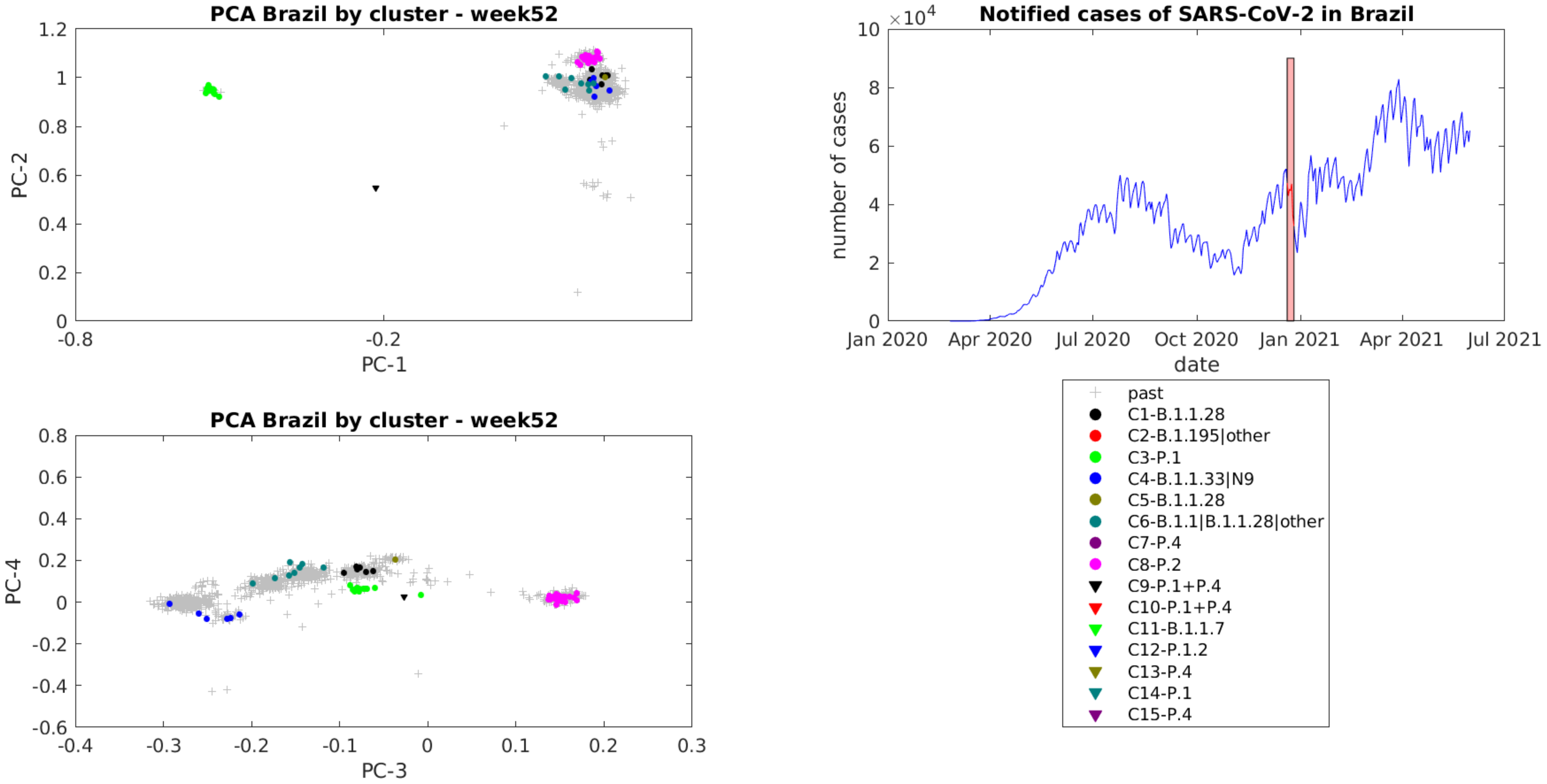
PCA Brazil by cluster - week52
PC-1
PC-2
-0.8
-0.2
1.2
1
0.8
0.6
0.4
0.2
0
Notified cases of SARS-CoV-2 in Brazil
×10^4
10
8
6
4
2
0
number of cases
Jan 2020
Apr 2020
Jul 2020
Oct 2020
Jan 2021
Apr 2021
Jul 2021
date
PCA Brazil by cluster - week52
PC-3
PC-4
0.8
0.6
0.4
0.2
0
-0.2
-0.4
-0.6
-0.4
-0.3
-0.2
-0.1
0
0.1
0.2
0.3
past
C1-B.1.1.28
C2-B.1.195|other
C3-P.1
C4-B.1.1.33|N9
C5-B.1.1.28
C6-B.1.1|B.1.1.28|other
C7-P.4
C8-P.2
C9-P.1+P.4
C10-P.1+P.4
C11-B.1.1.7
C12-P.1.2
C13-P.4
C14-P.1
C15-P.4

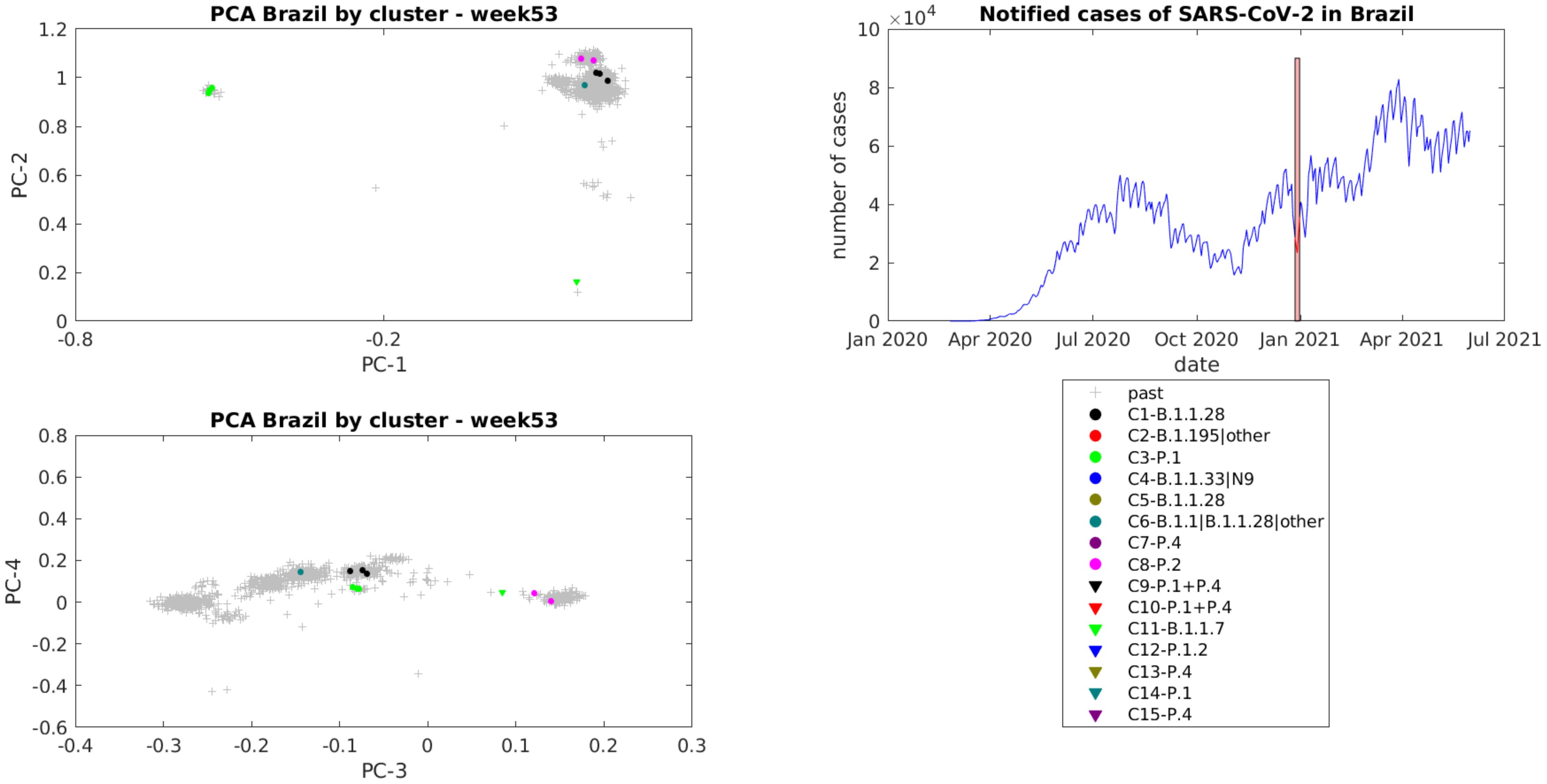
PCA Brazil by cluster - week53
PC-1
PC-2
Notified cases of SARS-CoV-2 in Brazil
×10^4
number of cases
date
Jan 2020
Apr 2020
Jul 2020
Oct 2020
Jan 2021
Apr 2021
Jul 2021
PCA Brazil by cluster - week53
PC-3
PC-4
past
C1-B.1.1.28
C2-B.1.195|other
C3-P.1
C4-B.1.1.33|N9
C5-B.1.1.28
C6-B.1.1|B.1.1.28|other
C7-P.4
C8-P.2
C9-P.1+P.4
C10-P.1+P.4
C11-B.1.1.7
C12-P.1.2
C13-P.4
C14-P.1
C15-P.4

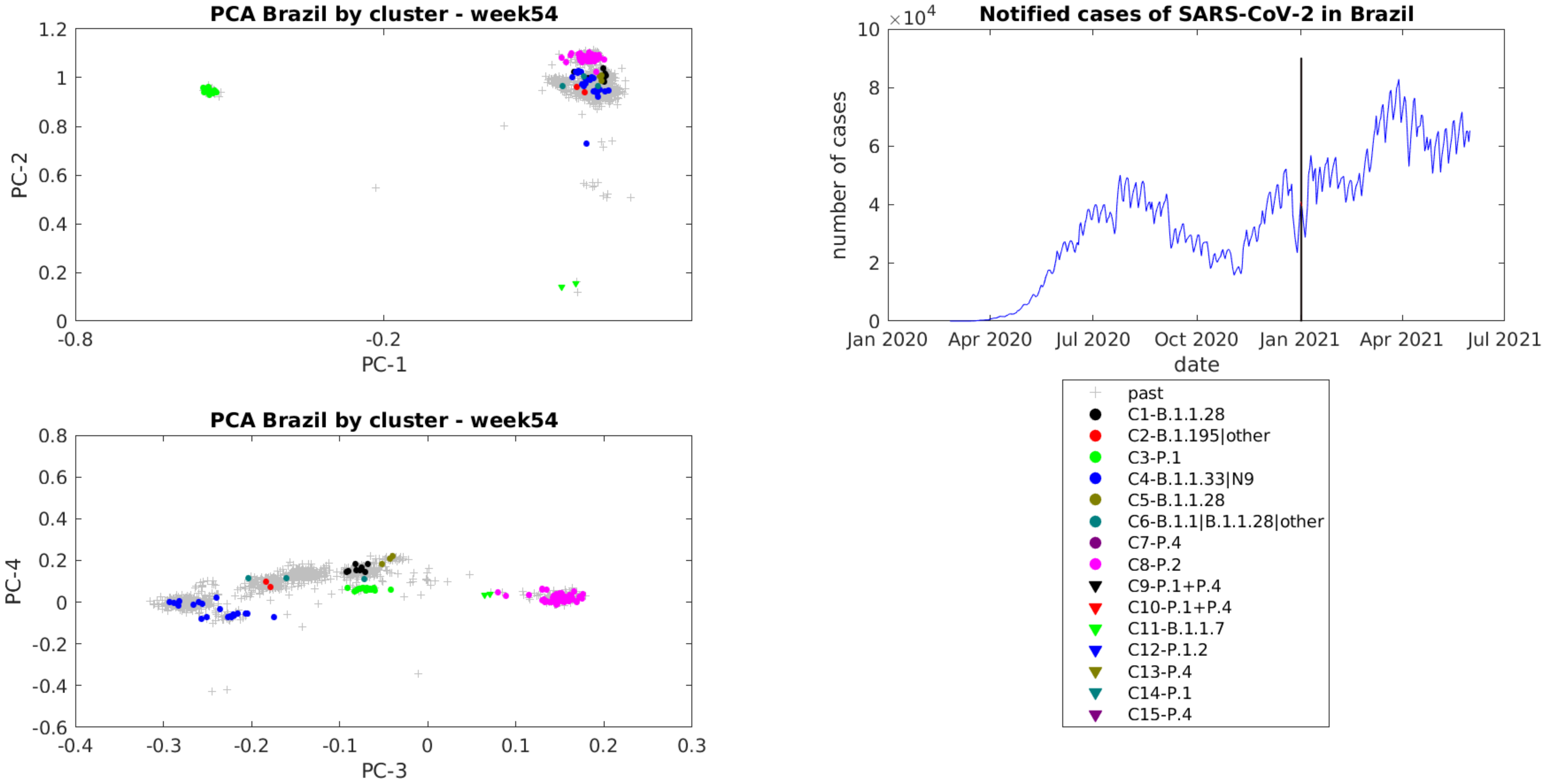
PCA Brazil by cluster - week54
PC-2
PC-1
1.2
1
0.8
0.6
0.4
0.2
0
-0.8
-0.2
Notified cases of SARS-CoV-2 in Brazil
$\times 10^4$
number of cases
date
10
8
6
4
2
0
Jan 2020
Apr 2020
Jul 2020
Oct 2020
Jan 2021
Apr 2021
Jul 2021
PCA Brazil by cluster - week54
PC-4
PC-3
0.8
0.6
0.4
0.2
0
-0.2
-0.4
-0.6
-0.4
-0.3
-0.2
-0.1
0
0.1
0.2
0.3
past
C1-B.1.1.28
C2-B.1.195|other
C3-P.1
C4-B.1.1.33|N9
C5-B.1.1.28
C6-B.1.1|B.1.1.28|other
C7-P.4
C8-P.2
C9-P.1+P.4
C10-P.1+P.4
C11-B.1.1.7
C12-P.1.2
C13-P.4
C14-P.1
C15-P.4

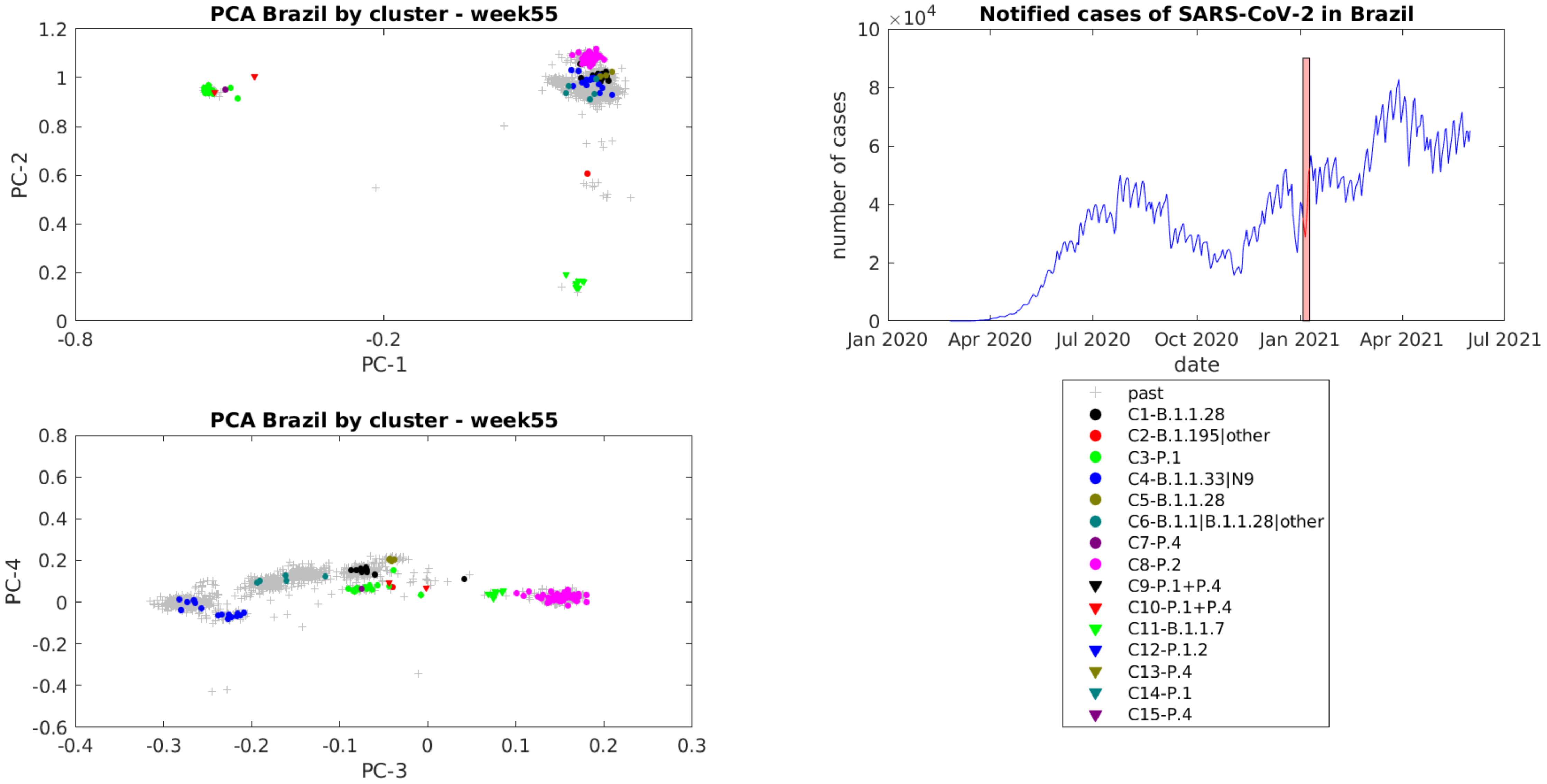

PCA Brazil by cluster - week55
PC-1
PC-2
PCA Brazil by cluster - week55
PC-3
PC-4
Notified cases of SARS-CoV-2 in Brazil
×10^4
number of cases
date
Jan 2020
Apr 2020
Jul 2020
Oct 2020
Jan 2021
Apr 2021
Jul 2021
past
C1-B.1.1.28
C2-B.1.195|other
C3-P.1
C4-B.1.1.33|N9
C5-B.1.1.28
C6-B.1.1|B.1.1.28|other
C7-P.4
C8-P.2
C9-P.1+P.4
C10-P.1+P.4
C11-B.1.1.7
C12-P.1.2
C13-P.4
C14-P.1
C15-P.4

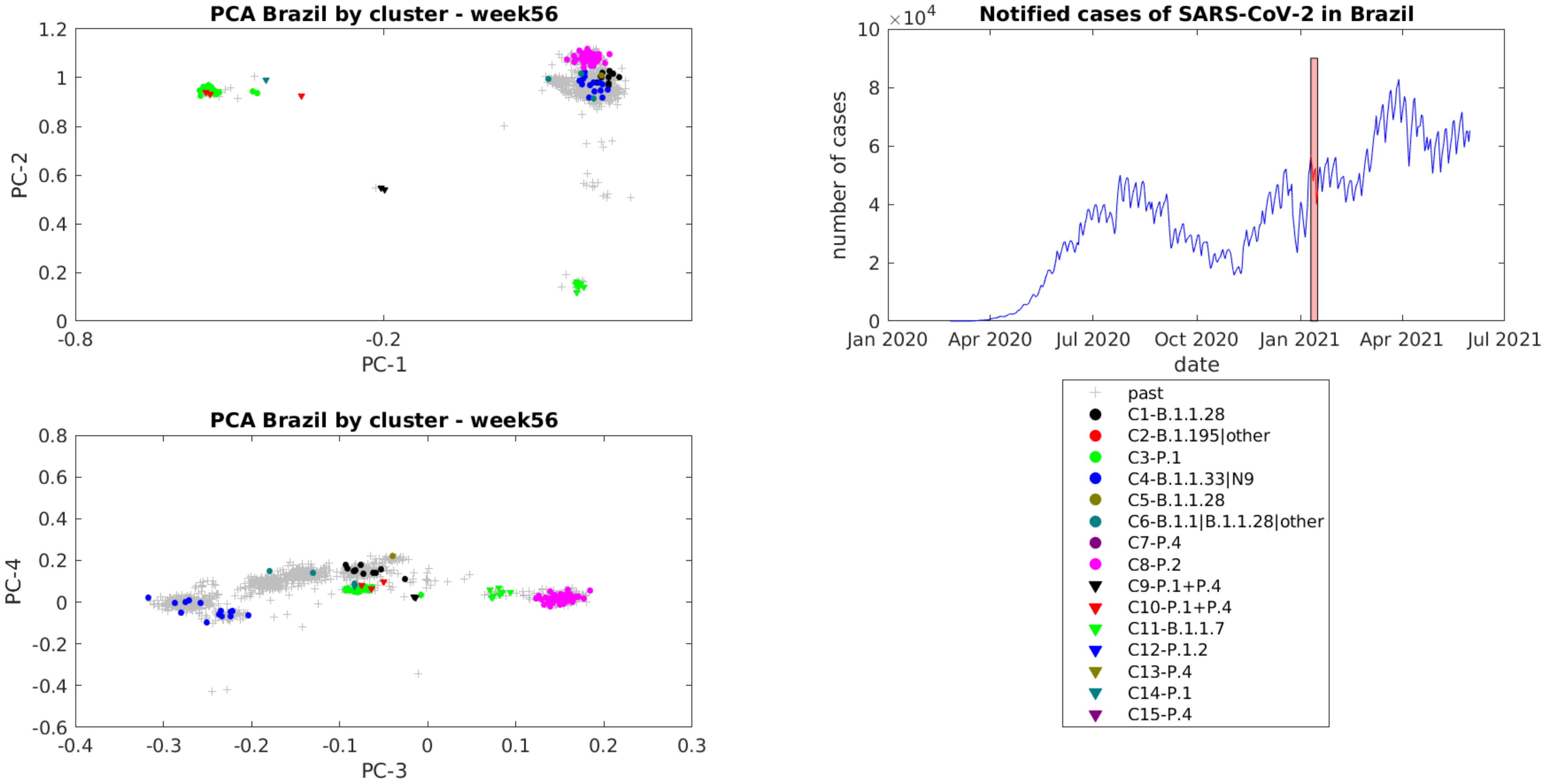

PCA Brazil by cluster - week56
PC-1
PC-2
Notified cases of SARS-CoV-2 in Brazil
×10^4
number of cases
date
Jan 2020
Apr 2020
Jul 2020
Oct 2020
Jan 2021
Apr 2021
Jul 2021
PCA Brazil by cluster - week56
PC-3
PC-4
past
C1-B.1.1.28
C2-B.1.195|other
C3-P.1
C4-B.1.1.33|N9
C5-B.1.1.28
C6-B.1.1|B.1.1.28|other
C7-P.4
C8-P.2
C9-P.1+P.4
C10-P.1+P.4
C11-B.1.1.7
C12-P.1.2
C13-P.4
C14-P.1
C15-P.4

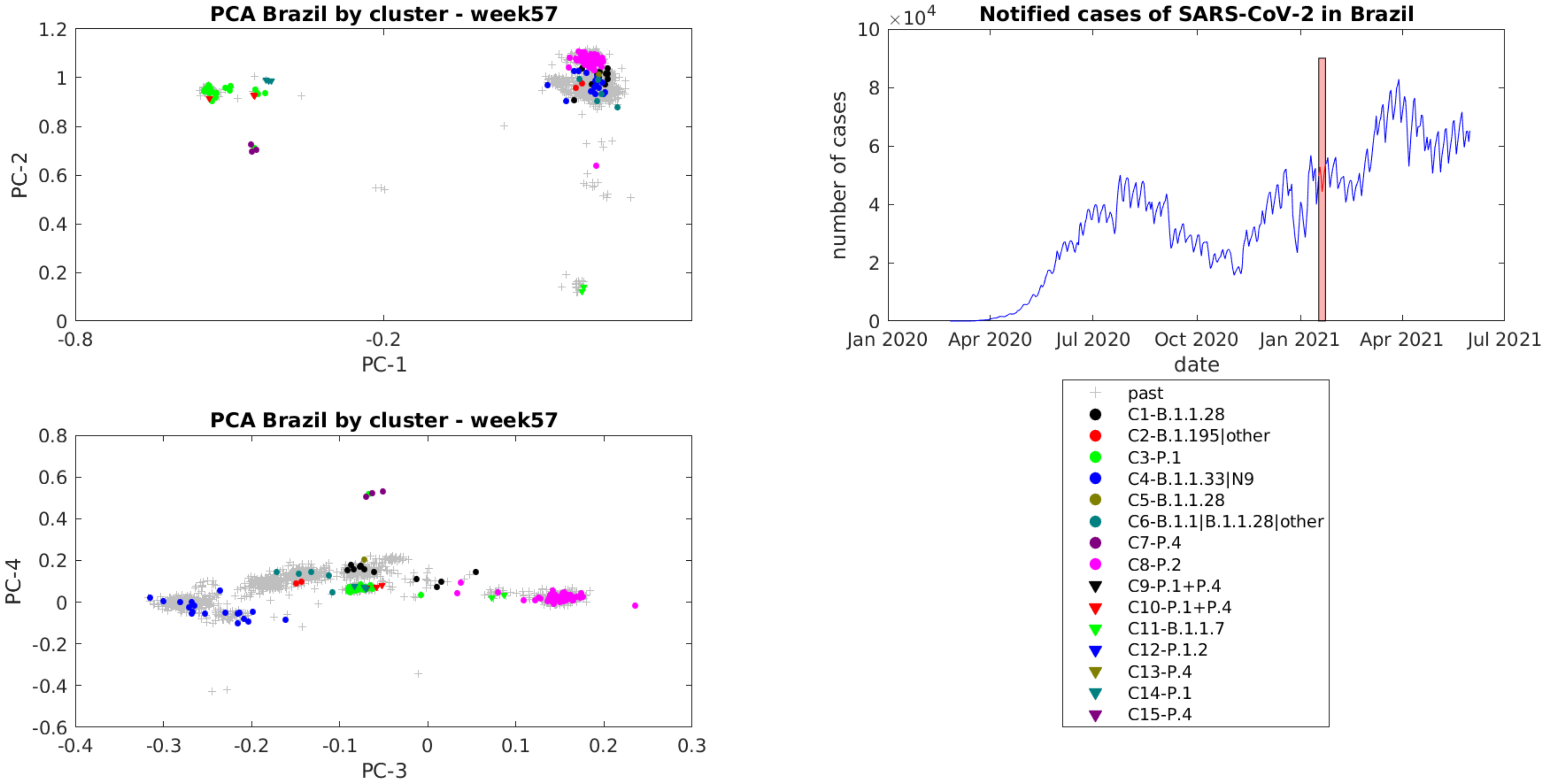

PCA Brazil by cluster - week57
PC-2
PC-1
1.2
1
0.8
0.6
0.4
0.2
0
-0.8
-0.2
PCA Brazil by cluster - week57
PC-4
PC-3
0.8
0.6
0.4
0.2
0
-0.2
-0.4
-0.6
-0.4
-0.3
-0.2
-0.1
0
0.1
0.2
0.3
Notified cases of SARS-CoV-2 in Brazil
×10^4
number of cases
date
10
8
6
4
2
0
Jan 2020
Apr 2020
Jul 2020
Oct 2020
Jan 2021
Apr 2021
Jul 2021
past
C1-B.1.1.28
C2-B.1.195|other
C3-P.1
C4-B.1.1.33|N9
C5-B.1.1.28
C6-B.1.1|B.1.1.28|other
C7-P.4
C8-P.2
C9-P.1+P.4
C10-P.1+P.4
C11-B.1.1.7
C12-P.1.2
C13-P.4
C14-P.1
C15-P.4

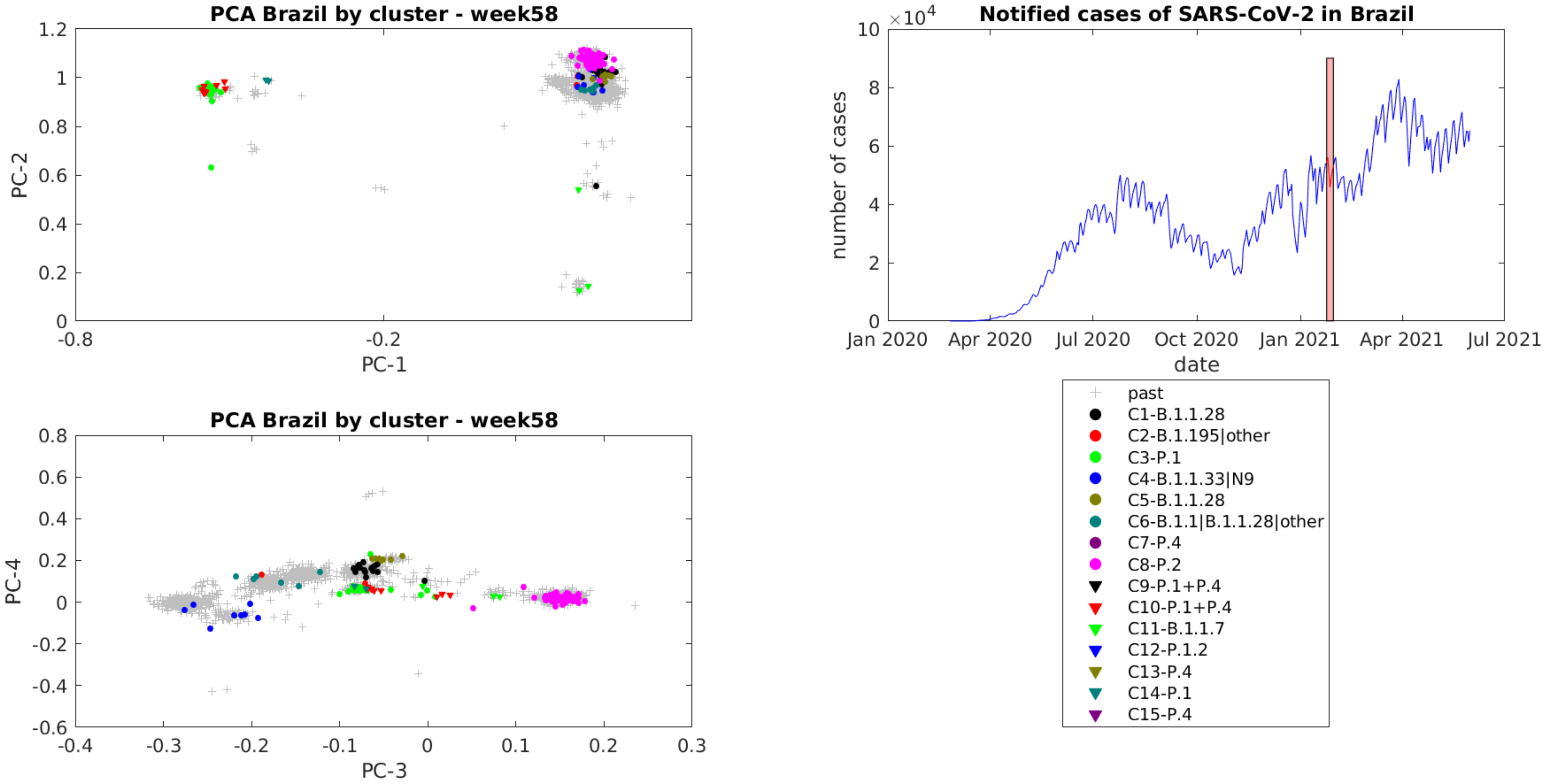
PCA Brazil by cluster - week58
PC-2
PC-1
1.2
1
0.8
0.6
0.4
0.2
0
-0.8
-0.2
Notified cases of SARS-CoV-2 in Brazil
×10^4
10
8
6
4
2
0
number of cases
Jan 2020
Apr 2020
Jul 2020
Oct 2020
Jan 2021
Apr 2021
Jul 2021
date
PCA Brazil by cluster - week58
PC-4
PC-3
0.8
0.6
0.4
0.2
0
-0.2
-0.4
-0.6
-0.4
-0.3
-0.2
-0.1
0
0.1
0.2
0.3
past
C1-B.1.1.28
C2-B.1.195|other
C3-P.1
C4-B.1.1.33|N9
C5-B.1.1.28
C6-B.1.1|B.1.1.28|other
C7-P.4
C8-P.2
C9-P.1+P.4
C10-P.1+P.4
C11-B.1.1.7
C12-P.1.2
C13-P.4
C14-P.1
C15-P.4

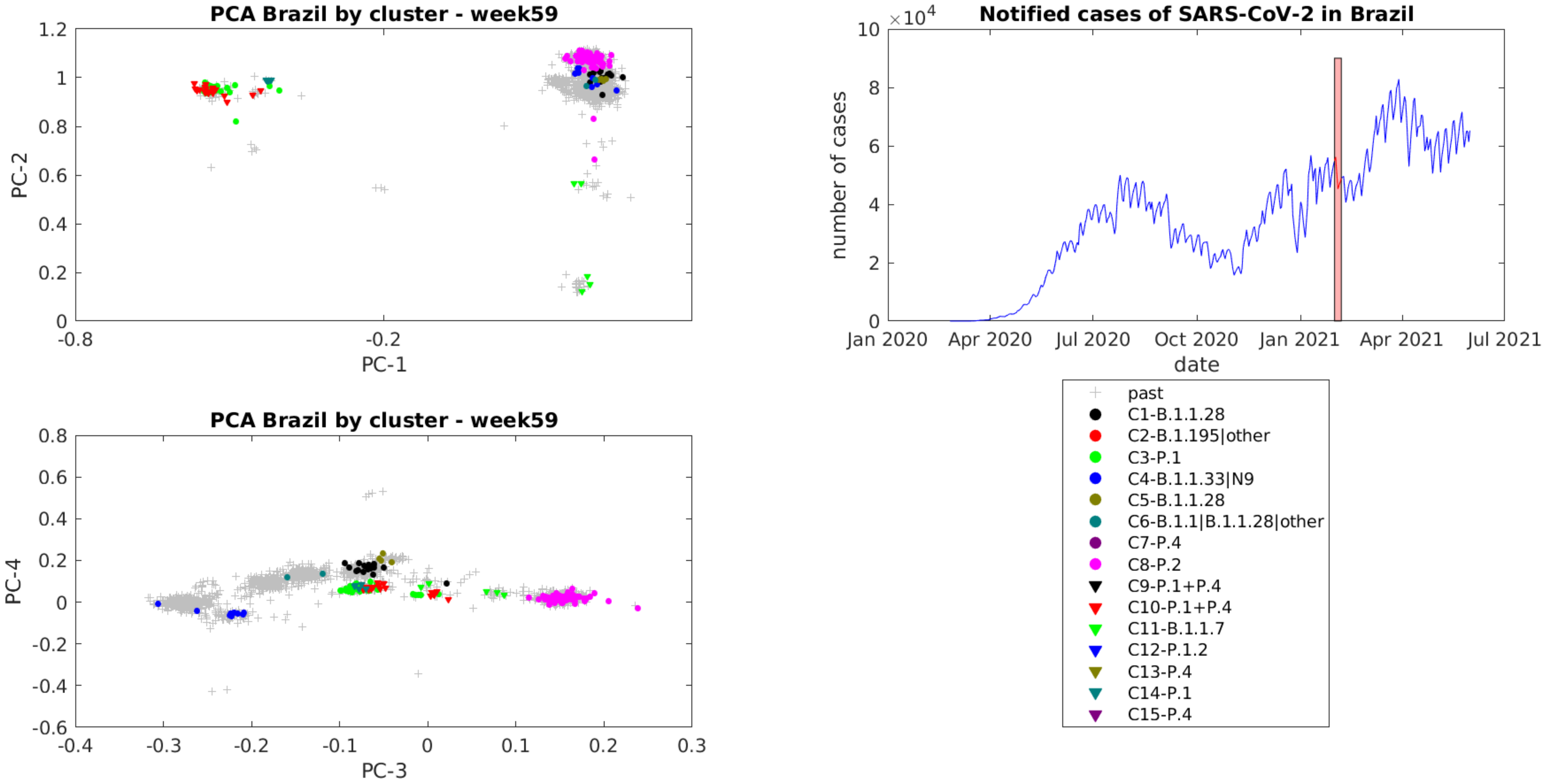
PCA Brazil by cluster - week59
PC-1
PC-2
Notified cases of SARS-CoV-2 in Brazil
$\times 10^4$
number of cases
date
Jan 2020
Apr 2020
Jul 2020
Oct 2020
Jan 2021
Apr 2021
Jul 2021
PCA Brazil by cluster - week59
PC-3
PC-4
past
C1-B.1.1.28
C2-B.1.195|other
C3-P.1
C4-B.1.1.33|N9
C5-B.1.1.28
C6-B.1.1|B.1.1.28|other
C7-P.4
C8-P.2
C9-P.1+P.4
C10-P.1+P.4
C11-B.1.1.7
C12-P.1.2
C13-P.4
C14-P.1
C15-P.4

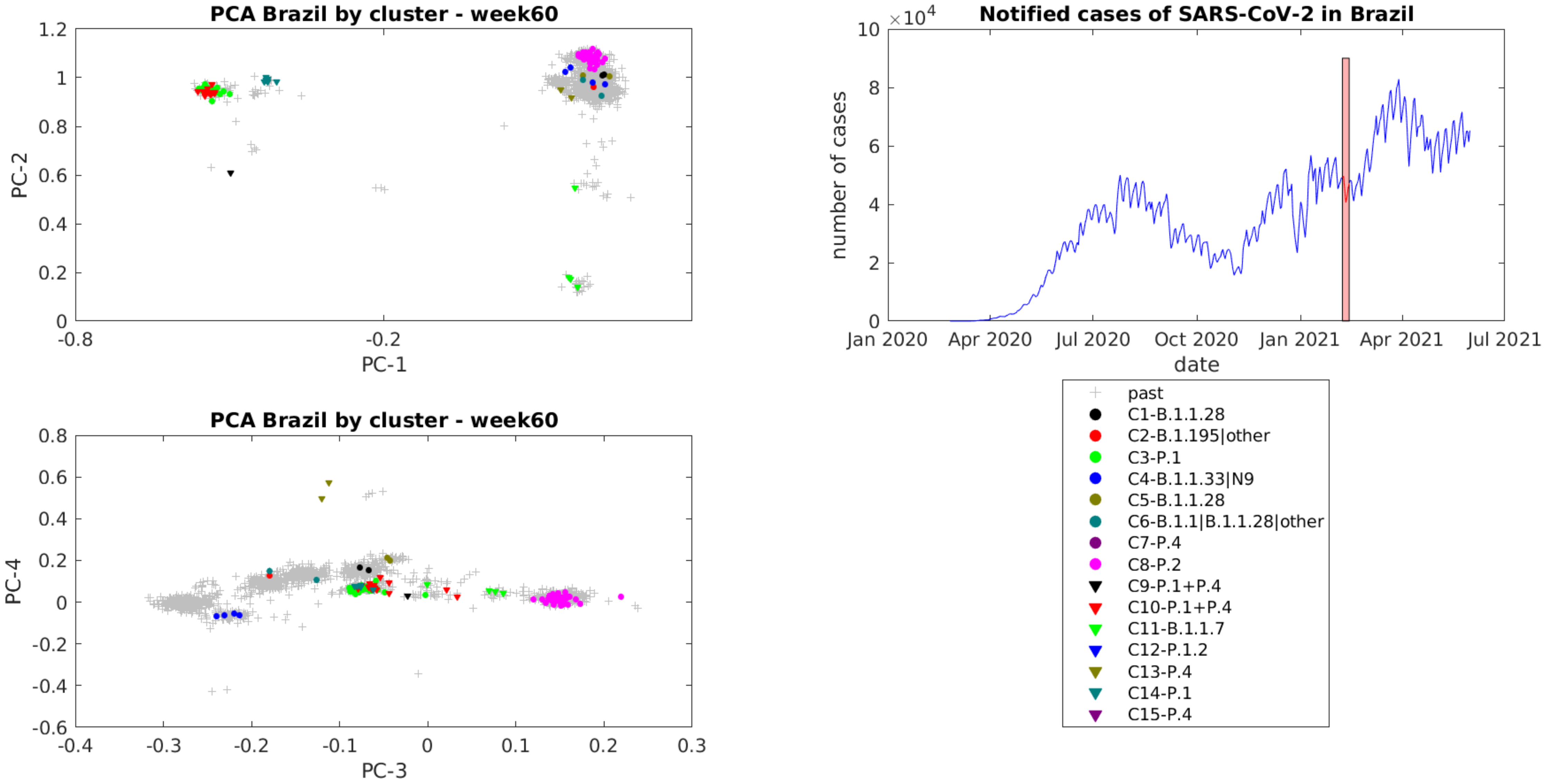

PCA Brazil by cluster - week60
PC-1
PC-2
PCA Brazil by cluster - week60
PC-3
PC-4
Notified cases of SARS-CoV-2 in Brazil
$\times 10^4$
number of cases
date
Jan 2020
Apr 2020
Jul 2020
Oct 2020
Jan 2021
Apr 2021
Jul 2021
past
C1-B.1.1.28
C2-B.1.195|other
C3-P.1
C4-B.1.1.33|N9
C5-B.1.1.28
C6-B.1.1|B.1.1.28|other
C7-P.4
C8-P.2
C9-P.1+P.4
C10-P.1+P.4
C11-B.1.1.7
C12-P.1.2
C13-P.4
C14-P.1
C15-P.4

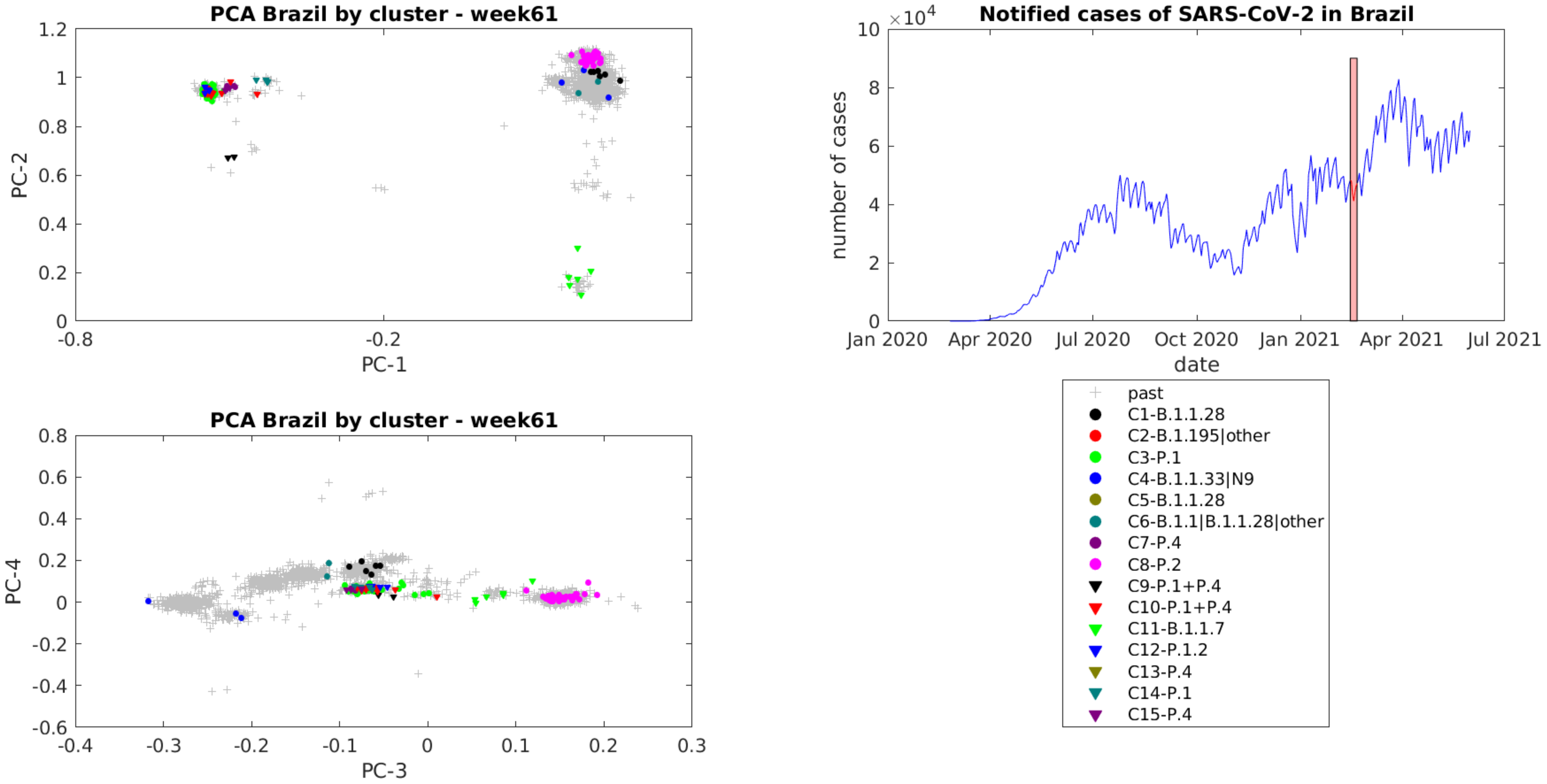
PCA Brazil by cluster - week61
PC-1
PC-2
Notified cases of SARS-CoV-2 in Brazil
×10^4
number of cases
date
Jan 2020
Apr 2020
Jul 2020
Oct 2020
Jan 2021
Apr 2021
Jul 2021
PCA Brazil by cluster - week61
PC-3
PC-4
past
C1-B.1.1.28
C2-B.1.195|other
C3-P.1
C4-B.1.1.33|N9
C5-B.1.1.28
C6-B.1.1|B.1.1.28|other
C7-P.4
C8-P.2
C9-P.1+P.4
C10-P.1+P.4
C11-B.1.1.7
C12-P.1.2
C13-P.4
C14-P.1
C15-P.4

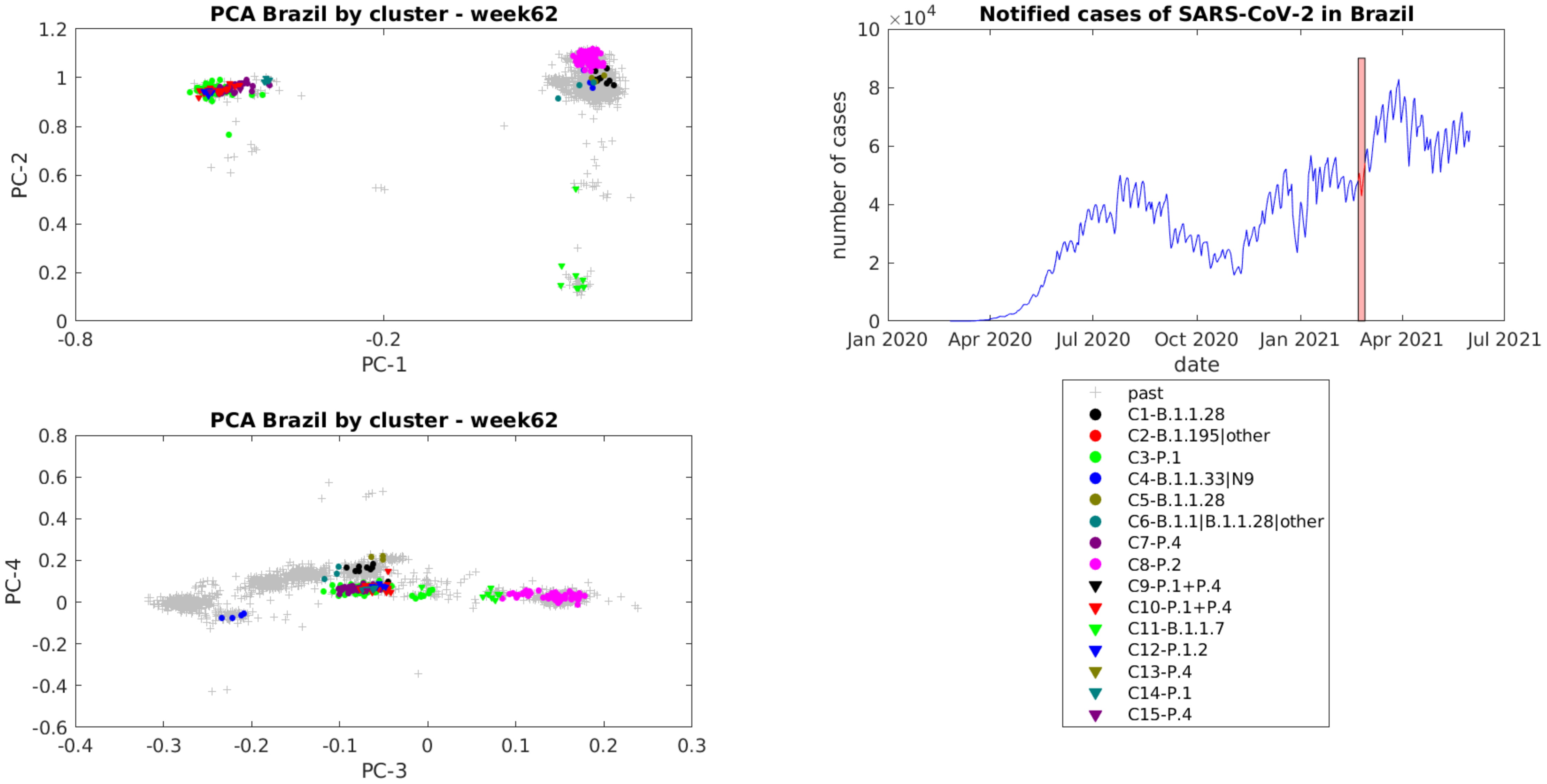
PCA Brazil by cluster - week62
PC-1
PC-2
Notified cases of SARS-CoV-2 in Brazil
number of cases
date
Jan 2020
Apr 2020
Jul 2020
Oct 2020
Jan 2021
Apr 2021
Jul 2021
PCA Brazil by cluster - week62
PC-3
PC-4
past
C1-B.1.1.28
C2-B.1.195|other
C3-P.1
C4-B.1.1.33|N9
C5-B.1.1.28
C6-B.1.1|B.1.1.28|other
C7-P.4
C8-P.2
C9-P.1+P.4
C10-P.1+P.4
C11-B.1.1.7
C12-P.1.2
C13-P.4
C14-P.1
C15-P.4

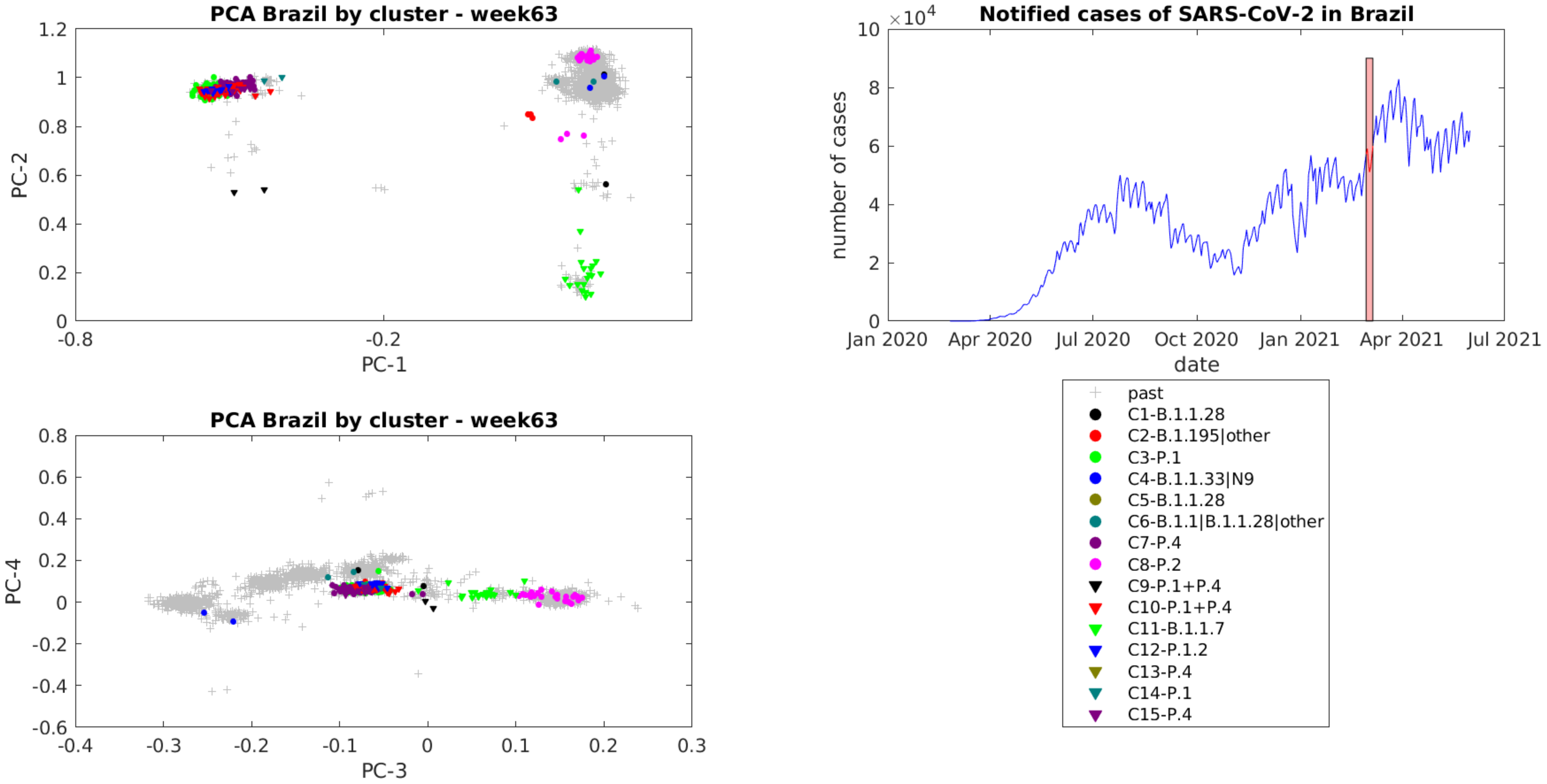
PCA Brazil by cluster - week63
PC-1
PC-2
-0.8
-0.2
0
0.2
0.4
0.6
0.8
1
1.2
PCA Brazil by cluster - week63
PC-3
PC-4
-0.4
-0.3
-0.2
-0.1
0
0.1
0.2
0.3
-0.6
-0.4
-0.2
0.2
0.4
0.6
0.8
Notified cases of SARS-CoV-2 in Brazil
×10^4
number of cases
date
Jan 2020
Apr 2020
Jul 2020
Oct 2020
Jan 2021
Apr 2021
Jul 2021
0
2
4
6
8
10
past
C1-B.1.1.28
C2-B.1.195|other
C3-P.1
C4-B.1.1.33|N9
C5-B.1.1.28
C6-B.1.1|B.1.1.28|other
C7-P.4
C8-P.2
C9-P.1+P.4
C10-P.1+P.4
C11-B.1.1.7
C12-P.1.2
C13-P.4
C14-P.1
C15-P.4

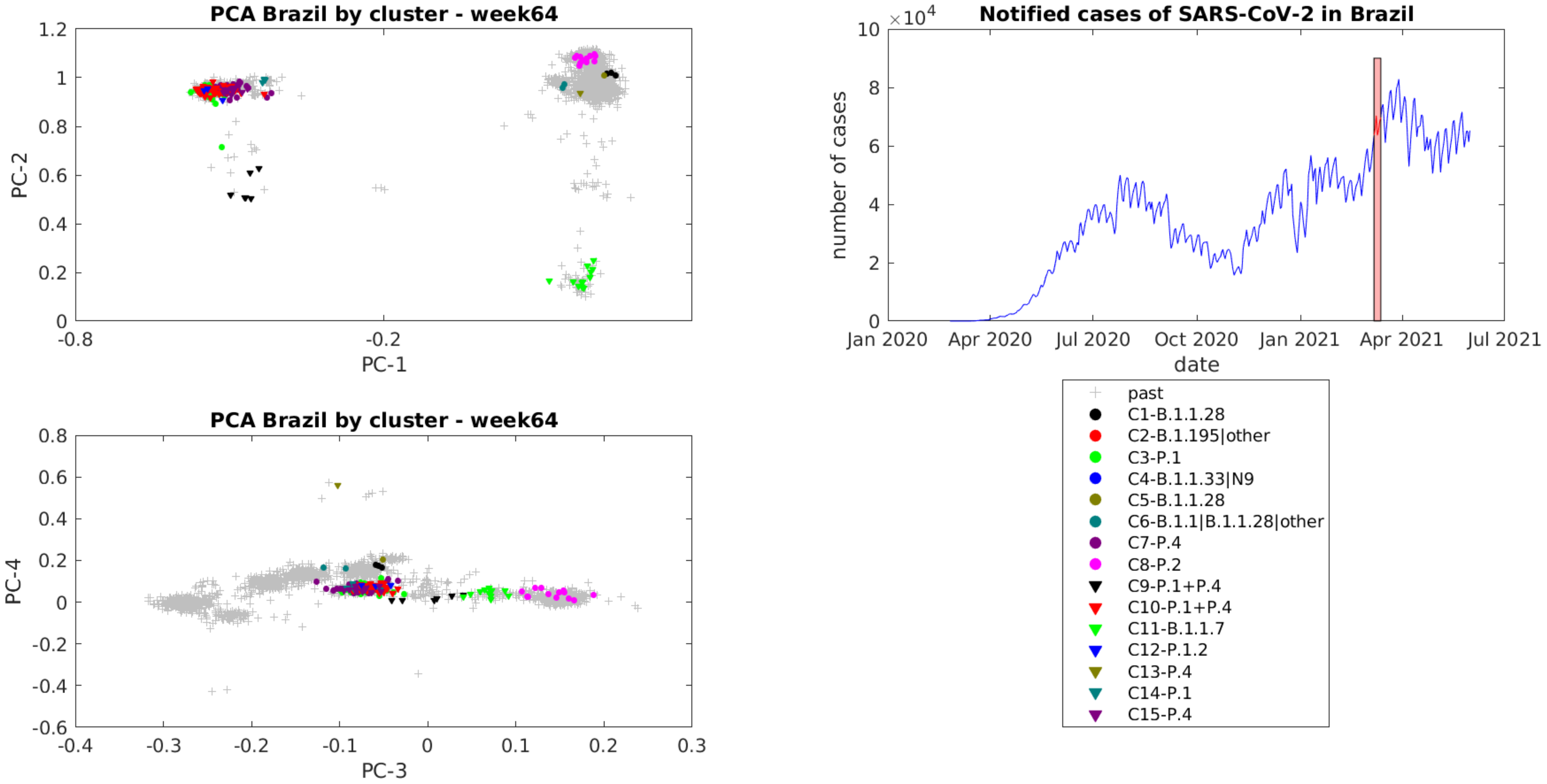
PCA Brazil by cluster - week64
PC-1
PC-2
PCA Brazil by cluster - week64
PC-3
PC-4
Notified cases of SARS-CoV-2 in Brazil
$\times 10^4$
number of cases
date
Jan 2020
Apr 2020
Jul 2020
Oct 2020
Jan 2021
Apr 2021
Jul 2021
past
C1-B.1.1.28
C2-B.1.195|other
C3-P.1
C4-B.1.1.33|N9
C5-B.1.1.28
C6-B.1.1|B.1.1.28|other
C7-P.4
C8-P.2
C9-P.1+P.4
C10-P.1+P.4
C11-B.1.1.7
C12-P.1.2
C13-P.4
C14-P.1
C15-P.4

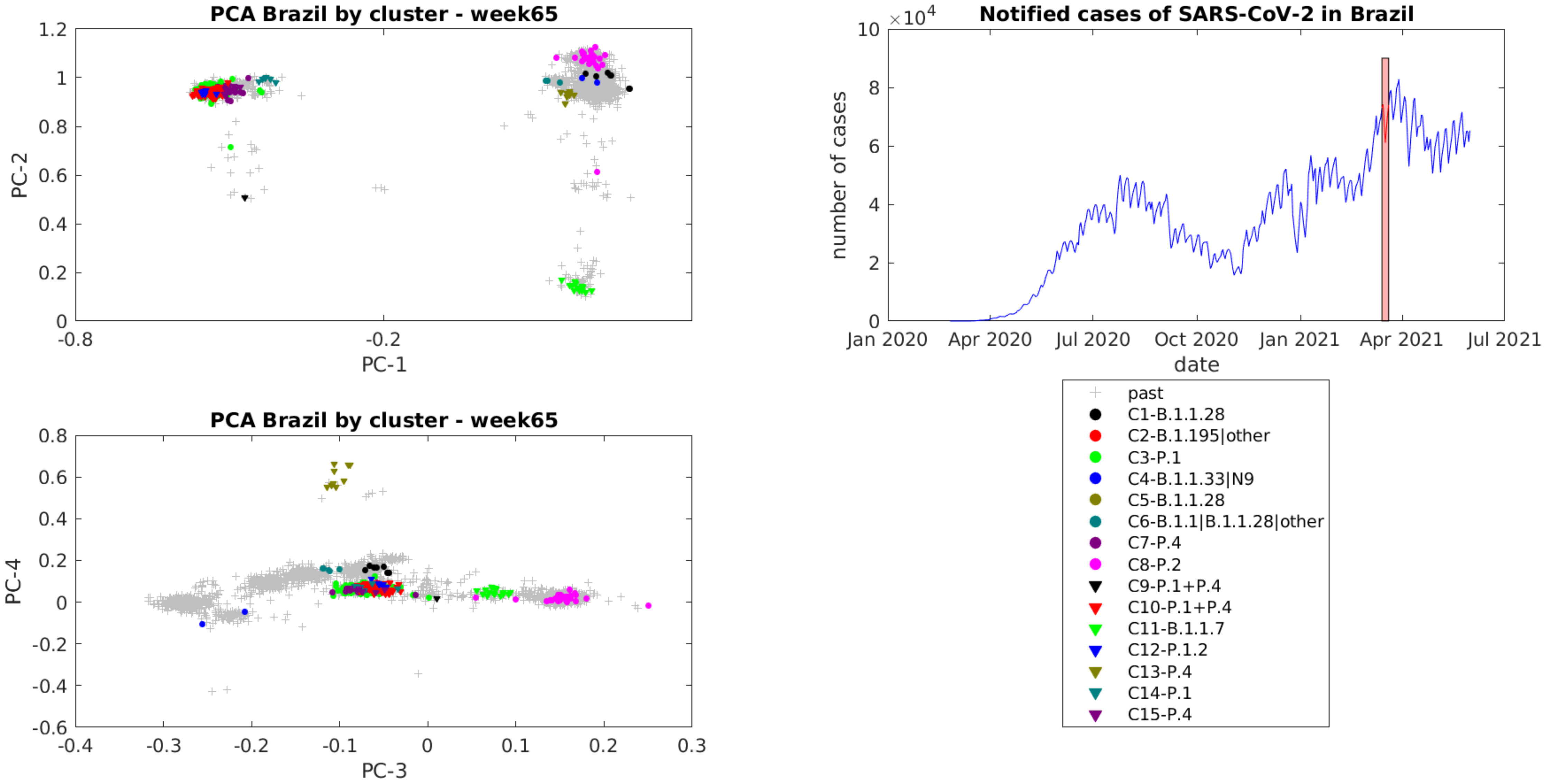
PCA Brazil by cluster - week65
PC-1
PC-2
-0.8
-0.2
0
0.2
0.4
0.6
0.8
1
1.2
Notified cases of SARS-CoV-2 in Brazil
$\times 10^4$
number of cases
date
0
2
4
6
8
10
Jan 2020
Apr 2020
Jul 2020
Oct 2020
Jan 2021
Apr 2021
Jul 2021
PCA Brazil by cluster - week65
PC-3
PC-4
-0.4
-0.3
-0.2
-0.1
0
0.1
0.2
0.3
-0.6
-0.4
-0.2
0
0.2
0.4
0.6
0.8
past
C1-B.1.1.28
C2-B.1.195|other
C3-P.1
C4-B.1.1.33|N9
C5-B.1.1.28
C6-B.1.1|B.1.1.28|other
C7-P.4
C8-P.2
C9-P.1+P.4
C10-P.1+P.4
C11-B.1.1.7
C12-P.1.2
C13-P.4
C14-P.1
C15-P.4

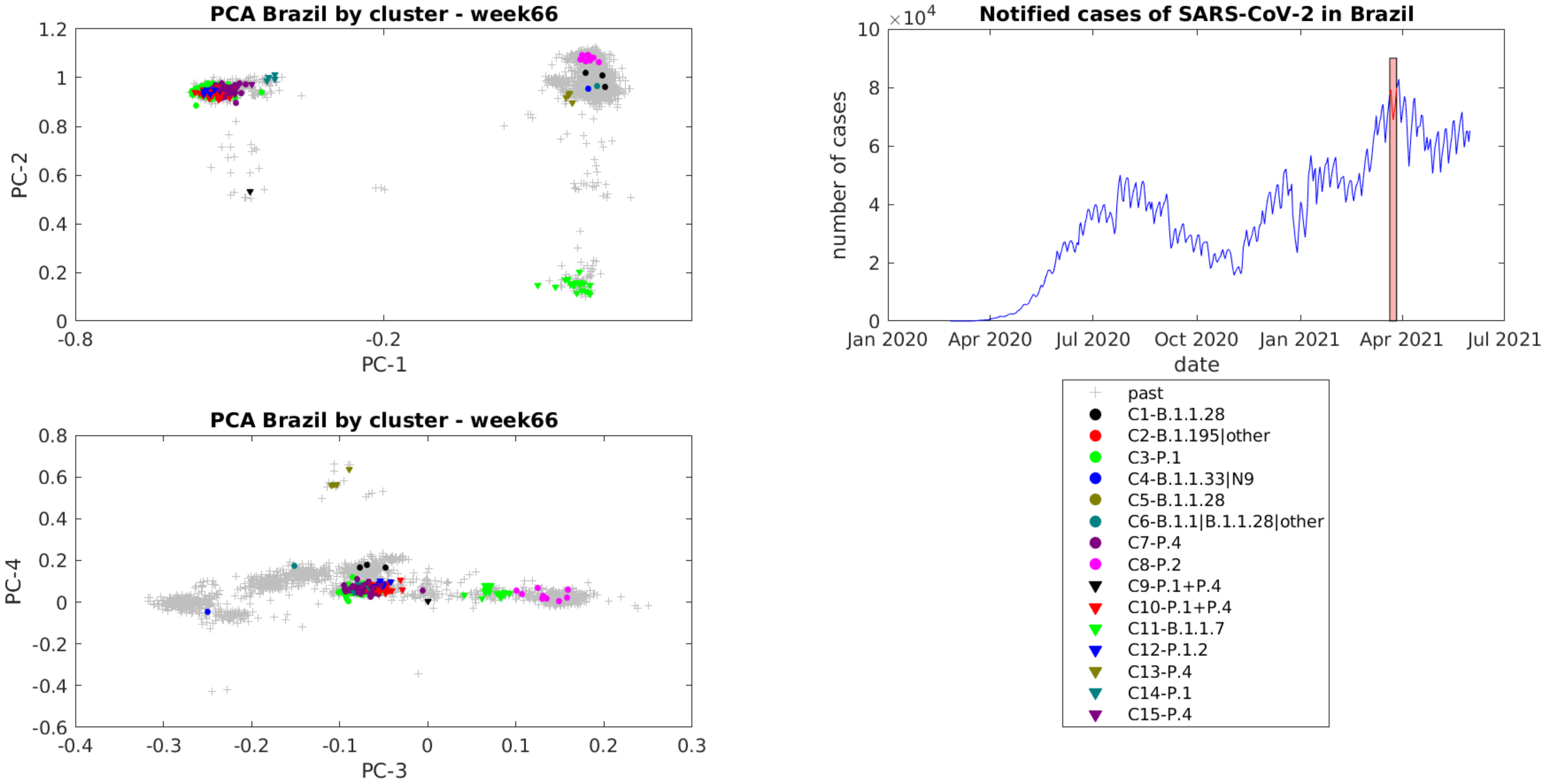

PCA Brazil by cluster - week66
PC-2
PC-1
PCA Brazil by cluster - week66
PC-4
PC-3
Notified cases of SARS-CoV-2 in Brazil
number of cases
date
$\times 10^4$
Jan 2020
Apr 2020
Jul 2020
Oct 2020
Jan 2021
Apr 2021
Jul 2021
past
C1-B.1.1.28
C2-B.1.195|other
C3-P.1
C4-B.1.1.33|N9
C5-B.1.1.28
C6-B.1.1|B.1.1.28|other
C7-P.4
C8-P.2
C9-P.1+P.4
C10-P.1+P.4
C11-B.1.1.7
C12-P.1.2
C13-P.4
C14-P.1
C15-P.4

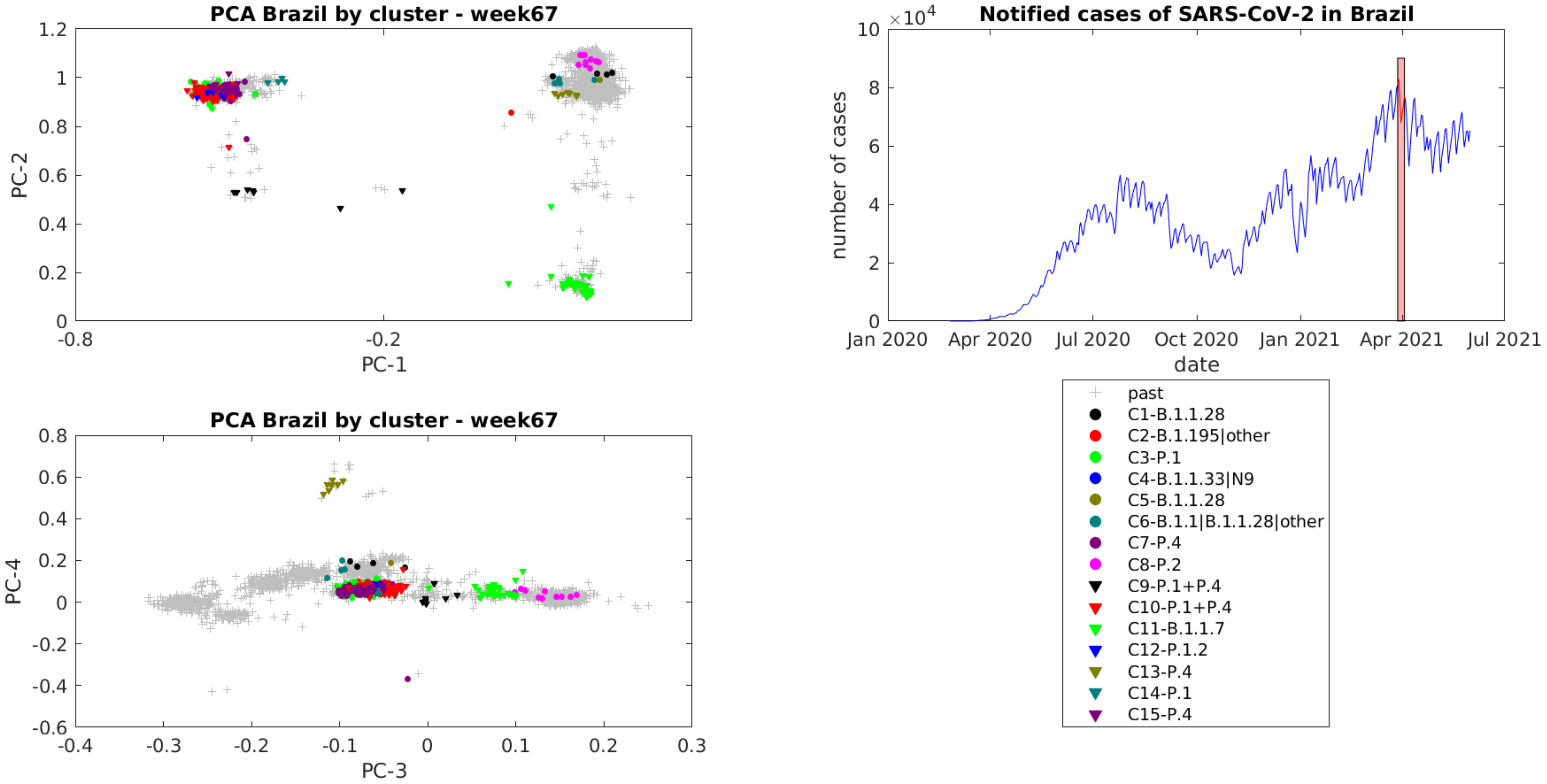

PCA Brazil by cluster - week67
PC-1
PC-2
PCA Brazil by cluster - week67
PC-3
PC-4
Notified cases of SARS-CoV-2 in Brazil
$\times 10^4$
number of cases
date
Jan 2020
Apr 2020
Jul 2020
Oct 2020
Jan 2021
Apr 2021
Jul 2021
past
C1-B.1.1.28
C2-B.1.195|other
C3-P.1
C4-B.1.1.33|N9
C5-B.1.1.28
C6-B.1.1|B.1.1.28|other
C7-P.4
C8-P.2
C9-P.1+P.4
C10-P.1+P.4
C11-B.1.1.7
C12-P.1.2
C13-P.4
C14-P.1
C15-P.4

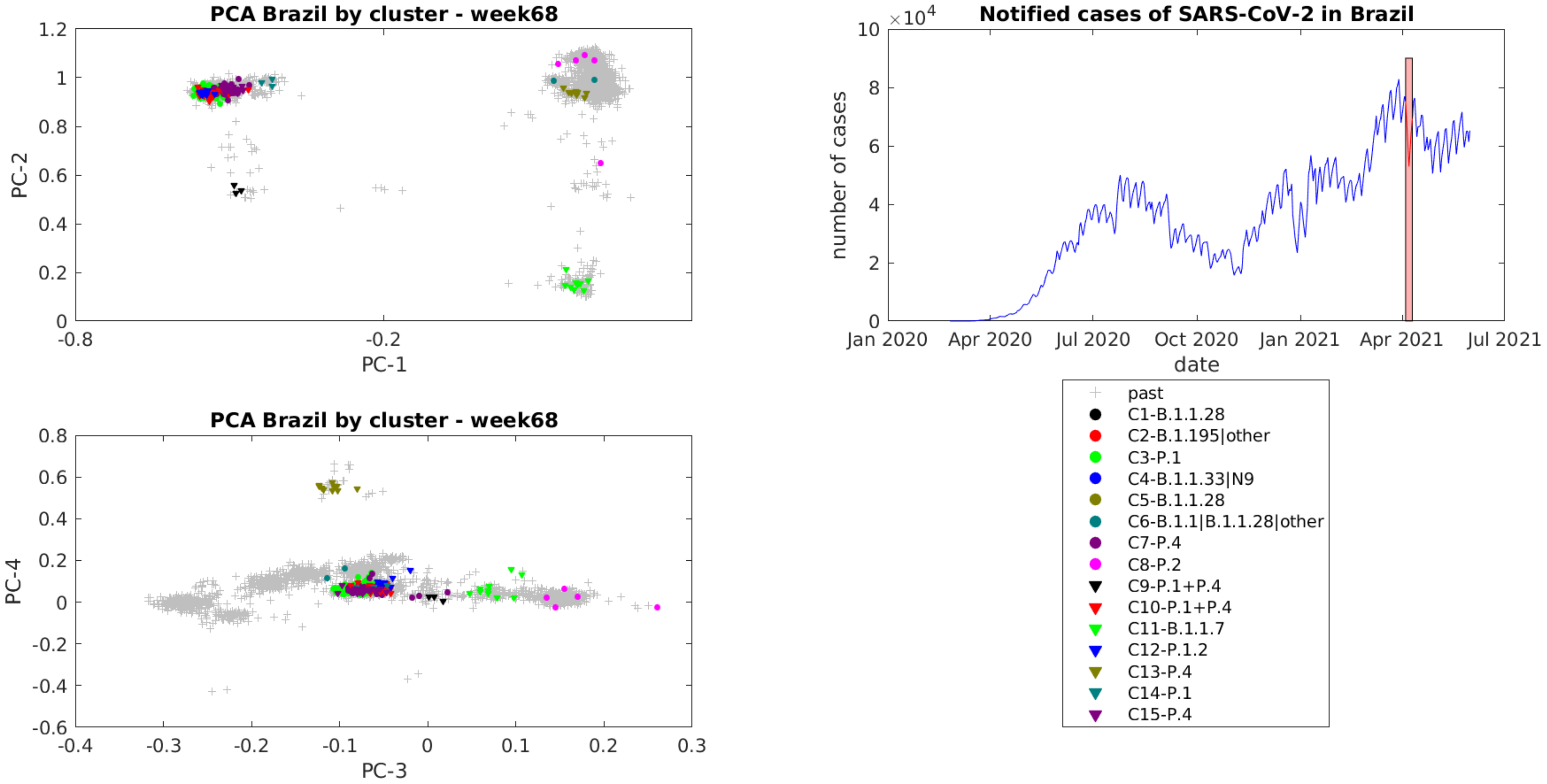
PCA Brazil by cluster - week68
PC-2
PC-1
Notified cases of SARS-CoV-2 in Brazil
$\times 10^4$
number of cases
date
Jan 2020
Apr 2020
Jul 2020
Oct 2020
Jan 2021
Apr 2021
Jul 2021
PCA Brazil by cluster - week68
PC-4
PC-3
past
C1-B.1.1.28
C2-B.1.195|other
C3-P.1
C4-B.1.1.33|N9
C5-B.1.1.28
C6-B.1.1|B.1.1.28|other
C7-P.4
C8-P.2
C9-P.1+P.4
C10-P.1+P.4
C11-B.1.1.7
C12-P.1.2
C13-P.4
C14-P.1
C15-P.4

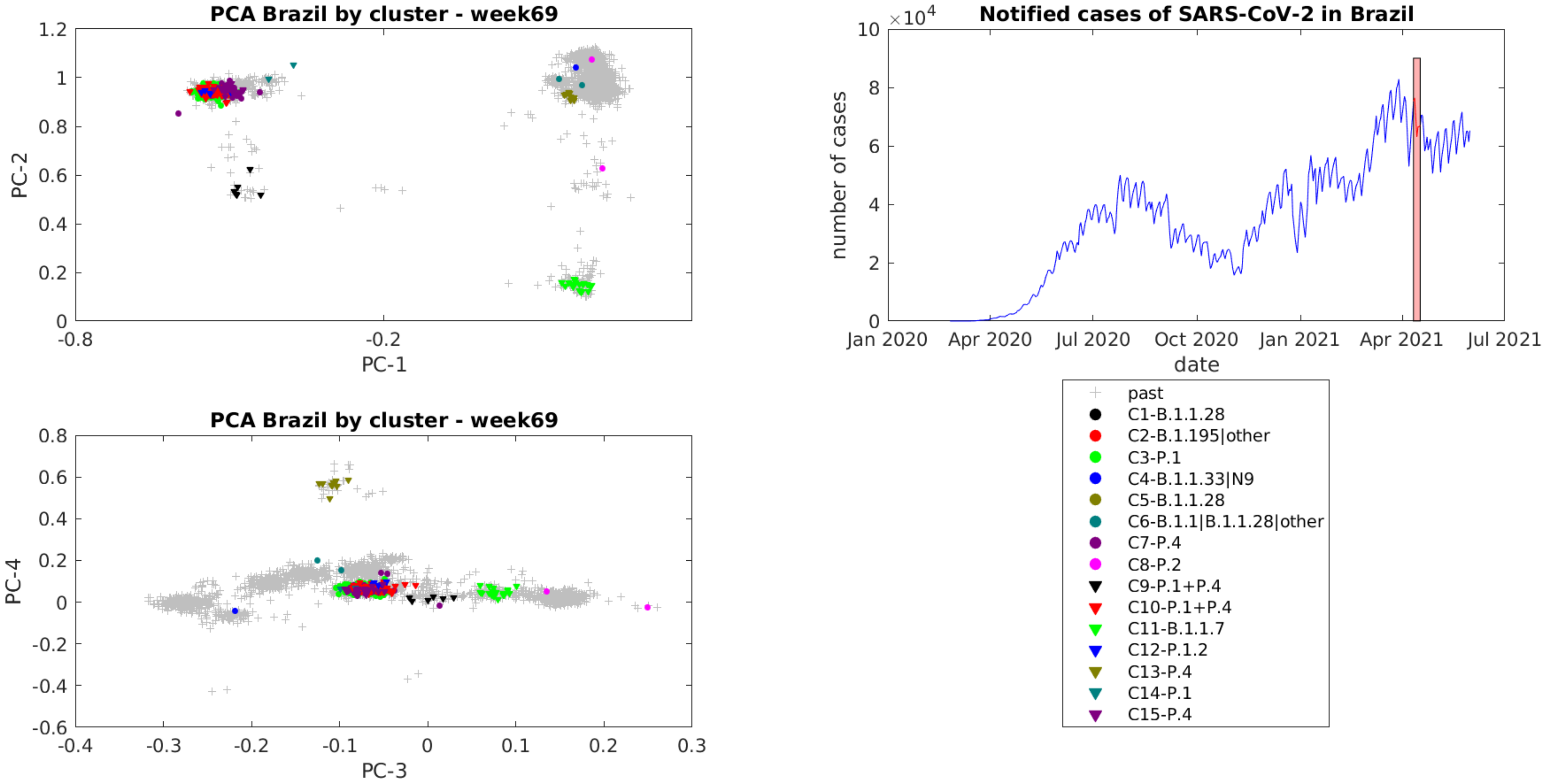
PCA Brazil by cluster - week69
PC-1
PC-2
1.2
1
0.8
0.6
0.4
0.2
0
-0.8
-0.2
PCA Brazil by cluster - week69
PC-3
PC-4
0.8
0.6
0.4
0.2
0
-0.2
-0.4
-0.6
-0.4
-0.3
-0.2
-0.1
0
0.1
0.2
0.3
Notified cases of SARS-CoV-2 in Brazil
$\times 10^4$
10
8
6
4
2
0
number of cases
date
Jan 2020
Apr 2020
Jul 2020
Oct 2020
Jan 2021
Apr 2021
Jul 2021
past
C1-B.1.1.28
C2-B.1.195|other
C3-P.1
C4-B.1.1.33|N9
C5-B.1.1.28
C6-B.1.1|B.1.1.28|other
C7-P.4
C8-P.2
C9-P.1+P.4
C10-P.1+P.4
C11-B.1.1.7
C12-P.1.2
C13-P.4
C14-P.1
C15-P.4

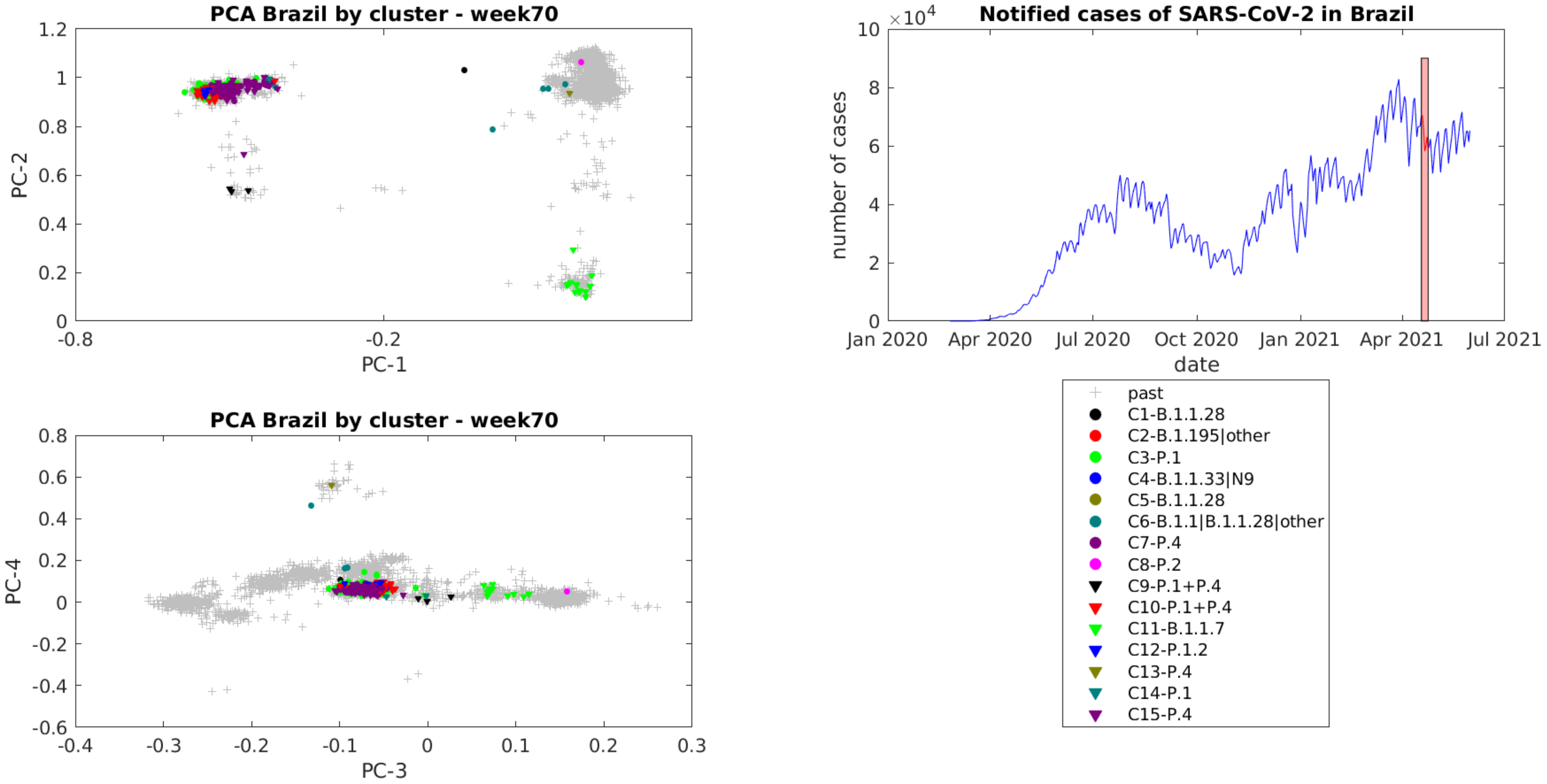
PCA Brazil by cluster - week70
PC-1
PC-2
PCA Brazil by cluster - week70
PC-3
PC-4
Notified cases of SARS-CoV-2 in Brazil
$\times 10^4$
number of cases
date
Jan 2020
Apr 2020
Jul 2020
Oct 2020
Jan 2021
Apr 2021
Jul 2021
past
C1-B.1.1.28
C2-B.1.195|other
C3-P.1
C4-B.1.1.33|N9
C5-B.1.1.28
C6-B.1.1|B.1.1.28|other
C7-P.4
C8-P.2
C9-P.1+P.4
C10-P.1+P.4
C11-B.1.1.7
C12-P.1.2
C13-P.4
C14-P.1
C15-P.4

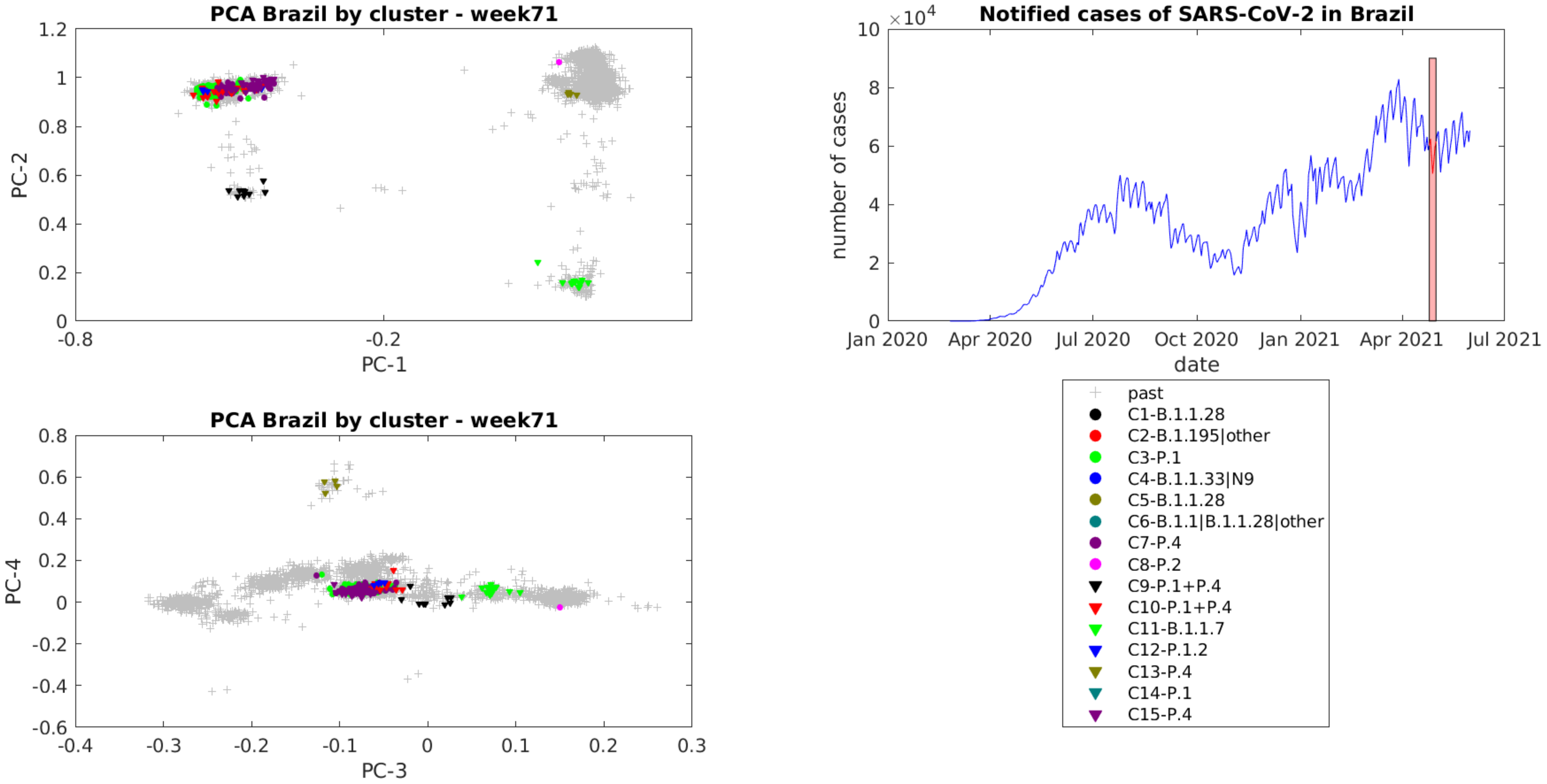
PCA Brazil by cluster - week71
PC-2
PC-1
1.2
1
0.8
0.6
0.4
0.2
0
-0.8
-0.2
Notified cases of SARS-CoV-2 in Brazil
$\times 10^4$
10
8
6
4
2
0
number of cases
date
Jan 2020
Apr 2020
Jul 2020
Oct 2020
Jan 2021
Apr 2021
Jul 2021
PCA Brazil by cluster - week71
PC-4
PC-3
0.8
0.6
0.4
0.2
0
-0.2
-0.4
-0.6
-0.4
-0.3
-0.2
-0.1
0
0.1
0.2
0.3
past
C1-B.1.1.28
C2-B.1.195|other
C3-P.1
C4-B.1.1.33|N9
C5-B.1.1.28
C6-B.1.1|B.1.1.28|other
C7-P.4
C8-P.2
C9-P.1+P.4
C10-P.1+P.4
C11-B.1.1.7
C12-P.1.2
C13-P.4
C14-P.1
C15-P.4

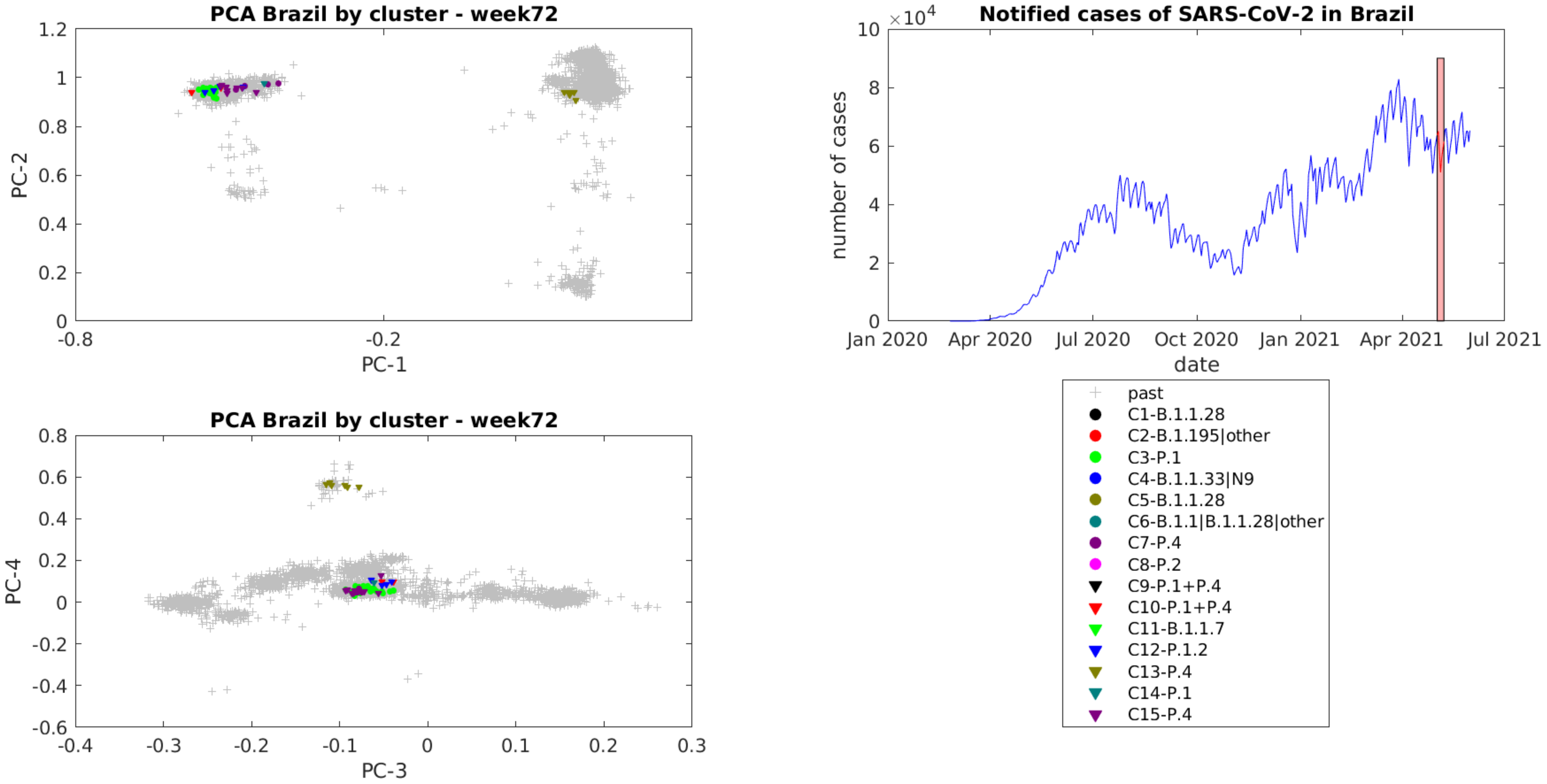
PCA Brazil by cluster - week72
PC-1
PC-2
1.2
1
0.8
0.6
0.4
0.2
0
-0.8
-0.2
Notified cases of SARS-CoV-2 in Brazil
×10^4
10
8
6
4
2
0
number of cases
Jan 2020
Apr 2020
Jul 2020
Oct 2020
Jan 2021
Apr 2021
Jul 2021
date
PCA Brazil by cluster - week72
PC-3
PC-4
0.8
0.6
0.4
0.2
0
-0.2
-0.4
-0.6
-0.4
-0.3
-0.2
-0.1
0
0.1
0.2
0.3
past
C1-B.1.1.28
C2-B.1.195|other
C3-P.1
C4-B.1.1.33|N9
C5-B.1.1.28
C6-B.1.1|B.1.1.28|other
C7-P.4
C8-P.2
C9-P.1+P.4
C10-P.1+P.4
C11-B.1.1.7
C12-P.1.2
C13-P.4
C14-P.1
C15-P.4
